# Whole-genome sequencing of 3,135 individuals representing the genetic diversity of the Japanese population

**DOI:** 10.1101/2025.10.02.25336626

**Authors:** Koichiro Higasa, Yoichiro Kamatani, Takahisa Kawaguchi, Shuji Kawaguchi, Saori Sakaue, Ta-yu Yang, Yukinori Okada, Yukihide Momozawa, Izumi Yamaguchi, Dominic Nelson, Simon Gravel, Yoshinori Murakami, Ryo Yamada, Keitaro Matsuo, Yoshihisa Yamano, Changhoon Kim, Jeong-sun Seo, Michiaki Kubo, Fumihiko Matsuda

**Affiliations:** Department of Genome Analysis, Institute of Biomedical Science, Kansai Medical University, Hirakata 573-1010, Japan; Human Disease Genomics, Center for Genomic Medicine, Kyoto University Graduate School of Medicine, Kyoto 606-8507, Japan; Kyoto-McGill International Collaborative Study in Genomic Medicine, Kyoto University Graduate School of Medicine, Kyoto 606-8507, Japan; Laboratory for Statistical Analysis, RIKEN Center for Integrative Medical Sciences, Yokohama 230-0045, Japan; Department of Statistical Genetics, Osaka University Graduate School of Medicine, Suita 565-0871, Japan; Department of Allergy and Rheumatology, Graduate School of Medicine, the University of Tokyo, Tokyo 113-8655, Japan; Laboratory of Statistical Immunology, Immunology Frontier Research Center (WPI-IFReC), Osaka University, Suita 565-0871, Japan; Integrated Frontier Research for Medical Science Division, Institute for Open and Transdisciplinary Research Initiatives, Osaka University, Suita 565-0871, Japan; Laboratory for Genotyping Development, RIKEN Center for Integrative Medical Sciences, Yokohama 230-0045, Japan; Department of Human Genetics, McGill University, Montreal, Canada, McGill University and Genome Quebec Innovation Centre, Montreal QC H3A0G4, Canada; Division of Molecular Pathology, The Institute of Medical Science, The University of Tokyo, Tokyo 113-8655, Japan; Statistical Genetics, Center for Genomic Medicine, Kyoto University Graduate School of Medicine, Kyoto 606-8507, Japan; Division of Cancer Epidemiology and Prevention, Aichi Cancer Center Research Institute, Aichi 464-8681, Japan; St. Marianna University School of Medicine, Kawasaki 216-8511, Japan; Bioinformatics Institute, Macrogen, Inc., Seoul 153781, Republic of Korea; Precision Medicine Center, Seoul National University Bundang Hospital, Seongnam 13620, Republic of Korea; RIKEN Center for Integrative Medical Sciences, Yokohama 230-0045, Japan

**Keywords:** Whole-genome sequence, genetic diversity of the Japanese population

## Abstract

Whole-genome sequence information currently available for large-scale sequencing studies is biased toward European descent populations. Such bias causes difficulties in identifying disease-associated genetic variations in non-European populations, including the Japanese. Here, to comprehensively identify genetic variants, we sequenced 3,135 individuals representing the genetic diversity of the Japanese population. Of the 44,757,785 identified variants, 31.0% exhibiting a minor allele frequency of < 1% were novel. Using these variants, we constructed a reference haplotype and graph-structured reference sequence to facilitate accurate imputation and variant characterization. Our findings suggest that integrating genetic variations from ethnically diverse populations into the prevailing catalogs is essential to achieve precision medicine for all populations.

## Introduction

The current large-scale genome sequencing programs intend to generate a complete list of genetic variants associated with disease susceptibility, pathogenicity, or drug response for precision medicine. However, most participants of such sequencing and genotyping projects have been of European ancestry. Consequently, approximately 80% of genetic studies have been based on populations of European descent [1, 2], although such individuals make up only 16% of the global population. This bias makes trans-ancestry portability of the polygenic risk score or genetic biomarkers intractable [3], especially for the side effects of drugs, because their frequencies are prone to drift in each population due to neutrality from selective pressure [4]. Thus, there is an urgent need to study other ethnic groups to avoid unexpected therapeutic effects in the under-represented non-European populations [5–8]. Furthermore, numerous contemporary human populations have an admixture of multiple ancestries in their evolutionary history. Therefore, understanding the specific ancestral components is crucial when evaluating the usability or transferability of clinically actionable variants [4].

In this study, we elaborately selected representative individuals of the Japanese population and performed whole-genome sequencing (WGS) and constructed a Japanese reference haplotype panel for accurate genotype imputation of genome-wide association studies and a graph structure of the reference genome sequence to improve variant identification. We incorporated the resources into a public database, the Human Genetic Variation Database (HGVD) [9], to promote the versatility of human genomic studies.

## Materials and Methods

### Participants

This study was approved by the Institutional Review Boards of Aichi Cancer Center, St. Marianna University, Kyoto University, and RIKEN Center for Integrative Medical Sciences (G751). According to institutional and national guidelines, written informed consent with permission to use the data in future research was obtained from all the participants. All data were de-identified to prevent the individual from being identified in the database.

### Sample selection

We applied a greedy algorithm to extract genetically diverse individuals for sequencing from 178,886 previously genotyped samples collected at Kyoto University [10, 11] and BioBank Japan (BBJ) [12]. As a score for relevance, the algorithm counts the number of individuals within a Euclidean distance radius of 3.5 in the high-dimensional principal component (PC) space. To avoid density-dependent oversampling, we initially selected the individuals with the highest score and removed those with the score from the PC space. After recalculating the scores for all other individuals, we selected those with the highest score. We repeated this process until we preserved the requested number of individuals. Owing to the limited available computational resources, we ran the process in the first six PC spaces. Finally, we extracted 3,148 individuals as a representative subset that captured the Japanese genetic background as comprehensively as possible (**Supplementary Fig. S1**).

### WGS and quality control (QC)

WGS was performed using the Illumina HiSeq X Ten sequencer (Illumina Inc., San Diego, CA, USA) for 3,148 individuals **(Supplementary Table S1)**. After aligning the sequence reads to the reference genome (GRCh37/hg19) using the Burrows-Wheeler Aligner [13], downstream analyses, including marking duplicates, base quality recalibration, haplotype calling, joint variant calling, and variant quality score recalibration (VQSR), were processed using Picard and GATK version 3.8 according to the GATK Best Practice recommendations. Of the 3,148 samples, 9 with a high rate of missing genotype in any chromosome (> 0.1), 2 with excess heterozygosity (> 0.05), 1 with a high singleton rate (> 0.001), and 1 with possible sample swapping were excluded from the analysis. For per-marker QC, we excluded 11,105,758 variants with no “PASS” flag or ExcessHet < 60 using VQSR, 274,898 with overlapped deletions, and 187 with significant differences in allele frequencies between BBJ and the other dataset (*p* < 1 × 10^−120^ using logistic regression).

### Population genetics analysis

We excluded 13 possibly related individuals (PI_HAT > 0.4 using PLINK [14]), leaving 3,122 individuals. After filtering out variants within segmental duplications or with low-complexity, strictly masked, or insufficient coverage (< 50% on average) regions, we extracted biallelic SNVs shared between our Japanese and the 1KGP datasets, except for singletons in the 1KGP to avoid ascertainment bias. We excluded variants with MAF of < 5% or > 10% difference in frequency between our Japanese and the JPT datasets. We then pruned SNVs with high LD using PLINK with the “--indep-pairwise 50 10 0.1” option, leaving 103,917 SNVs. PCA was performed using PLINK to identify the genetic makeup of the Japanese dataset. We traced genomic signatures of the population admixture through a global ancestry estimation using ADMIXTURE (version 1.3.0) [15]. ADMIXTURE estimates allele frequencies of variants in K ancestral populations and ancestry proportions for all individual genomes using a maximum likelihood approach. A five-fold cross-validation procedure was applied to determine the best K value. To minimize bias caused by variable sample sizes in the admixture analysis, we downsized the number of Hondo and Ryukyu individuals to a randomly selected subset of 100 individuals. The cutoff number (100) was determined to render the sample sizes even across populations without considerable sample variance loss. *F*_ST_, *F*_3_, and *f*_3_ statistics were calculated using the EIGENSOFT package [16] or AdmixTools (https://github.com/DReichLab/AdmixTools).

### Functional annotation of variants

Following the stringent QC (see above), the remaining 44,757,785 variants were annotated using ANNOVAR [17] and defined as severely damaging if predicted as damaging by the four *in silico* algorithms (PolyPhen2 [18], LRT [19], MutationTaster [20], and SIFT [21]). To identify pLOF variants, we used the LOFTEE v2.0 plugin of Variant Effect Predictor (VEP) v91.3 [22] with default parameters. The variants with MAF of < 1% and annotated as stop-gained, frameshift, or splice sites with high confidence were defined as pLOF.

### Analysis of pLOF variants

To compare pLOF constraints, we aggregated and assigned each pLOF variant onto a gene with a unique Ensembl Gene ID. For the gnomAD dataset, we collected the observed number of pLOF variants per Ensembl Transcript ID. We then converted them into mean values of the expected/observed pLOFs for the genes (Ensembl Gene IDs). We compared the per-gene burden of pLOFs between the Japanese and gnomAD datasets using Pearson’s correlation test. To evaluate the constraint of pLOF variants in drug target genes, we collected information for pharmacologically active targets from the DrugBank and Therapeutic Target databases (http://bidd.nus.edu.sg/group/cjttd). We merged the information with the pharmacological or therapeutic subgroup (2^nd^ level) of the ATC drug classification system to link these drug target genes to the drug categories based on the human diseases to be treated [23]. Using the association of the target genes in a given anatomical organ with the medications of interest for treating that organ, we compared the mean values of the observed/expected number of pLOF variants in the drug target with those in all genes using the Student’s *t*-test.

### Construction of the Japanese haplotype reference panel (JHRP)

Of the 44,757,785 identified variants, 21,033,874 SNVs were used to construct the JHRP with SHAPEIT2 (r904) [24] after filtering out 3,069,172 indels, 535,855 multiallelic SNVs, 18,944,892 singletons, and SNVs with < 95% genotyping success rates (1,116,971 SNVs) or with Hardy Weinberg equilibrium *p*-values (HWE-*p*) of < 1 × 10^−6^ (57,021 SNVs). For filtering indels and multiallelic SNVs, we used Bcftools (http://github.com/samtools/bcftools) with “-v SNP” and “-M 2” options, respectively.

### Power calculation of the JHRP

To assess the performance of the JHRP, we obtained an independent Japanese WGS data from the National Bioscience Database Center (NBDC) and AMED Genome group sharing Database with accession number hum0014-v2 (https://humandbs.biosciencedbc.jp/hum0014-v2) and agd0008-v1 (https://gr-sharingdbs.biosciencedbc.jp/agd0008-v1), respectively. We performed QC using the same criteria as JHRP, leaving 14,487,384 SNVs of 1,294 samples in the standard panel. As an imputation template, we extracted 496,177 autosomal SNVs to imitate the genotype data on Infinium Asian Screening Array version 1.0 (Illumina). Haplotype phasing using SHAPEIT2 [24] and genotype imputation using Minimac4 (https://github.com/statgen/Minimac4) were performed using one of the six reference panels: JHRP, 1KGP, JPT, GAsP, HRP, or TOPMed (**Supplementary Table S2**). Variant sites were divided into 14 bins based on alternative allele frequency in the test set. Imputation accuracy was measured within each bin as the aggregate squared Pearson correlation coefficient (R^2^) between imputed and actual allele dosages in each chromosome. We also used the rsq metric of Minimac4 to assess imputation quality.

### Analysis of SVs

SVs identified using Manta [25] were merged into a single VCF [26] and normalized into 438,065 SVs using Bcftools before being integrated with a collection of known 15,847,592 alternate alleles published from multiple population variation data sources (SBG.Graph.B37.V6.rc6.vcf.gz). The fraction of the SVs that overlapped with centromeres or segmental duplications was filtered. Genome annotations used for filtering were downloaded from the UCSC Genome Browser build hg19. Finally, 16,112,052 alternate alleles were formatted into the VCF file. The resulting VCF file has been deposited in the HGVD database under accession HGV0000016 [9].

### Graph alignment and variant calling

The Graph Genome Pipeline, developed by Seven Bridges Genomics Inc. [27], was downloaded from https://www.sevenbridges.com. Two docker files for graph aligner and variant caller were run on the Docker version 18.09 environment using 48 threads. The merged VCF file was specified using the graph aligner’s “--vcf” option.

## Results

### WGS and variant identification

We selected 3,148 Japanese individuals from 178,886 previously genome-scanned samples, based on principal component analysis (PCA), to widely cover the genetic diversity of the Japanese population (**Supplementary Fig. S1**). A total of 186.5 terabases of DNA sequences were generated and processed using the GATK Best Practice pipeline. On average, 99.5% of the reads were mapped to the reference genome (GRCh37/hg19), which corresponded to 94.3% of the bases covered with at least 10× depth (**Supplementary Table S3 and Supplementary Fig. S2**). We excluded 13 samples from the subsequent analysis owing to possible contamination or a low variant calling rate. From the remaining 3,135 individuals, we identified 44,757,785 variants (41,457,356 single nucleotide variants (SNVs) and 3,300,429 indels) after quality control filtering, of which 11,580,521 (25.9%) were not included in the public databases 3.5KJPNv2 [28] or dbSNP Build 154. Minor allele frequencies (MAFs) of most newly identified variants (11,527,747 or 99.5%) were < 1%.

### Population analysis

The contemporary Japanese population is considered an admixture of descendants of Japan’s early inhabitants called Jomon and more recent immigrants from continental population to form the Yayoi. Since the Yayoi are considered to have inhabited mainland Japan’s central area, the contemporary Japanese population contains a more prominent Chinese–Korean ancestry component. On the other hand, the Ryukyu and Ainu people, who dwell in the Southern and Northern ends of the Japanese archipelago, respectively, inherited more of the Jomon ancestry [29]. Consequently, a dual structure has been established and maintained [29].

To investigate the genetic background of individuals in the present study, we conducted a PCA using the WGS data. As shown in **Fig. 1a**, five clusters were observed from the sequenced individuals. The largest group (N = 2,688) overlapped with the Japanese samples (JPT) from the 1KGP, indicating the Japanese Hondo subpopulation [30]. The lower right cluster (**Fig. 1a**, orange, N = 321) corresponded to the Ryukyu subpopulation [30], and most individuals (91.51%) were from the Okinawa or Kyushu area. We named the other three clusters Pop1, Pop2, and Pop3, indicated in green (N = 51), red (N = 24), and purple (N = 38), respectively (**Fig. 1a**). Given the lower fixation index (*F*_ST_) values between Pop2 and Ryukyu or Hondo/JPT (*F*_ST_ = 0.001, **Fig. 2a**), Pop2 constituted an admixed population between Hondo and Ryukyu, hereinafter referred to as “H/R mix” subpopulation. When we included 39 Korean [31] and 504 East Asian individuals from the 1KGP into the PCA, Pop1 and one JPT individual (NA18976) overlapped with the Korean cluster (**Fig. 1b**), showing that Pop1 represented a subpopulation of Korean ancestry living in Japan.

**Fig. 1.**
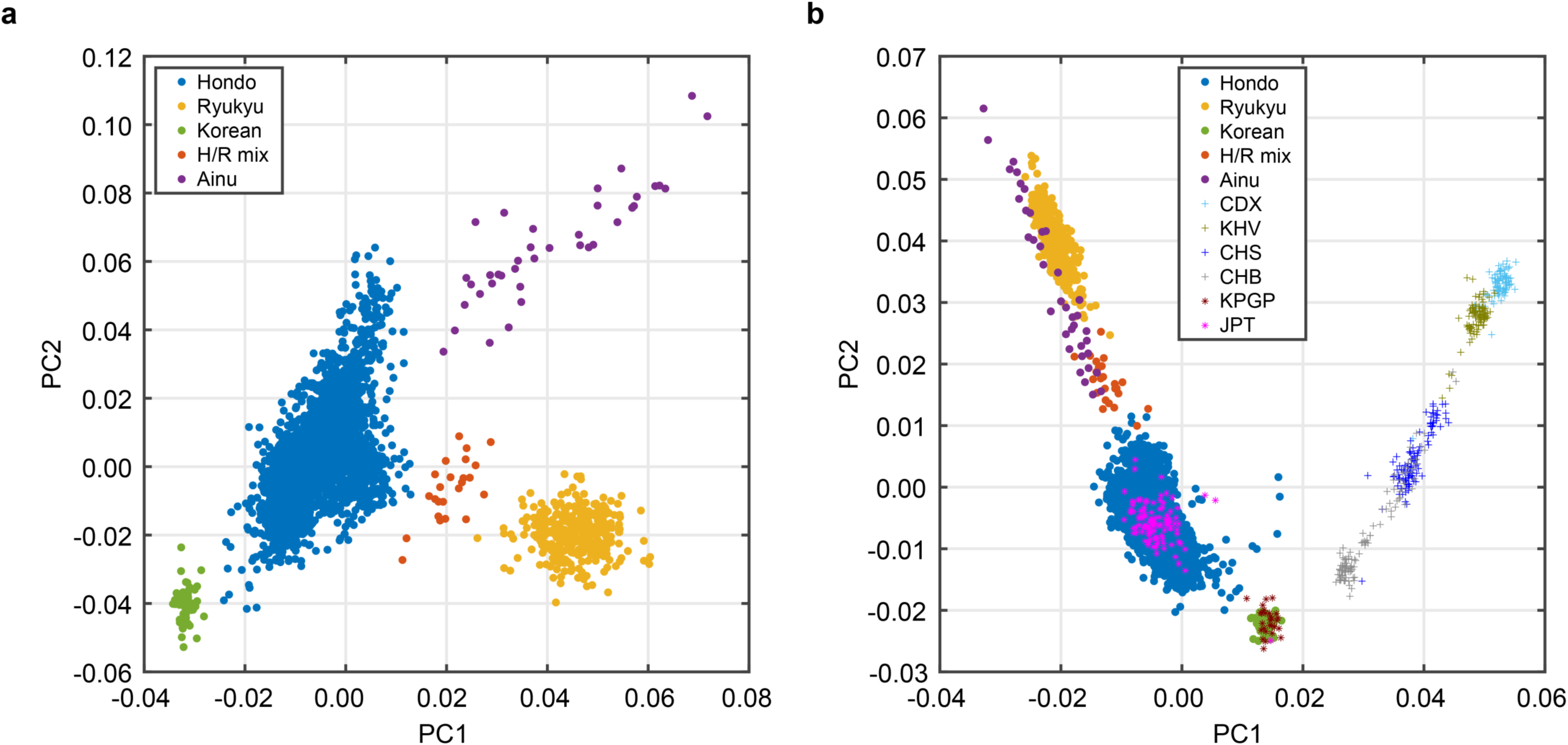
Principal component analysis (PCA) of the Japanese population. (**a**) Five clusters (Hondo, Ryukyu, Korean [Pop1], H/R mix [Pop2], and Ainu [Pop3]) of the Japanese population are shown. (**b**) PCA of East Asian samples from the 1000 Genome Project (1KGP) and Korean samples from the Korean Personal Genome Project (KPGP). JPT and KPGP samples overlapped with Hondo (blue) and Korean (green) clusters, respectively. JPT: Japanese in Tokyo, Japan; CHB: Han Chinese in Beijing, China; CHS: Southern Han Chinese; KHV: Kinh in Ho Chi Minh City, Vietnam; CDX: Chinese Dai in Xishuangbanna, China.

**Fig. 2.**
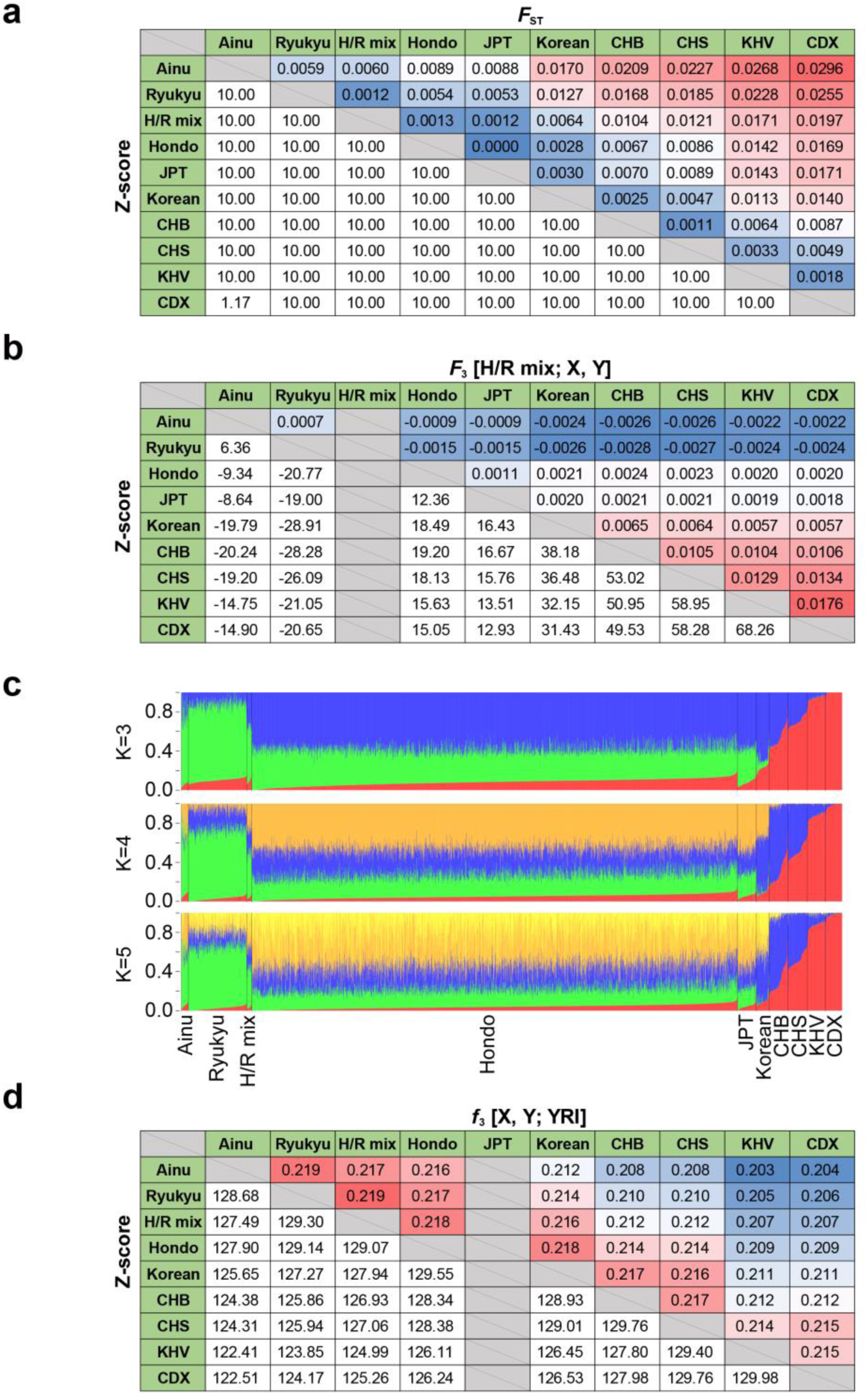
Admixture analysis of the Japanese population. Pairwise estimates of *F*_ST_ (**a**), *F*_3_ [H/R mix; X, Y] (**b**), and *f*_3_ [X, Y; YRI] (**d**) statistics are shown. Red and blue colors represent high and low values for each statistic, respectively. In (**a**), Z-scores are shown in the range of-10–10. (**c**) ADMIXTURE analysis. Error rates of five-fold cross-validation for K = 3, 4, and 5 ancestral components were 0.54850, 0.54845, and 0.54853, respectively. JPT: Japanese in Tokyo, Japan; CHB: Han Chinese in Beijing, China; CHS: Southern Han Chinese; KHV: Kinh in Ho Chi Minh City, Vietnam; CDX: Chinese Dai in Xishuangbanna, China.

To obtain further evidence regarding the five clusters’ admixture, we evaluated the *F*_3_-statistic, representing the shared genetic drift between two populations [32]. The *F*_3_-statistic for [C; A, B] becomes negative if gene flow occurred from both A and B ancestries to C’s ancestry. The *F*_3_-statistics for [H/R mix; Ryukyu, X] were substantively negative except when X was Pop3 (**Fig. 2b**). Consistent with this, *F*_3_-statistics for [H/R mix; Pop3, X] were negative except for Ryukyu. Similarly, the *F*_ST_ value revealed that the cluster most closely related to Pop3 was Ryukyu, the southernmost island in Japan (*F*_ST_ = 0.005947, **Fig. 2a**). However, 92.11% of individuals in Pop3 were from Hokkaido, the northernmost island. Considering that Ryukyu and Pop3 shared the same ancestry, Pop3 was most likely to be the Ainu subpopulation, which supports the dual structure model of the Japanese population [29, 33].

### ADMIXTURE analysis

To infer the Japanese population’s ancestry component, we performed genetic structure analysis with East Asian samples from the 1KGP using ADMIXTURE [15]. We identified a Japanese-specific component (**Fig. 2c**, green) that only appeared in Hondo, Ryukyu, H/R mix, Ainu, and 1KGP JPT samples but not others. This component was most predominant in the Ryukyu and Ainu. The second component, which was most dominant in Hondo (**Fig. 2c**, blue), was shared between the Korean and Northern Chinese populations but made a small or no contribution in the Ryukyu and Ainu, further supporting the dual structure model [29, 33]. To exclude the influence of biased sample size across populations in the estimations, we reduced the numbers of Hondo and Ryukyu individuals before the admixture analysis. Although the lowest cross-validation error rate was observed at K = 3, the difference for K = 4 was minimal (K = 3 in **Supplementary Fig. S3**).

We further evaluated the relative amount of shared genetic drift using the outgroup *f*_3_-statistic, i.e., the shared genetic drift of two populations related to a third [34]. Consistent with the above result, Ainu shared the most substantial history with Ryukyu (*f*_3_ = 0.219, **Fig. 2d**) regardless of the extensive geographical distance compared with the other clusters. A comparison of the f_3_-statistics obtained from 279 individuals from 130 populations in the Simons Genome Diversity Project [35] demonstrated that Hondo, Ryukyu, and Ainu shared the most substantial history with Korean (*f*_3_ = 0.217, 0.212, and 0.211, respectively) followed by Han (**Supplementary Table S4**). Among the populations in Central Asia/Siberia, Hondo shared a similar extent of history with the Northeast Eurasian populations such as Ulchi, Mongolia, Yakut, and Even (*f*_3_ = 0.212, 0.211, 0.202, and 0.202, respectively) (**Supplementary Table S4**). The *f*_3_-statistic did not exceed 0.20 with the other populations except three Oceanian populations, namely Igorot, Dusun, and Hawaiian (**Supplementary Table S4**).

### Characteristics of putative loss-of-function (pLOF) variants

We identified 21,913 pLOF variants in the 3,135 individuals in this study, using the loss-of-function transcript effect estimator (LOFTEE) package [22]. The majority (21,612; 98.6%) had a MAF of < 1% (**Supplementary Table S5**). The median pLOF burden per individual was 278, slightly higher than that in previous studies [36] (**Supplementary Table S5**). The individual pLOF burden was homogenous across the five clusters defined using PCA (data not shown). The number of observed pLOF variants per gene was similar to that in worldwide population genomes (n = 125,748) [36] (Pearson’s *r* = 0.55, *p* < 2.2 × 10^−16^). However, some genes showed substantial differences, e.g., *FLG* harbored 20 pLOF variants in the Japanese but only four in the worldwide population. The mean observed/expected number of pLOFs in drug target genes (0.033 [95% CI: 0.030–0.036]) was lower than that in all genes (0.050 [95% CI: 0.047–0.053]), consistent with the finding of a recent study [37].

When we classified the drug-target genes into 94 of 2^nd^ level categories by the definition of the Anatomical Therapeutic Classification (ATC),[23] the most significant depletion of pLOF was found in genes targeted by antineoplastic and immunomodulating agents (*p* = 1.1 × 10^−10^ using Student’s *t*-test). Besides, individuals who harbor homozygous pLOF variants can be considered “human knock-outs” for a given gene [38]. In our dataset, 240 pLOF variants in 227 genes were homozygous in at least one individual. Although no significant difference in pLOF enrichment was observed between the knock-out-tolerant and drug target genes (Odds Ratio = 0.76, *p* = 0.33 by Fisher’s exact test), 19 of the knock-out-tolerant genes were targets of already approved or developed drugs (**Supplementary Table S6**). These presumptive knock-out individuals’ clinical information might help understand the phenotypic consequences or side effects of the respective medications.

### The increased power of a haplotype reference panel of the Japanese population

As the 3,135 individuals analyzed in this study were selected to cover the Japanese population’s genetic diversity, the data constitute a promising resource for constructing the JHRP for genotype imputation. As expected, the imputation accuracy using the JHRP outperformed that of other panels constructed from a large number of Asian or global population samples [6, 39, 40], especially for variants with lower MAF (**Fig. 3**).

**Fig. 3.**
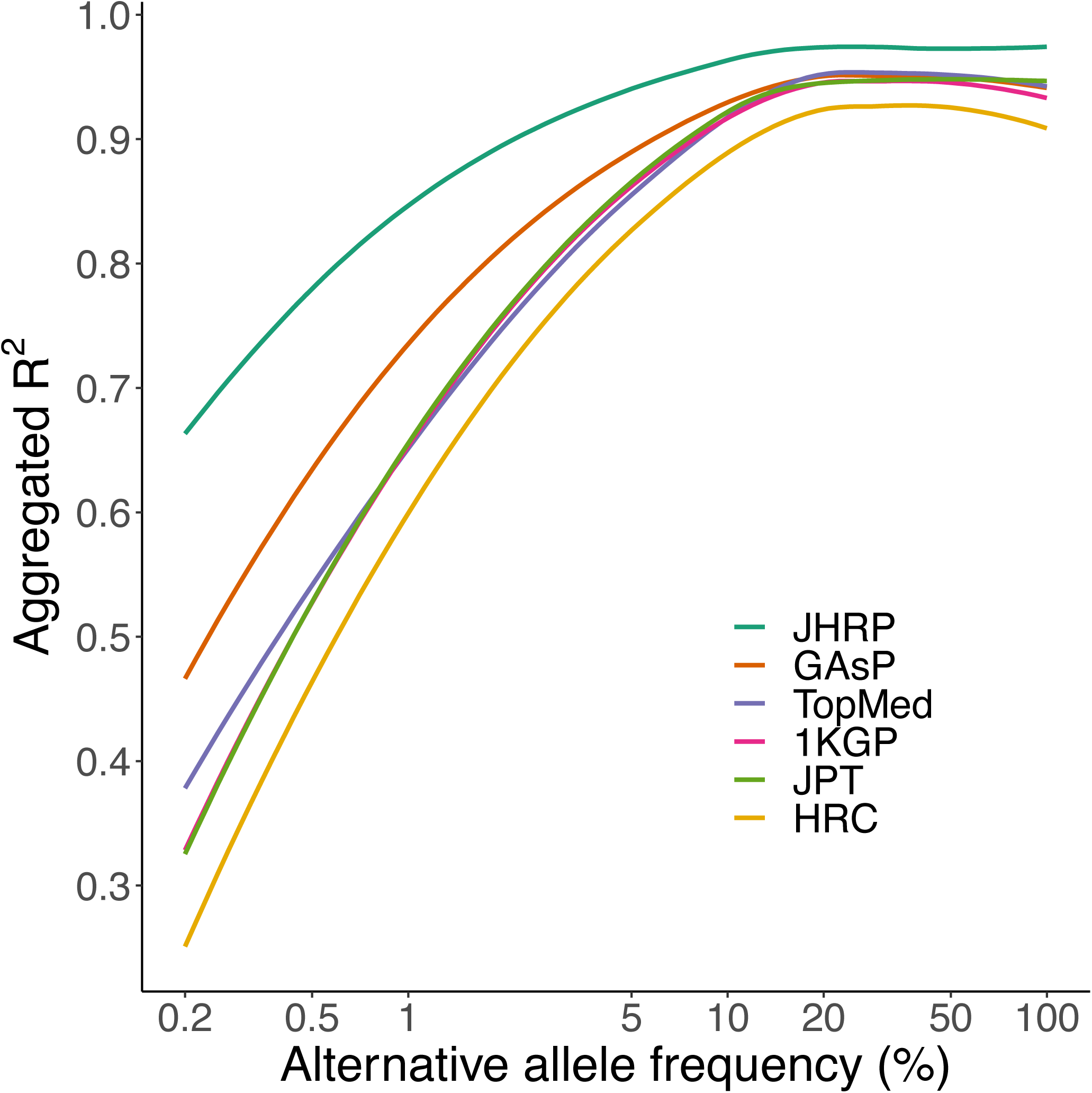
Power of the Japanese haplotype reference panel. Imputation accuracies of the three haplotype reference panels are shown. Aggregated R^2^ values against alternative allele frequency bins of the variants were plotted when the JHRP, 1KGP, JPT, GAsP, HRC or TOPMed was used as the haplotype reference panel. JHRP, Japanese haplotype reference panel; 1KGP, 1000 Genome Project; JPT: Japanese in Tokyo, Japan, GAsP: Genome Asia Pilot, HRC: Haplotype Reference Consortium, TOPMed: Trans-Omics for Precision Medicine.

### Construction of a graph structure of the Japanese reference sequence

Previous studies showed that a graph structure for a reference sequence could improve mapping and variant calling [27, 41–43]. We constructed a graph structure of the Japanese reference sequence by integrating a collection of known alternate alleles in multiple population variation data sources (global; GLB) [27] and 438,056 structural variants (SVs) identified in this study. Consistent with a previous study [27], the accuracy of variant calling within or around SVs was significantly increased owing to the reduction of misalignments and enhancement of coverage uniformity at the boundaries of SVs by placing the reads along the graph path (**Supplementary Fig. S4**). Although the GLB improved the mapping quality compared to using the linear reference, further integration of the Japanese population graph yielded the best performance (*p* < 0.0011 using ANOVA) (**Fig. 4**). Thus, iterative integration of accumulating knowledge of SVs is necessary for optimal analysis using a reference sequence graph structure.

**Fig. 4.**
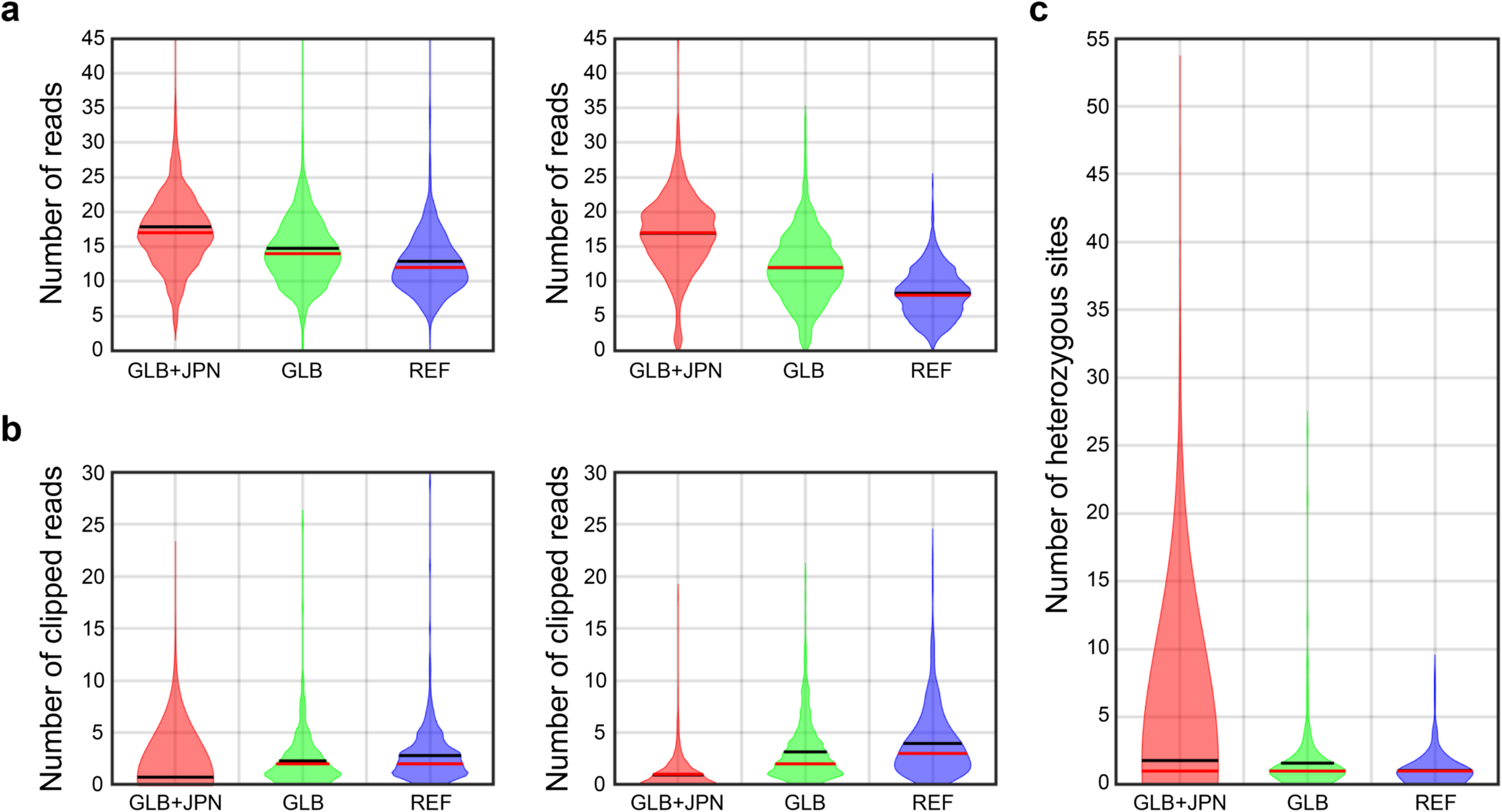
Comparison of the three references. Numbers of aligned reads (**a**), clipped reads (**b**), and correctly genotyped variants per structural variant site (**c**) are shown through the analysis of 691 homozygous and 491 heterozygous deletions with linear (REF), global graph (GLB), and global and Japanese graph (GLB+JPN) references. The mean and median are indicated by black and red lines, respectively.

## Discussion

In the present study, we performed WGS on 3,135 individuals from the Japanese population, which has long been under-represented in public databases. PCA and admixture analysis showed that these individuals covered the genetic diversity of the Japanese population. The constructed haplotype reference panel and graph structure of the reference sequence could facilitate genetic studies and interpretation of variants identified from genetically closer East Asians.

Genotype imputation is a cost-efficient tactic to improve the power and resolution of genome-wide association studies. The imputation accuracy heavily depends on the number of haplotypes of diverse populations included in the reference panel or the imputation template. In particular, rarer or more population-specific variants exacerbate the inaccuracy [44]. The results of the present study showed that the population-matched haplotype reference panel had substantial power, especially for low-frequency and rare variants, at > 90% precision for non-reference homozygous and heterozygous variants. We expect that this high-performance panel will enable the discovery of pathogenic variants in patients with rare diseases.

The current interpretation of genetic variants largely depends on the linear haploid sequence of the human reference genome provided by the international consortium in 2003 [45]. Efforts to upgrade the reference genome are ongoing [46, 47]; however, a considerable proportion in the number and size of genomic variation is difficult to compile into a single reference genome because compatibility issues remain to be addressed [48, 49]. As demonstrated in the present study, the graph structure represents a promising solution to compile the accumulating genetic variations into a higher dimension of reference structure, through which accurate identification and interpretation of variants would be possible. Furthermore, recent potent and intelligent technologies such as graphics processing units and Docker (https://www.docker.com/) have enabled the training of algorithms to decipher and share more extensive and complex structures of genomes [50]. Combining such new technologies and strategies will improve the understanding and interpretation of genomic information and increase clinical usage by shortening the analysis time and establishing new paradigms.

Previously, we developed a genetic variation database, HGVD, to provide information on exonic genetic variations of the Japanese population identified by the whole-exome sequencing of 1,208 Japanese individuals [9]. We have further reinforced the comprehensiveness of HGVD by including additional genetic variations obtained in this study to provide the most substantial catalog of genetic diversity in the Japanese population to date. We have also deposited resources for the imputation and construction of a graph reference. This information could offer enormous benefits for screening and interpreting clinically relevant variants in future precision medicine for all populations.

## Data Availability

All data produced are available online.

https://www.hgvd.genome.med.kyoto-u.ac.jp/

## Acknowledgements

We acknowledge the Nagahama City Office and the nonprofit organization “Zeroji Club” for their assistance in recruiting control individuals. We are grateful to the staff of BBJ for collecting samples.

## Author contributions

RY, JS, MK, and FM designed the study. IY, KM, YY, MK, and FM collected DNA and clinical information. KH, YK, TK, SK, SS, TY, YO, YMO, DN, SG, YMU, and CK analyzed the data. KH, YK, TK, SK, SS, YO, and FM wrote the manuscript. All authors critically reviewed the final version of the manuscript.

## Competing interests

The authors declare no competing financial interests.

## Data availability

The data of this study are available from HGVD (https://www.hgvd.genome.med.kyoto-u.ac.jp/). Phase 3 data of the 1000 Genomes Project are publicly available at ftp://ftp.1000genomes.ebi.ac.uk/vol1/ftp/release/20130502/.

## Funding

This research was funded by JSPS, Grant-in-Aid for Scientific Research (C), KAKENHI Grant Numbers JP17K07255, JP17KT0125, and JP19K07349, Grant-in-Aid for Scientific Research (B), KAKENHI Grant Number JP19H03209 and JP21H03548, and Grant-in-Aid for Young Scientists (B), KAKENHI Grant Number JP16K18474, the Practical Research Project for Rare/Intractable Diseases from the Ministry of Health, Labour and Welfare of Japan, under Grant Numbers 201238002A and 201324049B, and the Practical Research Project for Rare/Intractable Diseases from Japan Agency for Medical Research and Development, AMED, under Grant Numbers JP16ek0109070h0003, JP18kk0205008h0003, JP18kk0205001s0703 JP19ek0109283h0003, and JP19ek0109348h0002.

## Notes

### Competing Interest Statement

The authors have declared no competing interest.

### Author Declarations

Central IRB at Kyoto University gave ethical approval for this work.

