## Supplemental Figures for "Whole-genome sequencing of 3,135 individuals representing the genetic diversity of the Japanese population"

### Supplementary figures

**Supplementary Figure S1**

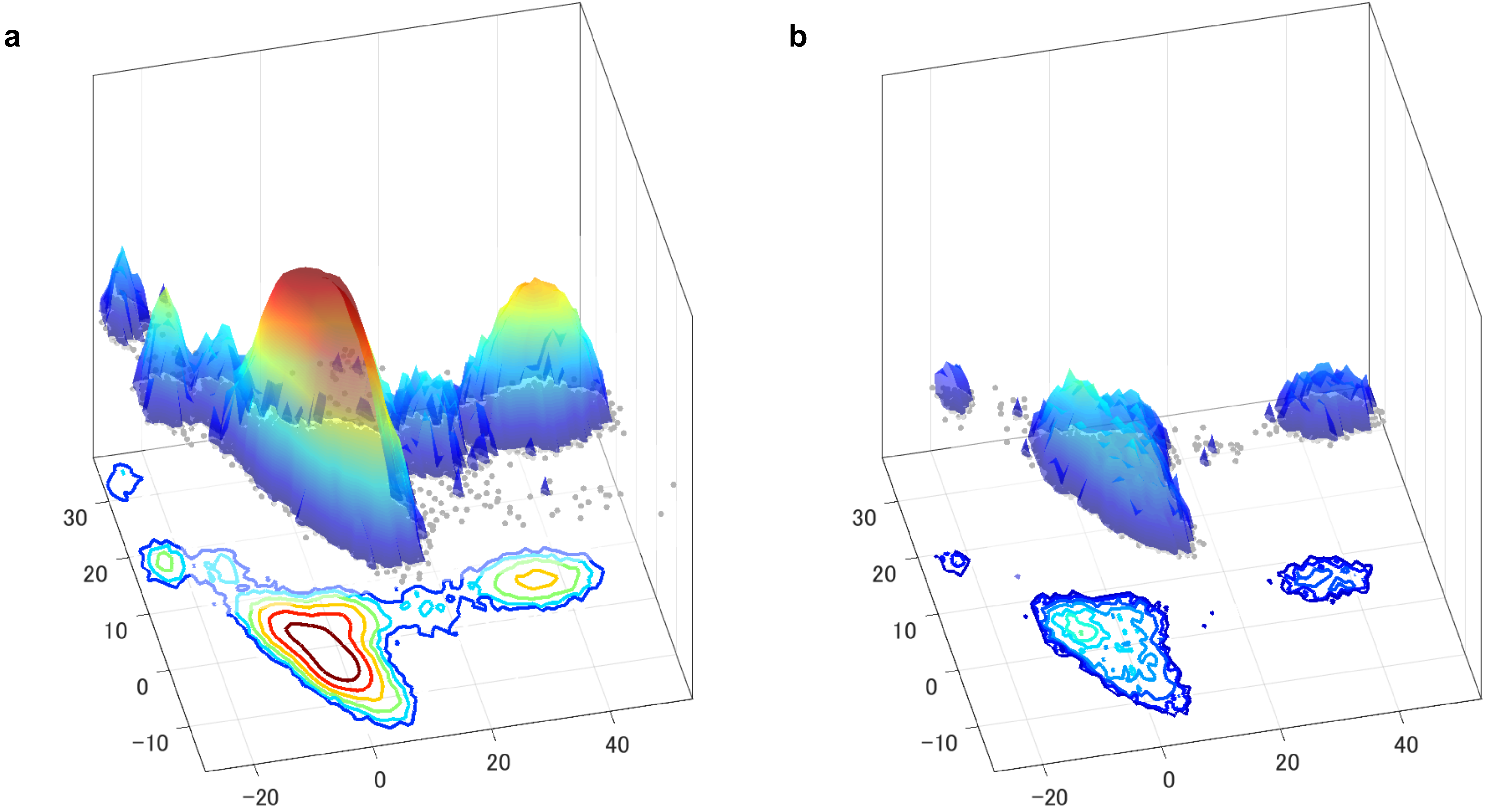

**Supplementary Figure S2**

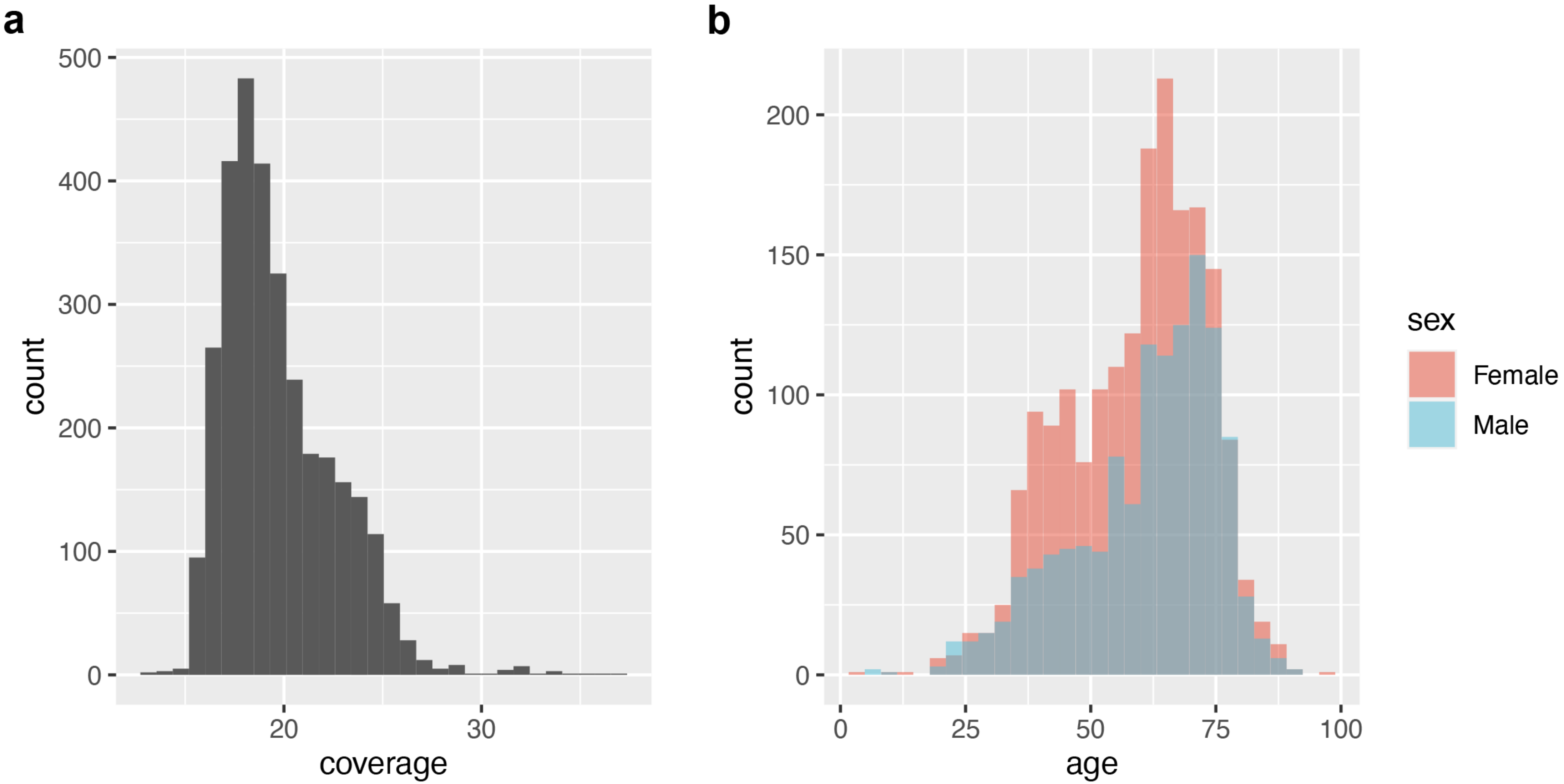

**Supplementary Figure S3**

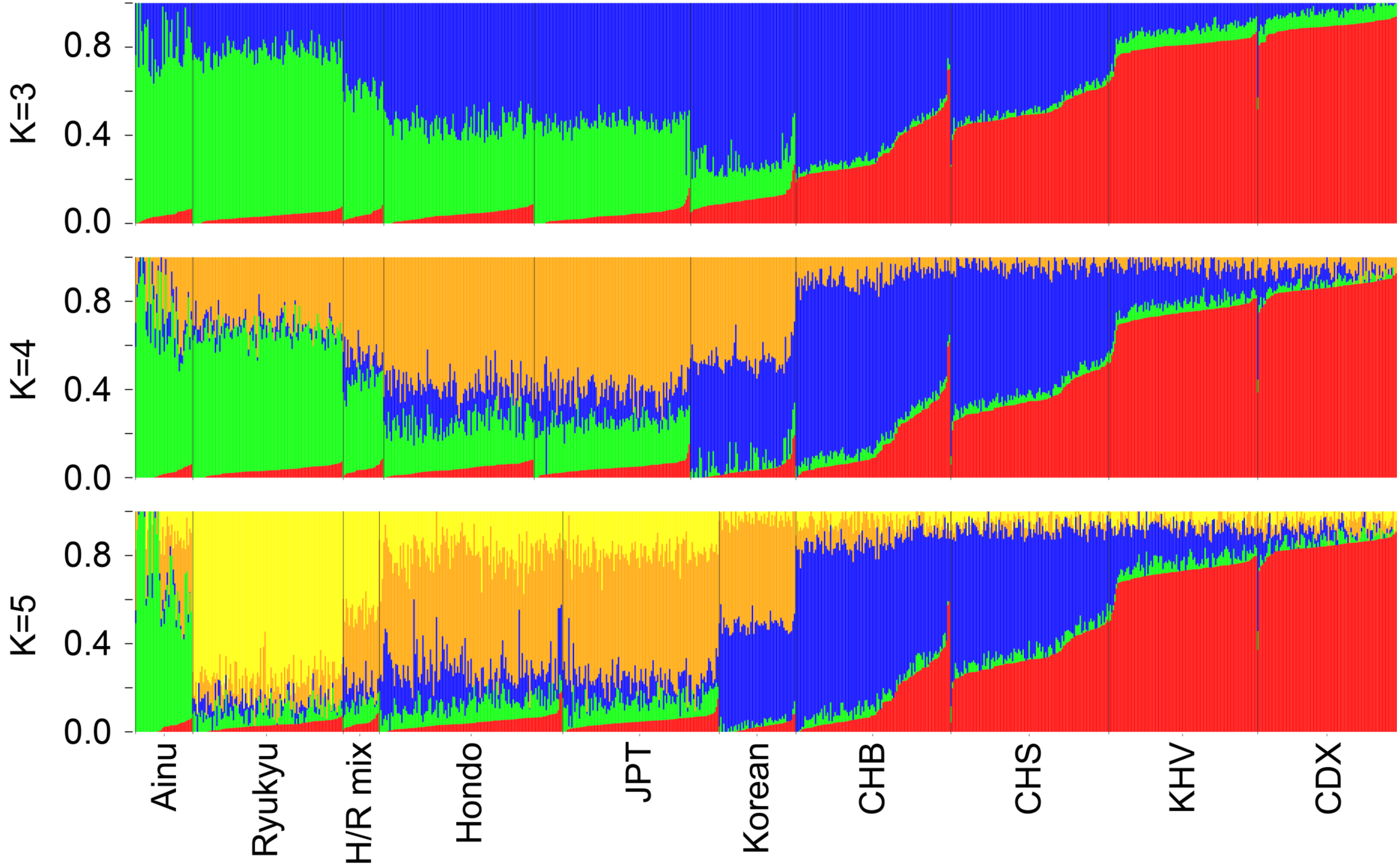

#### Supplementary Figure S4

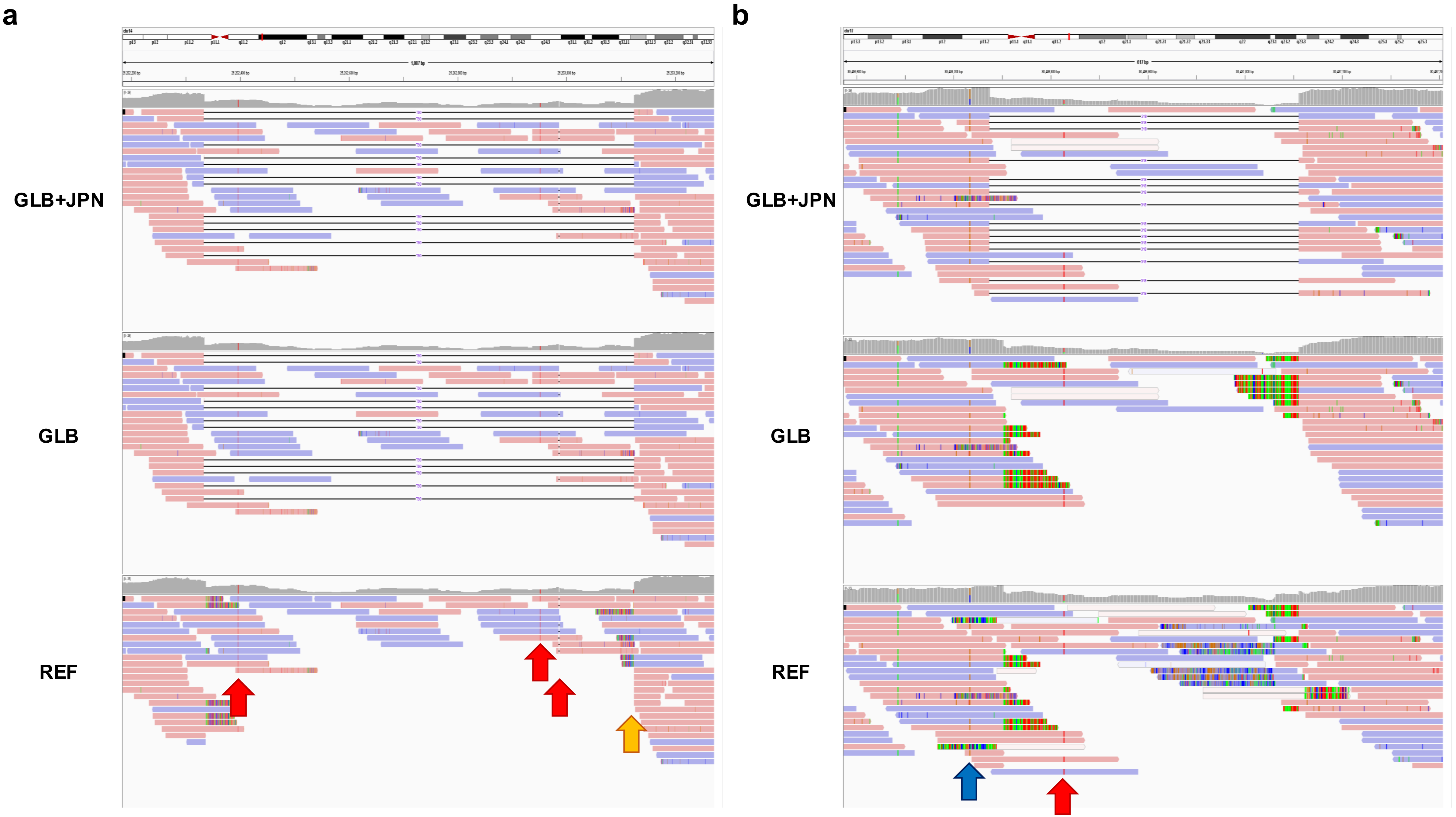

**Supplemental information titles and legends**

**Supplementary Figure S1.** Principal component analysis of the Japanese population. The first and second principal components are plotted with densities for (**a**) 178,886 samples and (**b**) 3,148 representative samples of the Japanese population.

**Supplementary Figure S2.** Basic summary of sequencing across 3,148 individuals. The distribution of (**a**) mean genome-wide coverage and (**b**) age of males and females are shown.

**Supplementary Figure S3.** ADMIXTURE analysis after adjusting the sample size of each cluster. A total of 100 individuals were selected randomly from each Hondo and Ryukyu cluster to avoid overrepresentation bias. Error rates of five-fold cross-validation for K = 2, 3, 4, and 5 ancestral components were 0.56517, 0.56435, 0.56475, and 0.56566, respectively. JPT: Japanese in Tokyo, Japan, CHB: Han Chinese in Beijing, China, CHS: Southern Han Chinese, KHV: Kinh in Ho Chi Minh City, Vietnam, CDX: Chinese Dai in Xishuangbanna, China.

**Supplementary Figure S4.** Advantages of the reference graph structure. Examples of heterozygous deletion sites at chromosome 14 position 25,261,884–25,263,574 (**a**) and chromosome 17 position 30,436,287–30,437,505 (**b**) are apparent through the analysis with linear (REF), global graph (GLB), and global and Japanese graph (GLB+JPN) references. The

GLB+JPN bridges short reads to span SVs in both regions, whereas the GLB only supports region A. Correction of genotypes, increase of supporting reads, and removal of miscalls as observed through analysis of the graph structures, but not the linear reference, are indicated in red, blue, and gold arrows.
