## Supplemental Table for "Whole-genome sequencing of 3,135 individuals representing the genetic diversity of the Japanese population"

Supplementary Table S1. Mapping statistics of individuals

| SampleID | Institute | Geographical region | Number of Read | | Total length(bp) | | % of read | | Coverage | Target Covered >10X |
|---|---|---|---|---|---|---|---|---|---|---|
| | | | Total | Mapped | Total | Mapped | Mapped | Duplicates | | |
| 0000063134 | BBJ | Kanto | 525,697,626 | 523,613,666 | 78,854,643,900 | 78,213,547,191 | 99.60 | 10.84 | 26.07 | 97.31 |
| 0000063140 | BBJ | Kanto | 488,517,806 | 487,000,449 | 73,277,670,900 | 72,742,857,137 | 99.69 | 8.93 | 24.25 | 98.71 |
| 0000063144 | BBJ | Kanto | 473,926,535 | 472,290,793 | 71,088,980,250 | 70,533,496,069 | 99.65 | 7.22 | 23.51 | 98.69 |
| 0000063150 | BBJ | Kanto | 511,374,088 | 508,559,429 | 76,706,113,200 | 75,913,613,216 | 99.45 | 9.96 | 25.30 | 98.88 |
| 0000063154 | BBJ | Kanto | 449,380,780 | 447,156,056 | 67,407,117,000 | 66,777,568,355 | 99.50 | 9.17 | 22.26 | 98.06 |
| 0000063160 | BBJ | Kanto | 483,182,271 | 480,709,425 | 72,477,340,650 | 71,806,489,839 | 99.49 | 11.02 | 23.94 | 96.34 |
| 0000063170 | BBJ | Kanto | 444,117,650 | 442,298,422 | 66,617,647,500 | 66,026,702,105 | 99.59 | 8.61 | 22.01 | 97.88 |
| 0000063174 | BBJ | Kanto | 383,843,849 | 381,985,504 | 57,576,577,350 | 57,027,612,159 | 99.52 | 4.04 | 19.01 | 96.18 |
| 0000063180 | BBJ | Kanto | 415,295,714 | 413,775,621 | 62,294,357,100 | 61,788,971,075 | 99.63 | 9.25 | 20.60 | 94.17 |
| 0000063184 | BBJ | Kanto | 450,331,759 | 448,537,230 | 67,549,763,850 | 66,961,579,513 | 99.60 | 6.32 | 22.32 | 98.33 |
| 0000063190 | BBJ | Kanto | 457,339,703 | 455,404,348 | 68,600,955,450 | 67,985,276,500 | 99.58 | 5.03 | 22.66 | 98.28 |
| 0000063194 | BBJ | Kanto | 460,665,335 | 459,004,042 | 69,099,800,250 | 68,499,786,045 | 99.64 | 7.78 | 22.83 | 96.05 |
| 0000063204 | BBJ | Kanto | 426,132,723 | 424,230,172 | 63,919,908,450 | 63,351,091,919 | 99.55 | 6.62 | 21.12 | 97.65 |
| 0000063210 | BBJ | Kanto | 442,554,342 | 440,163,204 | 66,383,151,300 | 65,651,948,513 | 99.46 | 4.73 | 21.88 | 95.71 |
| 0000063220 | BBJ | Kanto | 338,954,811 | 338,099,565 | 50,843,221,650 | 50,466,333,715 | 99.75 | 5.27 | 16.82 | 91.86 |
| 0000063230 | BBJ | Kanto | 402,878,628 | 401,681,099 | 60,431,794,200 | 59,978,871,546 | 99.70 | 6.73 | 19.99 | 96.83 |
| 0000063630 | BBJ | Kanto | 502,834,779 | 498,866,132 | 75,425,216,850 | 74,442,609,469 | 99.21 | 7.44 | 24.81 | 97.14 |
| 0000063640 | BBJ | Kanto | 447,008,530 | 444,209,276 | 67,051,279,500 | 66,314,873,798 | 99.37 | 5.95 | 22.10 | 97.78 |
| 0000064272 | BBJ | Kanto | 445,726,494 | 443,628,917 | 66,858,974,100 | 66,282,144,644 | 99.53 | 5.36 | 22.09 | 98.40 |
| 0000064282 | BBJ | Kanto | 443,739,318 | 440,749,259 | 66,560,897,700 | 65,846,608,111 | 99.33 | 5.41 | 21.95 | 98.28 |
| 0000064292 | BBJ | Kanto | 488,863,113 | 487,123,317 | 73,329,466,950 | 72,729,080,179 | 99.64 | 11.42 | 24.24 | 96.56 |
| 0000064302 | BBJ | Kanto | 486,449,174 | 484,770,550 | 72,967,376,100 | 72,400,756,535 | 99.65 | 8.98 | 24.13 | 96.79 |
| 0000064312 | BBJ | Kanto | 453,970,070 | 452,201,824 | 68,095,510,500 | 67,523,830,281 | 99.61 | 6.86 | 22.51 | 96.33 |
| 0000064322 | BBJ | Kanto | 448,660,677 | 445,367,063 | 67,299,101,550 | 66,523,102,963 | 99.27 | 8.15 | 22.17 | 95.41 |
| 0000064332 | BBJ | Kanto | 399,818,088 | 398,059,550 | 59,972,713,200 | 59,417,995,997 | 99.56 | 4.79 | 19.81 | 94.11 |
| 0000064342 | BBJ | Kanto | 393,481,119 | 391,969,443 | 59,022,167,850 | 58,508,477,782 | 99.62 | 8.07 | 19.50 | 93.08 |
| 0000064352 | BBJ | Kanto | 380,993,931 | 379,268,452 | 57,149,089,650 | 56,620,192,696 | 99.55 | 6.49 | 18.87 | 95.52 |
| 0000064362 | BBJ | Kanto | 429,247,764 | 427,146,828 | 64,387,164,600 | 63,784,454,382 | 99.51 | 6.71 | 21.26 | 95.13 |
| 0000064372 | BBJ | Kanto | 407,377,744 | 405,850,750 | 61,106,661,600 | 60,593,577,508 | 99.63 | 5.50 | 20.20 | 94.47 |
| 0000064382 | BBJ | Kanto | 483,230,226 | 481,111,641 | 72,484,533,900 | 71,803,322,871 | 99.56 | 10.50 | 23.93 | 98.43 |
| 0000064392 | BBJ | Kanto | 433,416,559 | 430,278,116 | 65,012,483,850 | 64,237,566,671 | 99.28 | 5.64 | 21.41 | 95.60 |
| 0000064402 | BBJ | Kanto | 508,803,265 | 505,763,430 | 76,320,489,750 | 75,487,875,982 | 99.40 | 10.04 | 25.16 | 96.93 |
| 0000064412 | BBJ | Kanto | 428,187,501 | 425,159,624 | 64,228,125,150 | 63,473,048,316 | 99.29 | 5.80 | 21.16 | 97.49 |
| 0000064422 | BBJ | Kanto | 431,417,617 | 429,070,685 | 64,712,642,550 | 64,054,014,345 | 99.46 | 7.99 | 21.35 | 97.59 |
| 0000064442 | BBJ | Kanto | 416,142,575 | 414,730,290 | 62,421,386,250 | 61,907,514,238 | 99.66 | 8.39 | 20.64 | 94.50 |
| 0000064452 | BBJ | Kanto | 395,628,935 | 393,944,398 | 59,344,340,250 | 58,806,146,883 | 99.57 | 6.32 | 19.60 | 96.60 |
| 0000064886 | BBJ | Kanto | 383,526,277 | 381,688,837 | 57,528,941,550 | 56,997,034,691 | 99.52 | 6.09 | 19.00 | 96.02 |
| 0000064896 | BBJ | Kanto | 382,530,561 | 380,011,425 | 57,379,584,150 | 56,741,349,521 | 99.34 | 4.53 | 18.91 | 93.42 |
| 0000064906 | BBJ | Kanto | 457,286,075 | 455,468,635 | 68,592,911,250 | 68,021,966,409 | 99.60 | 7.30 | 22.67 | 98.46 |
| 0000064916 | BBJ | Kanto | 408,647,255 | 406,382,305 | 61,297,088,250 | 60,699,095,783 | 99.45 | 4.98 | 20.23 | 94.89 |
| 0000064926 | BBJ | Kanto | 415,215,675 | 413,626,735 | 62,282,351,250 | 61,746,496,555 | 99.62 | 8.79 | 20.58 | 94.30 |
| 0000064932 | BBJ | Kanto | 400,832,029 | 398,387,400 | 60,124,804,350 | 59,484,141,762 | 99.39 | 6.26 | 19.83 | 96.61 |
| 0000064936 | BBJ | Kanto | 450,049,176 | 446,855,736 | 67,507,376,400 | 66,725,231,939 | 99.29 | 7.98 | 22.24 | 95.57 |
| 0000065858 | BBJ | Kanto | 388,674,546 | 386,900,512 | 58,301,181,900 | 57,776,481,048 | 99.54 | 6.96 | 19.26 | 96.21 |
| 0000065868 | BBJ | Kanto | 498,322,630 | 496,739,692 | 74,748,394,500 | 74,198,424,719 | 99.68 | 10.88 | 24.73 | 98.81 |
| 0000065878 | BBJ | Kanto | 499,768,724 | 497,727,669 | 74,965,308,600 | 74,339,884,248 | 99.59 | 7.72 | 24.78 | 98.86 |
| 0000065888 | BBJ | Kanto | 445,054,436 | 443,005,599 | 66,758,165,400 | 66,140,808,277 | 99.54 | 8.36 | 22.05 | 98.02 |
| 0000065898 | BBJ | Kanto | 401,641,681 | 400,295,615 | 60,246,252,150 | 59,762,530,593 | 99.66 | 8.78 | 19.92 | 93.50 |
| 0000065908 | BBJ | Kanto | 389,647,392 | 387,877,895 | 58,447,108,800 | 57,887,353,949 | 99.55 | 3.59 | 19.30 | 93.71 |
| 0000065918 | BBJ | Kanto | 429,056,525 | 426,676,372 | 64,358,478,750 | 63,711,759,072 | 99.45 | 8.56 | 21.24 | 94.84 |
| 0000065928 | BBJ | Kanto | 455,472,466 | 452,455,966 | 68,320,869,900 | 67,569,916,895 | 99.34 | 5.65 | 22.52 | 98.23 |
| 0000065938 | BBJ | Kanto | 503,868,125 | 500,111,610 | 75,580,218,750 | 74,669,857,796 | 99.25 | 8.80 | 24.89 | 97.06 |
| 0000065948 | BBJ | Kanto | 464,503,002 | 462,224,664 | 69,675,450,300 | 69,001,100,366 | 99.51 | 8.02 | 23.00 | 98.35 |
| 0000065958 | BBJ | Kanto | 428,224,436 | 425,497,198 | 64,233,665,400 | 63,531,586,929 | 99.36 | 7.59 | 21.18 | 97.42 |
| 0000065968 | BBJ | Kanto | 376,747,844 | 375,309,972 | 56,512,176,600 | 56,029,981,072 | 99.62 | 7.48 | 18.68 | 94.92 |
| 0999932959 | BBJ | Kanto | 416,738,528 | 414,040,287 | 62,510,779,200 | 61,804,822,409 | 99.35 | 7.62 | 20.60 | 94.05 |
| 0999932979 | BBJ | Kanto | 449,683,113 | 447,567,005 | 67,452,466,950 | 66,787,988,898 | 99.53 | 6.92 | 22.26 | 98.05 |
| 0999933439 | BBJ | Kanto | 441,980,312 | 440,392,039 | 66,297,046,800 | 65,741,644,189 | 99.64 | 10.63 | 21.91 | 97.60 |
| 0999933449 | BBJ | Kanto | 408,454,989 | 404,531,987 | 61,268,248,350 | 60,405,759,958 | 99.04 | 4.03 | 20.14 | 97.13 |
| 0999933459 | BBJ | Kanto | 453,263,432 | 450,454,003 | 67,989,514,800 | 67,274,469,167 | 99.38 | 6.07 | 22.42 | 96.04 |
| 0999933469 | BBJ | Kanto | 490,312,550 | 487,095,152 | 73,546,882,500 | 72,716,662,764 | 99.34 | 5.64 | 24.24 | 97.02 |
| 0999933479 | BBJ | Kanto | 499,309,432 | 496,808,579 | 74,896,414,800 | 74,170,092,778 | 99.50 | 9.16 | 24.72 | 98.77 |
| 0999933489 | BBJ | Kanto | 394,666,381 | 393,327,185 | 59,199,957,150 | 58,698,442,216 | 99.66 | 6.88 | 19.57 | 93.50 |
| 0999933499 | BBJ | Kanto | 480,783,339 | 479,134,155 | 72,117,500,850 | 71,565,967,024 | 99.66 | 7.87 | 23.86 | 98.72 |
| 0999933509 | BBJ | Kanto | 486,080,303 | 483,987,715 | 72,912,045,450 | 72,284,289,652 | 99.57 | 9.63 | 24.09 | 96.61 |
| 0999933519 | BBJ | Kanto | 494,882,651 | 492,040,811 | 74,232,397,650 | 73,486,483,113 | 99.43 | 8.51 | 24.50 | 97.01 |
| 0999934233 | BBJ | Kanto | 409,926,927 | 407,920,252 | 61,489,039,050 | 60,893,661,391 | 99.51 | 6.46 | 20.30 | 94.32 |
| 0999934651 | BBJ | Kanto | 513,765,032 | 509,674,929 | 77,064,754,800 | 76,078,886,918 | 99.20 | 7.95 | 25.36 | 98.85 |
| 0999934671 | BBJ | Kanto | 508,134,235 | 503,050,711 | 76,220,135,250 | 75,117,612,051 | 99.00 | 10.11 | 25.04 | 98.74 |
| 0999934713 | BBJ | Kanto | 393,289,053 | 391,704,067 | 58,993,357,950 | 58,473,513,467 | 99.60 | 7.67 | 19.49 | 95.96 |
| 0999934723 | BBJ | Kanto | 366,804,929 | 365,377,065 | 55,020,739,350 | 54,541,835,949 | 99.61 | 7.02 | 18.18 | 91.53 |
| 0999934733 | BBJ | Kanto | 464,821,586 | 462,765,955 | 69,723,237,900 | 69,057,155,904 | 99.56 | 5.33 | 23.02 | 98.66 |
| 0999934743 | BBJ | Kanto | 485,111,382 | 482,257,849 | 72,766,707,300 | 71,927,178,937 | 99.41 | 4.63 | 23.98 | 98.83 |
| 0999934753 | BBJ | Kanto | 457,512,929 | 453,940,128 | 68,626,939,350 | 67,805,443,892 | 99.22 | 8.58 | 22.60 | 98.10 |
| 0999934763 | BBJ | Kanto | 454,222,802 | 450,106,127 | 68,133,420,300 | 67,193,529,327 | 99.09 | 5.00 | 22.40 | 96.22 |
| 0999934773 | BBJ | Kanto | 403,226,149 | 401,850,459 | 60,483,922,350 | 59,954,634,562 | 99.66 | 7.53 | 19.98 | 93.71 |
| 0999934783 | BBJ | Kanto | 419,064,063 | 417,797,716 | 62,859,609,450 | 62,371,872,080 | 99.70 | 7.64 | 20.79 | 97.47 |
| 0999934793 | BBJ | Kanto | 407,554,062 | 403,860,459 | 61,133,109,300 | 60,313,392,761 | 99.09 | 4.68 | 20.10 | 97.05 |
| 0999934803 | BBJ | Kanto | 531,022,081 | 528,488,652 | 79,653,312,150 | 78,947,272,155 | 99.52 | 6.65 | 26.32 | 99.09 |
| 0999934813 | BBJ | Kanto | 490,764,803 | 486,936,070 | 73,614,720,450 | 72,742,626,738 | 99.22 | 6.79 | 24.25 | 97.04 |
| 0999934917 | BBJ | Kanto | 411,137,563 | 408,213,061 | 61,670,634,450 | 60,944,490,081 | 99.29 | 7.17 | 20.31 | 96.68 |
| 0999934927 | BBJ | Kanto | 387,612,881 | 384,576,114 | 58,141,932,150 | 57,442,780,607 | 99.22 | 7.01 | 19.15 | 95.12 |
| 0999934947 | BBJ | Kanto | 503,182,065 | 500,815,035 | 75,477,309,750 | 74,758,483,479 | 99.53 | 7.29 | 24.92 | 97.16 |
| 0999934957 | BBJ | Kanto | 520,567,023 | 516,873,296 | 78,085,053,450 | 77,124,291,848 | 99.29 | 10.12 | 25.71 | 96.95 |
| 0999935031 | BBJ | Kanto | 365,952,463 | 364,646,176 | 54,892,869,450 | 54,409,982,459 | 99.64 | 6.45 | 18.14 | 91.31 |
| 0999935041 | BBJ | Kanto | 493,522,449 | 491,102,367 | 74,028,367,350 | 73,326,186,197 | 99.51 | 8.67 | 24.44 | 98.66 |
| 0999935051 | BBJ | Kanto | 427,487,372 | 425,965,127 | 64,123,105,800 | 63,572,214,066 | 99.64 | 7.81 | 21.19 | 94.92 |
| 0999935061 | BBJ | Kanto | 391,604,980 | 389,979,044 | 58,740,747,000 | 58,219,985,064 | 99.58 | 6.62 | 19.41 | 96.18 |
| 0999935071 | BBJ | Kanto | 440,765,796 | 437,064,349 | 66,114,869,400 | 65,247,813,491 | 99.16 | 7.39 | 21.75 | 95.25 |
| 0999935081 | BBJ | Kanto | 450,163,352 | 447,472,427 | 67,524,502,800 | 66,825,285,759 | 99.40 | 5.71 | 22.28 | 95.86 |
| 0999935407 | BBJ | Kanto | 436,646,292 | 434,661,016 | 65,496,943,800 | 64,839,647,041 | 99.55 | 8.84 | 21.61 | 97.53 |
| 0999935437 | BBJ | Kanto | 589,164,849 | 585,195,910 | 88,374,727,350 | 87,365,824,871 | 99.33 | 12.79 | 29.12 | 97.96 |
| 0999935447 | BBJ | Kanto | 459,241,665 | 456,648,994 | 68,886,249,750 | 68,159,471,471 | 99.44 | 7.37 | 22.72 | 95.99 |
| 0999935457 | BBJ | Kanto | 355,665,381 | 354,142,043 | 53,349,807,150 | 52,852,223,671 | 99.57 | 3.91 | 17.62 | 94.53 |
| 0999935467 | BBJ | Kanto | 517,581,064 | 515,436,281 | 77,637,159,600 | 76,978,379,090 | 99.59 | 7.29 | 25.66 | 99.02 |
| 0999935477 | BBJ | Kanto | 475,896,815 | 473,982,382 | 71,384,522,250 | 70,775,162,124 | 99.60 | 5.84 | 23.59 | 96.91 |

| | | | | | | | | | | |
|---|---|---|---|---|---|---|---|---|---|---|
| 0999935495 | BBJ | Kanto | 384,695,710 | 382,627,265 | 57,704,356,500 | 57,124,158,919 | 99.46 | 6.11 | 19.04 | 93.01 |
| 0999935505 | BBJ | Kanto | 472,266,146 | 469,961,768 | 70,839,921,900 | 70,174,567,685 | 99.51 | 8.40 | 23.39 | 98.42 |
| 0999936075 | BBJ | Kanto | 487,496,691 | 485,500,866 | 73,124,503,650 | 72,490,177,466 | 99.59 | 8.96 | 24.16 | 96.77 |
| 0999936085 | BBJ | Kanto | 447,967,698 | 445,045,182 | 67,195,154,700 | 66,448,217,077 | 99.35 | 5.20 | 22.15 | 95.93 |
| 0999936095 | BBJ | Kanto | 474,001,113 | 469,243,226 | 71,100,166,950 | 70,042,976,038 | 99.00 | 4.94 | 23.35 | 98.58 |
| 0999936105 | BBJ | Kanto | 346,869,486 | 345,616,299 | 52,030,422,900 | 51,594,683,742 | 99.64 | 6.99 | 17.20 | 89.30 |
| 0999936115 | BBJ | Kanto | 425,317,526 | 423,939,750 | 63,797,628,900 | 63,281,789,880 | 99.68 | 10.02 | 21.09 | 97.27 |
| 0999936135 | BBJ | Kanto | 375,367,153 | 371,320,134 | 56,305,072,950 | 55,457,373,405 | 98.92 | 4.46 | 18.49 | 95.11 |
| 0999936145 | BBJ | Kanto | 426,041,402 | 423,782,265 | 63,906,210,300 | 63,282,774,166 | 99.47 | 10.03 | 21.09 | 97.31 |
| 0999936155 | BBJ | Kanto | 398,312,006 | 396,237,653 | 59,746,800,900 | 59,143,920,466 | 99.48 | 5.13 | 19.71 | 94.20 |
| 0999936165 | BBJ | Kanto | 378,570,976 | 376,593,891 | 56,785,646,400 | 56,241,013,175 | 99.48 | 6.18 | 18.75 | 95.73 |
| 0999936175 | BBJ | Kanto | 439,221,514 | 435,486,068 | 65,883,227,100 | 65,030,263,071 | 99.15 | 6.46 | 21.68 | 98.04 |
| 1000063688 | BBJ | Kanto | 377,568,234 | 375,602,883 | 56,635,235,100 | 56,065,489,131 | 99.48 | 7.15 | 18.69 | 94.94 |
| 1000063698 | BBJ | Kanto | 411,494,259 | 409,778,376 | 61,724,138,850 | 61,191,268,287 | 99.58 | 8.22 | 20.40 | 94.24 |
| 1000063708 | BBJ | Kanto | 416,270,390 | 414,970,729 | 62,440,558,500 | 61,932,911,663 | 99.69 | 10.28 | 20.64 | 96.82 |
| 1000063718 | BBJ | Kanto | 480,067,523 | 477,037,958 | 72,010,128,450 | 71,260,766,334 | 99.37 | 6.92 | 23.75 | 98.74 |
| 1000063728 | BBJ | Kanto | 649,245,497 | 645,592,017 | 97,386,824,550 | 96,401,332,674 | 99.44 | 5.25 | 32.13 | 98.82 |
| 1000063738 | BBJ | Kanto | 378,115,903 | 376,244,428 | 56,717,385,450 | 56,188,967,781 | 99.51 | 5.81 | 18.73 | 95.43 |
| 1000063748 | BBJ | Kanto | 449,884,578 | 446,929,656 | 67,482,686,700 | 66,753,041,088 | 99.34 | 9.19 | 22.25 | 95.50 |
| 1000063768 | BBJ | Kanto | 420,470,399 | 418,812,809 | 63,070,559,850 | 62,549,611,222 | 99.61 | 7.66 | 20.85 | 97.28 |
| 1000063778 | BBJ | Kanto | 382,244,633 | 380,955,088 | 57,336,694,950 | 56,846,493,823 | 99.66 | 9.67 | 18.95 | 94.05 |
| 1000064906 | BBJ | Kanto | 404,790,041 | 403,075,183 | 60,718,506,150 | 60,180,021,165 | 99.58 | 7.07 | 20.06 | 96.55 |
| 1000064916 | BBJ | Kanto | 389,331,764 | 387,761,950 | 58,399,764,600 | 57,910,826,823 | 99.60 | 6.43 | 19.30 | 93.38 |
| 1000064926 | BBJ | Kanto | 398,999,832 | 397,049,312 | 59,849,974,800 | 59,291,493,474 | 99.51 | 6.74 | 19.76 | 93.80 |
| 1000064956 | BBJ | Kanto | 414,062,716 | 411,887,857 | 62,109,407,400 | 61,494,314,786 | 99.47 | 8.18 | 20.50 | 94.37 |
| 1000064966 | BBJ | Kanto | 434,943,813 | 433,218,305 | 65,241,571,950 | 64,705,482,340 | 99.60 | 5.35 | 21.57 | 98.13 |
| 1000064976 | BBJ | Kanto | 390,765,116 | 388,948,594 | 58,614,767,400 | 58,068,526,985 | 99.54 | 4.10 | 19.36 | 93.77 |
| 1000064986 | BBJ | Kanto | 433,641,722 | 431,960,545 | 65,046,258,300 | 64,489,828,283 | 99.61 | 4.84 | 21.50 | 98.01 |
| 1000064996 | BBJ | Kanto | 466,631,808 | 463,759,271 | 69,994,771,200 | 69,263,459,539 | 99.38 | 4.69 | 23.09 | 96.55 |
| 1000065016 | BBJ | Kanto | 504,700,512 | 500,397,725 | 75,705,076,800 | 74,702,411,596 | 99.15 | 7.57 | 24.90 | 98.82 |
| 1000065026 | BBJ | Kanto | 371,701,951 | 370,241,792 | 55,755,292,650 | 55,282,757,168 | 99.61 | 6.49 | 18.43 | 95.00 |
| 1000065460 | BBJ | Kanto | 444,296,377 | 442,653,167 | 66,644,456,550 | 66,096,494,771 | 99.63 | 5.70 | 22.03 | 96.16 |
| 1000065480 | BBJ | Kanto | 470,313,808 | 468,027,036 | 70,547,071,200 | 69,917,010,565 | 99.51 | 6.81 | 23.31 | 98.68 |
| 1000065500 | BBJ | Kanto | 384,928,789 | 383,056,377 | 57,739,318,350 | 57,204,343,171 | 99.51 | 5.44 | 19.07 | 92.97 |
| 1000065510 | BBJ | Kanto | 439,453,506 | 437,315,417 | 65,918,025,900 | 65,297,654,847 | 99.51 | 6.94 | 21.77 | 95.26 |
| 1000065520 | BBJ | Kanto | 467,731,591 | 464,052,400 | 70,159,738,650 | 69,281,034,293 | 99.21 | 6.46 | 23.09 | 98.36 |
| 1000065530 | BBJ | Kanto | 498,988,586 | 495,837,059 | 74,848,287,900 | 74,017,165,898 | 99.37 | 11.12 | 24.67 | 96.60 |
| 1000065550 | BBJ | Kanto | 470,881,636 | 469,026,564 | 70,632,245,400 | 69,965,833,540 | 99.61 | 9.27 | 23.32 | 98.38 |
| 1000065560 | BBJ | Kanto | 430,106,534 | 428,497,489 | 64,515,980,100 | 63,930,736,486 | 99.63 | 9.95 | 21.31 | 94.69 |
| 1000065570 | BBJ | Kanto | 407,918,457 | 406,263,284 | 61,187,768,550 | 60,638,957,098 | 99.59 | 10.20 | 20.21 | 93.25 |
| 1000065734 | BBJ | Kanto | 461,597,002 | 456,912,283 | 69,239,550,300 | 68,214,915,368 | 98.99 | 7.06 | 22.74 | 96.28 |
| 1000065744 | BBJ | Kanto | 490,479,941 | 488,445,591 | 73,571,991,150 | 72,962,752,105 | 99.59 | 7.44 | 24.32 | 97.06 |
| 1000065764 | BBJ | Kanto | 407,203,525 | 405,234,086 | 61,080,528,750 | 60,489,340,915 | 99.52 | 6.27 | 20.16 | 94.00 |
| 1000065774 | BBJ | Kanto | 451,469,758 | 449,967,839 | 67,720,463,700 | 67,151,864,643 | 99.67 | 6.16 | 22.38 | 96.04 |
| 1000065992 | BBJ | Kanto | 467,891,778 | 465,376,635 | 70,183,766,700 | 69,491,759,792 | 99.46 | 8.25 | 23.16 | 96.41 |
| 1000066002 | BBJ | Kanto | 425,426,952 | 423,550,123 | 63,814,042,800 | 63,252,099,309 | 99.56 | 4.79 | 21.08 | 98.03 |
| 1000066012 | BBJ | Kanto | 489,191,601 | 487,219,133 | 73,378,740,150 | 72,732,641,446 | 99.60 | 11.23 | 24.24 | 96.56 |
| 1000066022 | BBJ | Kanto | 519,558,863 | 517,387,842 | 77,933,829,450 | 77,258,998,179 | 99.58 | 7.02 | 25.75 | 97.54 |
| 1000066042 | BBJ | Kanto | 452,267,549 | 450,050,942 | 67,840,132,350 | 67,187,315,567 | 99.51 | 8.02 | 22.40 | 95.74 |
| 1000066062 | BBJ | Kanto | 425,751,999 | 423,828,893 | 63,862,799,850 | 63,282,004,298 | 99.55 | 4.54 | 21.09 | 97.96 |
| 1000066072 | BBJ | Kanto | 434,167,074 | 432,321,609 | 65,125,061,100 | 64,548,030,059 | 99.57 | 6.64 | 21.52 | 97.83 |
| 1000066082 | BBJ | Kanto | 461,391,411 | 458,429,587 | 69,208,711,650 | 68,449,716,078 | 99.36 | 6.58 | 22.82 | 98.24 |
| 1000066092 | BBJ | Kanto | 437,227,601 | 435,654,887 | 65,584,140,150 | 65,041,179,306 | 99.64 | 8.25 | 21.68 | 97.73 |
| 1000066102 | BBJ | Kanto | 480,264,886 | 478,298,212 | 72,039,732,900 | 71,435,826,564 | 99.59 | 6.95 | 23.81 | 98.70 |
| 1000066224 | BBJ | Kanto | 501,534,485 | 498,197,749 | 75,230,172,750 | 74,376,966,136 | 99.33 | 7.41 | 24.79 | 98.74 |
| 1000066562 | BBJ | Kanto | 502,014,157 | 499,157,152 | 75,302,123,550 | 74,535,290,223 | 99.43 | 6.22 | 24.85 | 98.92 |
| 1999934671 | BBJ | Kanto | 421,347,109 | 417,780,883 | 63,202,066,350 | 62,397,058,042 | 99.15 | 6.66 | 20.80 | 94.45 |
| 1999934681 | BBJ | Kanto | 383,100,878 | 380,149,552 | 57,465,131,700 | 56,754,558,856 | 99.23 | 6.84 | 18.92 | 95.32 |
| 1999934691 | BBJ | Kanto | 451,039,136 | 448,548,618 | 67,655,870,400 | 66,946,531,300 | 99.45 | 8.18 | 22.32 | 95.81 |
| 1999935231 | BBJ | Kanto | 449,244,200 | 446,382,984 | 67,386,630,000 | 66,608,042,722 | 99.36 | 7.29 | 22.20 | 98.06 |
| 1999935241 | BBJ | Kanto | 424,521,447 | 421,551,202 | 63,678,217,050 | 62,944,138,140 | 99.30 | 7.03 | 20.98 | 94.81 |
| 1999935251 | BBJ | Kanto | 437,366,157 | 433,553,810 | 65,604,923,550 | 64,743,692,594 | 99.13 | 8.48 | 21.58 | 97.41 |
| 1999935261 | BBJ | Kanto | 385,225,770 | 383,975,505 | 57,783,865,500 | 57,301,652,837 | 99.68 | 8.90 | 19.10 | 92.56 |
| 1999935271 | BBJ | Kanto | 555,017,057 | 552,657,140 | 83,252,558,550 | 82,517,216,348 | 99.57 | 21.49 | 27.51 | 98.66 |
| 1999935281 | BBJ | Kanto | 491,194,871 | 489,111,088 | 73,679,230,650 | 73,015,758,306 | 99.58 | 8.57 | 24.34 | 96.64 |
| 1999935295 | BBJ | Kanto | 497,976,475 | 494,544,421 | 74,696,471,250 | 73,833,592,230 | 99.31 | 9.08 | 24.61 | 96.77 |
| 1999935305 | BBJ | Kanto | 449,702,365 | 447,123,686 | 67,455,354,750 | 66,750,110,801 | 99.43 | 7.69 | 22.25 | 98.05 |
| 1999935315 | BBJ | Kanto | 532,244,389 | 529,093,063 | 79,836,658,350 | 78,993,525,083 | 99.41 | 11.85 | 26.33 | 97.29 |
| 1999935845 | BBJ | Kanto | 326,543,163 | 325,710,770 | 48,981,474,450 | 48,637,199,532 | 99.75 | 4.70 | 16.21 | 90.57 |
| 1999935855 | BBJ | Kanto | 462,481,300 | 459,263,405 | 69,372,195,000 | 68,574,465,061 | 99.30 | 6.48 | 22.86 | 96.14 |
| 1999935865 | BBJ | Kanto | 743,743,263 | 741,369,836 | 111,561,489,450 | 110,640,644,776 | 99.68 | 6.82 | 36.88 | 99.15 |
| 1999935875 | BBJ | Kanto | 492,892,489 | 490,961,678 | 73,933,873,350 | 73,298,671,299 | 99.61 | 4.75 | 24.43 | 98.72 |
| 1999935885 | BBJ | Kanto | 402,181,918 | 401,127,639 | 60,327,287,700 | 59,885,299,277 | 99.74 | 9.99 | 19.96 | 93.58 |
| 1999935895 | BBJ | Kanto | 506,348,562 | 504,208,094 | 75,952,284,300 | 75,275,631,944 | 99.58 | 15.80 | 25.09 | 96.22 |
| 1999935973 | BBJ | Kanto | 522,841,634 | 519,943,103 | 78,426,245,100 | 77,655,623,476 | 99.45 | 9.27 | 25.89 | 98.95 |
| 1999936047 | BBJ | Kanto | 416,900,699 | 414,355,891 | 62,535,104,850 | 61,880,874,392 | 99.39 | 7.16 | 20.63 | 97.09 |
| 1999936057 | BBJ | Kanto | 451,552,440 | 445,408,446 | 67,732,866,000 | 66,471,993,600 | 98.64 | 6.85 | 22.16 | 95.63 |
| 1999936067 | BBJ | Kanto | 466,011,227 | 462,965,890 | 69,901,684,050 | 69,084,705,812 | 99.35 | 7.09 | 23.03 | 96.28 |
| 1999936383 | BBJ | Kanto | 433,073,592 | 429,495,766 | 64,961,038,800 | 64,144,838,285 | 99.17 | 6.29 | 21.38 | 95.26 |
| 1999936393 | BBJ | Kanto | 488,132,781 | 484,728,514 | 73,219,917,150 | 72,397,351,654 | 99.30 | 9.37 | 24.13 | 96.59 |
| 1999936403 | BBJ | Kanto | 409,232,958 | 407,721,129 | 61,384,943,700 | 60,843,820,098 | 99.63 | 6.50 | 20.28 | 94.21 |
| 1999936413 | BBJ | Kanto | 459,805,737 | 456,766,027 | 68,970,860,550 | 68,226,026,625 | 99.34 | 6.91 | 22.74 | 98.11 |
| 1999936423 | BBJ | Kanto | 500,381,936 | 497,920,309 | 75,057,290,400 | 74,357,331,819 | 99.51 | 14.81 | 24.79 | 96.22 |
| 1999936433 | BBJ | Kanto | 532,741,276 | 530,296,133 | 79,911,191,400 | 79,190,061,757 | 99.54 | 13.09 | 26.40 | 98.88 |
| 1999936443 | BBJ | Kanto | 452,170,226 | 450,290,107 | 67,825,533,900 | 67,245,794,085 | 99.58 | 6.15 | 22.42 | 96.29 |
| 1999936453 | BBJ | Kanto | 451,866,023 | 445,107,291 | 67,779,903,450 | 66,479,809,438 | 98.50 | 5.87 | 22.16 | 98.35 |
| 1999936597 | BBJ | Kanto | 466,876,576 | 465,330,685 | 70,031,486,400 | 69,496,663,637 | 99.67 | 6.56 | 23.17 | 98.42 |
| 1999936607 | BBJ | Kanto | 426,645,144 | 425,496,942 | 63,996,771,600 | 63,496,299,832 | 99.73 | 5.04 | 21.17 | 95.21 |
| 1999936617 | BBJ | Kanto | 497,410,826 | 495,214,889 | 74,611,623,900 | 73,937,060,064 | 99.56 | 9.92 | 24.65 | 98.68 |
| 1999936619 | BBJ | Kanto | 486,027,548 | 484,672,914 | 72,904,132,200 | 72,376,998,022 | 99.72 | 4.57 | 24.13 | 98.77 |
| 1999936627 | BBJ | Kanto | 434,376,864 | 432,187,779 | 65,156,529,600 | 64,538,949,236 | 99.50 | 4.80 | 21.51 | 95.74 |
| 1999936629 | BBJ | Kanto | 454,709,558 | 452,147,666 | 68,206,433,700 | 67,520,686,455 | 99.44 | 6.36 | 22.51 | 96.15 |
| 1999936639 | BBJ | Kanto | 509,428,906 | 506,904,300 | 76,414,335,900 | 75,672,363,929 | 99.50 | 7.96 | 25.22 | 98.92 |
| 1999936647 | BBJ | Kanto | 469,353,088 | 467,108,826 | 70,402,963,200 | 69,724,992,447 | 99.52 | 4.65 | 23.24 | 96.84 |
| 1999936649 | BBJ | Kanto | 513,567,248 | 510,208,676 | 77,035,087,200 | 76,041,776,917 | 99.35 | 8.49 | 25.35 | 98.70 |
| 1999936657 | BBJ | Kanto | 532,200,722 | 529,568,622 | 79,830,108,300 | 79,048,104,272 | 99.51 | 11.73 | 26.35 | 97.34 |
| 1999936659 | BBJ | Kanto | 475,738,602 | 472,997,662 | 71,360,790,300 | 70,641,376,023 | 99.42 | 7.00 | 23.55 | 96.62 |
| 1999936667 | BBJ | Kanto | 438,492,232 | 436,624,047 | 65,773,834,800 | 65,193,467,275 | 99.57 | 5.56 | 21.73 | 96.02 |
| 1999936669 | BBJ | Kanto | 414,674,201 | 412,560,263 | 62,201,130,150 | 61,631,597,746 | 99.49 | 6.13 | 20.54 | 97.47 |
| 1999936677 | BBJ | Kanto | 467,153,496 | 462,264,148 | 70,073,024,400 | 69,033,424,633 | 98.95 | 7.33 | 23.01 | 98.54 |

| | | | | | | | | | | |
|---|---|---|---|---|---|---|---|---|---|---|
| 1999936679 | BBJ | Kanto | 371,285,753 | 369,645,933 | 55,692,862,950 | 55,174,151,575 | 99.56 | 5.56 | 18.39 | 95.04 |
| 1999936689 | BBJ | Kanto | 401,573,364 | 399,841,055 | 60,236,004,600 | 59,671,426,545 | 99.57 | 6.40 | 19.89 | 93.90 |
| 1999936697 | BBJ | Kanto | 483,280,394 | 480,500,891 | 72,492,059,100 | 71,769,731,579 | 99.42 | 5.53 | 23.92 | 98.84 |
| 1999936699 | BBJ | Kanto | 389,158,267 | 387,700,394 | 58,373,740,050 | 57,863,563,918 | 99.63 | 7.45 | 19.29 | 95.73 |
| 1999936707 | BBJ | Kanto | 436,715,467 | 434,700,113 | 65,507,320,050 | 64,907,508,008 | 99.54 | 6.14 | 21.64 | 95.68 |
| 1999936709 | BBJ | Kanto | 396,438,622 | 395,111,842 | 59,465,793,300 | 58,982,835,718 | 99.67 | 7.79 | 19.66 | 96.37 |
| 1999936719 | BBJ | Kanto | 439,098,992 | 434,071,695 | 65,864,848,800 | 64,828,725,400 | 98.86 | 5.64 | 21.61 | 97.74 |
| 1999936727 | BBJ | Kanto | 465,308,716 | 463,526,136 | 69,796,307,400 | 69,234,600,899 | 99.62 | 7.39 | 23.08 | 96.53 |
| 1999936729 | BBJ | Kanto | 519,720,934 | 508,036,283 | 77,958,140,100 | 75,903,177,021 | 97.75 | 7.66 | 25.30 | 98.93 |
| 1999936737 | BBJ | Kanto | 417,267,896 | 414,555,992 | 62,590,184,400 | 61,899,505,010 | 99.35 | 5.52 | 20.63 | 97.73 |
| 1999936749 | BBJ | Kanto | 511,368,990 | 508,551,979 | 76,705,348,500 | 75,941,583,997 | 99.45 | 8.05 | 25.31 | 97.33 |
| 1999936759 | BBJ | Kanto | 560,200,365 | 558,272,948 | 84,030,054,750 | 83,347,530,786 | 99.66 | 14.12 | 27.78 | 99.03 |
| 1999936769 | BBJ | Kanto | 374,541,653 | 372,968,679 | 56,181,247,950 | 55,709,117,387 | 99.58 | 6.23 | 18.57 | 95.12 |
| 2000063962 | BBJ | Kanto | 365,397,637 | 363,703,170 | 54,809,645,550 | 54,311,400,405 | 99.54 | 6.89 | 18.10 | 94.46 |
| 2000063972 | BBJ | Kanto | 365,031,778 | 363,766,175 | 54,754,766,700 | 54,316,725,642 | 99.65 | 6.60 | 18.11 | 91.73 |
| 2000063982 | BBJ | Kanto | 523,301,281 | 518,681,527 | 78,495,192,150 | 77,489,212,402 | 99.12 | 12.83 | 25.83 | 97.02 |
| 2000063992 | BBJ | Kanto | 502,124,379 | 499,265,200 | 75,318,656,850 | 74,556,592,527 | 99.43 | 10.66 | 24.85 | 96.88 |
| 2000064012 | BBJ | Kanto | 504,683,957 | 500,688,451 | 75,702,593,550 | 74,736,321,577 | 99.21 | 6.76 | 24.91 | 97.05 |
| 2000064022 | BBJ | Kanto | 449,067,942 | 445,347,423 | 67,360,191,300 | 66,501,807,286 | 99.17 | 3.96 | 22.17 | 96.11 |
| 2000064042 | BBJ | Kanto | 419,444,460 | 418,091,721 | 62,916,669,000 | 62,420,520,018 | 99.68 | 8.17 | 20.81 | 97.53 |
| 2000064196 | BBJ | Kanto | 545,469,169 | 542,050,976 | 81,820,375,350 | 80,931,678,095 | 99.37 | 9.93 | 26.98 | 99.08 |
| 2000064206 | BBJ | Kanto | 500,450,773 | 497,977,850 | 75,067,615,950 | 74,388,084,246 | 99.51 | 9.59 | 24.80 | 96.91 |
| 2000064220 | BBJ | Kanto | 449,556,491 | 447,223,994 | 67,433,473,650 | 66,793,587,193 | 99.48 | 8.59 | 22.26 | 95.88 |
| 2000064226 | BBJ | Kanto | 451,920,244 | 448,904,065 | 67,788,036,600 | 67,035,203,239 | 99.33 | 8.95 | 22.35 | 95.59 |
| 2000064230 | BBJ | Kanto | 505,544,680 | 502,609,661 | 75,831,702,000 | 75,072,925,037 | 99.42 | 7.62 | 25.02 | 98.94 |
| 2000064236 | BBJ | Kanto | 545,460,885 | 542,780,038 | 81,819,132,750 | 81,038,173,104 | 99.51 | 11.02 | 27.01 | 97.56 |
| 2000064246 | BBJ | Kanto | 418,967,601 | 417,018,695 | 62,845,140,150 | 62,256,513,686 | 99.53 | 6.20 | 20.75 | 97.37 |
| 2000064250 | BBJ | Kanto | 441,125,238 | 438,959,920 | 66,168,785,700 | 65,553,480,339 | 99.51 | 6.23 | 21.85 | 95.89 |
| 2000064256 | BBJ | Kanto | 466,395,718 | 462,244,449 | 69,959,357,700 | 68,919,496,018 | 99.11 | 7.83 | 22.97 | 95.77 |
| 2000064260 | BBJ | Kanto | 500,017,337 | 497,863,707 | 75,002,600,550 | 74,279,404,880 | 99.57 | 11.48 | 24.76 | 96.69 |
| 2000064266 | BBJ | Kanto | 458,239,482 | 454,908,571 | 68,735,922,300 | 67,903,481,209 | 99.27 | 8.99 | 22.63 | 95.73 |
| 2000064270 | BBJ | Kanto | 501,619,263 | 499,466,040 | 75,242,889,450 | 74,608,362,674 | 99.57 | 10.35 | 24.87 | 98.81 |
| 2000064280 | BBJ | Kanto | 516,459,065 | 512,941,814 | 77,468,859,750 | 76,605,692,716 | 99.32 | 13.69 | 25.54 | 96.79 |
| 2000064286 | BBJ | Kanto | 470,708,032 | 468,787,093 | 70,606,204,800 | 69,948,139,660 | 99.59 | 8.15 | 23.32 | 98.19 |
| 2000064290 | BBJ | Kanto | 443,771,298 | 433,742,784 | 66,565,694,700 | 64,764,209,461 | 97.74 | 6.05 | 21.59 | 95.05 |
| 2000064300 | BBJ | Kanto | 425,709,456 | 423,408,055 | 63,856,418,400 | 63,208,107,586 | 99.46 | 5.92 | 21.07 | 94.89 |
| 2000064310 | BBJ | Kanto | 403,381,505 | 401,524,261 | 60,507,225,750 | 59,961,961,061 | 99.54 | 6.32 | 19.99 | 96.86 |
| 2000064320 | BBJ | Kanto | 374,454,415 | 373,015,165 | 56,168,162,250 | 55,709,484,187 | 99.62 | 6.10 | 18.57 | 95.30 |
| 2000064464 | BBJ | Kanto | 467,343,806 | 465,817,228 | 70,101,570,900 | 69,547,650,873 | 99.67 | 8.60 | 23.18 | 96.39 |
| 2000064474 | BBJ | Kanto | 492,981,639 | 488,483,742 | 73,947,245,850 | 72,959,321,358 | 99.09 | 7.92 | 24.32 | 97.00 |
| 2000064484 | BBJ | Kanto | 354,307,131 | 352,913,254 | 53,146,069,650 | 52,706,688,488 | 99.61 | 6.74 | 17.57 | 93.51 |
| 2000064494 | BBJ | Kanto | 504,190,917 | 501,657,242 | 75,628,637,550 | 74,946,733,028 | 99.50 | 6.68 | 24.98 | 98.95 |
| 2000064504 | BBJ | Kanto | 446,039,308 | 442,241,436 | 66,905,896,200 | 66,047,092,145 | 99.15 | 8.00 | 22.02 | 95.38 |
| 2000064524 | BBJ | Kanto | 467,396,110 | 465,548,978 | 70,109,416,500 | 69,495,492,787 | 99.60 | 9.55 | 23.17 | 98.25 |
| 2000064534 | BBJ | Kanto | 391,458,618 | 389,903,683 | 58,718,792,700 | 58,209,508,577 | 99.60 | 9.49 | 19.40 | 95.63 |
| 2000064552 | BBJ | Kanto | 466,085,137 | 463,893,578 | 69,912,770,550 | 69,244,905,484 | 99.53 | 8.38 | 23.08 | 98.39 |
| 2000064800 | BBJ | Kanto | 531,146,571 | 528,051,553 | 79,671,985,650 | 78,846,422,673 | 99.42 | 8.40 | 26.28 | 97.46 |
| 2000064810 | BBJ | Kanto | 444,778,151 | 441,468,730 | 66,716,722,650 | 65,937,403,773 | 99.26 | 7.06 | 21.98 | 97.81 |
| 2000065628 | BBJ | Kanto | 461,382,981 | 459,572,758 | 69,207,447,150 | 68,624,354,232 | 99.61 | 8.54 | 22.87 | 96.27 |
| 2000065638 | BBJ | Kanto | 450,197,727 | 448,159,549 | 67,529,659,050 | 66,912,500,783 | 99.55 | 8.08 | 22.30 | 95.77 |
| 2000065648 | BBJ | Kanto | 491,493,004 | 489,458,833 | 73,723,950,600 | 73,105,186,927 | 99.59 | 5.71 | 24.37 | 98.91 |
| 2000065658 | BBJ | Kanto | 451,234,711 | 448,477,786 | 67,685,206,650 | 66,973,557,743 | 99.39 | 10.76 | 22.32 | 94.95 |
| 2000065668 | BBJ | Kanto | 361,738,088 | 360,255,329 | 54,260,713,200 | 53,784,389,678 | 99.59 | 6.74 | 17.93 | 94.00 |
| 2000065678 | BBJ | Kanto | 481,405,159 | 478,761,445 | 72,210,773,850 | 71,498,522,976 | 99.45 | 5.73 | 23.83 | 96.79 |
| 2000065698 | BBJ | Kanto | 448,551,202 | 446,066,874 | 67,282,680,300 | 66,519,513,564 | 99.45 | 5.84 | 22.17 | 98.31 |
| 2000066168 | BBJ | Kanto | 528,944,496 | 526,460,243 | 79,341,674,400 | 78,556,997,420 | 99.53 | 9.25 | 26.19 | 97.37 |
| 2999934281 | BBJ | Kanto | 447,465,011 | 444,722,125 | 67,119,751,650 | 66,387,651,296 | 99.39 | 5.87 | 22.13 | 98.21 |
| 2999934721 | BBJ | Kanto | 314,336,595 | 312,964,870 | 47,150,489,250 | 46,716,895,632 | 99.56 | 5.41 | 15.57 | 87.25 |
| 2999934741 | BBJ | Kanto | 391,109,810 | 387,538,598 | 58,666,471,500 | 57,884,505,849 | 99.09 | 6.27 | 19.29 | 92.78 |
| 2999934949 | BBJ | Kanto | 444,061,582 | 441,792,529 | 66,609,237,300 | 65,981,364,307 | 99.49 | 7.69 | 21.99 | 98.10 |
| 2999934959 | BBJ | Kanto | 485,577,908 | 481,754,029 | 72,836,686,200 | 71,913,230,582 | 99.21 | 7.89 | 23.97 | 98.62 |
| 2999935005 | BBJ | Kanto | 411,466,756 | 409,701,356 | 61,720,013,400 | 61,135,640,608 | 99.57 | 8.08 | 20.38 | 93.95 |
| 2999935015 | BBJ | Kanto | 448,854,336 | 447,092,730 | 67,328,150,400 | 66,715,811,757 | 99.61 | 8.26 | 22.24 | 95.45 |
| 2999935023 | BBJ | Kanto | 452,939,358 | 444,409,592 | 67,940,903,700 | 66,350,136,297 | 98.12 | 8.86 | 22.12 | 97.78 |
| 2999935035 | BBJ | Kanto | 462,649,020 | 457,938,919 | 69,397,353,000 | 68,332,422,069 | 98.98 | 5.05 | 22.78 | 96.19 |
| 2999935045 | BBJ | Kanto | 452,322,073 | 448,015,332 | 67,848,310,950 | 66,902,604,977 | 99.05 | 4.50 | 22.30 | 98.40 |
| 2999935055 | BBJ | Kanto | 405,596,282 | 404,037,495 | 60,839,442,300 | 60,305,748,252 | 99.62 | 6.84 | 20.10 | 94.13 |
| 2999935065 | BBJ | Kanto | 426,311,136 | 424,966,822 | 63,946,670,400 | 63,408,117,000 | 99.68 | 6.79 | 21.14 | 97.61 |
| 2999935075 | BBJ | Kanto | 435,159,438 | 433,244,998 | 65,273,915,700 | 64,679,197,186 | 99.56 | 5.03 | 21.56 | 95.62 |
| 2999935085 | BBJ | Kanto | 421,669,020 | 420,163,321 | 63,250,353,000 | 62,700,222,194 | 99.64 | 8.37 | 20.90 | 94.60 |
| 2999935095 | BBJ | Kanto | 402,104,166 | 398,965,517 | 60,315,624,900 | 59,567,373,296 | 99.22 | 5.44 | 19.86 | 96.34 |
| 2999935469 | BBJ | Kanto | 412,097,077 | 410,490,996 | 61,814,561,550 | 61,257,038,359 | 99.61 | 10.33 | 20.42 | 93.54 |
| 2999935479 | BBJ | Kanto | 415,004,877 | 413,408,206 | 62,250,731,550 | 61,707,812,796 | 99.62 | 6.73 | 20.57 | 97.07 |
| 2999935489 | BBJ | Kanto | 480,067,774 | 476,894,800 | 72,010,166,100 | 71,193,657,399 | 99.34 | 5.34 | 23.73 | 96.92 |
| 2999935499 | BBJ | Kanto | 461,445,710 | 458,577,642 | 69,216,856,500 | 68,461,815,250 | 99.38 | 5.99 | 22.82 | 98.50 |
| 2999935519 | BBJ | Kanto | 493,090,784 | 490,956,578 | 73,963,617,600 | 73,287,317,648 | 99.57 | 10.33 | 24.43 | 98.55 |
| 2999935529 | BBJ | Kanto | 470,668,097 | 467,506,291 | 70,600,214,550 | 69,812,568,780 | 99.33 | 5.19 | 23.27 | 96.64 |
| 2999935539 | BBJ | Kanto | 495,696,886 | 492,094,297 | 74,354,532,900 | 73,433,685,412 | 99.27 | 7.45 | 24.48 | 97.01 |
| 2999935549 | BBJ | Kanto | 433,945,671 | 431,638,820 | 65,091,850,650 | 64,471,988,880 | 99.47 | 6.46 | 21.49 | 95.61 |
| 2999935559 | BBJ | Kanto | 385,053,620 | 383,569,143 | 57,758,043,000 | 57,264,630,202 | 99.61 | 6.36 | 19.09 | 96.06 |
| 2999935569 | BBJ | Kanto | 325,389,164 | 323,994,030 | 48,808,374,600 | 48,385,017,838 | 99.57 | 3.75 | 16.13 | 87.87 |
| 2999935579 | BBJ | Kanto | 416,444,006 | 414,604,237 | 62,466,600,900 | 61,891,606,592 | 99.56 | 4.74 | 20.63 | 94.87 |
| 2999935589 | BBJ | Kanto | 509,302,160 | 506,499,596 | 76,395,324,000 | 75,643,474,998 | 99.45 | 6.50 | 25.21 | 97.47 |
| 2999935599 | BBJ | Kanto | 431,678,750 | 429,208,528 | 64,751,812,500 | 64,091,152,895 | 99.43 | 4.72 | 21.36 | 95.80 |
| 2999935609 | BBJ | Kanto | 500,920,353 | 498,473,394 | 75,138,052,950 | 74,458,345,515 | 99.51 | 5.96 | 24.82 | 98.76 |
| 2999935623 | BBJ | Kanto | 285,205,953 | 283,717,899 | 42,780,892,950 | 42,361,944,050 | 99.48 | 3.39 | 14.12 | 82.39 |
| 2999935633 | BBJ | Kanto | 489,895,929 | 487,176,264 | 73,484,389,350 | 72,687,443,826 | 99.44 | 9.78 | 24.23 | 96.45 |
| 2999935643 | BBJ | Kanto | 504,847,422 | 501,851,995 | 75,727,113,300 | 74,929,777,445 | 99.41 | 11.37 | 24.98 | 96.78 |
| 2999935653 | BBJ | Kanto | 482,023,738 | 478,766,246 | 72,303,560,700 | 71,478,241,659 | 99.32 | 7.33 | 23.83 | 96.56 |
| 2999935663 | BBJ | Kanto | 449,188,901 | 446,758,249 | 67,378,335,150 | 66,729,352,799 | 99.46 | 5.52 | 22.24 | 96.12 |
| 2999935673 | BBJ | Kanto | 468,062,950 | 464,758,179 | 70,209,442,500 | 69,407,041,700 | 99.29 | 6.28 | 23.14 | 98.34 |
| 2999935693 | BBJ | Kanto | 459,620,365 | 457,249,298 | 68,943,054,750 | 68,259,121,238 | 99.48 | 9.54 | 22.75 | 98.29 |
| 2999935703 | BBJ | Kanto | 517,330,051 | 514,903,196 | 77,599,507,650 | 76,915,639,908 | 99.53 | 6.30 | 25.64 | 97.63 |
| 2999935713 | BBJ | Kanto | 458,429,566 | 455,738,252 | 68,764,434,900 | 68,079,678,604 | 99.41 | 7.86 | 22.69 | 96.31 |
| 2999935723 | BBJ | Kanto | 348,550,205 | 347,161,801 | 52,282,530,750 | 51,835,474,178 | 99.60 | 6.07 | 17.28 | 89.94 |
| 2999935733 | BBJ | Kanto | 355,056,367 | 353,851,115 | 53,258,455,050 | 52,834,313,569 | 99.66 | 7.04 | 17.61 | 92.93 |
| 2999935743 | BBJ | Kanto | 410,608,567 | 408,375,803 | 61,591,285,050 | 60,970,369,038 | 99.46 | 4.81 | 20.32 | 97.62 |
| 2999935753 | BBJ | Kanto | 527,480,695 | 525,199,686 | 79,122,104,250 | 78,406,308,035 | 99.57 | 9.23 | 26.14 | 97.48 |
| 2999936407 | BBJ | Kanto | 511,976,393 | 507,092,847 | 76,796,458,950 | 75,694,488,863 | 99.05 | 9.25 | 25.23 | 97.09 |
| 2999936417 | BBJ | Kanto | 414,214,763 | 412,508,238 | 62,132,214,450 | 61,576,868,481 | 99.59 | 6.82 | 20.53 | 94.30 |

| | | | | | | | | | | |
|---|---|---|---|---|---|---|---|---|---|---|
| 2999936427 | BBJ | Kanto | 403,535,657 | 401,769,227 | 60,530,348,550 | 59,976,785,310 | 99.56 | 6.36 | 19.99 | 94.06 |
| 2999936437 | BBJ | Kanto | 373,902,310 | 372,607,355 | 56,085,346,500 | 55,663,196,047 | 99.65 | 4.36 | 18.55 | 96.04 |
| 2999936447 | BBJ | Kanto | 387,248,464 | 385,444,127 | 58,087,269,600 | 57,537,996,340 | 99.53 | 3.89 | 19.18 | 93.73 |
| 2999936457 | BBJ | Kanto | 368,247,821 | 366,739,222 | 55,237,173,150 | 54,698,365,894 | 99.59 | 4.44 | 18.23 | 95.00 |
| 2999936467 | BBJ | Kanto | 500,435,501 | 498,397,314 | 75,065,325,150 | 74,447,312,376 | 99.59 | 8.12 | 24.82 | 97.16 |
| 2999936477 | BBJ | Kanto | 500,514,645 | 498,577,821 | 75,077,196,750 | 74,449,754,370 | 99.61 | 8.53 | 24.82 | 97.15 |
| 2999936487 | BBJ | Kanto | 407,417,935 | 405,824,429 | 61,112,690,250 | 60,586,383,861 | 99.61 | 7.46 | 20.20 | 97.07 |
| 3000062912 | BBJ | Kanto | 497,607,746 | 495,127,412 | 74,641,161,900 | 73,911,670,195 | 99.50 | 7.68 | 24.64 | 97.10 |
| 3000062922 | BBJ | Kanto | 365,086,294 | 363,389,643 | 54,762,944,100 | 54,254,474,114 | 99.54 | 6.64 | 18.08 | 91.02 |
| 3000062932 | BBJ | Kanto | 440,571,320 | 437,627,511 | 66,085,698,000 | 65,352,979,614 | 99.33 | 6.41 | 21.78 | 95.52 |
| 3000062942 | BBJ | Kanto | 548,410,509 | 546,315,237 | 82,261,576,350 | 81,592,421,741 | 99.62 | 10.91 | 27.20 | 97.71 |
| 3000062952 | BBJ | Kanto | 460,847,540 | 458,955,759 | 69,127,131,000 | 68,476,358,434 | 99.59 | 7.11 | 22.83 | 96.01 |
| 3000062962 | BBJ | Kanto | 467,073,014 | 464,171,157 | 70,060,952,100 | 69,312,955,952 | 99.38 | 6.15 | 23.10 | 96.44 |
| 3000062972 | BBJ | Kanto | 488,932,788 | 485,897,435 | 73,339,918,200 | 72,582,864,084 | 99.38 | 8.83 | 24.19 | 98.67 |
| 3000062982 | BBJ | Kanto | 460,548,972 | 457,269,111 | 69,082,345,800 | 68,273,657,046 | 99.29 | 5.68 | 22.76 | 98.43 |
| 3000062992 | BBJ | Kanto | 459,425,859 | 456,290,400 | 68,913,878,850 | 68,144,318,260 | 99.32 | 10.55 | 22.71 | 95.70 |
| 3000063002 | BBJ | Kanto | 512,180,304 | 507,987,504 | 76,827,045,600 | 75,836,615,167 | 99.18 | 7.65 | 25.28 | 98.86 |
| 3000064054 | BBJ | Kanto | 431,252,973 | 429,851,882 | 64,687,945,950 | 64,224,672,925 | 99.68 | 8.28 | 21.41 | 97.95 |
| 3000064064 | BBJ | Kanto | 477,609,897 | 475,773,671 | 71,641,484,550 | 71,054,063,551 | 99.62 | 7.46 | 23.68 | 96.85 |
| 3000064068 | BBJ | Kanto | 403,216,554 | 399,809,423 | 60,482,483,100 | 59,703,560,060 | 99.16 | 5.51 | 19.90 | 93.70 |
| 3000064074 | BBJ | Kanto | 507,317,962 | 504,414,227 | 76,097,694,300 | 75,313,084,014 | 99.43 | 9.38 | 25.10 | 97.12 |
| 3000064078 | BBJ | Kanto | 453,919,187 | 451,872,245 | 68,087,878,050 | 67,502,055,470 | 99.55 | 6.29 | 22.50 | 98.53 |
| 3000064080 | BBJ | Kanto | 500,660,141 | 498,594,104 | 75,099,021,150 | 74,480,123,542 | 99.59 | 10.34 | 24.83 | 98.84 |
| 3000064084 | BBJ | Kanto | 505,202,431 | 503,079,756 | 75,780,364,650 | 75,102,781,839 | 99.58 | 8.92 | 25.03 | 98.86 |
| 3000064088 | BBJ | Kanto | 469,561,115 | 466,040,056 | 70,434,167,250 | 69,583,543,969 | 99.25 | 9.81 | 23.19 | 95.75 |
| 3000064090 | BBJ | Kanto | 570,513,210 | 567,801,755 | 85,576,981,500 | 84,738,769,561 | 99.52 | 9.24 | 28.25 | 97.96 |
| 3000064094 | BBJ | Kanto | 497,724,640 | 494,242,053 | 74,658,696,000 | 73,805,814,766 | 99.30 | 8.87 | 24.60 | 96.76 |
| 3000064098 | BBJ | Kanto | 491,347,241 | 488,718,191 | 73,702,086,150 | 72,982,518,107 | 99.46 | 5.71 | 24.33 | 98.83 |
| 3000064100 | BBJ | Kanto | 538,081,325 | 534,258,944 | 80,712,198,750 | 79,778,149,671 | 99.29 | 9.18 | 26.59 | 97.55 |
| 3000064104 | BBJ | Kanto | 551,331,453 | 548,337,306 | 82,699,717,950 | 81,889,855,664 | 99.46 | 16.48 | 27.30 | 98.85 |
| 3000064108 | BBJ | Kanto | 476,676,603 | 474,536,814 | 71,501,490,450 | 70,867,585,230 | 99.55 | 8.53 | 23.62 | 96.62 |
| 3000064110 | BBJ | Kanto | 454,205,299 | 450,842,645 | 68,130,794,850 | 67,320,094,220 | 99.26 | 5.77 | 22.44 | 95.98 |
| 3000064114 | BBJ | Kanto | 374,704,740 | 373,280,262 | 56,205,711,000 | 55,701,746,765 | 99.62 | 8.24 | 18.57 | 94.60 |
| 3000064120 | BBJ | Kanto | 463,888,735 | 460,667,743 | 69,583,310,250 | 68,770,840,355 | 99.31 | 6.15 | 22.92 | 96.31 |
| 3000064124 | BBJ | Kanto | 480,400,933 | 478,722,910 | 72,060,139,950 | 71,461,539,269 | 99.65 | 5.52 | 23.82 | 96.66 |
| 3000064128 | BBJ | Kanto | 424,352,629 | 422,617,063 | 63,652,894,350 | 63,092,387,624 | 99.59 | 8.43 | 21.03 | 97.44 |
| 3000064130 | BBJ | Kanto | 443,853,871 | 441,148,003 | 66,578,080,650 | 65,873,796,094 | 99.39 | 6.21 | 21.96 | 97.97 |
| 3000064134 | BBJ | Kanto | 433,588,016 | 431,872,922 | 65,038,202,400 | 64,485,847,143 | 99.60 | 6.81 | 21.50 | 95.33 |
| 3000064138 | BBJ | Kanto | 425,554,197 | 423,537,081 | 63,833,129,550 | 63,243,167,395 | 99.53 | 6.62 | 21.08 | 95.07 |
| 3000064144 | BBJ | Kanto | 444,291,124 | 441,780,088 | 66,643,668,600 | 66,000,299,237 | 99.43 | 10.05 | 22.00 | 97.71 |
| 3000064148 | BBJ | Kanto | 452,248,639 | 449,690,839 | 67,837,295,850 | 67,136,105,582 | 99.43 | 5.63 | 22.38 | 96.12 |
| 3000064154 | BBJ | Kanto | 434,749,184 | 430,530,148 | 65,212,377,600 | 64,257,882,889 | 99.03 | 7.86 | 21.42 | 95.21 |
| 3000064158 | BBJ | Kanto | 564,348,437 | 552,943,271 | 84,652,265,550 | 82,550,659,750 | 97.98 | 3.07 | 27.52 | 98.17 |
| 3000064164 | BBJ | Kanto | 448,903,530 | 445,408,496 | 67,335,529,500 | 66,497,004,530 | 99.22 | 9.04 | 22.17 | 95.28 |
| 3000064168 | BBJ | Kanto | 464,193,762 | 462,330,720 | 69,629,064,300 | 68,993,603,288 | 99.60 | 7.83 | 23.00 | 95.99 |
| 3000064174 | BBJ | Kanto | 403,112,940 | 401,573,628 | 60,466,941,000 | 59,904,047,426 | 99.62 | 7.65 | 19.97 | 95.53 |
| 3000064606 | BBJ | Kanto | 335,118,656 | 333,826,293 | 50,267,798,400 | 49,842,907,663 | 99.61 | 6.81 | 16.61 | 91.22 |
| 3000064616 | BBJ | Kanto | 417,020,940 | 415,436,615 | 62,553,141,000 | 62,022,954,842 | 99.62 | 7.25 | 20.67 | 94.76 |
| 3000064626 | BBJ | Kanto | 438,770,654 | 437,046,258 | 65,815,598,100 | 65,228,398,487 | 99.61 | 7.21 | 21.74 | 95.62 |
| 3000064636 | BBJ | Kanto | 421,343,129 | 419,792,149 | 63,201,469,350 | 62,654,702,595 | 99.63 | 4.62 | 20.88 | 95.27 |
| 3000064646 | BBJ | Kanto | 474,511,282 | 472,402,682 | 71,176,692,300 | 70,526,803,876 | 99.56 | 4.47 | 23.51 | 96.73 |
| 3000064656 | BBJ | Kanto | 458,574,591 | 456,524,115 | 68,786,188,650 | 68,020,748,372 | 99.55 | 7.76 | 22.67 | 79.20 |
| 3000064666 | BBJ | Kanto | 410,046,598 | 408,534,402 | 61,506,989,700 | 60,984,383,946 | 99.63 | 6.64 | 20.33 | 97.03 |
| 3000064676 | BBJ | Kanto | 468,156,385 | 465,280,684 | 70,223,457,750 | 69,457,609,856 | 99.39 | 8.26 | 23.15 | 98.50 |
| 3000064686 | BBJ | Kanto | 484,353,686 | 481,592,314 | 72,653,052,900 | 71,901,034,146 | 99.43 | 5.80 | 23.97 | 96.92 |
| 3000064696 | BBJ | Kanto | 423,263,843 | 420,781,714 | 63,489,576,450 | 62,849,555,369 | 99.41 | 4.77 | 20.95 | 97.56 |
| 3000064706 | BBJ | Kanto | 506,422,547 | 503,871,335 | 75,963,382,050 | 75,206,751,515 | 99.50 | 8.36 | 25.07 | 97.14 |
| 3000064716 | BBJ | Kanto | 457,527,612 | 454,016,256 | 68,629,141,800 | 67,776,245,776 | 99.23 | 6.94 | 22.59 | 98.18 |
| 3999934573 | BBJ | Kanto | 368,279,801 | 365,678,064 | 55,241,970,150 | 54,612,216,230 | 99.29 | 6.62 | 18.20 | 94.21 |
| 3999934811 | BBJ | Kanto | 390,436,723 | 389,105,443 | 58,565,508,450 | 58,092,326,189 | 99.66 | 5.93 | 19.36 | 96.32 |
| 3999934841 | BBJ | Kanto | 479,460,397 | 476,986,269 | 71,919,059,550 | 71,164,091,280 | 99.48 | 10.04 | 23.72 | 98.34 |
| 3999934851 | BBJ | Kanto | 487,658,368 | 485,113,043 | 73,148,755,200 | 72,427,340,876 | 99.48 | 9.10 | 24.14 | 96.57 |
| 3999934861 | BBJ | Kanto | 386,879,659 | 385,484,713 | 58,031,948,850 | 57,531,835,440 | 99.64 | 6.99 | 19.18 | 92.78 |
| 3999934871 | BBJ | Kanto | 284,110,240 | 282,966,128 | 42,616,536,000 | 42,246,620,595 | 99.60 | 4.22 | 14.08 | 81.47 |
| 3999934881 | BBJ | Kanto | 439,099,764 | 436,820,591 | 65,864,964,600 | 65,248,168,452 | 99.48 | 7.56 | 21.75 | 95.56 |
| 3999934891 | BBJ | Kanto | 464,940,455 | 459,051,958 | 69,741,068,250 | 68,562,024,303 | 98.73 | 5.32 | 22.85 | 96.34 |
| 3999934901 | BBJ | Kanto | 413,667,266 | 412,171,794 | 62,050,089,900 | 61,510,706,089 | 99.64 | 8.09 | 20.50 | 94.40 |
| 3999934911 | BBJ | Kanto | 373,334,160 | 371,457,617 | 56,000,124,000 | 55,437,598,095 | 99.50 | 6.91 | 18.48 | 94.74 |
| 3999934921 | BBJ | Kanto | 486,089,063 | 484,058,606 | 72,913,359,450 | 72,280,053,402 | 99.58 | 6.83 | 24.09 | 97.01 |
| 3999934931 | BBJ | Kanto | 432,111,578 | 429,083,927 | 64,816,736,700 | 64,073,660,549 | 99.30 | 7.16 | 21.36 | 95.43 |
| 3999935005 | BBJ | Kanto | 479,752,472 | 477,416,436 | 71,962,870,800 | 71,198,534,783 | 99.51 | 9.34 | 23.73 | 96.25 |
| 3999935015 | BBJ | Kanto | 461,500,654 | 458,726,490 | 69,225,098,100 | 68,506,645,983 | 99.40 | 5.48 | 22.84 | 98.45 |
| 3999935025 | BBJ | Kanto | 454,315,848 | 451,653,437 | 68,147,377,200 | 67,458,943,119 | 99.41 | 6.03 | 22.49 | 98.35 |
| 3999935035 | BBJ | Kanto | 437,219,884 | 435,400,088 | 65,582,982,600 | 65,003,112,372 | 99.58 | 6.94 | 21.67 | 95.41 |
| 3999935045 | BBJ | Kanto | 413,557,632 | 412,037,224 | 62,033,644,800 | 61,509,625,790 | 99.63 | 7.11 | 20.50 | 94.41 |
| 3999935055 | BBJ | Kanto | 511,677,404 | 509,123,871 | 76,751,610,600 | 76,006,451,817 | 99.50 | 8.15 | 25.34 | 97.33 |
| 3999935065 | BBJ | Kanto | 487,374,578 | 485,296,056 | 73,106,186,700 | 72,484,204,871 | 99.57 | 6.83 | 24.16 | 98.85 |
| 3999935075 | BBJ | Kanto | 492,094,389 | 489,806,623 | 73,814,158,350 | 73,127,109,451 | 99.54 | 11.44 | 24.38 | 96.62 |
| 3999935085 | BBJ | Kanto | 484,696,262 | 482,450,036 | 72,704,439,300 | 72,044,653,121 | 99.54 | 5.94 | 24.01 | 98.89 |
| 3999935173 | BBJ | Kanto | 420,636,947 | 419,116,098 | 63,095,542,050 | 62,583,188,235 | 99.64 | 8.44 | 20.86 | 97.35 |
| 3999935193 | BBJ | Kanto | 478,403,930 | 476,725,192 | 71,760,589,500 | 71,189,468,441 | 99.65 | 8.26 | 23.73 | 98.60 |
| 3999935203 | BBJ | Kanto | 417,757,793 | 416,082,276 | 62,663,668,950 | 62,109,229,146 | 99.60 | 8.01 | 20.70 | 94.61 |
| 3999936569 | BBJ | Kanto | 365,292,442 | 364,228,974 | 54,793,866,300 | 54,343,180,162 | 99.71 | 4.97 | 18.11 | 91.61 |
| 3999936579 | BBJ | Kanto | 425,331,102 | 422,520,195 | 63,799,665,300 | 63,068,372,122 | 99.34 | 6.91 | 21.02 | 94.57 |
| 3999936589 | BBJ | Kanto | 358,097,980 | 354,492,089 | 53,714,697,000 | 52,951,101,695 | 98.99 | 3.63 | 17.65 | 93.94 |
| 3999936599 | BBJ | Kanto | 455,395,154 | 452,278,537 | 68,309,273,100 | 67,522,879,848 | 99.32 | 7.38 | 22.51 | 98.26 |
| 3999936609 | BBJ | Kanto | 462,062,752 | 459,164,536 | 69,309,412,800 | 68,524,355,957 | 99.37 | 9.08 | 22.84 | 95.88 |
| 3999936619 | BBJ | Kanto | 375,516,537 | 374,027,899 | 56,327,480,550 | 55,831,528,935 | 99.60 | 5.28 | 18.61 | 92.42 |
| 3999936629 | BBJ | Kanto | 578,182,168 | 576,280,824 | 86,727,325,200 | 86,007,714,476 | 99.67 | 6.63 | 28.67 | 99.12 |
| 3999936639 | BBJ | Kanto | 380,474,196 | 378,140,812 | 57,071,129,400 | 56,498,597,447 | 99.39 | 5.59 | 18.83 | 92.41 |
| 3999936777 | BBJ | Kanto | 523,670,470 | 520,222,901 | 78,550,570,500 | 77,684,276,611 | 99.34 | 9.72 | 25.89 | 98.93 |
| 3999937217 | BBJ | Kanto | 386,637,723 | 385,507,215 | 57,995,658,450 | 57,559,970,928 | 99.71 | 5.77 | 19.19 | 96.40 |
| 3999937227 | BBJ | Kanto | 443,250,170 | 439,629,093 | 66,487,525,500 | 65,630,484,585 | 99.18 | 10.93 | 21.88 | 95.95 |
| 3999937237 | BBJ | Kanto | 492,404,766 | 489,976,265 | 73,860,714,900 | 73,202,628,759 | 99.51 | 9.51 | 24.40 | 96.80 |
| 3999937247 | BBJ | Kanto | 446,811,187 | 444,393,009 | 67,021,678,050 | 66,399,986,609 | 99.46 | 5.84 | 22.13 | 98.29 |
| 3999937257 | BBJ | Kanto | 380,050,149 | 378,520,684 | 57,007,522,350 | 56,522,174,923 | 99.60 | 6.62 | 18.84 | 92.48 |
| 3999937267 | BBJ | Kanto | 468,746,269 | 466,991,502 | 70,311,940,350 | 69,740,820,286 | 99.63 | 9.01 | 23.25 | 96.24 |
| 3999937277 | BBJ | Kanto | 502,351,987 | 500,351,038 | 75,352,798,050 | 74,736,025,792 | 99.60 | 7.64 | 24.91 | 97.13 |
| 3999937287 | BBJ | Kanto | 416,837,588 | 414,924,899 | 62,525,638,200 | 61,921,602,469 | 99.54 | 7.78 | 20.64 | 94.48 |
| 3999937297 | BBJ | Kanto | 446,993,893 | 445,002,151 | 67,049,083,950 | 66,419,379,368 | 99.55 | 7.73 | 22.14 | 95.79 |

| | | | | | | | | | | |
|---|---|---|---|---|---|---|---|---|---|---|
| 4000063876 | BBJ | Kanto | 398,874,062 | 397,389,045 | 59,831,109,300 | 59,332,890,810 | 99.63 | 8.85 | 19.78 | 96.28 |
| 4000063886 | BBJ | Kanto | 520,965,712 | 517,720,624 | 78,144,856,800 | 77,304,597,283 | 99.38 | 8.86 | 25.77 | 97.35 |
| 4000063896 | BBJ | Kanto | 518,555,127 | 515,657,189 | 77,783,269,050 | 76,913,749,430 | 99.44 | 11.51 | 25.64 | 98.72 |
| 4000063906 | BBJ | Kanto | 413,798,258 | 412,602,459 | 62,069,738,700 | 61,582,610,670 | 99.71 | 9.85 | 20.53 | 96.94 |
| 4000063926 | BBJ | Kanto | 424,352,071 | 423,020,804 | 63,652,810,650 | 63,116,659,229 | 99.69 | 5.92 | 21.04 | 97.62 |
| 4000063936 | BBJ | Kanto | 381,305,401 | 379,576,348 | 57,195,810,150 | 56,669,167,566 | 99.55 | 5.68 | 18.89 | 95.65 |
| 4000063946 | BBJ | Kanto | 416,821,152 | 415,204,175 | 62,523,172,800 | 61,996,894,405 | 99.61 | 6.59 | 20.67 | 94.73 |
| 4000063956 | BBJ | Kanto | 512,552,524 | 509,267,394 | 76,882,878,600 | 76,034,827,727 | 99.36 | 9.76 | 25.34 | 98.78 |
| 4000063966 | BBJ | Kanto | 440,921,001 | 438,566,848 | 66,138,150,150 | 65,477,831,084 | 99.47 | 4.57 | 21.83 | 96.06 |
| 4000063976 | BBJ | Kanto | 433,570,906 | 431,850,562 | 65,035,635,900 | 64,434,755,241 | 99.60 | 11.11 | 21.48 | 94.37 |
| 4000064240 | BBJ | Kanto | 449,713,532 | 447,656,702 | 67,457,029,800 | 66,842,399,658 | 99.54 | 5.54 | 22.28 | 98.49 |
| 4000064250 | BBJ | Kanto | 430,748,989 | 429,029,846 | 64,612,348,350 | 64,058,557,806 | 99.60 | 7.56 | 21.35 | 95.11 |
| 4000064260 | BBJ | Kanto | 493,623,940 | 491,671,837 | 74,043,591,000 | 73,422,637,851 | 99.60 | 9.59 | 24.47 | 96.82 |
| 4000064270 | BBJ | Kanto | 507,587,601 | 505,790,599 | 76,138,140,150 | 75,536,595,487 | 99.65 | 6.75 | 25.18 | 98.97 |
| 4000064280 | BBJ | Kanto | 482,293,891 | 479,061,581 | 72,344,083,650 | 71,530,910,162 | 99.33 | 13.33 | 23.84 | 98.08 |
| 4000064290 | BBJ | Kanto | 488,244,396 | 484,861,525 | 73,236,659,400 | 72,397,737,233 | 99.31 | 5.57 | 24.13 | 96.83 |
| 4000064300 | BBJ | Kanto | 477,573,306 | 475,444,234 | 71,635,995,900 | 71,021,425,706 | 99.55 | 8.83 | 23.67 | 96.43 |
| 4000064310 | BBJ | Kanto | 447,646,945 | 444,441,171 | 67,147,041,750 | 66,181,544,844 | 99.28 | 10.31 | 22.06 | 96.23 |
| 4000064320 | BBJ | Kanto | 392,652,245 | 391,008,674 | 58,897,836,750 | 58,374,502,902 | 99.58 | 5.99 | 19.46 | 93.49 |
| 4000064330 | BBJ | Kanto | 446,990,918 | 444,218,495 | 67,048,637,700 | 66,350,541,194 | 99.38 | 6.75 | 22.12 | 95.91 |
| 4000064340 | BBJ | Kanto | 454,900,410 | 452,268,920 | 68,235,061,500 | 67,554,399,035 | 99.42 | 7.21 | 22.52 | 98.32 |
| 4000064350 | BBJ | Kanto | 408,197,866 | 405,381,002 | 61,229,679,900 | 60,525,989,692 | 99.31 | 4.76 | 20.18 | 97.29 |
| 4000064360 | BBJ | Kanto | 396,163,158 | 391,643,833 | 59,424,473,700 | 58,447,193,668 | 98.86 | 4.00 | 19.48 | 96.78 |
| 4000064370 | BBJ | Kanto | 502,133,300 | 498,952,570 | 75,319,995,000 | 74,523,238,651 | 99.37 | 8.55 | 24.84 | 98.68 |
| 4000064380 | BBJ | Kanto | 454,358,078 | 450,676,752 | 68,153,711,700 | 67,275,397,842 | 99.19 | 6.42 | 22.43 | 96.04 |
| 4000064390 | BBJ | Kanto | 461,532,058 | 455,971,572 | 69,229,808,700 | 68,090,161,617 | 98.80 | 7.83 | 22.70 | 95.84 |
| 4000064400 | BBJ | Kanto | 436,195,779 | 434,833,664 | 65,429,366,850 | 64,912,394,299 | 99.69 | 7.43 | 21.64 | 97.66 |
| 4000064410 | BBJ | Kanto | 422,036,733 | 420,559,571 | 63,305,509,950 | 62,779,127,706 | 99.65 | 9.04 | 20.93 | 94.50 |
| 4000064420 | BBJ | Kanto | 411,382,816 | 409,878,369 | 61,707,422,400 | 61,177,385,500 | 99.63 | 7.31 | 20.39 | 97.21 |
| 4000064430 | BBJ | Kanto | 529,275,404 | 527,638,163 | 79,391,310,600 | 78,768,268,276 | 99.69 | 5.19 | 26.26 | 98.93 |
| 4000064516 | BBJ | Kanto | 486,991,955 | 483,905,920 | 73,048,793,250 | 72,248,411,874 | 99.37 | 8.49 | 24.08 | 98.62 |
| 4000064870 | BBJ | Kanto | 513,552,060 | 508,949,540 | 77,032,809,000 | 75,977,757,715 | 99.10 | 8.66 | 25.33 | 98.88 |
| 4000065288 | BBJ | Kanto | 505,729,648 | 503,144,657 | 75,859,447,200 | 75,137,270,617 | 99.49 | 10.86 | 25.05 | 96.93 |
| 4000065298 | BBJ | Kanto | 342,289,029 | 340,366,993 | 51,343,354,350 | 50,804,450,646 | 99.44 | 5.34 | 16.93 | 89.07 |
| 4000065308 | BBJ | Kanto | 449,766,075 | 448,164,509 | 67,464,911,250 | 66,884,773,803 | 99.64 | 7.56 | 22.29 | 98.03 |
| 4000065318 | BBJ | Kanto | 383,260,808 | 381,553,666 | 57,489,121,200 | 56,963,500,692 | 99.55 | 7.97 | 18.99 | 92.38 |
| 4000065328 | BBJ | Kanto | 390,866,191 | 388,149,179 | 58,629,928,650 | 57,984,743,646 | 99.30 | 4.55 | 19.33 | 93.97 |
| 4000065338 | BBJ | Kanto | 481,848,389 | 479,439,669 | 72,277,258,350 | 71,579,944,381 | 99.50 | 5.65 | 23.86 | 98.88 |
| 4000065348 | BBJ | Kanto | 463,424,901 | 461,184,651 | 69,513,735,150 | 68,863,525,738 | 99.52 | 4.91 | 22.95 | 96.75 |
| 4000065358 | BBJ | Kanto | 404,855,219 | 403,515,525 | 60,728,282,850 | 60,256,894,998 | 99.67 | 8.80 | 20.09 | 96.61 |
| 4000065368 | BBJ | Kanto | 395,051,874 | 393,397,471 | 59,257,781,100 | 58,731,656,690 | 99.58 | 7.20 | 19.58 | 96.20 |
| 4000065378 | BBJ | Kanto | 444,634,592 | 442,166,773 | 66,695,188,800 | 66,040,646,222 | 99.44 | 4.55 | 22.01 | 98.24 |
| 4000065388 | BBJ | Kanto | 486,033,058 | 483,520,371 | 72,904,958,700 | 72,200,080,124 | 99.48 | 9.36 | 24.07 | 98.48 |
| 4000065398 | BBJ | Kanto | 526,274,556 | 523,382,469 | 78,941,183,400 | 78,103,619,896 | 99.45 | 24.74 | 26.03 | 94.83 |
| 4000065862 | BBJ | Kanto | 420,599,629 | 418,161,276 | 63,089,944,350 | 62,469,782,724 | 99.42 | 6.17 | 20.82 | 97.43 |
| 4000065868 | BBJ | Kanto | 455,554,404 | 452,686,950 | 68,333,160,600 | 67,594,492,220 | 99.37 | 5.51 | 22.53 | 96.21 |
| 4000065872 | BBJ | Kanto | 406,150,748 | 403,545,863 | 60,922,612,200 | 60,285,174,074 | 99.36 | 5.15 | 20.10 | 94.49 |
| 4000065878 | BBJ | Kanto | 542,919,806 | 539,254,945 | 81,437,970,900 | 80,516,139,525 | 99.32 | 10.99 | 26.84 | 97.34 |
| 4000065882 | BBJ | Kanto | 496,554,406 | 494,156,894 | 74,483,160,900 | 73,806,200,470 | 99.52 | 9.14 | 24.60 | 96.99 |
| 4000065892 | BBJ | Kanto | 469,551,162 | 467,361,346 | 70,432,674,300 | 69,797,995,904 | 99.53 | 5.77 | 23.27 | 98.69 |
| 4000065902 | BBJ | Kanto | 518,819,132 | 516,308,107 | 77,822,869,800 | 77,124,720,827 | 99.52 | 17.75 | 25.71 | 98.47 |
| 4000065912 | BBJ | Kanto | 430,417,677 | 428,803,273 | 64,562,651,550 | 64,031,743,532 | 99.62 | 7.42 | 21.34 | 97.92 |
| 4000065922 | BBJ | Kanto | 423,742,206 | 419,296,641 | 63,561,330,900 | 62,614,613,289 | 98.95 | 7.87 | 20.87 | 94.22 |
| 4000065932 | BBJ | Kanto | 358,377,129 | 356,827,501 | 53,756,569,350 | 53,271,228,427 | 99.57 | 3.78 | 17.76 | 94.57 |
| 4000065942 | BBJ | Kanto | 399,680,425 | 397,959,494 | 59,952,063,750 | 59,409,439,218 | 99.57 | 7.30 | 19.80 | 93.60 |
| 4000065952 | BBJ | Kanto | 408,460,629 | 405,939,544 | 61,269,094,350 | 60,625,332,114 | 99.38 | 5.43 | 20.21 | 94.48 |
| 4000065962 | BBJ | Kanto | 422,466,469 | 419,736,495 | 63,369,970,350 | 62,684,206,729 | 99.35 | 8.33 | 20.89 | 94.62 |
| 4000065972 | BBJ | Kanto | 478,026,137 | 474,613,115 | 71,703,920,550 | 70,885,147,293 | 99.29 | 9.22 | 23.63 | 96.29 |
| 4000065982 | BBJ | Kanto | 455,181,343 | 452,444,306 | 68,277,201,450 | 67,569,871,981 | 99.40 | 6.40 | 22.52 | 98.32 |
| 4000065992 | BBJ | Kanto | 452,733,120 | 450,396,193 | 67,909,968,000 | 67,247,058,195 | 99.48 | 6.15 | 22.42 | 95.97 |
| 4000066002 | BBJ | Kanto | 469,458,960 | 466,917,499 | 70,418,844,000 | 69,716,206,637 | 99.46 | 11.53 | 23.24 | 95.83 |
| 4000066012 | BBJ | Kanto | 400,678,106 | 397,499,393 | 60,101,715,900 | 59,358,776,034 | 99.21 | 4.84 | 19.79 | 93.85 |
| 4000066372 | BBJ | Kanto | 437,395,456 | 434,416,635 | 65,609,318,400 | 64,910,224,033 | 99.32 | 6.18 | 21.64 | 98.14 |
| 4000066744 | BBJ | Kanto | 473,823,212 | 471,750,145 | 71,073,481,800 | 70,458,970,537 | 99.56 | 8.88 | 23.49 | 96.51 |
| 4000066754 | BBJ | Kanto | 369,023,766 | 367,219,017 | 55,353,564,900 | 54,799,800,538 | 99.51 | 6.51 | 18.27 | 91.60 |
| 4000066764 | BBJ | Kanto | 499,435,784 | 496,949,770 | 74,915,367,600 | 74,220,723,235 | 99.50 | 14.60 | 24.74 | 98.48 |
| 4000066774 | BBJ | Kanto | 466,067,149 | 464,667,639 | 69,910,072,350 | 69,340,708,489 | 99.70 | 8.80 | 23.11 | 96.28 |
| 4000066784 | BBJ | Kanto | 352,729,933 | 351,264,853 | 52,909,489,950 | 52,439,706,592 | 99.58 | 3.63 | 17.48 | 94.14 |
| 4000066794 | BBJ | Kanto | 367,335,545 | 365,613,778 | 55,100,331,750 | 54,595,402,725 | 99.53 | 6.37 | 18.20 | 94.63 |
| 4000066804 | BBJ | Kanto | 485,099,653 | 482,558,268 | 72,764,947,950 | 72,076,662,924 | 99.48 | 9.30 | 24.03 | 96.64 |
| 4000066814 | BBJ | Kanto | 469,464,484 | 466,610,990 | 70,419,672,600 | 69,713,463,096 | 99.39 | 8.64 | 23.24 | 98.34 |
| 4000066824 | BBJ | Kanto | 439,689,015 | 436,608,248 | 65,953,352,250 | 65,186,600,550 | 99.30 | 4.82 | 21.73 | 96.04 |
| 4000066834 | BBJ | Kanto | 380,661,784 | 379,513,742 | 57,099,267,600 | 56,647,334,086 | 99.70 | 4.91 | 18.88 | 93.07 |
| 4000067374 | BBJ | Kanto | 438,137,329 | 435,173,255 | 65,720,599,350 | 64,979,413,860 | 99.32 | 7.52 | 21.66 | 97.61 |
| 4999934013 | BBJ | Kanto | 448,189,038 | 445,619,599 | 67,228,355,700 | 66,549,424,198 | 99.43 | 6.26 | 22.18 | 98.36 |
| 4999934593 | BBJ | Kanto | 429,825,781 | 428,386,027 | 64,473,867,150 | 63,971,637,532 | 99.67 | 8.19 | 21.32 | 97.74 |
| 4999934603 | BBJ | Kanto | 464,964,612 | 462,221,664 | 69,744,691,800 | 69,025,380,858 | 99.41 | 5.60 | 23.01 | 96.45 |
| 4999934613 | BBJ | Kanto | 365,738,282 | 364,250,181 | 54,860,742,300 | 54,356,011,573 | 99.59 | 4.38 | 18.12 | 95.09 |
| 4999934623 | BBJ | Kanto | 405,892,992 | 404,289,636 | 60,883,948,800 | 60,339,773,796 | 99.60 | 4.37 | 20.11 | 97.20 |
| 4999934643 | BBJ | Kanto | 475,432,237 | 473,607,979 | 71,314,835,550 | 70,740,379,422 | 99.62 | 7.12 | 23.58 | 96.80 |
| 4999934653 | BBJ | Kanto | 457,843,301 | 455,824,707 | 68,676,495,150 | 68,074,817,341 | 99.56 | 5.22 | 22.69 | 98.58 |
| 4999934663 | BBJ | Kanto | 484,740,562 | 482,123,393 | 72,711,084,300 | 72,009,846,617 | 99.46 | 5.66 | 24.00 | 96.99 |
| 4999934673 | BBJ | Kanto | 468,197,698 | 466,330,549 | 70,229,654,700 | 69,665,862,286 | 99.60 | 7.18 | 23.22 | 98.65 |
| 4999934683 | BBJ | Kanto | 467,241,041 | 464,703,443 | 70,086,156,150 | 69,417,244,950 | 99.46 | 8.86 | 23.14 | 98.42 |
| 4999934775 | BBJ | Kanto | 413,474,787 | 410,944,591 | 62,021,218,050 | 61,363,776,116 | 99.39 | 6.95 | 20.45 | 97.12 |
| 4999934785 | BBJ | Kanto | 406,203,372 | 403,555,685 | 60,930,505,800 | 60,255,170,664 | 99.35 | 5.22 | 20.09 | 97.24 |
| 4999934811 | BBJ | Kanto | 435,800,361 | 434,207,643 | 65,370,054,150 | 64,811,690,655 | 99.63 | 9.75 | 21.60 | 94.95 |
| 4999934821 | BBJ | Kanto | 475,219,100 | 472,921,135 | 71,282,865,000 | 70,620,617,943 | 99.52 | 12.52 | 23.54 | 95.97 |
| 4999934831 | BBJ | Kanto | 474,426,925 | 472,419,533 | 71,164,038,750 | 70,574,159,568 | 99.58 | 9.26 | 23.52 | 98.58 |
| 4999934841 | BBJ | Kanto | 375,019,206 | 372,381,865 | 56,252,880,900 | 55,585,664,900 | 99.30 | 5.36 | 18.53 | 91.96 |
| 4999934851 | BBJ | Kanto | 496,645,336 | 495,004,511 | 74,496,800,400 | 73,939,333,843 | 99.67 | 9.60 | 24.65 | 98.76 |
| 4999934861 | BBJ | Kanto | 460,573,921 | 458,604,874 | 69,086,088,150 | 68,494,780,984 | 99.57 | 7.99 | 22.83 | 98.43 |
| 4999935289 | BBJ | Kanto | 382,126,064 | 380,495,270 | 57,318,909,600 | 56,787,409,050 | 99.57 | 9.92 | 18.93 | 91.59 |
| 4999935299 | BBJ | Kanto | 392,216,685 | 390,445,582 | 58,832,502,750 | 58,286,332,358 | 99.55 | 7.79 | 19.43 | 95.93 |
| 4999935305 | BBJ | Kanto | 402,745,787 | 400,831,153 | 60,411,868,050 | 59,822,662,927 | 99.52 | 3.78 | 19.94 | 94.26 |
| 4999935309 | BBJ | Kanto | 462,517,223 | 460,380,350 | 69,377,583,450 | 68,712,797,767 | 99.54 | 10.24 | 22.90 | 95.75 |
| 4999935319 | BBJ | Kanto | 442,939,422 | 439,100,252 | 66,440,913,300 | 65,555,622,059 | 99.13 | 5.06 | 21.85 | 95.91 |
| 4999935325 | BBJ | Kanto | 503,895,062 | 501,887,031 | 75,584,259,300 | 74,939,330,146 | 99.60 | 6.00 | 24.98 | 97.46 |
| 4999935329 | BBJ | Kanto | 439,915,904 | 436,858,977 | 65,987,385,600 | 65,245,688,282 | 99.31 | 5.56 | 21.75 | 95.66 |
| 4999935335 | BBJ | Kanto | 467,321,181 | 465,598,939 | 70,098,177,150 | 69,526,999,420 | 99.63 | 11.33 | 23.18 | 98.32 |

| | | | | | | | | | | |
|---|---|---|---|---|---|---|---|---|---|---|
| 4999935339 | BBJ | Kanto | 507,216,093 | 505,133,762 | 76,082,413,950 | 75,476,889,301 | 99.59 | 13.83 | 25.16 | 96.52 |
| 4999935345 | BBJ | Kanto | 400,467,187 | 397,283,104 | 60,070,078,050 | 59,351,252,495 | 99.20 | 5.96 | 19.78 | 93.64 |
| 4999935349 | BBJ | Kanto | 445,423,595 | 443,572,099 | 66,813,539,250 | 66,223,739,295 | 99.58 | 7.26 | 22.07 | 95.96 |
| 4999935355 | BBJ | Kanto | 406,778,876 | 404,560,848 | 61,016,831,400 | 60,447,610,939 | 99.45 | 5.48 | 20.15 | 94.78 |
| 4999935365 | BBJ | Kanto | 466,220,937 | 462,770,368 | 69,933,140,550 | 69,098,480,646 | 99.26 | 6.78 | 23.03 | 96.54 |
| 4999935369 | BBJ | Kanto | 384,095,859 | 381,410,010 | 57,614,378,850 | 56,942,808,573 | 99.30 | 5.77 | 18.98 | 92.55 |
| 4999935375 | BBJ | Kanto | 462,026,119 | 459,870,429 | 69,303,917,850 | 68,668,880,658 | 99.53 | 6.97 | 22.89 | 96.44 |
| 4999935379 | BBJ | Kanto | 434,827,211 | 432,226,419 | 65,224,081,650 | 64,546,325,842 | 99.40 | 4.50 | 21.52 | 96.07 |
| 4999936497 | BBJ | Kanto | 487,015,540 | 482,064,930 | 73,052,331,000 | 71,964,591,204 | 98.98 | 9.61 | 23.99 | 96.45 |
| 4999936507 | BBJ | Kanto | 473,112,273 | 470,395,657 | 70,966,840,950 | 70,239,238,898 | 99.43 | 6.61 | 23.41 | 96.54 |
| 4999936967 | BBJ | Kanto | 438,174,269 | 434,706,195 | 65,726,140,350 | 64,910,520,305 | 99.21 | 5.45 | 21.64 | 97.64 |
| 4999936977 | BBJ | Kanto | 456,845,169 | 453,937,111 | 68,526,775,350 | 67,775,504,354 | 99.36 | 8.41 | 22.59 | 98.12 |
| 4999936987 | BBJ | Kanto | 433,801,298 | 431,864,129 | 65,070,194,700 | 64,483,842,923 | 99.55 | 6.51 | 21.49 | 95.38 |
| 4999936997 | BBJ | Kanto | 399,920,998 | 398,345,295 | 59,988,149,700 | 59,432,921,818 | 99.61 | 5.86 | 19.81 | 93.82 |
| 4999937007 | BBJ | Kanto | 444,607,758 | 442,636,388 | 66,691,163,700 | 66,067,872,483 | 99.56 | 4.07 | 22.02 | 98.21 |
| 4999937017 | BBJ | Kanto | 422,821,053 | 420,798,636 | 63,423,157,950 | 62,813,258,387 | 99.52 | 4.39 | 20.94 | 95.11 |
| 4999937027 | BBJ | Kanto | 263,792,613 | 262,464,184 | 39,568,891,950 | 39,191,294,210 | 99.50 | 4.30 | 13.06 | 72.54 |
| 4999937037 | BBJ | Kanto | 455,446,985 | 452,071,536 | 68,317,047,750 | 67,506,323,251 | 99.26 | 7.83 | 22.50 | 98.01 |
| 4999937047 | BBJ | Kanto | 422,301,057 | 418,415,163 | 63,345,158,550 | 62,498,527,753 | 99.08 | 5.29 | 20.83 | 97.47 |
| 4999937057 | BBJ | Kanto | 458,011,315 | 455,591,981 | 68,701,697,250 | 68,037,675,673 | 99.47 | 8.16 | 22.68 | 96.14 |
| 4999937067 | BBJ | Kanto | 474,013,240 | 472,398,068 | 71,101,986,000 | 70,509,453,910 | 99.66 | 8.92 | 23.50 | 96.45 |
| 5000063436 | BBJ | Kanto | 588,203,578 | 584,140,935 | 88,230,536,700 | 87,213,113,543 | 99.31 | 9.72 | 29.07 | 98.09 |
| 5000063446 | BBJ | Kanto | 388,813,837 | 387,363,577 | 58,322,075,550 | 57,791,767,415 | 99.63 | 7.03 | 19.26 | 95.94 |
| 5000063456 | BBJ | Kanto | 410,042,260 | 408,655,240 | 61,506,339,000 | 60,976,726,729 | 99.66 | 5.17 | 20.33 | 97.25 |
| 5000063466 | BBJ | Kanto | 446,390,149 | 444,194,228 | 66,958,522,350 | 66,329,482,829 | 99.51 | 7.28 | 22.11 | 95.71 |
| 5000063476 | BBJ | Kanto | 443,250,600 | 441,322,541 | 66,487,590,000 | 65,876,171,626 | 99.57 | 9.68 | 21.96 | 95.32 |
| 5000063486 | BBJ | Kanto | 480,463,127 | 476,045,351 | 72,069,469,050 | 71,049,300,702 | 99.08 | 7.58 | 23.68 | 96.56 |
| 5000063496 | BBJ | Kanto | 482,343,605 | 480,127,830 | 72,351,540,750 | 71,622,203,386 | 99.54 | 5.26 | 23.87 | 96.89 |
| 5000064002 | BBJ | Kanto | 466,658,465 | 463,006,499 | 69,998,769,750 | 69,104,783,027 | 99.22 | 6.96 | 23.03 | 96.45 |
| 5000064012 | BBJ | Kanto | 402,797,583 | 401,160,605 | 60,419,637,450 | 59,880,535,749 | 99.59 | 7.27 | 19.96 | 94.02 |
| 5000064016 | BBJ | Kanto | 476,623,731 | 473,636,391 | 71,493,559,650 | 70,720,437,911 | 99.37 | 11.25 | 23.57 | 96.10 |
| 5000064026 | BBJ | Kanto | 480,969,839 | 476,396,863 | 72,145,475,850 | 71,111,805,053 | 99.05 | 7.90 | 23.70 | 98.51 |
| 5000064032 | BBJ | Kanto | 508,667,126 | 506,750,822 | 76,300,068,900 | 75,691,429,087 | 99.62 | 8.34 | 25.23 | 98.94 |
| 5000064036 | BBJ | Kanto | 443,803,118 | 441,258,181 | 66,570,467,700 | 65,877,986,869 | 99.43 | 7.16 | 21.96 | 97.93 |
| 5000064062 | BBJ | Kanto | 482,754,090 | 480,097,439 | 72,413,113,500 | 71,709,889,297 | 99.45 | 9.84 | 23.90 | 96.55 |
| 5000064072 | BBJ | Kanto | 476,776,534 | 472,593,261 | 71,516,480,100 | 70,571,892,968 | 99.12 | 5.25 | 23.52 | 96.54 |
| 5000064082 | BBJ | Kanto | 488,443,770 | 485,338,120 | 73,266,565,500 | 72,435,878,013 | 99.36 | 6.68 | 24.15 | 98.73 |
| 5000064092 | BBJ | Kanto | 478,398,207 | 476,082,383 | 71,759,731,050 | 71,062,673,478 | 99.52 | 6.18 | 23.69 | 98.57 |
| 5000064122 | BBJ | Kanto | 375,082,167 | 373,813,941 | 56,262,325,050 | 55,787,886,319 | 99.66 | 6.49 | 18.60 | 92.28 |
| 5000064524 | BBJ | Kanto | 495,717,611 | 490,660,616 | 74,357,641,650 | 73,278,356,946 | 98.98 | 5.78 | 24.43 | 97.24 |
| 5000064534 | BBJ | Kanto | 493,945,224 | 492,217,935 | 74,091,783,600 | 73,487,921,854 | 99.65 | 7.88 | 24.50 | 97.06 |
| 5000064544 | BBJ | Kanto | 446,528,849 | 444,660,966 | 66,979,327,350 | 66,423,421,006 | 99.58 | 5.68 | 22.14 | 98.46 |
| 5000064554 | BBJ | Kanto | 518,481,611 | 516,299,477 | 77,772,241,650 | 77,127,027,164 | 99.58 | 7.92 | 25.71 | 98.97 |
| 5000064564 | BBJ | Kanto | 404,178,731 | 402,413,482 | 60,626,809,650 | 60,080,148,842 | 99.56 | 9.21 | 20.03 | 96.23 |
| 5000064584 | BBJ | Kanto | 457,093,332 | 454,705,698 | 68,563,999,800 | 67,892,062,557 | 99.48 | 6.79 | 22.63 | 96.10 |
| 5000064594 | BBJ | Kanto | 499,796,933 | 498,088,799 | 74,969,539,950 | 74,398,635,581 | 99.66 | 7.91 | 24.80 | 97.15 |
| 5000064604 | BBJ | Kanto | 429,824,633 | 427,290,703 | 64,473,694,950 | 63,751,747,026 | 99.41 | 6.12 | 21.25 | 95.18 |
| 5000064612 | BBJ | Kanto | 445,977,899 | 440,777,386 | 66,896,684,850 | 65,793,234,802 | 98.83 | 7.11 | 21.93 | 95.50 |
| 5000064614 | BBJ | Kanto | 399,253,131 | 395,784,342 | 59,887,969,650 | 59,088,486,930 | 99.13 | 5.07 | 19.70 | 93.93 |
| 5000064620 | BBJ | Kanto | 367,186,641 | 365,044,047 | 55,077,996,150 | 54,492,964,833 | 99.42 | 5.51 | 18.16 | 91.51 |
| 5000064630 | BBJ | Kanto | 413,599,176 | 411,388,450 | 62,039,876,400 | 61,435,867,150 | 99.47 | 6.98 | 20.48 | 94.41 |
| 5000064634 | BBJ | Kanto | 337,663,367 | 336,603,745 | 50,649,505,050 | 50,250,307,194 | 99.69 | 5.08 | 16.75 | 88.63 |
| 5000064640 | BBJ | Kanto | 492,545,521 | 490,712,320 | 73,881,828,150 | 73,268,140,047 | 99.63 | 6.87 | 24.42 | 97.15 |
| 5000064650 | BBJ | Kanto | 466,495,518 | 464,786,715 | 69,974,327,700 | 69,404,680,552 | 99.63 | 6.21 | 23.13 | 98.72 |
| 5000064660 | BBJ | Kanto | 508,225,752 | 504,657,629 | 76,233,862,800 | 75,331,356,732 | 99.30 | 8.62 | 25.11 | 98.88 |
| 5000064670 | BBJ | Kanto | 407,681,963 | 406,467,085 | 61,152,294,450 | 60,678,596,683 | 99.70 | 9.70 | 20.23 | 93.83 |
| 5000064680 | BBJ | Kanto | 424,967,846 | 422,902,182 | 63,745,176,900 | 63,135,559,340 | 99.51 | 4.66 | 21.05 | 95.22 |
| 5000064690 | BBJ | Kanto | 428,962,311 | 427,400,388 | 64,344,346,650 | 63,814,800,185 | 99.64 | 5.67 | 21.27 | 95.30 |
| 5000064700 | BBJ | Kanto | 477,674,465 | 474,275,405 | 71,651,169,750 | 70,810,354,102 | 99.29 | 7.19 | 23.60 | 96.65 |
| 5000064710 | BBJ | Kanto | 471,438,926 | 466,868,926 | 70,715,838,900 | 69,620,461,962 | 99.03 | 3.91 | 23.21 | 98.66 |
| 5000065140 | BBJ | Kanto | 476,617,730 | 474,481,871 | 71,492,659,500 | 70,819,383,505 | 99.55 | 7.63 | 23.61 | 98.59 |
| 5000065150 | BBJ | Kanto | 464,314,089 | 459,006,133 | 69,647,113,350 | 68,520,333,658 | 98.86 | 6.84 | 22.84 | 96.25 |
| 5000065160 | BBJ | Kanto | 500,306,757 | 497,108,033 | 75,046,013,550 | 74,209,124,433 | 99.36 | 7.42 | 24.74 | 97.10 |
| 5000065170 | BBJ | Kanto | 472,238,705 | 469,672,261 | 70,835,805,750 | 70,114,664,090 | 99.46 | 8.85 | 23.37 | 96.26 |
| 5000065174 | BBJ | Kanto | 510,806,320 | 506,270,569 | 76,620,948,000 | 75,602,001,670 | 99.11 | 10.16 | 25.20 | 98.82 |
| 5000065184 | BBJ | Kanto | 482,166,303 | 477,991,782 | 72,324,945,450 | 71,315,120,222 | 99.13 | 7.37 | 23.77 | 96.30 |
| 5000065188 | BBJ | Kanto | 463,789,512 | 460,488,182 | 69,568,426,800 | 68,732,855,739 | 99.29 | 7.30 | 22.91 | 98.57 |
| 5000065194 | BBJ | Kanto | 477,942,714 | 472,657,290 | 71,691,407,100 | 70,557,624,383 | 98.89 | 6.32 | 23.52 | 98.56 |
| 5000065198 | BBJ | Kanto | 523,671,379 | 521,484,749 | 78,550,706,850 | 77,892,282,682 | 99.58 | 7.40 | 25.96 | 99.03 |
| 5000065208 | BBJ | Kanto | 504,752,851 | 501,489,025 | 75,712,927,650 | 74,831,994,806 | 99.35 | 9.97 | 24.94 | 96.94 |
| 5000065218 | BBJ | Kanto | 399,169,578 | 397,535,182 | 59,875,436,700 | 59,344,885,376 | 99.59 | 7.98 | 19.78 | 93.49 |
| 5000065228 | BBJ | Kanto | 381,088,962 | 379,824,440 | 57,163,344,300 | 56,649,344,292 | 99.67 | 6.13 | 18.88 | 92.25 |
| 5000065238 | BBJ | Kanto | 438,525,394 | 435,706,086 | 65,778,809,100 | 65,074,411,867 | 99.36 | 10.02 | 21.69 | 95.04 |
| 5000065248 | BBJ | Kanto | 452,573,162 | 449,484,158 | 67,885,974,300 | 67,112,418,479 | 99.32 | 5.61 | 22.37 | 96.19 |
| 5000065258 | BBJ | Kanto | 363,195,651 | 362,192,165 | 54,479,347,650 | 54,044,190,299 | 99.72 | 4.58 | 18.01 | 91.71 |
| 5000065268 | BBJ | Kanto | 355,685,744 | 354,725,532 | 53,352,861,600 | 52,914,209,697 | 99.73 | 4.53 | 17.64 | 90.86 |
| 5000065598 | BBJ | Kanto | 521,181,195 | 518,103,343 | 78,177,179,250 | 77,315,351,994 | 99.41 | 8.03 | 25.77 | 98.96 |
| 5999934563 | BBJ | Kanto | 341,429,588 | 340,614,300 | 51,214,438,200 | 50,849,790,202 | 99.76 | 4.91 | 16.95 | 92.36 |
| 5999934573 | BBJ | Kanto | 369,353,772 | 367,864,651 | 55,403,065,800 | 54,916,449,204 | 99.60 | 8.13 | 18.31 | 94.01 |
| 5999934593 | BBJ | Kanto | 454,894,146 | 452,569,198 | 68,234,121,900 | 67,557,979,866 | 99.49 | 5.60 | 22.52 | 96.32 |
| 5999934603 | BBJ | Kanto | 521,365,441 | 515,384,035 | 78,204,816,150 | 76,936,721,443 | 98.85 | 13.75 | 25.65 | 96.72 |
| 5999934613 | BBJ | Kanto | 434,078,009 | 431,535,259 | 65,111,701,350 | 64,462,287,726 | 99.41 | 6.32 | 21.49 | 95.78 |
| 5999934633 | BBJ | Kanto | 474,607,668 | 471,824,039 | 71,191,150,200 | 70,441,791,394 | 99.41 | 6.96 | 23.48 | 96.67 |
| 5999935101 | BBJ | Kanto | 474,745,647 | 472,186,644 | 71,211,847,050 | 70,454,508,948 | 99.46 | 6.29 | 23.48 | 98.66 |
| 5999935111 | BBJ | Kanto | 468,667,704 | 466,076,943 | 70,300,155,600 | 69,599,899,223 | 99.45 | 7.51 | 23.20 | 98.41 |
| 5999935275 | BBJ | Kanto | 452,583,781 | 450,577,564 | 67,887,567,150 | 67,231,888,168 | 99.56 | 7.31 | 22.41 | 97.20 |
| 5999935285 | BBJ | Kanto | 380,627,762 | 379,325,487 | 57,094,164,300 | 56,608,252,030 | 99.66 | 6.00 | 18.87 | 92.71 |
| 5999935295 | BBJ | Kanto | 390,789,982 | 389,587,109 | 58,618,497,300 | 58,155,916,661 | 99.69 | 8.76 | 19.39 | 92.72 |
| 5999935305 | BBJ | Kanto | 401,665,253 | 398,454,370 | 60,249,787,950 | 59,495,390,232 | 99.20 | 5.55 | 19.83 | 93.75 |
| 5999935315 | BBJ | Kanto | 497,124,908 | 495,143,735 | 74,568,736,200 | 73,937,438,959 | 99.60 | 13.34 | 24.65 | 98.57 |
| 5999935335 | BBJ | Kanto | 453,786,669 | 451,869,188 | 68,068,000,350 | 67,489,607,061 | 99.58 | 7.31 | 22.50 | 98.37 |
| 5999935537 | BBJ | Kanto | 475,813,382 | 470,846,443 | 71,372,007,300 | 70,293,409,115 | 98.96 | 10.45 | 23.43 | 98.29 |
| 5999935641 | BBJ | Kanto | 385,579,485 | 384,485,016 | 57,836,922,750 | 57,385,396,557 | 99.72 | 5.07 | 19.13 | 93.15 |
| 5999935651 | BBJ | Kanto | 396,284,147 | 394,818,542 | 59,442,622,050 | 58,946,935,222 | 99.63 | 9.84 | 19.65 | 95.84 |
| 5999935661 | BBJ | Kanto | 547,055,755 | 543,576,758 | 82,058,363,250 | 81,172,215,768 | 99.36 | 11.50 | 27.06 | 98.93 |
| 5999935671 | BBJ | Kanto | 413,683,264 | 412,162,363 | 62,052,489,600 | 61,542,600,175 | 99.63 | 7.06 | 20.51 | 97.30 |
| 5999935681 | BBJ | Kanto | 398,446,604 | 396,634,133 | 59,766,990,600 | 59,201,068,831 | 99.55 | 3.73 | 19.73 | 94.05 |
| 5999935701 | BBJ | Kanto | 493,644,203 | 491,452,252 | 74,046,630,450 | 73,391,094,589 | 99.56 | 10.37 | 24.46 | 98.69 |
| 5999935711 | BBJ | Kanto | 525,241,089 | 523,482,650 | 78,786,163,350 | 78,177,358,037 | 99.67 | 8.72 | 26.06 | 99.02 |
| 5999935721 | BBJ | Kanto | 422,342,440 | 420,876,250 | 63,351,366,000 | 62,826,883,551 | 99.65 | 6.80 | 20.94 | 97.58 |

| | | | | | | | | | | |
|---|---|---|---|---|---|---|---|---|---|---|
| 5999935829 | BBJ | Kanto | 387,758,791 | 386,356,231 | 58,163,818,650 | 57,656,282,940 | 99.64 | 7.40 | 19.22 | 92.33 |
| 5999935839 | BBJ | Kanto | 348,104,441 | 346,986,890 | 52,215,666,150 | 51,806,974,385 | 99.68 | 5.35 | 17.27 | 93.19 |
| 5999935849 | BBJ | Kanto | 482,837,053 | 480,725,772 | 72,425,557,950 | 71,768,139,323 | 99.56 | 10.93 | 23.92 | 98.39 |
| 5999935859 | BBJ | Kanto | 476,018,110 | 473,771,686 | 71,402,716,500 | 70,618,870,709 | 99.53 | 10.59 | 23.54 | 98.22 |
| 5999935869 | BBJ | Kanto | 501,089,155 | 497,927,440 | 75,163,373,250 | 74,378,072,503 | 99.37 | 9.47 | 24.79 | 96.77 |
| 5999935879 | BBJ | Kanto | 394,341,955 | 392,678,109 | 59,151,293,250 | 58,628,491,455 | 99.58 | 6.69 | 19.54 | 93.53 |
| 5999935899 | BBJ | Kanto | 467,042,011 | 465,202,160 | 70,056,301,650 | 69,488,445,412 | 99.61 | 4.93 | 23.16 | 96.81 |
| 5999935909 | BBJ | Kanto | 393,806,271 | 392,212,998 | 59,070,940,650 | 58,547,206,557 | 99.60 | 7.62 | 19.52 | 93.43 |
| 5999935919 | BBJ | Kanto | 407,190,773 | 405,443,170 | 61,078,615,950 | 60,538,151,666 | 99.57 | 7.09 | 20.18 | 94.23 |
| 5999935927 | BBJ | Kanto | 414,567,437 | 412,944,383 | 62,185,115,550 | 61,629,725,285 | 99.61 | 8.69 | 20.54 | 96.87 |
| 5999935937 | BBJ | Kanto | 453,192,845 | 450,987,451 | 67,978,926,750 | 67,340,566,822 | 99.51 | 7.14 | 22.45 | 98.11 |
| 5999935947 | BBJ | Kanto | 429,703,225 | 426,792,071 | 64,455,483,750 | 63,737,505,823 | 99.32 | 6.81 | 21.25 | 94.99 |
| 5999935957 | BBJ | Kanto | 426,570,606 | 424,542,105 | 63,985,590,900 | 63,350,064,123 | 99.52 | 4.81 | 21.12 | 95.26 |
| 5999935967 | BBJ | Kanto | 510,902,171 | 508,277,879 | 76,635,325,650 | 75,907,418,582 | 99.49 | 6.10 | 25.30 | 97.48 |
| 5999935987 | BBJ | Kanto | 437,529,420 | 435,786,614 | 65,629,413,000 | 65,069,107,816 | 99.60 | 9.22 | 21.69 | 97.78 |
| 6000063748 | BBJ | Kanto | 451,728,370 | 448,343,062 | 67,759,255,500 | 66,958,207,278 | 99.25 | 5.83 | 22.32 | 96.21 |
| 6000063758 | BBJ | Kanto | 480,544,408 | 477,784,634 | 72,081,661,200 | 71,263,936,168 | 99.43 | 11.91 | 23.75 | 98.31 |
| 6000063768 | BBJ | Kanto | 409,614,817 | 407,912,355 | 61,442,222,550 | 60,920,178,896 | 99.58 | 5.13 | 20.31 | 95.12 |
| 6000063778 | BBJ | Kanto | 467,966,476 | 466,451,450 | 70,194,971,400 | 69,677,403,818 | 99.68 | 5.45 | 23.23 | 98.80 |
| 6000063788 | BBJ | Kanto | 402,873,625 | 401,358,334 | 60,431,043,750 | 59,899,290,267 | 99.62 | 8.33 | 19.97 | 93.79 |
| 6000063798 | BBJ | Kanto | 495,749,561 | 492,499,674 | 74,362,434,150 | 73,531,894,805 | 99.34 | 7.99 | 24.51 | 96.88 |
| 6000063808 | BBJ | Kanto | 457,647,713 | 454,296,496 | 68,647,156,950 | 67,776,407,865 | 99.27 | 9.42 | 22.59 | 98.27 |
| 6000063818 | BBJ | Kanto | 464,439,806 | 462,058,834 | 69,665,970,900 | 69,021,794,936 | 99.49 | 5.59 | 23.01 | 96.62 |
| 6000063838 | BBJ | Kanto | 462,031,102 | 460,207,807 | 69,304,665,300 | 68,742,921,239 | 99.61 | 11.21 | 22.91 | 98.12 |
| 6000063848 | BBJ | Kanto | 413,293,113 | 411,683,539 | 61,993,966,950 | 61,446,258,178 | 99.61 | 6.47 | 20.48 | 94.47 |
| 6000063858 | BBJ | Kanto | 501,381,043 | 498,377,732 | 75,207,156,450 | 74,393,207,808 | 99.40 | 6.62 | 24.80 | 97.12 |
| 6000063868 | BBJ | Kanto | 353,106,051 | 351,989,919 | 52,965,907,650 | 52,529,087,385 | 99.68 | 4.81 | 17.51 | 90.79 |
| 6000064258 | BBJ | Kanto | 556,382,678 | 551,398,666 | 83,457,401,700 | 82,330,119,330 | 99.10 | 9.17 | 27.44 | 96.64 |
| 6000064266 | BBJ | Kanto | 494,786,679 | 492,465,600 | 74,218,001,850 | 73,561,349,467 | 99.53 | 8.83 | 24.52 | 97.00 |
| 6000064276 | BBJ | Kanto | 470,428,075 | 468,438,625 | 70,564,211,250 | 69,941,978,822 | 99.58 | 6.55 | 23.31 | 98.66 |
| 6000064286 | BBJ | Kanto | 461,385,739 | 458,849,934 | 69,207,860,850 | 68,537,745,276 | 99.45 | 7.59 | 22.85 | 98.38 |
| 6000064296 | BBJ | Kanto | 358,422,339 | 357,064,500 | 53,763,350,850 | 53,296,505,293 | 99.62 | 4.08 | 17.77 | 91.43 |
| 6000064306 | BBJ | Kanto | 389,891,208 | 388,548,519 | 58,483,681,200 | 57,990,299,211 | 99.66 | 5.19 | 19.33 | 96.28 |
| 6000064316 | BBJ | Kanto | 423,362,204 | 421,892,048 | 63,504,330,600 | 62,980,040,945 | 99.65 | 8.44 | 20.99 | 97.41 |
| 6000064326 | BBJ | Kanto | 460,616,023 | 457,663,357 | 69,092,403,450 | 68,364,775,176 | 99.36 | 5.00 | 22.79 | 96.26 |
| 6000064336 | BBJ | Kanto | 372,798,013 | 371,476,922 | 55,919,701,950 | 55,445,325,744 | 99.65 | 6.31 | 18.48 | 92.27 |
| 6000064370 | BBJ | Kanto | 472,214,992 | 470,093,116 | 70,832,248,800 | 70,221,901,042 | 99.55 | 7.65 | 23.41 | 98.55 |
| 6000064380 | BBJ | Kanto | 403,023,285 | 398,413,911 | 60,453,492,750 | 59,526,735,375 | 98.86 | 5.26 | 19.84 | 93.97 |
| 6000064390 | BBJ | Kanto | 532,882,881 | 529,560,971 | 79,932,432,150 | 79,103,309,618 | 99.38 | 13.67 | 26.37 | 98.83 |
| 6000064400 | BBJ | Kanto | 523,703,132 | 520,846,347 | 78,555,469,800 | 77,783,270,213 | 99.45 | 7.67 | 25.93 | 98.98 |
| 6000064410 | BBJ | Kanto | 391,637,053 | 390,304,031 | 58,745,557,950 | 58,263,115,261 | 99.66 | 7.16 | 19.42 | 96.02 |
| 6000064420 | BBJ | Kanto | 491,258,072 | 488,623,796 | 73,688,710,800 | 72,908,487,747 | 99.46 | 23.53 | 24.30 | 93.75 |
| 6000064950 | BBJ | Kanto | 445,915,020 | 443,146,303 | 66,887,253,000 | 66,131,344,272 | 99.38 | 8.05 | 22.04 | 97.87 |
| 6000064960 | BBJ | Kanto | 484,164,782 | 481,049,016 | 72,624,717,300 | 71,827,907,934 | 99.36 | 5.75 | 23.94 | 96.83 |
| 6000064966 | BBJ | Kanto | 431,940,036 | 428,242,222 | 64,791,005,400 | 63,931,588,675 | 99.14 | 5.91 | 21.31 | 97.61 |
| 6000065024 | BBJ | Kanto | 517,699,659 | 514,638,333 | 77,654,948,850 | 76,855,758,847 | 99.41 | 10.00 | 25.62 | 97.20 |
| 6000065034 | BBJ | Kanto | 502,917,982 | 500,990,150 | 75,437,697,300 | 74,810,856,579 | 99.62 | 11.19 | 24.94 | 96.90 |
| 6000065044 | BBJ | Kanto | 458,630,755 | 455,837,072 | 68,794,613,250 | 68,095,123,007 | 99.39 | 8.19 | 22.70 | 98.48 |
| 6000065054 | BBJ | Kanto | 524,077,502 | 521,364,809 | 78,611,625,300 | 77,841,872,374 | 99.48 | 10.78 | 25.95 | 97.31 |
| 6000065064 | BBJ | Kanto | 505,353,547 | 502,027,020 | 75,803,032,050 | 74,937,319,164 | 99.34 | 8.71 | 24.98 | 97.06 |
| 6000065074 | BBJ | Kanto | 464,472,960 | 461,087,842 | 69,670,944,000 | 68,880,533,912 | 99.27 | 6.14 | 22.96 | 98.45 |
| 6000065094 | BBJ | Kanto | 437,527,253 | 435,757,155 | 65,629,087,950 | 65,042,960,053 | 99.60 | 8.36 | 21.68 | 94.99 |
| 6000065434 | BBJ | Kanto | 472,793,617 | 470,626,467 | 70,919,042,550 | 70,285,170,840 | 99.54 | 8.64 | 23.43 | 96.43 |
| 6000065444 | BBJ | Kanto | 404,359,595 | 400,784,438 | 60,653,939,250 | 59,861,330,408 | 99.12 | 5.90 | 19.95 | 93.39 |
| 6000065772 | BBJ | Kanto | 484,489,501 | 482,053,753 | 72,673,425,150 | 72,028,327,595 | 99.50 | 5.40 | 24.01 | 96.97 |
| 6000065782 | BBJ | Kanto | 493,745,643 | 491,192,643 | 74,061,846,450 | 73,324,251,580 | 99.48 | 8.20 | 24.44 | 97.00 |
| 6000065792 | BBJ | Kanto | 474,905,850 | 472,285,626 | 71,235,877,500 | 70,521,398,784 | 99.45 | 7.72 | 23.51 | 96.63 |
| 6000065802 | BBJ | Kanto | 367,564,395 | 364,159,945 | 55,134,659,250 | 54,365,844,127 | 99.07 | 6.43 | 18.12 | 90.45 |
| 6000065812 | BBJ | Kanto | 350,774,665 | 348,789,024 | 52,616,199,750 | 52,086,310,669 | 99.43 | 7.19 | 17.36 | 89.06 |
| 6000065822 | BBJ | Kanto | 524,719,503 | 522,740,051 | 78,707,925,450 | 78,069,023,409 | 99.62 | 6.65 | 26.02 | 97.74 |
| 6000065832 | BBJ | Kanto | 454,149,074 | 450,180,061 | 68,122,361,100 | 67,239,201,650 | 99.13 | 4.90 | 22.41 | 96.24 |
| 6000065842 | BBJ | Kanto | 477,971,954 | 476,396,251 | 71,695,793,100 | 71,131,810,819 | 99.67 | 8.03 | 23.71 | 98.56 |
| 6000065852 | BBJ | Kanto | 429,274,876 | 427,808,970 | 64,391,231,400 | 63,861,816,478 | 99.66 | 9.30 | 21.29 | 97.43 |
| 6000065862 | BBJ | Kanto | 442,864,523 | 441,465,722 | 66,429,678,450 | 65,895,788,809 | 99.68 | 9.18 | 21.97 | 95.39 |
| 6000065872 | BBJ | Kanto | 378,979,381 | 377,021,280 | 56,846,907,150 | 56,269,720,220 | 99.48 | 3.83 | 18.76 | 95.74 |
| 6000065882 | BBJ | Kanto | 436,128,950 | 433,893,922 | 65,419,342,500 | 64,812,103,446 | 99.49 | 5.59 | 21.60 | 98.08 |
| 6000065892 | BBJ | Kanto | 535,987,530 | 532,335,046 | 80,398,129,500 | 79,467,284,494 | 99.32 | 8.08 | 26.49 | 99.02 |
| 6000065902 | BBJ | Kanto | 458,553,849 | 455,525,327 | 68,783,077,350 | 68,018,785,170 | 99.34 | 8.31 | 22.67 | 98.31 |
| 6000065912 | BBJ | Kanto | 421,598,458 | 420,218,050 | 63,239,768,700 | 62,738,262,137 | 99.67 | 8.26 | 20.91 | 94.72 |
| 6000066322 | BBJ | Kanto | 574,983,586 | 571,268,544 | 86,247,537,900 | 85,262,955,157 | 99.35 | 10.52 | 28.42 | 99.15 |
| 6000066332 | BBJ | Kanto | 523,793,929 | 521,500,336 | 78,569,089,350 | 77,840,708,329 | 99.56 | 9.46 | 25.95 | 98.96 |
| 6000066342 | BBJ | Kanto | 508,075,723 | 504,174,925 | 76,211,358,450 | 75,303,603,324 | 99.23 | 7.89 | 25.10 | 98.71 |
| 6999934693 | BBJ | Kanto | 358,508,486 | 356,835,362 | 53,776,272,900 | 53,277,050,195 | 99.53 | 6.23 | 17.76 | 93.64 |
| 6999934703 | BBJ | Kanto | 503,903,582 | 496,573,064 | 75,585,537,300 | 74,126,084,123 | 98.55 | 10.26 | 24.71 | 96.63 |
| 6999934713 | BBJ | Kanto | 465,154,636 | 461,814,262 | 69,773,195,400 | 68,931,287,490 | 99.28 | 11.15 | 22.98 | 98.27 |
| 6999935183 | BBJ | Kanto | 508,660,376 | 506,505,638 | 76,299,056,400 | 75,618,205,854 | 99.58 | 7.78 | 25.21 | 97.35 |
| 6999935193 | BBJ | Kanto | 450,882,609 | 448,572,446 | 67,632,391,350 | 66,986,270,128 | 99.49 | 6.77 | 22.33 | 95.89 |
| 6999935203 | BBJ | Kanto | 439,247,305 | 437,607,960 | 65,887,095,750 | 65,345,272,422 | 99.63 | 7.34 | 21.78 | 97.95 |
| 6999935213 | BBJ | Kanto | 406,366,356 | 404,808,189 | 60,954,953,400 | 60,399,199,755 | 99.62 | 6.98 | 20.13 | 96.77 |
| 6999935223 | BBJ | Kanto | 544,387,110 | 538,495,282 | 81,658,066,500 | 80,349,748,461 | 98.92 | 10.15 | 26.78 | 97.53 |
| 6999935233 | BBJ | Kanto | 444,730,513 | 443,166,727 | 66,709,576,950 | 66,203,964,928 | 99.65 | 5.67 | 22.07 | 98.44 |
| 6999935243 | BBJ | Kanto | 394,887,726 | 392,026,103 | 59,233,158,900 | 58,564,191,863 | 99.28 | 5.29 | 19.52 | 93.75 |
| 6999935253 | BBJ | Kanto | 394,044,621 | 392,449,087 | 59,106,693,150 | 58,571,515,356 | 99.60 | 7.39 | 19.52 | 96.46 |
| 6999935267 | BBJ | Kanto | 449,014,791 | 445,882,512 | 67,352,218,650 | 66,557,389,070 | 99.30 | 5.89 | 22.19 | 95.92 |
| 6999935277 | BBJ | Kanto | 487,274,548 | 484,925,747 | 73,091,182,200 | 72,416,493,884 | 99.52 | 7.94 | 24.14 | 96.79 |
| 6999935287 | BBJ | Kanto | 494,440,351 | 491,975,227 | 74,166,052,650 | 73,429,364,800 | 99.50 | 11.02 | 24.48 | 98.65 |
| 6999935381 | BBJ | Kanto | 481,447,853 | 476,945,563 | 72,217,177,950 | 71,160,149,534 | 99.06 | 8.14 | 23.72 | 96.41 |
| 6999935797 | BBJ | Kanto | 406,863,468 | 405,086,859 | 61,029,520,200 | 60,470,008,843 | 99.56 | 9.64 | 20.16 | 96.26 |
| 6999935801 | BBJ | Kanto | 494,760,798 | 492,100,840 | 74,214,119,700 | 73,451,031,953 | 99.46 | 8.56 | 24.48 | 96.96 |
| 6999935807 | BBJ | Kanto | 536,073,880 | 533,416,847 | 80,411,082,000 | 79,619,004,990 | 99.50 | 9.95 | 26.54 | 98.98 |
| 6999935811 | BBJ | Kanto | 440,356,090 | 437,864,495 | 66,053,413,500 | 65,381,213,834 | 99.43 | 7.76 | 21.79 | 98.01 |
| 6999935817 | BBJ | Kanto | 374,127,245 | 372,065,364 | 56,119,086,750 | 55,532,769,034 | 99.45 | 6.00 | 18.51 | 91.86 |
| 6999935821 | BBJ | Kanto | 450,289,535 | 448,162,429 | 67,543,430,250 | 66,928,075,494 | 99.53 | 6.56 | 22.31 | 96.17 |
| 6999935827 | BBJ | Kanto | 480,735,688 | 479,061,961 | 72,110,353,200 | 71,496,342,108 | 99.65 | 5.56 | 23.83 | 96.57 |
| 6999935831 | BBJ | Kanto | 469,541,647 | 466,002,565 | 70,431,247,050 | 69,577,753,909 | 99.25 | 6.05 | 23.19 | 96.62 |
| 6999935837 | BBJ | Kanto | 383,351,571 | 382,075,923 | 57,502,735,650 | 57,044,278,600 | 99.67 | 4.40 | 19.01 | 96.50 |
| 6999935841 | BBJ | Kanto | 443,726,879 | 440,805,776 | 66,559,031,850 | 65,835,757,012 | 99.34 | 7.23 | 21.95 | 95.62 |
| 6999935851 | BBJ | Kanto | 409,116,480 | 407,755,319 | 61,367,472,000 | 60,849,381,217 | 99.67 | 6.47 | 20.28 | 97.11 |
| 6999935857 | BBJ | Kanto | 426,508,033 | 421,367,315 | 63,976,204,950 | 62,925,668,399 | 98.79 | 5.00 | 20.98 | 94.70 |
| 6999935861 | BBJ | Kanto | 388,846,462 | 387,481,790 | 58,326,969,300 | 57,822,053,641 | 99.65 | 7.33 | 19.27 | 93.07 |

| | | | | | | | | | | |
|---|---|---|---|---|---|---|---|---|---|---|
| 6999935867 | BBJ | Kanto | 432,015,666 | 429,932,596 | 64,802,349,900 | 64,203,366,721 | 99.52 | 5.24 | 21.40 | 95.82 |
| 6999935871 | BBJ | Kanto | 525,687,245 | 523,520,304 | 78,853,086,750 | 78,157,652,954 | 99.59 | 6.42 | 26.05 | 99.08 |
| 6999935877 | BBJ | Kanto | 397,326,818 | 395,792,542 | 59,599,022,700 | 59,074,631,774 | 99.61 | 8.13 | 19.69 | 93.45 |
| 6999935887 | BBJ | Kanto | 397,763,472 | 395,865,880 | 59,664,520,800 | 59,083,611,178 | 99.52 | 7.63 | 19.69 | 96.38 |
| 6999935897 | BBJ | Kanto | 485,276,165 | 482,955,701 | 72,791,424,750 | 72,118,377,206 | 99.52 | 10.33 | 24.04 | 96.54 |
| 6999936755 | BBJ | Kanto | 465,317,052 | 462,401,788 | 69,797,557,800 | 69,046,954,820 | 99.37 | 6.09 | 23.02 | 98.54 |
| 6999936929 | BBJ | Kanto | 460,977,121 | 458,120,347 | 69,146,568,150 | 68,369,003,171 | 99.38 | 9.21 | 22.79 | 98.16 |
| 6999936939 | BBJ | Kanto | 464,505,094 | 462,785,550 | 69,675,764,100 | 69,091,531,925 | 99.63 | 9.14 | 23.03 | 98.32 |
| 6999936949 | BBJ | Kanto | 442,959,862 | 440,491,993 | 66,443,979,300 | 65,768,185,947 | 99.44 | 8.58 | 21.92 | 97.90 |
| 6999936959 | BBJ | Kanto | 404,043,708 | 402,585,227 | 60,606,556,200 | 60,083,646,435 | 99.64 | 8.22 | 20.03 | 96.13 |
| 6999936969 | BBJ | Kanto | 419,655,728 | 418,172,526 | 62,948,359,200 | 62,423,112,606 | 99.65 | 8.99 | 20.81 | 97.38 |
| 6999936979 | BBJ | Kanto | 429,821,960 | 428,501,321 | 64,473,294,000 | 63,949,563,142 | 99.69 | 10.13 | 21.32 | 94.78 |
| 6999936989 | BBJ | Kanto | 468,662,177 | 465,770,764 | 70,299,326,550 | 69,551,400,299 | 99.38 | 6.62 | 23.18 | 96.36 |
| 6999936999 | BBJ | Kanto | 437,457,483 | 435,471,557 | 65,618,622,450 | 65,028,937,563 | 99.55 | 8.75 | 21.68 | 95.14 |
| 6999937009 | BBJ | Kanto | 465,927,648 | 463,976,736 | 69,889,147,200 | 69,302,132,298 | 99.58 | 5.19 | 23.10 | 96.87 |
| 6999937019 | BBJ | Kanto | 481,770,272 | 479,796,657 | 72,265,540,800 | 71,649,495,935 | 99.59 | 8.08 | 23.88 | 96.66 |
| 6999937325 | BBJ | Kanto | 376,609,919 | 374,998,221 | 56,491,487,850 | 55,946,952,123 | 99.57 | 7.79 | 18.65 | 91.92 |
| 6999937335 | BBJ | Kanto | 400,059,310 | 398,332,838 | 60,008,896,500 | 59,446,065,639 | 99.57 | 6.18 | 19.82 | 93.74 |
| 6999937345 | BBJ | Kanto | 499,688,216 | 498,115,746 | 74,953,232,400 | 74,328,348,417 | 99.69 | 4.92 | 24.78 | 97.22 |
| 6999937365 | BBJ | Kanto | 457,230,329 | 454,155,770 | 68,584,549,350 | 67,796,282,685 | 99.33 | 7.93 | 22.60 | 95.97 |
| 6999937375 | BBJ | Kanto | 446,982,649 | 444,249,841 | 67,047,397,350 | 66,206,909,509 | 99.39 | 9.16 | 22.07 | 97.73 |
| 6999937385 | BBJ | Kanto | 444,563,953 | 441,414,487 | 66,684,592,950 | 65,906,081,562 | 99.29 | 4.95 | 21.97 | 98.11 |
| 6999937395 | BBJ | Kanto | 414,336,059 | 410,931,814 | 62,150,408,850 | 61,385,626,813 | 99.18 | 4.02 | 20.46 | 97.54 |
| 6999937405 | BBJ | Kanto | 476,188,889 | 472,402,567 | 71,428,333,350 | 70,556,076,117 | 99.20 | 8.65 | 23.52 | 98.38 |
| 6999937415 | BBJ | Kanto | 480,570,273 | 476,508,240 | 72,085,540,950 | 71,108,682,084 | 99.15 | 8.57 | 23.70 | 96.41 |
| 6999937425 | BBJ | Kanto | 401,657,296 | 399,997,495 | 60,248,594,400 | 59,697,212,719 | 99.59 | 6.23 | 19.90 | 93.89 |
| 6999937435 | BBJ | Kanto | 471,277,026 | 469,340,321 | 70,691,553,900 | 70,094,945,748 | 99.59 | 14.16 | 23.36 | 98.06 |
| 6999937445 | BBJ | Kanto | 537,046,229 | 533,863,308 | 80,556,934,350 | 79,745,118,354 | 99.41 | 10.44 | 26.58 | 98.98 |
| 6999937455 | BBJ | Kanto | 446,835,594 | 444,229,016 | 67,025,339,100 | 66,274,785,374 | 99.42 | 8.51 | 22.09 | 95.59 |
| 6999937465 | BBJ | Kanto | 382,008,672 | 379,666,098 | 57,301,300,800 | 56,689,711,191 | 99.39 | 5.39 | 18.90 | 93.25 |
| 6999937475 | BBJ | Kanto | 439,272,746 | 437,119,521 | 65,890,911,900 | 65,278,383,628 | 99.51 | 9.28 | 21.76 | 97.88 |
| 7000063092 | BBJ | Kanto | 468,685,903 | 466,265,382 | 70,302,885,450 | 69,624,745,692 | 99.48 | 8.53 | 23.21 | 96.48 |
| 7000063102 | BBJ | Kanto | 517,472,133 | 514,444,871 | 77,620,819,950 | 76,829,702,494 | 99.41 | 14.38 | 25.61 | 96.73 |
| 7000063112 | BBJ | Kanto | 457,186,573 | 454,998,424 | 68,577,985,950 | 67,942,596,033 | 99.52 | 5.81 | 22.65 | 96.46 |
| 7000063132 | BBJ | Kanto | 382,566,524 | 381,348,904 | 57,384,978,600 | 56,935,670,149 | 99.68 | 8.83 | 18.98 | 95.16 |
| 7000063142 | BBJ | Kanto | 417,411,335 | 415,833,753 | 62,611,700,250 | 62,036,867,879 | 99.62 | 7.36 | 20.68 | 94.50 |
| 7000063152 | BBJ | Kanto | 475,405,794 | 472,983,547 | 71,310,869,100 | 70,654,998,125 | 99.49 | 6.99 | 23.55 | 96.65 |
| 7000063162 | BBJ | Kanto | 438,854,863 | 437,008,828 | 65,828,229,450 | 65,223,396,405 | 99.58 | 8.14 | 21.74 | 97.57 |
| 7000063210 | BBJ | Kanto | 465,556,254 | 463,608,236 | 69,833,438,100 | 69,225,284,617 | 99.58 | 8.57 | 23.08 | 96.27 |
| 7000063220 | BBJ | Kanto | 481,558,049 | 478,917,624 | 72,233,707,350 | 71,525,658,231 | 99.45 | 8.35 | 23.84 | 96.77 |
| 7000063230 | BBJ | Kanto | 431,357,268 | 426,663,829 | 64,703,590,200 | 63,718,783,980 | 98.91 | 8.48 | 21.24 | 94.28 |
| 7000063260 | BBJ | Kanto | 331,954,598 | 330,938,546 | 49,793,189,700 | 49,401,057,057 | 99.69 | 4.89 | 16.47 | 90.97 |
| 7000063702 | BBJ | Kanto | 463,578,284 | 461,084,528 | 69,536,742,600 | 68,858,570,410 | 99.46 | 10.57 | 22.95 | 95.86 |
| 7000063786 | BBJ | Kanto | 498,043,215 | 495,569,319 | 74,706,482,250 | 74,016,375,123 | 99.50 | 5.46 | 24.67 | 97.40 |
| 7000063796 | BBJ | Kanto | 510,163,639 | 506,617,479 | 76,524,545,850 | 75,670,752,002 | 99.30 | 10.34 | 25.22 | 97.03 |
| 7000063800 | BBJ | Kanto | 483,621,351 | 480,579,287 | 72,543,202,650 | 71,725,254,814 | 99.37 | 10.69 | 23.91 | 98.45 |
| 7000063810 | BBJ | Kanto | 462,140,138 | 459,457,962 | 69,321,020,700 | 68,615,586,243 | 99.42 | 7.30 | 22.87 | 98.45 |
| 7000063816 | BBJ | Kanto | 497,388,930 | 494,432,372 | 74,608,339,500 | 73,805,602,877 | 99.41 | 9.25 | 24.60 | 98.64 |
| 7000063820 | BBJ | Kanto | 478,030,434 | 475,488,269 | 71,704,565,100 | 71,002,175,436 | 99.47 | 7.65 | 23.67 | 98.60 |
| 7000063830 | BBJ | Kanto | 493,926,331 | 490,361,053 | 74,088,949,650 | 73,198,747,834 | 99.28 | 8.08 | 24.40 | 98.62 |
| 7000063840 | BBJ | Kanto | 469,949,454 | 463,282,091 | 70,492,418,100 | 69,150,003,708 | 98.58 | 7.47 | 23.05 | 98.36 |
| 7000064308 | BBJ | Kanto | 399,476,928 | 398,150,572 | 59,921,539,200 | 59,427,772,786 | 99.67 | 7.41 | 19.81 | 96.64 |
| 7000064328 | BBJ | Kanto | 497,860,749 | 493,691,426 | 74,679,112,350 | 73,687,452,941 | 99.16 | 9.81 | 24.56 | 98.76 |
| 7000064338 | BBJ | Kanto | 443,812,349 | 441,802,114 | 66,571,852,350 | 65,909,188,762 | 99.55 | 6.57 | 21.97 | 95.53 |
| 7000064358 | BBJ | Kanto | 519,368,609 | 516,588,365 | 77,905,291,350 | 77,080,342,974 | 99.46 | 11.37 | 25.69 | 98.73 |
| 7000064368 | BBJ | Kanto | 389,045,480 | 387,700,841 | 58,356,822,000 | 57,859,189,644 | 99.65 | 6.23 | 19.29 | 96.12 |
| 7000064378 | BBJ | Kanto | 427,187,981 | 425,626,930 | 64,078,197,150 | 63,523,373,371 | 99.63 | 9.27 | 21.17 | 97.26 |
| 7000064406 | BBJ | Kanto | 498,440,171 | 496,577,322 | 74,766,025,650 | 74,108,282,958 | 99.63 | 9.24 | 24.70 | 98.80 |
| 7000064416 | BBJ | Kanto | 424,137,742 | 421,188,776 | 63,620,661,300 | 62,910,074,393 | 99.30 | 5.14 | 20.97 | 97.79 |
| 7000064914 | BBJ | Kanto | 516,709,507 | 514,389,519 | 77,506,426,050 | 76,839,878,561 | 99.55 | 11.03 | 25.61 | 97.19 |
| 7000064924 | BBJ | Kanto | 423,316,340 | 421,055,967 | 63,497,451,000 | 62,877,291,602 | 99.47 | 6.45 | 20.96 | 97.85 |
| 7000064944 | BBJ | Kanto | 531,664,427 | 528,975,453 | 79,749,664,050 | 78,993,297,438 | 99.49 | 15.42 | 26.33 | 98.80 |
| 7000064964 | BBJ | Kanto | 356,403,370 | 354,970,625 | 53,460,505,500 | 52,979,102,645 | 99.60 | 4.63 | 17.66 | 90.99 |
| 7000064974 | BBJ | Kanto | 494,650,515 | 492,884,366 | 74,197,577,250 | 73,551,536,555 | 99.64 | 6.52 | 24.52 | 97.16 |
| 7000064984 | BBJ | Kanto | 435,777,917 | 434,232,951 | 65,366,687,550 | 64,797,918,421 | 99.65 | 7.85 | 21.60 | 95.22 |
| 7000064994 | BBJ | Kanto | 463,941,695 | 460,993,783 | 69,591,254,250 | 68,851,518,650 | 99.36 | 7.45 | 22.95 | 96.26 |
| 7000065004 | BBJ | Kanto | 519,643,194 | 516,521,274 | 77,946,479,100 | 77,077,880,891 | 99.40 | 8.58 | 25.69 | 97.37 |
| 7000065014 | BBJ | Kanto | 473,421,572 | 470,634,774 | 71,013,235,800 | 70,246,249,124 | 99.41 | 7.31 | 23.42 | 98.51 |
| 7000065024 | BBJ | Kanto | 442,516,493 | 439,713,622 | 66,377,473,950 | 65,646,466,923 | 99.37 | 6.60 | 21.88 | 95.69 |
| 7000065044 | BBJ | Kanto | 366,377,508 | 365,177,938 | 54,956,626,200 | 54,479,041,301 | 99.67 | 4.73 | 18.16 | 91.39 |
| 7999934265 | BBJ | Kanto | 352,728,086 | 351,755,611 | 52,909,212,900 | 52,474,438,885 | 99.72 | 4.51 | 17.49 | 90.61 |
| 7999934275 | BBJ | Kanto | 453,951,108 | 451,942,241 | 68,092,666,200 | 67,470,556,442 | 99.56 | 8.10 | 22.49 | 95.94 |
| 7999934285 | BBJ | Kanto | 429,957,384 | 427,688,988 | 64,493,607,600 | 63,839,241,395 | 99.47 | 5.23 | 21.28 | 97.76 |
| 7999934295 | BBJ | Kanto | 416,455,358 | 412,042,577 | 62,468,303,700 | 61,548,556,786 | 98.94 | 7.80 | 20.52 | 96.39 |
| 7999934305 | BBJ | Kanto | 473,340,267 | 470,838,404 | 71,001,040,050 | 70,317,291,679 | 99.47 | 7.98 | 23.44 | 98.57 |
| 7999934315 | BBJ | Kanto | 472,212,633 | 467,856,966 | 70,831,894,950 | 69,862,177,385 | 99.08 | 7.32 | 23.29 | 96.44 |
| 7999934325 | BBJ | Kanto | 484,627,361 | 482,532,640 | 72,694,104,150 | 72,078,675,395 | 99.57 | 7.81 | 24.03 | 98.76 |
| 7999934335 | BBJ | Kanto | 497,837,143 | 495,384,598 | 74,675,571,450 | 73,979,434,461 | 99.51 | 7.94 | 24.66 | 97.15 |
| 7999934345 | BBJ | Kanto | 519,370,275 | 514,412,168 | 77,905,541,250 | 76,732,800,465 | 99.05 | 10.04 | 25.58 | 97.13 |
| 7999934467 | BBJ | Kanto | 460,281,472 | 455,657,279 | 69,042,220,800 | 67,965,956,518 | 99.00 | 7.82 | 22.66 | 95.87 |
| 7999934937 | BBJ | Kanto | 469,020,499 | 464,402,164 | 70,353,074,850 | 69,326,955,548 | 99.02 | 6.72 | 23.11 | 98.46 |
| 7999934967 | BBJ | Kanto | 461,734,767 | 459,250,504 | 69,260,215,050 | 68,547,041,183 | 99.46 | 8.25 | 22.85 | 98.36 |
| 7999934977 | BBJ | Kanto | 487,240,943 | 484,557,888 | 73,086,141,450 | 72,335,012,067 | 99.45 | 9.68 | 24.11 | 98.42 |
| 7999934987 | BBJ | Kanto | 405,374,583 | 404,092,569 | 60,806,187,450 | 60,307,125,751 | 99.68 | 5.97 | 20.10 | 96.87 |
| 7999934997 | BBJ | Kanto | 406,038,555 | 404,694,973 | 60,905,783,250 | 60,363,180,644 | 99.67 | 6.72 | 20.12 | 94.07 |
| 7999935007 | BBJ | Kanto | 495,699,566 | 493,824,989 | 74,354,934,900 | 73,762,554,395 | 99.62 | 16.64 | 24.59 | 98.25 |
| 7999935017 | BBJ | Kanto | 435,305,494 | 431,722,001 | 65,295,824,100 | 64,485,849,856 | 99.18 | 8.59 | 21.50 | 95.14 |
| 7999935027 | BBJ | Kanto | 446,710,573 | 444,239,003 | 67,006,585,950 | 66,335,118,905 | 99.45 | 9.27 | 22.11 | 95.50 |
| 7999935037 | BBJ | Kanto | 348,323,966 | 346,782,356 | 52,248,594,900 | 51,778,751,942 | 99.56 | 5.97 | 17.26 | 92.94 |
| 7999935047 | BBJ | Kanto | 507,638,191 | 505,331,986 | 76,145,728,650 | 75,456,497,093 | 99.55 | 6.54 | 25.15 | 97.42 |
| 7999935073 | BBJ | Kanto | 496,773,026 | 494,162,863 | 74,515,953,900 | 73,789,923,453 | 99.47 | 8.90 | 24.60 | 96.93 |
| 7999935493 | BBJ | Kanto | 385,061,296 | 383,669,809 | 57,759,194,400 | 57,280,249,747 | 99.64 | 9.23 | 19.09 | 92.17 |
| 7999935503 | BBJ | Kanto | 438,828,359 | 435,919,321 | 65,824,253,850 | 65,071,533,247 | 99.34 | 7.28 | 21.69 | 95.33 |
| 7999935513 | BBJ | Kanto | 489,450,085 | 486,971,145 | 73,417,512,750 | 72,676,517,854 | 99.49 | 10.89 | 24.23 | 96.36 |
| 7999935523 | BBJ | Kanto | 521,278,729 | 518,946,842 | 78,191,809,350 | 77,374,450,035 | 99.55 | 9.29 | 25.79 | 98.79 |
| 7999935533 | BBJ | Kanto | 441,268,774 | 438,870,390 | 66,190,316,100 | 65,508,315,317 | 99.46 | 8.20 | 21.84 | 95.12 |
| 7999935543 | BBJ | Kanto | 489,262,510 | 486,989,871 | 73,389,376,500 | 72,730,437,423 | 99.54 | 21.29 | 24.24 | 94.66 |
| 7999935553 | BBJ | Kanto | 546,671,858 | 544,297,592 | 82,000,778,700 | 81,277,720,288 | 99.57 | 11.43 | 27.09 | 97.47 |
| 7999936051 | BBJ | Kanto | 409,998,985 | 408,808,406 | 61,499,847,750 | 61,009,342,702 | 99.71 | 6.82 | 20.34 | 97.14 |
| 7999936061 | BBJ | Kanto | 443,949,652 | 442,558,411 | 66,592,447,800 | 66,051,546,988 | 99.69 | 8.58 | 22.02 | 97.98 |

| | | | | | | | | | | |
|---|---|---|---|---|---|---|---|---|---|---|
| 7999936071 | BBJ | Kanto | 452,667,244 | 450,501,983 | 67,900,086,600 | 67,191,005,108 | 99.52 | 4.66 | 22.40 | 96.19 |
| 7999936081 | BBJ | Kanto | 364,131,886 | 362,363,726 | 54,619,782,900 | 54,082,287,473 | 99.51 | 19.35 | 18.03 | 83.73 |
| 7999936091 | BBJ | Kanto | 450,485,467 | 447,925,031 | 67,572,820,050 | 66,865,488,384 | 99.43 | 4.77 | 22.29 | 96.23 |
| 7999936101 | BBJ | Kanto | 469,448,032 | 464,182,135 | 70,417,204,800 | 69,329,431,245 | 98.88 | 9.07 | 23.11 | 96.15 |
| 7999936111 | BBJ | Kanto | 412,722,688 | 411,120,119 | 61,908,403,200 | 61,367,356,033 | 99.61 | 7.66 | 20.46 | 96.96 |
| 7999936121 | BBJ | Kanto | 402,173,679 | 400,652,001 | 60,326,051,850 | 59,814,876,612 | 99.62 | 6.47 | 19.94 | 96.85 |
| 7999936131 | BBJ | Kanto | 448,171,900 | 445,065,004 | 67,225,785,000 | 66,439,541,022 | 99.31 | 9.83 | 22.15 | 97.67 |
| 7999936409 | BBJ | Kanto | 401,764,431 | 399,832,011 | 60,264,664,650 | 59,685,327,264 | 99.52 | 7.02 | 19.90 | 93.63 |
| 7999936419 | BBJ | Kanto | 464,759,692 | 463,221,312 | 69,713,953,800 | 69,146,454,728 | 99.67 | 7.07 | 23.05 | 98.33 |
| 7999936429 | BBJ | Kanto | 485,560,423 | 481,884,386 | 72,834,063,450 | 71,925,273,472 | 99.24 | 8.02 | 23.98 | 96.54 |
| 7999936439 | BBJ | Kanto | 444,471,749 | 440,801,245 | 66,670,762,350 | 65,781,289,631 | 99.17 | 4.98 | 21.93 | 98.12 |
| 7999936449 | BBJ | Kanto | 410,235,320 | 408,419,799 | 61,535,298,000 | 60,953,601,913 | 99.56 | 4.62 | 20.32 | 97.21 |
| 7999936459 | BBJ | Kanto | 434,512,837 | 433,102,824 | 65,176,925,550 | 64,643,137,126 | 99.68 | 8.08 | 21.55 | 97.78 |
| 7999936469 | BBJ | Kanto | 459,095,442 | 457,127,054 | 68,864,316,300 | 68,288,005,374 | 99.57 | 5.74 | 22.76 | 98.64 |
| 7999936479 | BBJ | Kanto | 465,312,561 | 462,952,998 | 69,796,884,150 | 69,155,713,950 | 99.49 | 6.80 | 23.05 | 96.54 |
| 7999936489 | BBJ | Kanto | 481,532,042 | 479,751,038 | 72,229,806,300 | 71,654,211,595 | 99.63 | 8.18 | 23.88 | 98.64 |
| 7999936499 | BBJ | Kanto | 407,274,074 | 405,243,705 | 61,091,111,100 | 60,530,879,793 | 99.50 | 4.27 | 20.18 | 97.54 |
| 8000063300 | BBJ | Kanto | 495,803,798 | 493,430,164 | 74,370,569,700 | 73,690,879,906 | 99.52 | 10.21 | 24.56 | 98.74 |
| 8000063320 | BBJ | Kanto | 480,451,401 | 478,299,767 | 72,067,710,150 | 71,434,332,713 | 99.55 | 8.02 | 23.81 | 96.81 |
| 8000063330 | BBJ | Kanto | 418,139,175 | 416,169,750 | 62,720,876,250 | 62,105,972,503 | 99.53 | 5.26 | 20.70 | 94.81 |
| 8000063340 | BBJ | Kanto | 430,058,809 | 428,209,519 | 64,508,821,350 | 63,905,855,516 | 99.57 | 3.93 | 21.30 | 97.97 |
| 8000063350 | BBJ | Kanto | 373,068,734 | 371,622,769 | 55,960,310,100 | 55,488,320,858 | 99.61 | 3.99 | 18.50 | 93.03 |
| 8000063360 | BBJ | Kanto | 495,652,573 | 493,939,639 | 74,347,885,950 | 73,696,150,094 | 99.65 | 6.11 | 24.57 | 97.11 |
| 8000063370 | BBJ | Kanto | 558,189,030 | 556,615,177 | 83,728,354,500 | 83,089,403,305 | 99.72 | 5.92 | 27.70 | 99.14 |
| 8000063380 | BBJ | Kanto | 445,569,283 | 442,301,283 | 66,835,392,450 | 66,048,443,299 | 99.27 | 4.98 | 22.02 | 98.11 |
| 8000063688 | BBJ | Kanto | 518,930,763 | 515,857,682 | 77,839,614,450 | 76,927,372,315 | 99.41 | 10.92 | 25.64 | 97.08 |
| 8000063698 | BBJ | Kanto | 476,641,350 | 472,421,633 | 71,496,202,500 | 70,526,740,431 | 99.11 | 11.02 | 23.51 | 96.10 |
| 8000063708 | BBJ | Kanto | 383,933,170 | 382,720,752 | 57,589,975,500 | 57,118,926,339 | 99.68 | 10.62 | 19.04 | 94.95 |
| 8000063728 | BBJ | Kanto | 429,040,902 | 426,314,068 | 64,356,135,300 | 63,678,217,930 | 99.36 | 6.40 | 21.23 | 97.79 |
| 8000063738 | BBJ | Kanto | 464,924,720 | 461,935,165 | 69,738,708,000 | 68,982,513,250 | 99.36 | 5.01 | 22.99 | 98.55 |
| 8000063748 | BBJ | Kanto | 485,993,417 | 480,199,282 | 72,899,012,550 | 71,680,775,527 | 98.81 | 8.26 | 23.89 | 96.51 |
| 8000063758 | BBJ | Kanto | 425,904,561 | 423,488,748 | 63,885,684,150 | 63,257,601,195 | 99.43 | 4.97 | 21.09 | 97.74 |
| 8000063768 | BBJ | Kanto | 420,765,547 | 415,327,075 | 63,114,832,050 | 62,044,023,162 | 98.71 | 4.25 | 20.68 | 97.45 |
| 8000063778 | BBJ | Kanto | 479,230,836 | 477,298,682 | 71,884,625,400 | 71,277,342,648 | 99.60 | 9.69 | 23.76 | 98.38 |
| 8000063780 | BBJ | Kanto | 507,755,166 | 503,829,412 | 76,163,274,900 | 75,238,052,994 | 99.23 | 6.94 | 25.08 | 98.93 |
| 8000063792 | BBJ | Kanto | 472,315,213 | 470,725,303 | 70,847,281,950 | 70,321,002,330 | 99.66 | 6.16 | 23.44 | 98.76 |
| 8000063802 | BBJ | Kanto | 366,970,219 | 365,002,662 | 55,045,532,850 | 54,484,144,049 | 99.46 | 5.19 | 18.16 | 91.92 |
| 8000063812 | BBJ | Kanto | 436,655,939 | 433,931,842 | 65,498,390,850 | 64,793,233,994 | 99.38 | 12.63 | 21.60 | 94.51 |
| 8000063822 | BBJ | Kanto | 392,861,960 | 391,538,696 | 58,929,294,000 | 58,455,301,644 | 99.66 | 6.94 | 19.49 | 93.34 |
| 8000063832 | BBJ | Kanto | 423,678,729 | 422,361,298 | 63,551,809,350 | 63,068,074,091 | 99.69 | 8.19 | 21.02 | 94.93 |
| 8000063852 | BBJ | Kanto | 497,653,011 | 493,473,020 | 74,647,951,650 | 73,674,947,906 | 99.16 | 10.45 | 24.56 | 96.41 |
| 8000063862 | BBJ | Kanto | 393,228,217 | 391,998,297 | 58,984,232,550 | 58,502,467,081 | 99.69 | 8.06 | 19.50 | 95.84 |
| 8000064054 | BBJ | Kanto | 486,187,102 | 483,402,844 | 72,928,065,300 | 72,192,345,382 | 99.43 | 6.39 | 24.06 | 97.03 |
| 8000064064 | BBJ | Kanto | 390,362,408 | 388,066,055 | 58,554,361,200 | 57,974,542,863 | 99.41 | 6.07 | 19.32 | 96.22 |
| 8000064074 | BBJ | Kanto | 478,735,091 | 476,519,500 | 71,810,263,650 | 71,068,300,498 | 99.54 | 10.73 | 23.69 | 96.22 |
| 8000064084 | BBJ | Kanto | 491,490,729 | 488,639,309 | 73,723,609,350 | 72,970,515,563 | 99.42 | 5.92 | 24.32 | 97.00 |
| 8000064298 | BBJ | Kanto | 467,950,646 | 464,116,718 | 70,192,596,900 | 69,289,965,593 | 99.18 | 8.16 | 23.10 | 98.34 |
| 8000064308 | BBJ | Kanto | 463,106,310 | 459,121,802 | 69,465,946,500 | 68,555,500,764 | 99.14 | 7.11 | 22.85 | 98.46 |
| 8000064318 | BBJ | Kanto | 527,246,687 | 523,392,085 | 79,087,003,050 | 78,141,811,384 | 99.27 | 9.67 | 26.05 | 97.27 |
| 8000064706 | BBJ | Kanto | 474,833,844 | 473,001,983 | 71,225,076,600 | 70,675,033,250 | 99.61 | 4.85 | 23.56 | 98.69 |
| 8000064726 | BBJ | Kanto | 423,219,410 | 421,842,187 | 63,482,911,500 | 62,971,182,843 | 99.67 | 9.09 | 20.99 | 97.34 |
| 8000064736 | BBJ | Kanto | 460,229,671 | 458,729,982 | 69,034,450,650 | 68,490,814,290 | 99.67 | 13.33 | 22.83 | 95.46 |
| 8000064746 | BBJ | Kanto | 513,345,103 | 511,101,833 | 77,001,765,450 | 76,352,697,993 | 99.56 | 9.37 | 25.45 | 97.33 |
| 8000064766 | BBJ | Kanto | 408,406,450 | 406,678,709 | 61,260,967,500 | 60,703,742,938 | 99.58 | 6.49 | 20.23 | 94.35 |
| 8000064776 | BBJ | Kanto | 423,782,080 | 421,836,895 | 63,567,312,000 | 62,999,547,016 | 99.54 | 6.44 | 21.00 | 97.57 |
| 8000064786 | BBJ | Kanto | 503,171,132 | 500,033,620 | 75,475,669,800 | 74,622,425,631 | 99.38 | 11.40 | 24.87 | 98.63 |
| 8000064806 | BBJ | Kanto | 434,329,949 | 432,885,670 | 65,149,492,350 | 64,647,140,831 | 99.67 | 5.76 | 21.55 | 98.19 |
| 8000064816 | BBJ | Kanto | 423,069,189 | 421,575,657 | 63,460,378,350 | 62,937,878,270 | 99.65 | 7.10 | 20.98 | 97.64 |
| 8000065196 | BBJ | Kanto | 471,845,269 | 468,085,213 | 70,776,790,350 | 69,875,630,483 | 99.20 | 10.01 | 23.29 | 96.14 |
| 8000065216 | BBJ | Kanto | 391,792,701 | 389,387,482 | 58,768,905,150 | 58,136,385,329 | 99.39 | 5.23 | 19.38 | 93.50 |
| 8999935055 | BBJ | Kanto | 496,103,175 | 491,614,588 | 74,415,476,250 | 73,373,657,329 | 99.10 | 13.44 | 24.46 | 98.35 |
| 8999935065 | BBJ | Kanto | 494,199,717 | 490,958,281 | 74,129,957,550 | 73,274,357,401 | 99.34 | 12.02 | 24.42 | 98.49 |
| 8999935099 | BBJ | Kanto | 461,749,373 | 459,542,477 | 69,262,405,950 | 68,599,849,619 | 99.52 | 7.69 | 22.87 | 96.13 |
| 8999935223 | BBJ | Kanto | 413,228,321 | 410,488,334 | 61,984,248,150 | 61,295,406,647 | 99.34 | 5.78 | 20.43 | 97.27 |
| 8999935233 | BBJ | Kanto | 467,046,584 | 463,675,269 | 70,056,987,600 | 69,197,977,568 | 99.28 | 10.63 | 23.07 | 95.95 |
| 8999935243 | BBJ | Kanto | 510,329,552 | 505,963,188 | 76,549,432,800 | 75,544,487,799 | 99.14 | 9.45 | 25.18 | 98.88 |
| 8999935485 | BBJ | Kanto | 420,146,183 | 418,306,590 | 63,021,927,450 | 62,433,042,702 | 99.56 | 7.75 | 20.81 | 94.47 |
| 8999935495 | BBJ | Kanto | 441,199,806 | 439,076,136 | 66,179,970,900 | 65,478,513,177 | 99.52 | 4.09 | 21.83 | 95.97 |
| 8999935515 | BBJ | Kanto | 423,908,507 | 421,314,230 | 63,586,276,050 | 62,915,815,836 | 99.39 | 7.09 | 20.97 | 97.72 |
| 8999935525 | BBJ | Kanto | 468,001,195 | 464,327,652 | 70,200,179,250 | 69,327,615,169 | 99.22 | 6.52 | 23.11 | 96.39 |
| 8999935535 | BBJ | Kanto | 496,766,523 | 490,739,217 | 74,514,978,450 | 73,246,396,339 | 98.79 | 4.65 | 24.42 | 98.85 |
| 8999935545 | BBJ | Kanto | 405,299,413 | 403,287,851 | 60,794,911,950 | 60,187,480,766 | 99.50 | 6.12 | 20.06 | 93.97 |
| 8999935555 | BBJ | Kanto | 488,212,013 | 486,267,260 | 73,231,801,950 | 72,621,124,157 | 99.60 | 8.61 | 24.21 | 96.83 |
| 8999935565 | BBJ | Kanto | 426,652,138 | 424,649,884 | 63,997,820,700 | 63,421,745,212 | 99.53 | 5.06 | 21.14 | 95.63 |
| 8999935569 | BBJ | Kanto | 651,625,646 | 646,696,837 | 97,743,846,900 | 96,530,370,116 | 99.24 | 21.53 | 32.18 | 97.83 |
| 8999935575 | BBJ | Kanto | 455,585,256 | 453,152,340 | 68,337,788,400 | 67,679,893,572 | 99.47 | 8.00 | 22.56 | 96.05 |
| 8999935579 | BBJ | Kanto | 484,710,925 | 481,796,282 | 72,706,638,750 | 71,947,607,893 | 99.40 | 5.86 | 23.98 | 96.92 |
| 8999935585 | BBJ | Kanto | 490,146,123 | 488,191,917 | 73,521,918,450 | 72,917,863,221 | 99.60 | 10.65 | 24.31 | 98.71 |
| 8999935589 | BBJ | Kanto | 460,338,786 | 457,971,869 | 69,050,817,900 | 68,396,038,915 | 99.49 | 5.79 | 22.80 | 98.40 |
| 8999935595 | BBJ | Kanto | 530,265,253 | 527,322,624 | 79,539,787,950 | 78,740,299,702 | 99.45 | 11.27 | 26.25 | 98.97 |
| 8999935599 | BBJ | Kanto | 375,084,348 | 373,487,249 | 56,262,652,200 | 55,756,459,857 | 99.57 | 5.88 | 18.59 | 92.30 |
| 8999935605 | BBJ | Kanto | 373,589,438 | 371,313,224 | 56,038,415,700 | 55,422,547,539 | 99.39 | 6.75 | 18.47 | 91.71 |
| 8999935609 | BBJ | Kanto | 397,142,828 | 395,609,063 | 59,571,424,200 | 59,056,629,762 | 99.61 | 6.57 | 19.69 | 93.68 |
| 8999935615 | BBJ | Kanto | 403,975,739 | 401,451,920 | 60,596,360,850 | 59,950,119,672 | 99.38 | 7.18 | 19.98 | 93.67 |
| 8999935619 | BBJ | Kanto | 390,435,249 | 388,744,186 | 58,565,287,350 | 58,047,374,474 | 99.57 | 4.75 | 19.35 | 96.72 |
| 8999935625 | BBJ | Kanto | 410,771,807 | 408,345,506 | 61,615,771,050 | 60,988,639,450 | 99.41 | 4.98 | 20.33 | 97.62 |
| 8999935629 | BBJ | Kanto | 454,494,058 | 453,029,905 | 68,174,108,700 | 67,604,293,035 | 99.68 | 9.66 | 22.53 | 98.17 |
| 8999935639 | BBJ | Kanto | 417,983,218 | 416,481,692 | 62,697,482,700 | 62,147,463,080 | 99.64 | 8.68 | 20.72 | 97.09 |
| 8999935649 | BBJ | Kanto | 430,668,325 | 429,138,542 | 64,600,248,750 | 64,053,883,387 | 99.64 | 7.75 | 21.35 | 95.10 |
| 8999935659 | BBJ | Kanto | 499,847,623 | 497,253,153 | 74,977,143,450 | 74,250,840,952 | 99.48 | 10.04 | 24.75 | 96.87 |
| 8999935669 | BBJ | Kanto | 484,652,059 | 482,231,124 | 72,697,808,850 | 72,005,073,204 | 99.50 | 16.51 | 24.00 | 95.34 |
| 8999935679 | BBJ | Kanto | 468,048,784 | 465,040,917 | 70,207,317,600 | 69,463,218,308 | 99.36 | 12.47 | 23.15 | 98.12 |
| 8999935689 | BBJ | Kanto | 485,247,104 | 482,721,485 | 72,787,065,600 | 72,056,712,176 | 99.48 | 14.42 | 24.02 | 95.86 |
| 8999935699 | BBJ | Kanto | 555,272,531 | 552,343,548 | 83,290,879,650 | 82,473,185,982 | 99.47 | 4.70 | 27.49 | 98.12 |
| 8999935709 | BBJ | Kanto | 515,361,823 | 513,340,471 | 77,304,273,450 | 76,687,573,684 | 99.61 | 8.68 | 25.56 | 98.99 |
| 8999935719 | BBJ | Kanto | 482,483,693 | 480,210,727 | 72,372,553,950 | 71,728,320,068 | 99.53 | 7.18 | 23.91 | 96.94 |
| 8999935729 | BBJ | Kanto | 371,452,472 | 369,060,585 | 55,717,870,800 | 55,127,215,535 | 99.36 | 5.41 | 18.38 | 92.02 |
| 8999935749 | BBJ | Kanto | 490,152,875 | 488,410,026 | 73,522,931,250 | 72,935,942,928 | 99.64 | 7.69 | 24.31 | 98.84 |
| 8999935763 | BBJ | Kanto | 507,463,322 | 504,867,336 | 76,119,498,300 | 75,367,963,680 | 99.49 | 9.77 | 25.12 | 97.08 |
| 8999935773 | BBJ | Kanto | 460,459,592 | 457,838,619 | 69,068,938,800 | 68,359,240,945 | 99.43 | 7.38 | 22.79 | 98.38 |

| | | | | | | | | | | |
|---|---|---|---|---|---|---|---|---|---|---|
| 8999935777 | BBJ | Kanto | 473,706,095 | 467,945,062 | 71,055,914,250 | 69,857,570,567 | 98.78 | 8.62 | 23.29 | 96.18 |
| 8999935783 | BBJ | Kanto | 437,565,786 | 436,144,020 | 65,634,867,900 | 65,108,796,529 | 99.68 | 9.65 | 21.70 | 97.65 |
| 8999935793 | BBJ | Kanto | 489,290,798 | 487,291,640 | 73,393,619,700 | 72,768,361,704 | 99.59 | 9.02 | 24.26 | 96.89 |
| 8999935803 | BBJ | Kanto | 480,820,441 | 479,139,771 | 72,123,066,150 | 71,551,271,032 | 99.65 | 5.44 | 23.85 | 98.88 |
| 8999935901 | BBJ | Kanto | 448,335,531 | 444,755,714 | 67,250,329,650 | 66,364,834,334 | 99.20 | 7.39 | 22.12 | 95.66 |
| 8999936331 | BBJ | Kanto | 420,435,668 | 418,559,851 | 63,065,350,200 | 62,481,277,345 | 99.55 | 6.21 | 20.83 | 94.88 |
| 8999936341 | BBJ | Kanto | 393,355,982 | 391,851,392 | 59,003,397,300 | 58,485,211,010 | 99.62 | 5.71 | 19.50 | 93.63 |
| 8999936361 | BBJ | Kanto | 570,925,642 | 566,330,025 | 85,638,846,300 | 84,557,129,287 | 99.20 | 13.51 | 28.19 | 99.02 |
| 8999936367 | BBJ | Kanto | 370,813,810 | 368,346,365 | 55,622,071,500 | 54,979,031,882 | 99.33 | 6.67 | 18.33 | 91.31 |
| 8999936371 | BBJ | Kanto | 483,405,910 | 481,466,548 | 72,510,886,500 | 71,916,329,621 | 99.60 | 8.03 | 23.97 | 98.77 |
| 8999936377 | BBJ | Kanto | 428,717,984 | 426,048,526 | 64,307,697,600 | 63,637,167,314 | 99.38 | 4.37 | 21.21 | 97.80 |
| 8999936381 | BBJ | Kanto | 375,691,689 | 372,713,715 | 56,353,753,350 | 55,670,612,411 | 99.21 | 5.24 | 18.56 | 92.48 |
| 8999936387 | BBJ | Kanto | 460,468,968 | 457,237,293 | 69,070,345,200 | 68,261,831,013 | 99.30 | 6.68 | 22.75 | 96.04 |
| 8999936397 | BBJ | Kanto | 456,700,312 | 450,584,448 | 68,505,046,800 | 67,270,423,593 | 98.66 | 7.82 | 22.42 | 98.17 |
| 8999936407 | BBJ | Kanto | 458,234,172 | 455,701,371 | 68,735,125,800 | 68,039,767,061 | 99.45 | 11.22 | 22.68 | 95.34 |
| 8999936417 | BBJ | Kanto | 407,839,767 | 405,742,828 | 61,175,965,050 | 60,540,501,797 | 99.49 | 7.11 | 20.18 | 93.57 |
| 8999936427 | BBJ | Kanto | 463,257,632 | 461,580,975 | 69,488,644,800 | 68,929,191,933 | 99.64 | 7.21 | 22.98 | 96.46 |
| 9000063834 | BBJ | Kanto | 505,044,022 | 501,375,689 | 75,756,603,300 | 74,877,697,920 | 99.27 | 8.59 | 24.96 | 98.87 |
| 9000063844 | BBJ | Kanto | 505,015,322 | 502,339,267 | 75,752,298,300 | 75,029,162,873 | 99.47 | 8.97 | 25.01 | 97.17 |
| 9000063864 | BBJ | Kanto | 336,098,084 | 334,695,716 | 50,414,712,600 | 49,970,463,160 | 99.58 | 7.70 | 16.66 | 90.47 |
| 9000063874 | BBJ | Kanto | 375,997,643 | 374,261,425 | 56,399,646,450 | 55,859,178,407 | 99.54 | 6.19 | 18.62 | 92.25 |
| 9000063894 | BBJ | Kanto | 462,904,984 | 460,619,864 | 69,435,747,600 | 68,759,243,445 | 99.51 | 6.95 | 22.92 | 96.26 |
| 9000063904 | BBJ | Kanto | 478,034,842 | 476,102,258 | 71,705,226,300 | 71,043,933,011 | 99.60 | 7.82 | 23.68 | 98.42 |
| 9000063914 | BBJ | Kanto | 369,541,554 | 368,239,783 | 55,431,233,100 | 54,946,760,982 | 99.65 | 8.44 | 18.32 | 93.85 |
| 9000064026 | BBJ | Kanto | 494,071,276 | 492,483,037 | 74,110,691,400 | 73,592,612,999 | 99.68 | 10.96 | 24.53 | 96.79 |
| 9000064036 | BBJ | Kanto | 408,092,225 | 405,881,971 | 61,213,833,750 | 60,600,549,947 | 99.46 | 6.99 | 20.20 | 97.13 |
| 9000064046 | BBJ | Kanto | 376,417,174 | 374,592,167 | 56,462,576,100 | 55,920,783,515 | 99.52 | 6.04 | 18.64 | 92.06 |
| 9000064056 | BBJ | Kanto | 448,995,165 | 447,618,710 | 67,349,274,750 | 66,822,025,650 | 99.69 | 8.41 | 22.27 | 95.83 |
| 9000064066 | BBJ | Kanto | 393,605,497 | 392,295,096 | 59,040,824,550 | 58,586,868,184 | 99.67 | 9.89 | 19.53 | 95.86 |
| 9000064076 | BBJ | Kanto | 454,125,641 | 452,559,204 | 68,118,846,150 | 67,590,197,529 | 99.66 | 6.04 | 22.53 | 98.55 |
| 9000064086 | BBJ | Kanto | 500,364,053 | 498,085,813 | 75,054,607,950 | 74,369,143,166 | 99.54 | 8.82 | 24.79 | 98.72 |
| 9000064090 | BBJ | Kanto | 395,740,934 | 393,041,735 | 59,361,140,100 | 58,697,271,570 | 99.32 | 5.51 | 19.57 | 96.14 |
| 9000064096 | BBJ | Kanto | 390,854,192 | 389,619,194 | 58,628,128,800 | 58,149,928,116 | 99.68 | 8.27 | 19.38 | 93.04 |
| 9000064100 | BBJ | Kanto | 461,234,348 | 459,255,334 | 69,185,152,200 | 68,595,279,435 | 99.57 | 7.82 | 22.87 | 98.44 |
| 9000064106 | BBJ | Kanto | 377,949,980 | 376,492,358 | 56,692,497,000 | 56,213,562,170 | 99.61 | 6.29 | 18.74 | 95.53 |
| 9000064110 | BBJ | Kanto | 434,130,328 | 431,478,721 | 65,119,549,200 | 64,432,354,787 | 99.39 | 10.37 | 21.48 | 97.22 |
| 9000064116 | BBJ | Kanto | 487,262,782 | 484,378,875 | 73,089,417,300 | 72,326,742,218 | 99.41 | 5.76 | 24.11 | 98.73 |
| 9000064120 | BBJ | Kanto | 505,055,259 | 501,705,589 | 75,758,288,850 | 74,913,489,109 | 99.34 | 10.41 | 24.97 | 98.66 |
| 9000064140 | BBJ | Kanto | 409,808,322 | 406,548,680 | 61,471,248,300 | 60,706,256,463 | 99.20 | 7.87 | 20.24 | 96.19 |
| 9000064150 | BBJ | Kanto | 429,041,187 | 426,687,207 | 64,356,178,050 | 63,712,272,599 | 99.45 | 6.43 | 21.24 | 95.15 |
| 9000064160 | BBJ | Kanto | 460,454,550 | 458,805,452 | 69,068,182,500 | 68,486,414,737 | 99.64 | 9.31 | 22.83 | 98.19 |
| 9000064170 | BBJ | Kanto | 444,123,222 | 442,168,858 | 66,618,483,300 | 65,985,412,208 | 99.56 | 4.54 | 22.00 | 98.11 |
| 9000064180 | BBJ | Kanto | 451,699,109 | 448,427,873 | 67,754,866,350 | 66,981,653,023 | 99.28 | 10.26 | 22.33 | 95.47 |
| 9000064190 | BBJ | Kanto | 454,155,698 | 450,975,963 | 68,123,354,700 | 67,331,678,078 | 99.30 | 5.17 | 22.44 | 96.19 |
| 9000064508 | BBJ | Kanto | 439,563,489 | 437,246,451 | 65,934,523,350 | 65,304,702,085 | 99.47 | 5.35 | 21.77 | 95.94 |
| 9000064528 | BBJ | Kanto | 368,070,587 | 366,548,638 | 55,210,588,050 | 54,725,785,737 | 99.59 | 6.73 | 18.24 | 91.45 |
| 9000064538 | BBJ | Kanto | 450,133,458 | 447,929,661 | 67,520,018,700 | 66,902,339,894 | 99.51 | 8.65 | 22.30 | 95.70 |
| 9000064548 | BBJ | Kanto | 512,024,098 | 509,800,718 | 76,803,614,700 | 76,055,921,682 | 99.57 | 9.35 | 25.35 | 98.82 |
| 9000064558 | BBJ | Kanto | 354,794,513 | 353,854,223 | 53,219,176,950 | 52,838,739,052 | 99.73 | 5.33 | 17.61 | 90.91 |
| 9000064568 | BBJ | Kanto | 373,694,542 | 372,199,636 | 56,054,181,300 | 55,583,738,861 | 99.60 | 5.87 | 18.53 | 92.49 |
| 9000065112 | BBJ | Kanto | 411,760,004 | 409,865,050 | 61,764,000,600 | 61,201,724,296 | 99.54 | 6.44 | 20.40 | 94.74 |
| 9000065118 | BBJ | Kanto | 576,872,913 | 573,031,633 | 86,530,936,950 | 85,549,660,501 | 99.33 | 11.82 | 28.52 | 97.84 |
| 9000065122 | BBJ | Kanto | 484,925,429 | 482,553,757 | 72,738,814,350 | 72,040,072,012 | 99.51 | 7.56 | 24.01 | 96.90 |
| 9000065132 | BBJ | Kanto | 410,507,665 | 409,103,194 | 61,576,149,750 | 61,062,798,565 | 99.66 | 8.59 | 20.35 | 94.09 |
| 9000065142 | BBJ | Kanto | 477,518,936 | 473,872,134 | 71,627,840,400 | 70,757,657,679 | 99.24 | 7.48 | 23.59 | 96.53 |
| 9000065152 | BBJ | Kanto | 517,318,152 | 513,498,039 | 77,597,722,800 | 76,657,796,199 | 99.26 | 9.79 | 25.55 | 98.80 |
| 9000065162 | BBJ | Kanto | 424,450,682 | 422,709,993 | 63,667,602,300 | 63,108,093,585 | 99.59 | 8.49 | 21.04 | 97.25 |
| 9999934031 | BBJ | Kanto | 442,484,394 | 439,662,993 | 66,372,659,100 | 65,605,327,180 | 99.36 | 6.14 | 21.87 | 95.65 |
| 9999934095 | BBJ | Kanto | 415,168,688 | 412,356,732 | 62,275,303,200 | 61,593,207,686 | 99.32 | 6.27 | 20.53 | 97.00 |
| 9999934501 | BBJ | Kanto | 491,931,861 | 490,058,359 | 73,789,779,150 | 73,146,872,574 | 99.62 | 11.57 | 24.38 | 98.49 |
| 9999934511 | BBJ | Kanto | 471,415,997 | 467,377,928 | 70,712,399,550 | 69,797,369,642 | 99.14 | 7.15 | 23.27 | 96.51 |
| 9999934521 | BBJ | Kanto | 476,232,861 | 473,139,465 | 71,434,929,150 | 70,572,943,397 | 99.35 | 9.46 | 23.52 | 96.31 |
| 9999934531 | BBJ | Kanto | 514,500,411 | 512,530,815 | 77,175,061,650 | 76,530,479,875 | 99.62 | 7.51 | 25.51 | 98.99 |
| 9999934541 | BBJ | Kanto | 459,130,038 | 456,635,116 | 68,869,505,700 | 68,216,609,310 | 99.46 | 5.63 | 22.74 | 98.60 |
| 9999934551 | BBJ | Kanto | 492,542,905 | 489,656,583 | 73,881,435,750 | 73,134,972,863 | 99.41 | 7.01 | 24.38 | 98.87 |
| 9999934575 | BBJ | Kanto | 432,969,739 | 431,452,058 | 64,945,460,850 | 64,395,264,017 | 99.65 | 7.16 | 21.47 | 95.29 |
| 9999934585 | BBJ | Kanto | 392,375,729 | 391,235,229 | 58,856,359,350 | 58,379,867,368 | 99.71 | 6.31 | 19.46 | 96.40 |
| 9999934595 | BBJ | Kanto | 484,216,175 | 481,563,322 | 72,632,426,250 | 71,803,953,291 | 99.45 | 8.18 | 23.93 | 96.39 |
| 9999934605 | BBJ | Kanto | 400,361,278 | 398,334,978 | 60,054,191,700 | 59,459,729,596 | 99.49 | 11.98 | 19.82 | 92.25 |
| 9999934615 | BBJ | Kanto | 485,925,377 | 482,379,818 | 72,888,806,550 | 72,021,636,653 | 99.27 | 7.87 | 24.01 | 96.56 |
| 9999934625 | BBJ | Kanto | 474,604,951 | 472,245,038 | 71,190,742,650 | 70,516,940,103 | 99.50 | 7.35 | 23.51 | 98.43 |
| 9999934635 | BBJ | Kanto | 376,033,186 | 374,806,996 | 56,404,977,900 | 55,921,732,403 | 99.67 | 7.16 | 18.64 | 95.23 |
| 9999934645 | BBJ | Kanto | 425,915,429 | 423,119,964 | 63,887,314,350 | 63,184,235,674 | 99.34 | 6.17 | 21.06 | 97.32 |
| 9999934655 | BBJ | Kanto | 467,773,568 | 465,785,835 | 70,166,035,200 | 69,574,128,232 | 99.58 | 9.05 | 23.19 | 98.45 |
| 9999934675 | BBJ | Kanto | 409,627,298 | 405,495,360 | 61,444,094,700 | 60,582,342,265 | 98.99 | 4.82 | 20.19 | 94.61 |
| 9999934685 | BBJ | Kanto | 402,598,850 | 401,186,690 | 60,389,827,500 | 59,879,618,330 | 99.65 | 8.00 | 19.96 | 96.67 |
| 9999934695 | BBJ | Kanto | 388,957,503 | 387,003,555 | 58,343,625,450 | 57,786,346,177 | 99.50 | 6.04 | 19.26 | 96.35 |
| 9999934833 | BBJ | Kanto | 495,294,724 | 492,408,375 | 74,294,208,600 | 73,499,246,569 | 99.42 | 6.52 | 24.50 | 97.02 |
| 9999934843 | BBJ | Kanto | 469,918,358 | 466,342,836 | 70,487,753,700 | 69,614,527,910 | 99.24 | 6.71 | 23.20 | 98.58 |
| 9999935283 | BBJ | Kanto | 432,062,230 | 430,544,002 | 64,809,334,500 | 64,239,435,671 | 99.65 | 7.06 | 21.41 | 97.80 |
| 9999935293 | BBJ | Kanto | 480,819,506 | 478,282,660 | 72,122,925,900 | 71,302,328,209 | 99.47 | 5.14 | 23.77 | 96.76 |
| 9999935303 | BBJ | Kanto | 481,302,907 | 478,223,027 | 72,195,436,050 | 71,355,233,424 | 99.36 | 9.77 | 23.79 | 96.11 |
| 9999935307 | BBJ | Kanto | 412,760,150 | 411,635,806 | 61,914,022,500 | 61,399,591,308 | 99.73 | 4.85 | 20.47 | 94.37 |
| 9999935313 | BBJ | Kanto | 450,537,093 | 439,644,611 | 67,580,563,950 | 65,646,264,446 | 97.58 | 2.97 | 21.88 | 96.03 |
| 9999935323 | BBJ | Kanto | 489,890,665 | 485,433,121 | 73,483,599,750 | 72,486,839,996 | 99.09 | 5.45 | 24.16 | 98.75 |
| 9999935327 | BBJ | Kanto | 454,641,890 | 451,965,178 | 68,196,283,500 | 67,491,473,850 | 99.41 | 7.84 | 22.50 | 95.91 |
| 9999935333 | BBJ | Kanto | 459,418,895 | 456,810,593 | 68,912,834,250 | 68,224,834,148 | 99.43 | 7.27 | 22.74 | 98.20 |
| 9999935337 | BBJ | Kanto | 428,901,837 | 427,076,065 | 64,335,275,550 | 63,760,992,683 | 99.57 | 6.82 | 21.25 | 95.10 |
| 9999935343 | BBJ | Kanto | 458,768,576 | 456,406,620 | 68,815,286,400 | 68,177,946,193 | 99.49 | 5.07 | 22.73 | 98.36 |
| 9999935347 | BBJ | Kanto | 388,228,938 | 386,724,623 | 58,234,340,700 | 57,742,228,618 | 99.61 | 5.90 | 19.25 | 96.41 |
| 9999935353 | BBJ | Kanto | 385,981,800 | 384,735,194 | 57,897,270,000 | 57,433,247,422 | 99.68 | 7.49 | 19.14 | 95.78 |
| 9999935357 | BBJ | Kanto | 314,947,455 | 313,715,022 | 47,242,118,250 | 46,841,941,050 | 99.61 | 4.61 | 15.61 | 88.52 |
| 9999935363 | BBJ | Kanto | 417,480,722 | 414,777,153 | 62,622,108,300 | 61,929,517,635 | 99.35 | 6.89 | 20.64 | 94.42 |
| 9999935367 | BBJ | Kanto | 495,889,672 | 493,473,349 | 74,383,450,800 | 73,682,202,843 | 99.51 | 10.80 | 24.56 | 96.75 |
| 9999935373 | BBJ | Kanto | 493,900,784 | 485,579,857 | 74,085,117,600 | 72,464,985,859 | 98.32 | 9.47 | 24.15 | 98.59 |
| 9999935377 | BBJ | Kanto | 443,475,236 | 440,934,552 | 66,521,285,400 | 65,829,957,126 | 99.43 | 8.06 | 21.94 | 98.00 |
| 9999935383 | BBJ | Kanto | 457,773,883 | 455,852,280 | 68,666,082,450 | 68,090,206,540 | 99.58 | 7.64 | 22.70 | 98.40 |
| 9999935387 | BBJ | Kanto | 448,910,202 | 447,094,000 | 67,336,530,300 | 66,735,479,065 | 99.60 | 6.40 | 22.25 | 95.93 |
| 9999935403 | BBJ | Kanto | 432,660,498 | 427,442,397 | 64,899,074,700 | 63,860,312,195 | 98.79 | 4.44 | 21.29 | 98.00 |
| 9999935413 | BBJ | Kanto | 483,960,690 | 481,578,564 | 72,594,103,500 | 71,909,905,466 | 99.51 | 6.60 | 23.97 | 96.60 |

| | | | | | | | | | | |
|---|---|---|---|---|---|---|---|---|---|---|
| 9999935789 | BBJ | Kanto | 445,197,207 | 441,588,776 | 66,779,581,050 | 65,907,497,379 | 99.19 | 7.23 | 21.97 | 97.91 |
| 9999936289 | BBJ | Kanto | 366,768,575 | 365,509,195 | 55,015,286,250 | 54,555,937,900 | 99.66 | 5.82 | 18.19 | 91.69 |
| 9999936299 | BBJ | Kanto | 453,706,772 | 452,090,453 | 68,056,015,800 | 67,475,339,843 | 99.64 | 7.62 | 22.49 | 96.04 |
| 9999936309 | BBJ | Kanto | 399,736,015 | 397,923,708 | 59,960,402,250 | 59,413,915,240 | 99.55 | 6.64 | 19.80 | 96.56 |
| 9999936319 | BBJ | Kanto | 283,277,316 | 281,447,413 | 42,491,597,400 | 42,021,989,378 | 99.35 | 4.51 | 14.01 | 80.55 |
| 9999936329 | BBJ | Kanto | 471,571,065 | 469,530,824 | 70,735,659,750 | 70,118,117,545 | 99.57 | 8.29 | 23.37 | 96.37 |
| 9999936339 | BBJ | Kanto | 506,789,419 | 504,691,307 | 76,018,412,850 | 75,364,162,626 | 99.59 | 11.50 | 25.12 | 98.84 |
| 9999936349 | BBJ | Kanto | 372,996,365 | 370,662,005 | 55,949,454,750 | 55,373,815,253 | 99.37 | 6.08 | 18.46 | 91.85 |
| 9999936359 | BBJ | Kanto | 364,371,131 | 362,969,120 | 54,655,669,650 | 54,195,236,964 | 99.62 | 6.14 | 18.07 | 91.56 |
| 9999936369 | BBJ | Kanto | 369,792,916 | 368,381,526 | 55,468,937,400 | 54,984,619,910 | 99.62 | 6.01 | 18.33 | 94.43 |
| 9999936379 | BBJ | Kanto | 479,806,066 | 477,284,863 | 71,970,909,900 | 71,295,822,552 | 99.47 | 6.81 | 23.77 | 96.76 |
| AYUM0007 | St. Marianna Univ. | Kyushu | 356,230,552 | 354,954,240 | 53,434,582,800 | 53,356,294,628 | 99.64 | 7.82 | 17.79 | 92.57 |
| AYUM0008 | St. Marianna Univ. | Kyushu | 378,374,078 | 377,098,725 | 56,756,111,700 | 56,655,055,761 | 99.66 | 8.34 | 18.89 | 95.32 |
| AYUM0018 | St. Marianna Univ. | Kyushu | 350,659,880 | 348,350,587 | 52,598,982,000 | 52,346,340,762 | 99.34 | 5.84 | 17.45 | 93.03 |
| AYUM0024 | St. Marianna Univ. | Kyushu | 317,661,742 | 316,512,965 | 47,649,261,300 | 47,594,517,031 | 99.64 | 8.17 | 15.86 | 82.07 |
| AYUM0025 | St. Marianna Univ. | Kyushu | 325,994,909 | 324,787,023 | 48,899,236,350 | 48,818,990,095 | 99.63 | 8.15 | 16.27 | 88.93 |
| AYUM0030 | St. Marianna Univ. | Kyushu | 391,983,804 | 390,621,889 | 58,797,570,600 | 58,713,192,750 | 99.65 | 8.33 | 19.57 | 93.39 |
| AYUM0031 | St. Marianna Univ. | Kyushu | 349,540,010 | 348,349,751 | 52,431,001,500 | 52,352,414,127 | 99.66 | 7.53 | 17.45 | 89.84 |
| AYUM0034 | St. Marianna Univ. | Kyushu | 361,493,914 | 360,356,518 | 54,224,087,100 | 54,159,044,387 | 99.69 | 8.16 | 18.05 | 91.05 |
| AYUM0040 | St. Marianna Univ. | Kyushu | 346,390,643 | 344,431,865 | 51,958,596,450 | 51,693,815,476 | 99.43 | 6.02 | 17.23 | 91.49 |
| AYUM0043 | St. Marianna Univ. | Kyushu | 347,286,803 | 345,092,666 | 52,093,020,450 | 51,862,904,003 | 99.37 | 5.63 | 17.29 | 92.79 |
| AYUM0047 | St. Marianna Univ. | Kyushu | 355,116,786 | 353,805,088 | 53,267,517,900 | 53,202,045,533 | 99.63 | 6.45 | 17.73 | 94.56 |
| AYUM0054 | St. Marianna Univ. | Kyushu | 396,544,324 | 395,026,402 | 59,481,648,600 | 59,390,275,337 | 99.62 | 7.16 | 19.80 | 97.17 |
| AYUM0055 | St. Marianna Univ. | Kyushu | 373,928,435 | 372,608,359 | 56,089,265,250 | 56,027,823,545 | 99.65 | 7.77 | 18.68 | 94.13 |
| AYUM0057 | St. Marianna Univ. | Kyushu | 322,847,104 | 320,400,647 | 48,427,065,600 | 48,155,405,354 | 99.24 | 4.51 | 16.05 | 89.40 |
| AYUM0061 | St. Marianna Univ. | Kyushu | 368,856,436 | 367,511,055 | 55,328,465,400 | 55,224,762,251 | 99.64 | 6.55 | 18.41 | 95.49 |
| AYUM0063 | St. Marianna Univ. | Kyushu | 357,688,188 | 356,164,893 | 53,653,228,200 | 53,544,066,449 | 99.57 | 6.90 | 17.85 | 91.30 |
| AYUM0066 | St. Marianna Univ. | Kyushu | 397,983,617 | 396,296,127 | 59,697,542,550 | 59,600,533,650 | 99.58 | 6.61 | 19.87 | 97.30 |
| AYUM0076 | St. Marianna Univ. | Kyushu | 395,672,296 | 394,185,600 | 59,350,844,400 | 59,260,218,573 | 99.62 | 6.20 | 19.75 | 97.26 |
| AYUM0079 | St. Marianna Univ. | Kyushu | 357,142,582 | 355,498,315 | 53,571,387,300 | 53,435,781,107 | 99.54 | 6.63 | 17.81 | 91.23 |
| AYUM0084 | St. Marianna Univ. | Kyushu | 402,724,453 | 401,081,441 | 60,408,667,950 | 60,264,716,170 | 99.59 | 7.26 | 20.09 | 97.27 |
| AYUM0092 | St. Marianna Univ. | Kyushu | 362,557,188 | 361,185,075 | 54,383,578,200 | 54,295,404,738 | 99.62 | 6.90 | 18.10 | 91.76 |
| AYUM0093 | St. Marianna Univ. | Kyushu | 437,939,701 | 436,496,299 | 65,690,955,150 | 65,593,284,432 | 99.67 | 8.24 | 21.86 | 98.37 |
| AYUM0101 | St. Marianna Univ. | Kyushu | 400,180,301 | 398,541,149 | 60,027,045,150 | 59,894,189,789 | 99.59 | 7.67 | 19.96 | 97.17 |
| AYUM0110 | St. Marianna Univ. | Kyushu | 433,099,887 | 431,318,187 | 64,964,983,050 | 64,812,178,951 | 99.59 | 8.37 | 21.60 | 98.26 |
| AYUM0118 | St. Marianna Univ. | Kyushu | 466,219,125 | 464,487,036 | 69,932,868,750 | 69,815,795,430 | 99.63 | 9.09 | 23.27 | 98.80 |
| AYUM0119 | St. Marianna Univ. | Kyushu | 442,908,603 | 441,480,022 | 66,436,290,450 | 66,351,559,222 | 99.68 | 7.93 | 22.12 | 96.12 |
| AYUM0123 | St. Marianna Univ. | Kyushu | 413,379,061 | 411,824,806 | 62,006,859,150 | 61,865,677,386 | 99.62 | 8.11 | 20.62 | 97.67 |
| AYUM0156 | St. Marianna Univ. | Kyushu | 325,970,809 | 323,873,585 | 48,895,621,350 | 48,673,695,269 | 99.36 | 4.86 | 16.22 | 90.02 |
| AYUM0159 | St. Marianna Univ. | Kyushu | 438,286,969 | 436,630,426 | 65,743,045,350 | 65,609,260,481 | 99.62 | 8.95 | 21.87 | 95.59 |
| AYUM0160 | St. Marianna Univ. | Kyushu | 347,919,842 | 345,468,780 | 52,187,976,300 | 51,954,488,939 | 99.30 | 5.41 | 17.32 | 92.92 |
| AYUM0165 | St. Marianna Univ. | Kyushu | 320,977,998 | 318,627,945 | 48,146,699,700 | 47,871,249,976 | 99.27 | 5.41 | 15.96 | 88.98 |
| AYUM0166 | St. Marianna Univ. | Kyushu | 384,794,544 | 383,045,187 | 57,719,181,600 | 57,530,816,686 | 99.55 | 6.80 | 19.18 | 92.91 |
| AYUM0171 | St. Marianna Univ. | Kyushu | 331,695,830 | 329,433,368 | 49,754,374,500 | 49,432,849,546 | 99.32 | 5.07 | 16.48 | 85.43 |
| AYUM0198 | St. Marianna Univ. | Kyushu | 331,145,191 | 329,162,285 | 49,671,778,650 | 49,475,152,232 | 99.40 | 4.85 | 16.49 | 88.04 |
| AYUM0207 | St. Marianna Univ. | Kyushu | 373,427,501 | 370,670,211 | 56,014,125,150 | 55,701,529,175 | 99.26 | 4.93 | 18.57 | 95.31 |
| AYUM0219 | St. Marianna Univ. | Kyushu | 325,875,673 | 324,003,961 | 48,881,350,950 | 48,709,388,863 | 99.43 | 4.59 | 16.24 | 87.80 |
| AYUM0226 | St. Marianna Univ. | Kyushu | 328,916,122 | 327,297,202 | 49,337,418,300 | 49,179,658,634 | 99.51 | 4.78 | 16.39 | 91.11 |
| AYUM0232 | St. Marianna Univ. | Kyushu | 409,949,871 | 407,805,923 | 61,492,480,650 | 61,102,718,289 | 99.48 | 5.72 | 20.37 | 93.76 |
| AYUM0235 | St. Marianna Univ. | Kyushu | 362,540,333 | 360,398,232 | 54,381,049,950 | 54,158,861,035 | 99.41 | 4.72 | 18.05 | 94.93 |
| AYUM0238 | St. Marianna Univ. | Kyushu | 357,452,822 | 355,813,042 | 53,617,923,300 | 53,469,006,766 | 99.54 | 6.46 | 17.82 | 93.99 |
| AYUM0243 | St. Marianna Univ. | Kyushu | 340,756,523 | 338,956,217 | 51,113,478,450 | 50,922,614,580 | 99.47 | 4.51 | 16.97 | 89.51 |
| AYUM0244 | St. Marianna Univ. | Kyushu | 321,486,415 | 319,155,222 | 48,222,962,250 | 47,964,159,616 | 99.27 | 4.09 | 15.99 | 89.86 |
| AYUM0250 | St. Marianna Univ. | Kyushu | 395,978,721 | 393,074,371 | 59,396,808,150 | 59,093,014,942 | 99.27 | 6.08 | 19.70 | 96.32 |
| AYUM0254 | St. Marianna Univ. | Kyushu | 382,937,640 | 380,025,192 | 57,440,646,000 | 57,107,193,177 | 99.24 | 6.41 | 19.04 | 92.36 |
| AYUM0258 | St. Marianna Univ. | Kyushu | 330,670,542 | 328,593,965 | 49,600,581,300 | 49,369,489,044 | 99.37 | 5.70 | 16.46 | 87.37 |
| AYUM0264 | St. Marianna Univ. | Kyushu | 337,080,480 | 335,563,731 | 50,562,072,000 | 50,467,786,586 | 99.55 | 6.26 | 16.82 | 91.75 |
| AYUM0270 | St. Marianna Univ. | Kyushu | 307,624,168 | 305,744,851 | 46,143,625,200 | 45,994,686,592 | 99.39 | 4.01 | 15.33 | 87.71 |
| AYUM0277 | St. Marianna Univ. | Kyushu | 337,042,268 | 334,142,786 | 50,556,340,200 | 50,259,707,955 | 99.14 | 3.39 | 16.75 | 92.65 |
| AYUM0280 | St. Marianna Univ. | Kyushu | 339,701,482 | 336,854,867 | 50,955,222,300 | 50,631,793,012 | 99.16 | 3.82 | 16.88 | 92.39 |
| AYUM0281 | St. Marianna Univ. | Kyushu | 333,267,053 | 331,172,079 | 49,990,057,950 | 49,749,894,167 | 99.37 | 4.07 | 16.58 | 91.69 |
| AYUM0285 | St. Marianna Univ. | Kyushu | 331,066,311 | 328,133,233 | 49,659,946,650 | 49,318,079,759 | 99.11 | 3.24 | 16.44 | 89.66 |
| AYUM0300 | St. Marianna Univ. | Kyushu | 384,521,620 | 382,602,192 | 57,678,243,000 | 57,528,907,176 | 99.50 | 7.10 | 19.18 | 96.05 |
| AYUM0312 | St. Marianna Univ. | Kyushu | 323,217,993 | 321,212,092 | 48,482,698,950 | 48,284,968,248 | 99.38 | 6.09 | 16.09 | 87.58 |
| AYUM0321 | St. Marianna Univ. | Kyushu | 404,283,878 | 401,916,351 | 60,642,581,700 | 60,429,169,432 | 99.41 | 6.65 | 20.14 | 93.96 |
| AYUM0323 | St. Marianna Univ. | Kyushu | 343,272,610 | 341,506,848 | 51,490,891,500 | 51,346,958,426 | 99.49 | 6.62 | 17.12 | 92.61 |
| AYUM0325 | St. Marianna Univ. | Kyushu | 335,042,810 | 331,936,155 | 50,256,421,500 | 49,880,327,888 | 99.07 | 3.68 | 16.63 | 91.48 |
| AYUM0326 | St. Marianna Univ. | Kyushu | 359,570,710 | 356,651,084 | 53,935,606,500 | 53,578,548,248 | 99.19 | 3.01 | 17.86 | 94.31 |
| AYUM0328 | St. Marianna Univ. | Kyushu | 317,185,703 | 314,428,277 | 47,577,855,450 | 47,269,183,157 | 99.13 | 3.47 | 15.76 | 89.25 |
| AYUM0331 | St. Marianna Univ. | Kyushu | 364,324,440 | 361,981,844 | 54,648,666,000 | 54,429,757,707 | 99.36 | 5.67 | 18.14 | 94.49 |
| AYUM0333 | St. Marianna Univ. | Kyushu | 361,151,719 | 358,175,762 | 54,172,757,850 | 53,858,046,990 | 99.18 | 5.78 | 17.95 | 92.81 |
| AYUM0334 | St. Marianna Univ. | Kyushu | 332,247,529 | 329,461,132 | 49,837,129,350 | 49,517,922,410 | 99.16 | 4.25 | 16.51 | 90.95 |
| AYUM0358 | St. Marianna Univ. | Kyushu | 334,157,592 | 331,409,538 | 50,123,638,800 | 49,814,407,944 | 99.18 | 4.16 | 16.60 | 91.56 |
| AYUM0360 | St. Marianna Univ. | Kyushu | 353,113,717 | 351,625,633 | 52,967,057,550 | 52,851,138,632 | 99.58 | 6.79 | 17.62 | 93.98 |
| AYUM0362 | St. Marianna Univ. | Kyushu | 402,001,306 | 400,039,845 | 60,300,195,900 | 60,148,299,724 | 99.51 | 6.62 | 20.05 | 97.21 |
| AYUM0363 | St. Marianna Univ. | Kyushu | 369,286,980 | 367,355,920 | 55,393,047,000 | 55,215,897,501 | 99.48 | 6.41 | 18.41 | 92.23 |
| AYUM0374 | St. Marianna Univ. | Kyushu | 346,041,700 | 344,061,435 | 51,906,255,000 | 51,715,770,146 | 99.43 | 6.08 | 17.24 | 93.14 |
| AYUM0384 | St. Marianna Univ. | Kyushu | 365,895,132 | 363,169,807 | 54,884,269,800 | 54,582,915,421 | 99.26 | 5.96 | 18.19 | 90.94 |
| AYUM0400 | St. Marianna Univ. | Kyushu | 327,141,977 | 325,305,306 | 49,071,296,550 | 48,899,118,718 | 99.44 | 5.15 | 16.30 | 87.64 |
| AYUM0405 | St. Marianna Univ. | Kyushu | 361,991,599 | 359,790,497 | 54,298,739,850 | 54,078,556,995 | 99.39 | 5.37 | 18.03 | 94.71 |
| AYUM0406 | St. Marianna Univ. | Kyushu | 376,015,856 | 373,132,678 | 56,402,378,400 | 56,071,976,583 | 99.23 | 5.52 | 18.69 | 91.62 |
| AYUM0414 | St. Marianna Univ. | Kyushu | 364,885,201 | 362,694,316 | 54,732,780,150 | 54,515,886,530 | 99.40 | 6.84 | 18.17 | 94.63 |
| AYUM0437 | St. Marianna Univ. | Kyushu | 381,740,339 | 379,651,358 | 57,261,050,850 | 57,051,347,291 | 99.45 | 5.35 | 19.02 | 96.05 |
| AYUM0450 | St. Marianna Univ. | Kyushu | 365,165,679 | 362,634,175 | 54,774,851,850 | 54,518,740,045 | 99.31 | 6.09 | 18.17 | 93.72 |
| AYUM0455 | St. Marianna Univ. | Kyushu | 394,035,164 | 392,500,448 | 59,105,274,600 | 58,984,917,813 | 99.61 | 5.79 | 19.66 | 97.09 |
| AYUM0460 | St. Marianna Univ. | Kyushu | 347,471,039 | 345,541,910 | 52,120,655,850 | 51,930,306,511 | 99.44 | 5.71 | 17.31 | 93.10 |
| AYUM0467 | St. Marianna Univ. | Kyushu | 359,440,132 | 357,525,658 | 53,916,019,800 | 53,733,226,479 | 99.47 | 5.65 | 17.91 | 94.02 |
| AYUM0484 | St. Marianna Univ. | Kyushu | 349,416,521 | 347,411,197 | 52,412,478,150 | 52,195,996,449 | 99.43 | 5.22 | 17.40 | 93.68 |
| AYUM0488 | St. Marianna Univ. | Kyushu | 405,735,091 | 403,380,152 | 60,860,263,650 | 60,577,244,914 | 99.42 | 5.09 | 20.19 | 97.25 |
| AYUM0519 | St. Marianna Univ. | Kyushu | 427,275,998 | 424,973,660 | 64,091,399,700 | 63,807,478,728 | 99.46 | 4.77 | 21.27 | 97.86 |
| AYUM0549 | St. Marianna Univ. | Kyushu | 335,961,283 | 333,778,593 | 50,394,192,450 | 50,188,422,731 | 99.35 | 4.54 | 16.73 | 91.10 |
| AYUM0552 | St. Marianna Univ. | Kyushu | 381,209,356 | 378,420,400 | 57,181,403,400 | 56,860,584,459 | 99.27 | 5.00 | 18.95 | 95.30 |
| AYUM0560 | St. Marianna Univ. | Kyushu | 331,279,330 | 329,330,381 | 49,691,899,500 | 49,486,389,814 | 99.41 | 5.16 | 16.50 | 91.33 |
| AYUM0574 | St. Marianna Univ. | Kyushu | 422,553,829 | 419,251,704 | 63,383,074,350 | 63,000,934,300 | 99.22 | 5.23 | 21.00 | 97.67 |
| AYUM0579 | St. Marianna Univ. | Kyushu | 387,265,463 | 384,586,616 | 58,089,819,450 | 57,804,724,634 | 99.31 | 4.71 | 19.27 | 93.15 |
| AYUM0580 | St. Marianna Univ. | Kyushu | 340,119,435 | 337,612,188 | 51,017,915,250 | 50,747,761,496 | 99.26 | 4.46 | 16.92 | 91.90 |
| AYUM0586 | St. Marianna Univ. | Kyushu | 376,829,689 | 374,631,212 | 56,524,453,350 | 56,309,487,831 | 99.42 | 5.05 | 18.77 | 95.84 |
| AYUM0588 | St. Marianna Univ. | Kyushu | 339,216,952 | 337,080,820 | 50,882,542,800 | 50,658,545,565 | 99.37 | 4.54 | 16.89 | 88.73 |
| AYUM0593 | St. Marianna Univ. | Kyushu | 341,479,416 | 339,215,706 | 51,221,912,400 | 50,962,374,214 | 99.34 | 4.50 | 16.99 | 91.81 |
| AYUM0597 | St. Marianna Univ. | Kyushu | 368,687,311 | 366,393,042 | 55,303,096,650 | 55,045,273,655 | 99.38 | 4.88 | 18.35 | 95.17 |

| | | | | | | | | | | |
|---|---|---|---|---|---|---|---|---|---|---|
| AYUM0614 | St. Marianna Univ. | Kyushu | 342,027,586 | 340,167,374 | 51,304,137,900 | 51,118,354,028 | 99.46 | 4.86 | 17.04 | 92.63 |
| AYUM0615 | St. Marianna Univ. | Kyushu | 436,010,057 | 433,273,448 | 65,401,508,550 | 65,120,105,664 | 99.37 | 7.87 | 21.71 | 97.77 |
| AYUM0616 | St. Marianna Univ. | Kyushu | 326,488,951 | 324,207,079 | 48,973,342,650 | 48,709,385,636 | 99.30 | 6.93 | 16.24 | 84.77 |
| AYUM0621 | St. Marianna Univ. | Kyushu | 383,711,052 | 381,241,504 | 57,556,657,800 | 57,297,674,521 | 99.36 | 7.85 | 19.10 | 95.35 |
| AYUM0624 | St. Marianna Univ. | Kyushu | 355,337,381 | 352,655,765 | 53,300,607,150 | 52,995,466,574 | 99.25 | 7.59 | 17.67 | 89.12 |
| AYUM0626 | St. Marianna Univ. | Kyushu | 357,289,330 | 354,896,238 | 53,593,399,500 | 53,341,561,504 | 99.33 | 7.30 | 17.78 | 92.94 |
| AYUM0637 | St. Marianna Univ. | Kyushu | 349,812,285 | 347,540,855 | 52,471,842,750 | 52,215,088,910 | 99.35 | 7.42 | 17.41 | 91.89 |
| AYUM0638 | St. Marianna Univ. | Kyushu | 424,844,402 | 422,641,122 | 63,726,660,300 | 63,525,453,533 | 99.48 | 8.85 | 21.18 | 96.53 |
| AYUM0640 | St. Marianna Univ. | Kyushu | 365,055,652 | 362,849,842 | 54,758,347,800 | 54,528,591,697 | 99.40 | 7.47 | 18.18 | 93.51 |
| AYUM0643 | St. Marianna Univ. | Kyushu | 388,096,159 | 385,601,761 | 58,214,423,850 | 57,911,388,497 | 99.36 | 7.86 | 19.30 | 94.95 |
| AYUM0648 | St. Marianna Univ. | Kyushu | 440,083,638 | 437,250,576 | 66,012,545,700 | 65,637,144,429 | 99.36 | 8.06 | 21.88 | 94.78 |
| AYUM0655 | St. Marianna Univ. | Kyushu | 380,361,665 | 378,640,847 | 57,054,249,750 | 56,897,011,374 | 99.55 | 5.38 | 18.97 | 92.19 |
| AYUM0668 | St. Marianna Univ. | Kyushu | 346,991,843 | 344,567,995 | 52,048,776,450 | 51,798,345,490 | 99.30 | 6.71 | 17.27 | 91.64 |
| AYUM0672 | St. Marianna Univ. | Kyushu | 379,513,834 | 376,633,244 | 56,927,075,100 | 56,608,301,653 | 99.24 | 6.71 | 18.87 | 94.88 |
| AYUM0709 | St. Marianna Univ. | Kyushu | 377,830,682 | 375,245,973 | 56,674,602,300 | 56,389,959,422 | 99.32 | 6.79 | 18.80 | 94.73 |
| AYUM0715 | St. Marianna Univ. | Kyushu | 433,778,723 | 430,554,917 | 65,066,808,450 | 64,714,966,873 | 99.26 | 7.58 | 21.57 | 95.02 |
| AYUM0726 | St. Marianna Univ. | Kyushu | 363,975,331 | 361,299,539 | 54,596,299,650 | 54,303,761,266 | 99.26 | 7.50 | 18.10 | 89.81 |
| AYUM0727 | St. Marianna Univ. | Kyushu | 400,043,126 | 397,746,841 | 60,006,468,900 | 59,778,630,989 | 99.43 | 7.67 | 19.93 | 96.17 |
| AYUM0728 | St. Marianna Univ. | Kyushu | 365,221,591 | 363,028,518 | 54,783,238,650 | 54,593,173,330 | 99.40 | 7.44 | 18.20 | 93.90 |
| AYUM0744 | St. Marianna Univ. | Kyushu | 372,347,429 | 370,056,261 | 55,852,114,350 | 55,622,693,809 | 99.38 | 7.68 | 18.54 | 91.35 |
| AYUM1006 | St. Marianna Univ. | Kyushu | 357,203,540 | 355,490,100 | 53,580,531,000 | 53,490,050,855 | 99.52 | 4.68 | 17.83 | 94.23 |
| AYUM1007 | St. Marianna Univ. | Kyushu | 384,244,503 | 382,272,485 | 57,636,675,450 | 57,489,279,798 | 99.49 | 5.14 | 19.16 | 96.64 |
| AYUM1010 | St. Marianna Univ. | Kyushu | 361,399,802 | 359,326,549 | 54,209,970,300 | 54,007,621,267 | 99.43 | 5.82 | 18.00 | 91.53 |
| AYUM1014 | St. Marianna Univ. | Kyushu | 356,980,646 | 355,142,974 | 53,547,096,900 | 53,404,519,947 | 99.49 | 5.50 | 17.80 | 94.69 |
| AYUM1018 | St. Marianna Univ. | Kyushu | 368,434,712 | 366,497,777 | 55,265,206,800 | 55,087,872,810 | 99.47 | 5.52 | 18.36 | 95.56 |
| AYUM5017 | St. Marianna Univ. | Kyushu | 397,386,147 | 394,844,065 | 59,607,922,050 | 59,338,623,586 | 99.36 | 7.13 | 19.78 | 93.18 |
| AYUM5018 | St. Marianna Univ. | Kyushu | 372,495,606 | 369,753,840 | 55,874,340,900 | 55,560,765,555 | 99.26 | 7.00 | 18.52 | 93.79 |
| AYUM5019 | St. Marianna Univ. | Kyushu | 415,505,785 | 413,333,040 | 62,325,867,750 | 62,131,246,295 | 99.48 | 6.89 | 20.71 | 97.19 |
| AYUM5020 | St. Marianna Univ. | Kyushu | 337,200,900 | 335,386,311 | 50,580,135,000 | 50,398,707,824 | 99.46 | 6.62 | 16.80 | 90.43 |
| AYUM5024 | St. Marianna Univ. | Kyushu | 373,841,304 | 371,800,592 | 56,076,195,600 | 55,875,516,885 | 99.45 | 6.59 | 18.63 | 91.52 |
| AYUM5025 | St. Marianna Univ. | Kyushu | 342,615,106 | 340,850,379 | 51,392,265,900 | 51,220,740,878 | 99.48 | 6.44 | 17.07 | 87.83 |
| AYUM5026 | St. Marianna Univ. | Kyushu | 400,148,946 | 397,983,397 | 60,022,341,900 | 59,808,007,463 | 99.46 | 7.31 | 19.94 | 96.37 |
| AYUM5030 | St. Marianna Univ. | Kyushu | 403,611,053 | 401,494,869 | 60,541,657,950 | 60,347,821,758 | 99.48 | 7.21 | 20.12 | 96.58 |
| AYUM5035 | St. Marianna Univ. | Kyushu | 377,209,284 | 375,356,400 | 56,581,392,600 | 56,413,409,756 | 99.51 | 7.22 | 18.80 | 91.89 |
| AYUM5037 | St. Marianna Univ. | Kyushu | 388,105,719 | 385,976,028 | 58,215,857,850 | 58,016,272,355 | 99.45 | 6.78 | 19.34 | 92.83 |
| AYUM5038 | St. Marianna Univ. | Kyushu | 395,682,112 | 393,697,190 | 59,352,316,800 | 59,166,041,154 | 99.50 | 7.42 | 19.72 | 96.10 |
| AYUM5039 | St. Marianna Univ. | Kyushu | 411,214,718 | 409,279,995 | 61,682,207,700 | 61,529,933,312 | 99.53 | 7.07 | 20.51 | 97.08 |
| AYUM5040 | St. Marianna Univ. | Kyushu | 360,601,757 | 358,411,905 | 54,090,263,550 | 53,867,847,233 | 99.39 | 6.16 | 17.96 | 90.05 |
| AYUM5043 | St. Marianna Univ. | Kyushu | 387,500,574 | 385,487,481 | 58,125,086,100 | 57,943,822,849 | 99.48 | 7.23 | 19.31 | 95.79 |
| AYUM5044 | St. Marianna Univ. | Kyushu | 365,595,944 | 363,766,872 | 54,839,391,600 | 54,700,064,262 | 99.50 | 6.10 | 18.23 | 95.27 |
| AYUM5045 | St. Marianna Univ. | Kyushu | 390,862,519 | 388,713,302 | 58,629,377,850 | 58,427,624,959 | 99.45 | 6.49 | 19.48 | 95.94 |
| AYUM5046 | St. Marianna Univ. | Kyushu | 394,027,261 | 391,869,775 | 59,104,089,150 | 58,895,392,831 | 99.45 | 6.49 | 19.63 | 96.40 |
| AYUM5101 | St. Marianna Univ. | Kyushu | 365,987,773 | 363,812,653 | 54,898,165,950 | 54,692,114,704 | 99.41 | 5.77 | 18.23 | 90.58 |
| AYUM5102 | St. Marianna Univ. | Kyushu | 409,241,151 | 406,720,845 | 61,386,172,650 | 61,120,869,016 | 99.38 | 6.57 | 20.37 | 96.59 |
| AYUM5103 | St. Marianna Univ. | Kyushu | 404,299,973 | 401,827,780 | 60,644,995,950 | 60,394,065,640 | 99.39 | 6.76 | 20.13 | 93.65 |
| AYUM5104 | St. Marianna Univ. | Kyushu | 422,816,980 | 420,391,348 | 63,422,547,000 | 63,186,992,479 | 99.43 | 6.40 | 21.06 | 97.34 |
| AYUM5105 | St. Marianna Univ. | Kyushu | 344,391,505 | 342,429,487 | 51,658,725,750 | 51,484,957,924 | 99.43 | 6.55 | 17.16 | 88.76 |
| AYUM5111 | St. Marianna Univ. | Kyushu | 328,055,632 | 326,366,163 | 49,208,344,800 | 49,067,479,926 | 99.49 | 7.07 | 16.36 | 88.65 |
| AYUM5113 | St. Marianna Univ. | Kyushu | 346,096,257 | 344,459,201 | 51,914,438,550 | 51,794,962,598 | 99.53 | 6.02 | 17.26 | 93.42 |
| AYUM5117 | St. Marianna Univ. | Kyushu | 365,094,685 | 362,579,123 | 54,764,202,750 | 54,507,007,830 | 99.31 | 6.78 | 18.17 | 89.68 |
| AYUM5118 | St. Marianna Univ. | Kyushu | 339,984,656 | 338,177,757 | 50,997,698,400 | 50,844,649,712 | 99.47 | 5.95 | 16.95 | 92.58 |
| AYUM5119 | St. Marianna Univ. | Kyushu | 395,826,710 | 393,157,001 | 59,374,006,500 | 59,104,269,001 | 99.33 | 7.30 | 19.70 | 95.32 |
| AYUM5121 | St. Marianna Univ. | Kyushu | 421,130,450 | 418,362,712 | 63,169,567,500 | 62,891,879,323 | 99.34 | 7.49 | 20.96 | 96.73 |
| AYUM5123 | St. Marianna Univ. | Kyushu | 404,414,693 | 401,539,038 | 60,662,203,950 | 60,369,201,410 | 99.29 | 6.95 | 20.12 | 96.32 |
| AYUM5124 | St. Marianna Univ. | Kyushu | 374,180,630 | 371,569,204 | 56,127,094,500 | 55,842,084,928 | 99.30 | 7.22 | 18.61 | 93.68 |
| AYUM5127 | St. Marianna Univ. | Kyushu | 422,880,351 | 419,979,324 | 63,432,052,650 | 63,118,140,021 | 99.31 | 6.39 | 21.04 | 96.99 |
| AYUM5128 | St. Marianna Univ. | Kyushu | 353,668,716 | 351,356,877 | 53,050,307,400 | 52,808,164,343 | 99.35 | 7.62 | 17.60 | 87.90 |
| AYUM5129 | St. Marianna Univ. | Kyushu | 372,859,290 | 370,346,316 | 55,928,893,500 | 55,667,705,968 | 99.33 | 7.88 | 18.56 | 93.45 |
| AYUM5130 | St. Marianna Univ. | Kyushu | 361,523,601 | 358,973,091 | 54,228,540,150 | 53,949,685,231 | 99.29 | 6.87 | 17.98 | 92.48 |
| AYUM5131 | St. Marianna Univ. | Kyushu | 338,519,653 | 336,121,326 | 50,777,947,950 | 50,522,960,139 | 99.29 | 7.11 | 16.84 | 86.86 |
| AYUM5132 | St. Marianna Univ. | Kyushu | 416,685,692 | 414,272,750 | 62,502,853,800 | 62,245,552,231 | 99.42 | 5.66 | 20.75 | 96.44 |
| AYUM5133 | St. Marianna Univ. | Kyushu | 394,733,172 | 392,678,562 | 59,209,975,800 | 59,014,508,969 | 99.48 | 6.75 | 19.67 | 93.82 |
| AYUM5134 | St. Marianna Univ. | Kyushu | 384,123,420 | 382,943,863 | 57,618,513,000 | 57,577,346,845 | 99.69 | 7.30 | 19.19 | 96.50 |
| AYUM5139 | St. Marianna Univ. | Kyushu | 352,045,893 | 350,268,335 | 52,806,883,950 | 52,635,538,460 | 99.50 | 6.45 | 17.55 | 93.48 |
| AYUM5140 | St. Marianna Univ. | Kyushu | 340,393,527 | 338,930,015 | 51,059,029,050 | 50,918,535,249 | 99.57 | 5.21 | 16.97 | 92.57 |
| AYUM5141 | St. Marianna Univ. | Kyushu | 353,245,362 | 351,322,689 | 52,986,804,300 | 52,816,441,543 | 99.46 | 6.00 | 17.61 | 90.64 |
| AYUM5144 | St. Marianna Univ. | Kyushu | 354,394,126 | 352,542,468 | 53,159,118,900 | 52,964,669,184 | 99.48 | 6.35 | 17.65 | 90.15 |
| AYUM5145 | St. Marianna Univ. | Kyushu | 334,158,334 | 332,663,998 | 50,123,750,100 | 50,002,558,787 | 99.55 | 5.87 | 16.67 | 91.22 |
| AYUM5146 | St. Marianna Univ. | Kyushu | 366,709,895 | 364,839,333 | 55,006,484,250 | 54,837,801,882 | 99.49 | 6.00 | 18.28 | 91.97 |
| AYUM5149 | St. Marianna Univ. | Kyushu | 369,785,621 | 367,874,721 | 55,467,843,150 | 55,273,634,828 | 99.48 | 7.41 | 18.42 | 94.70 |
| AYUM5150 | St. Marianna Univ. | Kyushu | 368,217,920 | 367,142,661 | 55,232,688,000 | 55,198,017,403 | 99.71 | 6.56 | 18.40 | 95.54 |
| AYUM5151 | St. Marianna Univ. | Kyushu | 345,792,925 | 343,923,510 | 51,868,938,750 | 51,686,655,921 | 99.46 | 7.22 | 17.23 | 92.57 |
| AYUM5152 | St. Marianna Univ. | Kyushu | 367,666,492 | 365,675,511 | 55,149,973,800 | 54,955,922,469 | 99.46 | 6.47 | 18.32 | 91.91 |
| AYUM5154 | St. Marianna Univ. | Kyushu | 343,922,430 | 342,141,585 | 51,588,364,500 | 51,411,323,098 | 99.48 | 5.98 | 17.14 | 92.78 |
| AYUM5155 | St. Marianna Univ. | Kyushu | 356,204,886 | 353,727,794 | 53,430,732,900 | 53,158,175,770 | 99.30 | 8.77 | 17.72 | 87.45 |
| AYUM5156 | St. Marianna Univ. | Kyushu | 369,072,414 | 366,319,863 | 55,360,862,100 | 55,057,280,320 | 99.25 | 7.51 | 18.35 | 88.94 |
| AYUM5157 | St. Marianna Univ. | Kyushu | 348,070,007 | 345,506,536 | 52,210,501,050 | 51,960,560,233 | 99.26 | 8.77 | 17.32 | 91.93 |
| AYUM5158 | St. Marianna Univ. | Kyushu | 401,639,445 | 398,484,229 | 60,245,916,750 | 59,896,608,413 | 99.21 | 4.97 | 19.97 | 96.65 |
| AYUM5164 | St. Marianna Univ. | Kyushu | 385,240,626 | 382,036,377 | 57,786,093,900 | 57,407,867,015 | 99.17 | 5.12 | 19.14 | 92.32 |
| AYUM5165 | St. Marianna Univ. | Kyushu | 336,595,095 | 333,917,528 | 50,489,264,250 | 50,171,157,542 | 99.20 | 4.55 | 16.72 | 89.67 |
| HVUM0002 | St. Marianna Univ. | Kyushu | 359,378,995 | 357,229,221 | 53,906,849,250 | 53,635,800,718 | 99.40 | 7.30 | 17.88 | 93.60 |
| HVUM0003 | St. Marianna Univ. | Kyushu | 422,744,517 | 420,714,201 | 63,411,677,550 | 63,172,606,668 | 99.52 | 6.18 | 21.06 | 96.83 |
| HVUM0006 | St. Marianna Univ. | Kyushu | 393,534,519 | 390,668,437 | 59,030,177,850 | 58,676,518,901 | 99.27 | 7.30 | 19.56 | 96.36 |
| HVUM0009 | St. Marianna Univ. | Kyushu | 404,488,330 | 402,675,414 | 60,673,249,500 | 60,513,360,073 | 99.55 | 5.71 | 20.17 | 96.37 |
| HVUM0010 | St. Marianna Univ. | Kyushu | 431,042,945 | 429,267,325 | 64,656,441,750 | 64,536,034,330 | 99.59 | 4.94 | 21.51 | 98.51 |
| HVUM0011 | St. Marianna Univ. | Kyushu | 485,398,310 | 483,841,013 | 72,809,746,500 | 72,663,765,083 | 99.68 | 8.34 | 24.22 | 98.32 |
| HVUM0016 | St. Marianna Univ. | Kyushu | 388,049,231 | 385,774,950 | 58,207,384,650 | 57,926,032,002 | 99.41 | 8.82 | 19.31 | 92.35 |
| HVUM0017 | St. Marianna Univ. | Kyushu | 384,560,044 | 382,656,930 | 57,684,006,600 | 57,502,382,837 | 99.51 | 7.21 | 19.17 | 92.06 |
| HVUM0019 | St. Marianna Univ. | Kyushu | 367,933,720 | 365,920,826 | 55,190,058,000 | 55,011,725,256 | 99.45 | 7.93 | 18.34 | 94.74 |
| HVUM0020 | St. Marianna Univ. | Kyushu | 388,622,521 | 386,750,974 | 58,293,378,150 | 58,084,573,293 | 99.52 | 7.25 | 19.36 | 95.00 |
| HVUM0021 | St. Marianna Univ. | Kyushu | 350,748,902 | 349,156,240 | 52,612,335,300 | 52,479,106,458 | 99.55 | 6.73 | 17.49 | 89.15 |
| HVUM0022 | St. Marianna Univ. | Kyushu | 365,129,835 | 361,832,179 | 54,769,475,250 | 54,316,042,553 | 99.10 | 6.19 | 18.11 | 91.05 |
| HVUM0023 | St. Marianna Univ. | Kyushu | 402,022,789 | 400,611,094 | 60,303,418,350 | 60,130,182,641 | 99.65 | 5.78 | 20.04 | 96.25 |
| HVUM0027 | St. Marianna Univ. | Kyushu | 464,547,063 | 462,405,818 | 69,682,059,450 | 69,440,056,657 | 99.54 | 7.67 | 23.15 | 95.95 |
| HVUM0028 | St. Marianna Univ. | Kyushu | 426,726,075 | 424,039,367 | 64,008,911,250 | 63,670,827,970 | 99.37 | 7.57 | 21.22 | 97.68 |
| HVUM0032 | St. Marianna Univ. | Kyushu | 381,693,784 | 379,229,834 | 57,254,067,600 | 56,921,746,110 | 99.35 | 6.81 | 18.97 | 95.48 |
| HVUM0037 | St. Marianna Univ. | Kyushu | 388,599,664 | 385,208,431 | 58,289,949,600 | 57,998,054,577 | 99.13 | 3.27 | 19.33 | 95.83 |
| HVUM0039 | St. Marianna Univ. | Kyushu | 376,090,238 | 373,818,478 | 56,413,535,700 | 56,196,494,662 | 99.40 | 7.87 | 18.73 | 94.85 |
| HVUM0044 | St. Marianna Univ. | Kyushu | 410,608,016 | 408,452,701 | 61,591,202,400 | 61,377,398,373 | 99.48 | 8.12 | 20.46 | 91.70 |
| HVUM0046 | St. Marianna Univ. | Kyushu | 402,278,795 | 399,370,815 | 60,341,819,250 | 60,018,439,204 | 99.28 | 6.44 | 20.01 | 97.10 |

| | | | | | | | | | | |
|---|---|---|---|---|---|---|---|---|---|---|
| HVUM0048 | St. Marianna Univ. | Kyushu | 393,581,434 | 391,579,164 | 59,037,215,100 | 58,855,897,079 | 99.49 | 7.74 | 19.62 | 92.42 |
| HVUM0049 | St. Marianna Univ. | Kyushu | 363,597,064 | 360,464,984 | 54,539,559,600 | 54,078,629,854 | 99.14 | 7.51 | 18.03 | 92.90 |
| HVUM0050 | St. Marianna Univ. | Kyushu | 364,375,129 | 362,639,240 | 54,656,269,350 | 54,495,633,811 | 99.52 | 7.55 | 18.17 | 93.35 |
| HVUM0051 | St. Marianna Univ. | Kyushu | 361,071,135 | 358,053,704 | 54,160,670,250 | 53,790,900,207 | 99.16 | 6.46 | 17.93 | 90.72 |
| HVUM0056 | St. Marianna Univ. | Kyushu | 363,068,862 | 360,655,689 | 54,460,329,300 | 54,194,201,420 | 99.34 | 7.64 | 18.06 | 93.91 |
| HVUM0059 | St. Marianna Univ. | Kyushu | 339,182,266 | 336,556,677 | 50,877,339,900 | 50,555,963,884 | 99.23 | 7.12 | 16.85 | 90.43 |
| HVUM0060 | St. Marianna Univ. | Kyushu | 405,466,267 | 403,476,901 | 60,819,940,050 | 60,593,424,960 | 99.51 | 5.55 | 20.20 | 93.52 |
| HVUM0062 | St. Marianna Univ. | Kyushu | 380,358,388 | 378,742,834 | 57,053,758,200 | 56,850,366,631 | 99.58 | 7.06 | 18.95 | 94.54 |
| HVUM0064 | St. Marianna Univ. | Kyushu | 364,671,418 | 363,228,663 | 54,700,712,700 | 54,590,887,432 | 99.60 | 5.20 | 18.20 | 94.10 |
| HVUM0065 | St. Marianna Univ. | Kyushu | 361,065,845 | 357,966,633 | 54,159,876,750 | 53,794,831,321 | 99.14 | 7.45 | 17.93 | 93.46 |
| HVUM0077 | St. Marianna Univ. | Kyushu | 357,516,444 | 354,411,083 | 53,627,466,600 | 53,292,780,554 | 99.13 | 3.79 | 17.76 | 94.64 |
| HVUM0082 | St. Marianna Univ. | Kyushu | 404,434,730 | 401,155,475 | 60,665,209,500 | 60,234,831,501 | 99.19 | 8.88 | 20.08 | 96.27 |
| HVUM0083 | St. Marianna Univ. | Kyushu | 383,728,023 | 381,015,873 | 57,559,203,450 | 57,188,663,544 | 99.29 | 8.20 | 19.06 | 94.19 |
| HVUM0090 | St. Marianna Univ. | Kyushu | 366,462,461 | 361,170,417 | 54,969,369,150 | 54,202,465,107 | 98.56 | 7.17 | 18.07 | 92.91 |
| HVUM0091 | St. Marianna Univ. | Kyushu | 379,143,644 | 377,098,703 | 56,871,546,600 | 56,684,239,922 | 99.46 | 8.12 | 18.89 | 84.68 |
| HVUM0099 | St. Marianna Univ. | Kyushu | 408,473,857 | 406,600,871 | 61,271,078,550 | 61,086,929,291 | 99.54 | 6.96 | 20.36 | 96.55 |
| HVUM0102 | St. Marianna Univ. | Kyushu | 513,815,024 | 512,128,149 | 77,072,253,600 | 76,895,972,802 | 99.67 | 5.84 | 25.63 | 97.42 |
| HVUM0103 | St. Marianna Univ. | Kyushu | 415,874,079 | 413,507,560 | 62,381,111,850 | 62,151,538,988 | 99.43 | 8.66 | 20.72 | 89.94 |
| HVUM0105 | St. Marianna Univ. | Kyushu | 375,072,784 | 372,265,147 | 56,260,917,600 | 55,956,406,144 | 99.25 | 12.69 | 18.65 | 93.04 |
| HVUM0106 | St. Marianna Univ. | Kyushu | 368,033,516 | 365,051,613 | 55,205,027,400 | 54,861,203,468 | 99.19 | 8.33 | 18.29 | 93.39 |
| HVUM0108 | St. Marianna Univ. | Kyushu | 369,355,829 | 366,486,765 | 55,403,374,350 | 55,151,329,781 | 99.22 | 10.10 | 18.38 | 92.61 |
| HVUM0109 | St. Marianna Univ. | Kyushu | 382,294,194 | 380,655,363 | 57,344,129,100 | 57,205,403,757 | 99.57 | 8.91 | 19.07 | 92.00 |
| HVUM0112 | St. Marianna Univ. | Kyushu | 340,486,076 | 337,449,519 | 51,072,911,400 | 50,732,226,160 | 99.11 | 3.57 | 16.91 | 92.43 |
| HVUM0122 | St. Marianna Univ. | Kyushu | 364,450,611 | 361,866,273 | 54,667,591,650 | 54,339,370,308 | 99.29 | 8.09 | 18.11 | 92.73 |
| HVUM0142 | St. Marianna Univ. | Kyushu | 371,218,008 | 369,181,328 | 55,682,701,200 | 55,496,546,350 | 99.45 | 8.79 | 18.50 | 94.38 |
| HVUM0152 | St. Marianna Univ. | Kyushu | 407,257,279 | 405,559,429 | 61,088,591,850 | 60,942,314,021 | 99.58 | 6.64 | 20.31 | 96.91 |
| HVUM0154 | St. Marianna Univ. | Kyushu | 435,884,414 | 434,347,441 | 65,382,662,100 | 65,246,417,917 | 99.65 | 8.28 | 21.75 | 98.27 |
| HVUM0155 | St. Marianna Univ. | Kyushu | 369,477,592 | 367,624,866 | 55,421,638,800 | 55,236,683,766 | 99.50 | 5.40 | 18.41 | 94.11 |
| HVUM0157 | St. Marianna Univ. | Kyushu | 448,905,782 | 445,825,845 | 67,335,867,300 | 67,016,245,265 | 99.31 | 7.17 | 22.34 | 98.35 |
| HVUM0161 | St. Marianna Univ. | Kyushu | 441,259,247 | 438,988,181 | 66,188,887,050 | 65,979,653,467 | 99.49 | 5.54 | 21.99 | 97.39 |
| HVUM0162 | St. Marianna Univ. | Kyushu | 383,963,866 | 381,833,728 | 57,594,579,900 | 57,444,577,665 | 99.45 | 4.95 | 19.15 | 93.93 |
| HVUM0163 | St. Marianna Univ. | Kyushu | 477,409,642 | 474,571,573 | 71,611,446,300 | 71,249,270,308 | 99.41 | 5.30 | 23.75 | 98.80 |
| HVUM0169 | St. Marianna Univ. | Kyushu | 392,186,770 | 390,005,753 | 58,828,015,500 | 58,641,759,923 | 99.44 | 6.20 | 19.55 | 96.83 |
| HVUM0174 | St. Marianna Univ. | Kyushu | 409,765,571 | 406,165,802 | 61,464,835,650 | 61,034,357,558 | 99.12 | 8.28 | 20.34 | 97.09 |
| HVUM0196 | St. Marianna Univ. | Kyushu | 359,121,383 | 356,966,680 | 53,868,207,450 | 53,616,687,157 | 99.40 | 9.29 | 17.87 | 92.63 |
| HVUM0202 | St. Marianna Univ. | Kyushu | 384,166,435 | 381,646,870 | 57,624,965,250 | 57,332,739,139 | 99.34 | 6.66 | 19.11 | 94.02 |
| HVUM0262 | St. Marianna Univ. | Kyushu | 367,335,932 | 364,492,644 | 55,100,389,800 | 54,612,793,788 | 99.23 | 10.28 | 18.20 | 85.82 |
| HVUM0266 | St. Marianna Univ. | Kyushu | 402,519,905 | 400,329,399 | 60,377,985,750 | 60,122,370,823 | 99.46 | 9.85 | 20.04 | 96.08 |
| HVUM0271 | St. Marianna Univ. | Kyushu | 413,158,916 | 411,077,698 | 61,973,837,400 | 61,730,478,042 | 99.50 | 7.14 | 20.58 | 96.70 |
| HVUM0273 | St. Marianna Univ. | Kyushu | 371,595,142 | 369,217,662 | 55,739,271,300 | 55,477,503,618 | 99.36 | 8.54 | 18.49 | 91.10 |
| HVUM0274 | St. Marianna Univ. | Kyushu | 470,852,502 | 467,313,254 | 70,627,875,300 | 70,215,884,884 | 99.25 | 7.30 | 23.41 | 96.20 |
| HVUM0278 | St. Marianna Univ. | Kyushu | 402,289,320 | 399,511,083 | 60,343,398,000 | 60,037,883,865 | 99.31 | 6.85 | 20.01 | 96.69 |
| HVUM0283 | St. Marianna Univ. | Kyushu | 368,759,174 | 366,903,865 | 55,313,876,100 | 55,130,199,512 | 99.50 | 6.97 | 18.38 | 94.68 |
| HVUM0286 | St. Marianna Univ. | Kyushu | 366,395,219 | 364,496,408 | 54,959,282,850 | 54,857,685,236 | 99.48 | 8.27 | 18.29 | 94.29 |
| HVUM0290 | St. Marianna Univ. | Kyushu | 387,286,888 | 384,830,969 | 58,093,033,200 | 57,821,174,782 | 99.37 | 7.02 | 19.27 | 95.96 |
| HVUM0292 | St. Marianna Univ. | Kyushu | 372,441,813 | 370,234,604 | 55,866,271,950 | 55,661,576,842 | 99.41 | 6.95 | 18.55 | 92.42 |
| HVUM0295 | St. Marianna Univ. | Kyushu | 346,282,369 | 343,694,567 | 51,942,355,350 | 51,656,877,695 | 99.25 | 6.52 | 17.22 | 92.01 |
| HVUM0296 | St. Marianna Univ. | Kyushu | 373,776,754 | 371,419,642 | 56,066,513,100 | 55,765,813,821 | 99.37 | 6.00 | 18.59 | 94.94 |
| HVUM0303 | St. Marianna Univ. | Kyushu | 375,271,647 | 371,934,992 | 56,290,747,050 | 55,905,621,572 | 99.11 | 7.28 | 18.64 | 94.87 |
| HVUM0307 | St. Marianna Univ. | Kyushu | 380,502,987 | 377,097,075 | 57,075,448,050 | 56,674,784,320 | 99.10 | 7.44 | 18.89 | 95.14 |
| HVUM0314 | St. Marianna Univ. | Kyushu | 374,176,018 | 371,459,702 | 56,126,402,700 | 55,828,073,616 | 99.27 | 6.94 | 18.61 | 94.90 |
| HVUM0316 | St. Marianna Univ. | Kyushu | 357,050,586 | 353,381,791 | 53,557,587,900 | 53,080,211,106 | 98.97 | 6.49 | 17.69 | 92.94 |
| HVUM0317 | St. Marianna Univ. | Kyushu | 370,552,730 | 368,345,125 | 55,582,909,500 | 55,291,768,416 | 99.40 | 8.11 | 18.43 | 90.65 |
| HVUM0318 | St. Marianna Univ. | Kyushu | 361,857,396 | 356,022,745 | 54,278,609,400 | 53,502,772,840 | 98.39 | 5.68 | 17.83 | 92.43 |
| HVUM0322 | St. Marianna Univ. | Kyushu | 368,243,948 | 361,787,846 | 55,236,592,200 | 54,393,927,355 | 98.25 | 4.71 | 18.13 | 93.59 |
| HVUM0324 | St. Marianna Univ. | Kyushu | 415,497,466 | 413,081,537 | 62,324,619,900 | 62,029,230,069 | 99.42 | 10.46 | 20.68 | 96.59 |
| HVUM0332 | St. Marianna Univ. | Kyushu | 396,258,652 | 393,626,020 | 59,438,797,800 | 59,095,437,787 | 99.34 | 8.53 | 19.70 | 95.36 |
| HVUM0341 | St. Marianna Univ. | Kyushu | 440,251,899 | 437,216,219 | 66,037,784,850 | 65,721,473,149 | 99.31 | 8.37 | 21.91 | 94.60 |
| HVUM0346 | St. Marianna Univ. | Kyushu | 377,536,426 | 371,612,638 | 56,630,463,900 | 55,830,302,092 | 98.43 | 5.20 | 18.61 | 94.29 |
| HVUM0352 | St. Marianna Univ. | Kyushu | 396,718,619 | 393,829,465 | 59,507,792,850 | 59,181,500,880 | 99.27 | 13.55 | 19.73 | 93.35 |
| HVUM0359 | St. Marianna Univ. | Kyushu | 410,331,695 | 404,010,801 | 61,549,754,250 | 60,717,153,288 | 98.46 | 5.54 | 20.24 | 96.77 |
| HVUM0368 | St. Marianna Univ. | Kyushu | 349,448,381 | 346,836,651 | 52,417,257,150 | 52,036,875,626 | 99.25 | 6.56 | 17.35 | 91.96 |
| HVUM0375 | St. Marianna Univ. | Kyushu | 387,657,985 | 385,190,929 | 58,148,697,750 | 57,878,468,530 | 99.36 | 9.47 | 19.29 | 95.25 |
| HVUM0387 | St. Marianna Univ. | Kyushu | 394,284,466 | 392,766,335 | 59,142,669,900 | 58,970,468,447 | 99.61 | 5.67 | 19.66 | 95.96 |
| HVUM0388 | St. Marianna Univ. | Kyushu | 403,960,428 | 401,288,165 | 60,594,064,200 | 60,249,935,448 | 99.34 | 7.21 | 20.08 | 93.61 |
| HVUM0390 | St. Marianna Univ. | Kyushu | 393,527,369 | 387,996,280 | 59,029,105,350 | 58,303,113,104 | 98.59 | 5.59 | 19.43 | 95.76 |
| HVUM0392 | St. Marianna Univ. | Kyushu | 344,047,031 | 341,965,482 | 51,607,054,650 | 51,407,972,707 | 99.39 | 4.73 | 17.14 | 93.09 |
| HVUM0396 | St. Marianna Univ. | Kyushu | 359,577,467 | 357,449,956 | 53,936,620,050 | 53,698,729,458 | 99.41 | 9.22 | 17.90 | 91.63 |
| HVUM0402 | St. Marianna Univ. | Kyushu | 348,616,641 | 343,532,382 | 52,292,496,150 | 51,625,263,423 | 98.54 | 5.49 | 17.21 | 90.77 |
| HVUM0415 | St. Marianna Univ. | Kyushu | 385,230,547 | 382,003,287 | 57,784,582,050 | 57,342,435,586 | 99.16 | 7.05 | 19.11 | 92.33 |
| HVUM0417 | St. Marianna Univ. | Kyushu | 380,508,561 | 377,968,315 | 57,076,284,150 | 56,787,399,101 | 99.33 | 7.93 | 18.93 | 95.11 |
| HVUM0431 | St. Marianna Univ. | Kyushu | 374,227,093 | 371,549,468 | 56,134,063,950 | 55,767,854,852 | 99.28 | 6.86 | 18.59 | 94.34 |
| HVUM0432 | St. Marianna Univ. | Kyushu | 309,703,980 | 308,505,067 | 46,455,597,000 | 46,388,868,380 | 99.61 | 6.50 | 15.46 | 87.23 |
| HVUM0456 | St. Marianna Univ. | Kyushu | 401,857,524 | 400,430,730 | 60,278,628,600 | 60,139,078,293 | 99.64 | 7.39 | 20.05 | 96.29 |
| HVUM0457 | St. Marianna Univ. | Kyushu | 392,788,171 | 390,563,391 | 58,918,225,650 | 58,643,397,671 | 99.43 | 6.36 | 19.55 | 96.44 |
| HVUM0462 | St. Marianna Univ. | Kyushu | 345,931,398 | 340,801,066 | 51,889,709,700 | 51,210,495,896 | 98.52 | 4.83 | 17.07 | 90.96 |
| HVUM0468 | St. Marianna Univ. | Kyushu | 386,649,825 | 384,566,000 | 57,997,473,750 | 57,801,411,037 | 99.46 | 5.21 | 19.27 | 93.69 |
| HVUM0469 | St. Marianna Univ. | Kyushu | 386,237,085 | 380,207,872 | 57,935,562,750 | 57,139,765,776 | 98.44 | 5.32 | 19.05 | 95.17 |
| HVUM0475 | St. Marianna Univ. | Kyushu | 413,196,616 | 411,039,270 | 61,979,492,400 | 61,683,974,061 | 99.48 | 8.33 | 20.56 | 96.66 |
| HVUM0478 | St. Marianna Univ. | Kyushu | 388,104,144 | 386,051,384 | 58,215,621,600 | 57,996,512,813 | 99.47 | 6.59 | 19.33 | 93.47 |
| HVUM0480 | St. Marianna Univ. | Kyushu | 412,500,806 | 410,357,779 | 61,875,120,900 | 61,634,195,563 | 99.48 | 7.72 | 20.54 | 94.48 |
| HVUM0486 | St. Marianna Univ. | Kyushu | 397,712,836 | 396,211,029 | 59,656,925,400 | 59,546,155,055 | 99.62 | 6.20 | 19.85 | 95.89 |
| HVUM0490 | St. Marianna Univ. | Kyushu | 406,119,955 | 400,101,877 | 60,917,993,250 | 60,131,124,839 | 98.52 | 5.73 | 20.04 | 96.29 |
| HVUM0492 | St. Marianna Univ. | Kyushu | 368,324,622 | 362,844,359 | 55,248,693,300 | 54,511,027,225 | 98.51 | 5.08 | 18.17 | 93.33 |
| HVUM0497 | St. Marianna Univ. | Kyushu | 417,028,061 | 413,471,644 | 62,554,209,150 | 62,132,106,592 | 99.15 | 8.21 | 20.71 | 94.45 |
| HVUM0499 | St. Marianna Univ. | Kyushu | 508,020,930 | 504,513,288 | 76,203,139,500 | 75,720,943,056 | 99.31 | 18.03 | 25.24 | 97.96 |
| HVUM0502 | St. Marianna Univ. | Kyushu | 501,597,205 | 498,354,313 | 75,239,580,750 | 74,712,914,076 | 99.35 | 14.55 | 24.90 | 96.81 |
| HVUM0504 | St. Marianna Univ. | Kyushu | 431,438,784 | 429,011,270 | 64,715,817,600 | 64,507,266,806 | 99.44 | 5.47 | 21.50 | 98.38 |
| HVUM0508 | St. Marianna Univ. | Kyushu | 455,764,562 | 452,993,873 | 68,364,684,300 | 68,064,218,823 | 99.39 | 8.83 | 22.69 | 98.46 |
| HVUM0509 | St. Marianna Univ. | Kyushu | 337,037,671 | 334,493,395 | 50,555,650,650 | 50,263,984,993 | 99.25 | 7.80 | 16.75 | 90.12 |
| HVUM0510 | St. Marianna Univ. | Kyushu | 410,724,817 | 408,978,941 | 61,608,722,550 | 61,435,626,905 | 99.57 | 8.83 | 20.48 | 96.59 |
| HVUM0522 | St. Marianna Univ. | Kyushu | 424,719,806 | 421,662,886 | 63,707,970,900 | 62,306,100,326 | 99.28 | 6.46 | 20.77 | 73.13 |
| HVUM0523 | St. Marianna Univ. | Kyushu | 358,854,788 | 357,650,676 | 53,828,218,200 | 53,758,409,994 | 99.66 | 6.45 | 17.92 | 93.54 |
| HVUM0530 | St. Marianna Univ. | Kyushu | 379,929,024 | 377,624,449 | 56,989,353,600 | 56,760,690,087 | 99.39 | 8.24 | 18.92 | 95.35 |
| HVUM0538 | St. Marianna Univ. | Kyushu | 315,041,709 | 313,123,579 | 47,256,256,350 | 47,021,283,796 | 99.39 | 5.71 | 15.67 | 87.47 |
| HVUM0540 | St. Marianna Univ. | Kyushu | 419,322,277 | 416,396,714 | 62,898,341,550 | 62,586,599,848 | 99.30 | 8.52 | 20.86 | 97.40 |
| HVUM0541 | St. Marianna Univ. | Kyushu | 376,656,097 | 374,167,733 | 56,498,414,550 | 56,217,015,584 | 99.34 | 8.11 | 18.74 | 95.04 |
| HVUM0544 | St. Marianna Univ. | Kyushu | 402,356,855 | 399,818,105 | 60,353,528,250 | 60,076,806,347 | 99.37 | 8.24 | 20.03 | 93.57 |
| HVUM0545 | St. Marianna Univ. | Kyushu | 399,559,083 | 397,536,036 | 59,933,862,450 | 59,740,607,538 | 99.49 | 7.64 | 19.91 | 96.70 |
| HVUM0547 | St. Marianna Univ. | Kyushu | 353,271,006 | 351,287,153 | 52,990,650,900 | 52,766,288,472 | 99.44 | 6.17 | 17.59 | 93.41 |

| | | | | | | | | | | |
|---|---|---|---|---|---|---|---|---|---|---|
| HVUM0550 | St. Marianna Univ. | Kyushu | 411,955,973 | 409,616,347 | 61,793,395,950 | 61,515,031,797 | 99.43 | 8.18 | 20.51 | 96.76 |
| HVUM0551 | St. Marianna Univ. | Kyushu | 450,107,873 | 448,103,791 | 67,516,180,950 | 67,318,436,793 | 99.55 | 8.96 | 22.44 | 98.22 |
| HVUM0562 | St. Marianna Univ. | Kyushu | 361,769,920 | 360,180,040 | 54,265,488,000 | 54,097,505,664 | 99.56 | 6.63 | 18.03 | 93.39 |
| HVUM0564 | St. Marianna Univ. | Kyushu | 358,095,399 | 356,206,402 | 53,714,309,850 | 53,484,882,229 | 99.47 | 6.13 | 17.83 | 90.47 |
| HVUM0570 | St. Marianna Univ. | Kyushu | 334,325,743 | 332,650,749 | 50,148,861,450 | 49,984,795,193 | 99.50 | 6.24 | 16.66 | 90.99 |
| HVUM0572 | St. Marianna Univ. | Kyushu | 314,565,122 | 313,171,791 | 47,184,768,300 | 47,069,013,418 | 99.56 | 6.77 | 15.69 | 87.82 |
| HVUM0576 | St. Marianna Univ. | Kyushu | 332,881,111 | 331,110,241 | 49,932,166,650 | 49,753,223,097 | 99.47 | 5.34 | 16.58 | 90.93 |
| HVUM0585 | St. Marianna Univ. | Kyushu | 323,297,919 | 321,588,726 | 48,494,687,850 | 48,329,452,267 | 99.47 | 6.74 | 16.11 | 88.87 |
| HVUM0589 | St. Marianna Univ. | Kyushu | 390,430,139 | 388,431,188 | 58,564,520,850 | 58,395,809,484 | 99.49 | 5.54 | 19.47 | 96.69 |
| HVUM0590 | St. Marianna Univ. | Kyushu | 413,262,861 | 411,996,074 | 61,989,429,150 | 61,958,092,351 | 99.69 | 7.25 | 20.65 | 97.97 |
| HVUM0603 | St. Marianna Univ. | Kyushu | 389,557,600 | 387,221,260 | 58,433,640,000 | 58,211,223,403 | 99.40 | 4.94 | 19.40 | 94.06 |
| HVUM0604 | St. Marianna Univ. | Kyushu | 405,617,635 | 404,292,107 | 60,842,645,250 | 60,774,519,306 | 99.67 | 7.50 | 20.26 | 97.56 |
| HVUM0606 | St. Marianna Univ. | Kyushu | 470,792,082 | 469,170,340 | 70,618,812,300 | 70,528,503,788 | 99.66 | 7.78 | 23.51 | 97.01 |
| HVUM0607 | St. Marianna Univ. | Kyushu | 455,160,982 | 453,789,306 | 68,274,147,300 | 68,220,054,856 | 99.70 | 7.73 | 22.74 | 98.81 |
| HVUM0617 | St. Marianna Univ. | Kyushu | 416,531,756 | 414,968,430 | 62,479,763,400 | 62,385,370,661 | 99.62 | 7.21 | 20.80 | 95.10 |
| HVUM0619 | St. Marianna Univ. | Kyushu | 402,528,728 | 401,125,530 | 60,379,309,200 | 60,294,722,703 | 99.65 | 6.79 | 20.10 | 97.52 |
| HVUM0622 | St. Marianna Univ. | Kyushu | 441,820,161 | 440,161,271 | 66,273,024,150 | 66,169,370,649 | 99.62 | 7.36 | 22.06 | 96.20 |
| HVUM0625 | St. Marianna Univ. | Kyushu | 367,105,143 | 364,969,890 | 55,065,771,450 | 54,868,703,753 | 99.42 | 5.25 | 18.29 | 95.16 |
| HVUM0633 | St. Marianna Univ. | Kyushu | 339,481,971 | 337,742,872 | 50,922,295,650 | 50,761,466,281 | 99.49 | 4.74 | 16.92 | 89.54 |
| HVUM0634 | St. Marianna Univ. | Kyushu | 321,163,330 | 319,712,954 | 48,174,499,500 | 48,049,496,844 | 99.55 | 4.82 | 16.02 | 90.38 |
| HVUM0641 | St. Marianna Univ. | Kyushu | 306,489,305 | 304,833,548 | 45,973,395,750 | 45,819,965,769 | 99.46 | 4.19 | 15.27 | 87.55 |
| HVUM0642 | St. Marianna Univ. | Kyushu | 362,858,234 | 360,665,469 | 54,428,735,100 | 54,214,089,557 | 99.40 | 4.85 | 18.07 | 95.25 |
| HVUM0651 | St. Marianna Univ. | Kyushu | 412,872,781 | 411,012,557 | 61,930,917,150 | 61,762,428,953 | 99.55 | 4.49 | 20.59 | 95.28 |
| HVUM0657 | St. Marianna Univ. | Kyushu | 367,963,023 | 365,943,520 | 55,194,453,450 | 54,995,891,227 | 99.45 | 4.76 | 18.33 | 95.40 |
| HVUM0660 | St. Marianna Univ. | Kyushu | 362,328,909 | 360,203,189 | 54,349,336,350 | 54,121,216,025 | 99.41 | 4.72 | 18.04 | 95.03 |
| HVUM0669 | St. Marianna Univ. | Kyushu | 377,836,267 | 375,499,389 | 56,675,440,050 | 56,449,704,624 | 99.38 | 6.01 | 18.82 | 96.13 |
| HVUM0682 | St. Marianna Univ. | Kyushu | 360,133,612 | 357,936,307 | 54,020,041,800 | 53,798,289,293 | 99.39 | 5.21 | 17.93 | 91.31 |
| HVUM0684 | St. Marianna Univ. | Kyushu | 359,224,375 | 357,275,566 | 53,883,656,250 | 53,700,550,568 | 99.46 | 4.72 | 17.90 | 94.98 |
| HVUM0685 | St. Marianna Univ. | Kyushu | 332,118,648 | 330,246,258 | 49,817,797,200 | 49,640,894,466 | 99.44 | 4.87 | 16.55 | 91.58 |
| HVUM0687 | St. Marianna Univ. | Kyushu | 403,530,329 | 401,138,423 | 60,529,549,350 | 60,296,263,533 | 99.41 | 5.08 | 20.10 | 97.30 |
| HVUM0688 | St. Marianna Univ. | Kyushu | 366,029,196 | 364,142,952 | 54,904,379,400 | 54,727,456,273 | 99.48 | 5.01 | 18.24 | 95.43 |
| HVUM0695 | St. Marianna Univ. | Kyushu | 350,055,145 | 347,910,487 | 52,508,271,750 | 52,289,412,267 | 99.39 | 4.13 | 17.43 | 94.06 |
| HVUM0696 | St. Marianna Univ. | Kyushu | 322,397,976 | 320,735,938 | 48,359,696,400 | 48,187,649,256 | 99.48 | 3.43 | 16.06 | 86.89 |
| HVUM0698 | St. Marianna Univ. | Kyushu | 366,629,292 | 364,678,431 | 54,994,393,800 | 54,818,386,246 | 99.47 | 4.31 | 18.27 | 95.60 |
| HVUM0701 | St. Marianna Univ. | Kyushu | 328,222,866 | 326,365,563 | 49,233,429,900 | 49,058,930,025 | 99.43 | 5.80 | 16.35 | 90.51 |
| HVUM0707 | St. Marianna Univ. | Kyushu | 366,262,984 | 364,365,728 | 54,939,447,600 | 54,785,900,216 | 99.48 | 4.58 | 18.26 | 95.51 |
| HVUM0708 | St. Marianna Univ. | Kyushu | 404,132,583 | 402,160,166 | 60,619,887,450 | 60,456,809,228 | 99.51 | 4.82 | 20.15 | 97.57 |
| HVUM0710 | St. Marianna Univ. | Kyushu | 569,054,714 | 567,047,628 | 85,358,207,100 | 85,237,916,963 | 99.65 | 8.66 | 28.41 | 99.49 |
| HVUM0712 | St. Marianna Univ. | Kyushu | 386,322,519 | 384,207,529 | 57,948,377,850 | 57,762,608,311 | 99.45 | 5.03 | 19.25 | 96.54 |
| HVUM0713 | St. Marianna Univ. | Kyushu | 439,254,156 | 437,459,157 | 65,888,123,400 | 65,758,666,538 | 99.59 | 6.14 | 21.92 | 96.16 |
| HVUM0717 | St. Marianna Univ. | Kyushu | 391,974,244 | 389,629,235 | 58,796,136,600 | 58,595,808,264 | 99.40 | 5.11 | 19.53 | 97.15 |
| HVUM0718 | St. Marianna Univ. | Kyushu | 362,561,746 | 361,425,103 | 54,384,261,900 | 54,323,746,117 | 99.69 | 7.64 | 18.11 | 94.69 |
| HVUM0720 | St. Marianna Univ. | Kyushu | 384,854,569 | 383,274,194 | 57,728,185,350 | 57,626,244,480 | 99.59 | 6.01 | 19.21 | 96.72 |
| HVUM0721 | St. Marianna Univ. | Kyushu | 384,522,689 | 383,001,803 | 57,678,403,350 | 57,567,103,654 | 99.60 | 6.48 | 19.19 | 96.56 |
| HVUM0723 | St. Marianna Univ. | Kyushu | 345,052,377 | 343,858,990 | 51,757,856,550 | 51,686,270,778 | 99.65 | 5.73 | 17.23 | 93.54 |
| HVUM0724 | St. Marianna Univ. | Kyushu | 325,055,703 | 324,025,016 | 48,758,355,450 | 48,702,517,931 | 99.68 | 7.22 | 16.23 | 86.82 |
| HVUM0731 | St. Marianna Univ. | Kyushu | 353,845,106 | 352,717,938 | 53,076,765,900 | 53,033,348,486 | 99.68 | 7.54 | 17.68 | 93.60 |
| HVUM0733 | St. Marianna Univ. | Kyushu | 355,193,412 | 353,751,140 | 53,279,011,800 | 53,193,646,205 | 99.59 | 6.17 | 17.73 | 94.53 |
| HVUM0734 | St. Marianna Univ. | Kyushu | 391,033,129 | 389,762,673 | 58,654,969,350 | 58,609,324,908 | 99.68 | 7.75 | 19.54 | 93.22 |
| HVUM0735 | St. Marianna Univ. | Kyushu | 377,800,302 | 376,777,671 | 56,670,045,300 | 56,628,056,908 | 99.73 | 7.37 | 18.88 | 95.90 |
| HVUM0737 | St. Marianna Univ. | Kyushu | 412,405,063 | 411,305,466 | 61,860,759,450 | 61,844,673,614 | 99.73 | 6.82 | 20.61 | 97.95 |
| HVUM0746 | St. Marianna Univ. | Kyushu | 368,877,234 | 367,627,322 | 55,331,585,100 | 55,283,562,216 | 99.66 | 7.45 | 18.43 | 92.52 |
| HVUM0748 | St. Marianna Univ. | Kyushu | 367,051,624 | 365,952,183 | 55,057,743,600 | 55,043,439,166 | 99.70 | 7.50 | 18.35 | 95.05 |
| HVUM0750 | St. Marianna Univ. | Kyushu | 339,422,208 | 338,417,947 | 50,913,331,200 | 50,875,005,584 | 99.70 | 7.82 | 16.96 | 91.61 |
| HVUM0752 | St. Marianna Univ. | Kyushu | 349,598,290 | 348,471,906 | 52,439,743,500 | 52,388,027,263 | 99.68 | 6.98 | 17.46 | 90.56 |
| HVUM0754 | St. Marianna Univ. | Kyushu | 322,122,870 | 321,025,838 | 48,318,430,500 | 48,269,057,366 | 99.66 | 7.34 | 16.09 | 86.41 |
| HVUM0755 | St. Marianna Univ. | Kyushu | 365,505,647 | 364,245,229 | 54,825,847,050 | 54,754,721,283 | 99.66 | 8.23 | 18.25 | 94.31 |
| HVUM0761 | St. Marianna Univ. | Kyushu | 371,425,637 | 370,188,095 | 55,713,845,550 | 55,668,688,764 | 99.67 | 8.79 | 18.56 | 94.71 |
| HVUM0762 | St. Marianna Univ. | Kyushu | 364,905,635 | 363,702,876 | 54,735,845,250 | 54,681,974,507 | 99.67 | 8.04 | 18.23 | 94.54 |
| HVUM0765 | St. Marianna Univ. | Kyushu | 415,670,128 | 414,228,580 | 62,350,519,200 | 62,164,371,758 | 99.65 | 6.06 | 20.72 | 96.91 |
| HVUM0774 | St. Marianna Univ. | Kyushu | 390,015,785 | 388,595,352 | 58,502,367,750 | 58,331,212,949 | 99.64 | 5.56 | 19.44 | 95.70 |
| HVUM0776 | St. Marianna Univ. | Kyushu | 369,350,883 | 368,053,611 | 55,402,632,450 | 55,265,489,710 | 99.65 | 5.50 | 18.42 | 91.72 |
| HVUM0787 | St. Marianna Univ. | Kyushu | 404,979,637 | 403,575,263 | 60,746,945,550 | 60,539,778,810 | 99.65 | 5.85 | 20.18 | 93.66 |
| HVUM0788 | St. Marianna Univ. | Kyushu | 490,063,611 | 487,800,165 | 73,509,541,650 | 73,275,706,132 | 99.54 | 8.10 | 24.43 | 96.56 |
| HVUM0803 | St. Marianna Univ. | Kyushu | 340,864,408 | 338,293,102 | 51,129,661,200 | 50,188,149,283 | 99.25 | 5.13 | 16.73 | 74.47 |
| HVUM1007 | St. Marianna Univ. | Kyushu | 470,702,018 | 469,180,502 | 70,605,302,700 | 70,478,218,001 | 99.68 | 6.33 | 23.49 | 98.54 |
| HVUM1073 | St. Marianna Univ. | Kyushu | 409,671,310 | 407,869,393 | 61,450,696,500 | 61,298,505,905 | 99.56 | 8.12 | 20.43 | 93.67 |
| HVUM1078 | St. Marianna Univ. | Kyushu | 354,541,227 | 351,902,018 | 53,181,184,050 | 52,876,866,198 | 99.26 | 6.27 | 17.63 | 93.31 |
| HVUM1080 | St. Marianna Univ. | Kyushu | 347,030,831 | 345,097,168 | 52,054,624,650 | 51,871,513,318 | 99.44 | 6.33 | 17.29 | 89.60 |
| HVUM1081 | St. Marianna Univ. | Kyushu | 342,283,659 | 340,193,032 | 51,342,548,850 | 51,102,653,160 | 99.39 | 4.90 | 17.03 | 92.15 |
| HVUM1084 | St. Marianna Univ. | Kyushu | 311,070,051 | 308,690,446 | 46,660,507,650 | 46,479,956,807 | 99.24 | 4.45 | 15.49 | 84.58 |
| HVUM1085 | St. Marianna Univ. | Kyushu | 398,864,160 | 397,201,020 | 59,829,624,000 | 59,692,167,343 | 99.58 | 6.75 | 19.90 | 97.03 |
| C_AC0006 | Aichi Cancer Center | Kansai | 430,558,110 | 428,851,918 | 64,583,716,500 | 64,470,346,095 | 99.60 | 5.86 | 21.49 | 98.40 |
| C_AC0020 | Aichi Cancer Center | Kansai | 405,725,356 | 403,816,158 | 60,858,803,400 | 60,714,290,193 | 99.53 | 5.52 | 20.24 | 97.71 |
| C_AC0032 | Aichi Cancer Center | Kansai | 357,565,728 | 355,885,775 | 53,634,859,200 | 53,495,315,240 | 99.53 | 5.22 | 17.83 | 94.88 |
| C_AC0038 | Aichi Cancer Center | Kansai | 330,633,549 | 329,482,398 | 49,595,032,350 | 49,533,683,542 | 99.65 | 5.64 | 16.51 | 88.67 |
| C_AC0043 | Aichi Cancer Center | Kansai | 345,999,113 | 344,635,320 | 51,899,866,950 | 51,824,515,614 | 99.61 | 5.38 | 17.27 | 90.65 |
| C_AC0045 | Aichi Cancer Center | Kansai | 425,417,744 | 423,625,198 | 63,812,661,600 | 63,705,627,907 | 99.58 | 6.31 | 21.24 | 98.22 |
| C_AC0064 | Aichi Cancer Center | Kansai | 433,901,278 | 432,141,521 | 65,085,191,700 | 64,994,398,301 | 99.59 | 5.13 | 21.66 | 98.48 |
| C_AC0067 | Aichi Cancer Center | Kansai | 385,199,821 | 383,520,145 | 57,779,973,150 | 57,661,508,982 | 99.56 | 5.74 | 19.22 | 96.63 |
| C_AC0069 | Aichi Cancer Center | Kansai | 442,161,206 | 440,189,001 | 66,324,180,900 | 66,159,269,626 | 99.55 | 5.90 | 22.05 | 96.18 |
| C_AC0071 | Aichi Cancer Center | Kansai | 392,743,320 | 390,893,227 | 58,911,498,000 | 58,760,668,181 | 99.53 | 5.08 | 19.59 | 97.13 |
| C_AC0082 | Aichi Cancer Center | Kansai | 441,143,297 | 438,705,700 | 66,171,494,550 | 65,943,651,284 | 99.45 | 4.94 | 21.98 | 98.45 |
| C_AC0084 | Aichi Cancer Center | Kansai | 393,462,742 | 391,818,346 | 59,019,411,300 | 58,909,590,482 | 99.58 | 5.11 | 19.64 | 97.29 |
| C_AC0086 | Aichi Cancer Center | Kansai | 347,353,822 | 346,420,995 | 52,103,073,300 | 52,077,574,894 | 99.73 | 4.89 | 17.36 | 94.33 |
| C_AC0087 | Aichi Cancer Center | Kansai | 344,409,712 | 343,494,296 | 51,661,456,800 | 51,626,419,918 | 99.73 | 4.91 | 17.21 | 94.09 |
| C_AC0091 | Aichi Cancer Center | Kansai | 313,501,008 | 312,608,773 | 47,025,151,200 | 46,980,594,055 | 99.72 | 4.35 | 15.66 | 86.76 |
| C_AC0092 | Aichi Cancer Center | Kansai | 330,969,121 | 330,067,989 | 49,645,368,150 | 49,614,100,199 | 99.73 | 4.00 | 16.54 | 92.69 |
| C_AC0097 | Aichi Cancer Center | Kansai | 436,704,254 | 434,707,044 | 65,505,638,100 | 65,350,005,022 | 99.54 | 10.10 | 21.78 | 95.12 |
| C_AC0103 | Aichi Cancer Center | Kansai | 360,030,889 | 359,031,787 | 54,004,633,350 | 53,938,779,055 | 99.72 | 4.48 | 17.98 | 95.38 |
| C_AC0104 | Aichi Cancer Center | Kansai | 335,244,135 | 334,284,476 | 50,286,620,250 | 50,250,193,300 | 99.71 | 4.26 | 16.75 | 89.91 |
| C_AC0118 | Aichi Cancer Center | Kansai | 318,796,134 | 317,501,168 | 47,819,420,100 | 47,726,519,678 | 99.59 | 4.02 | 15.91 | 87.40 |
| C_AC0121 | Aichi Cancer Center | Kansai | 391,529,613 | 390,439,936 | 58,729,441,950 | 58,686,636,815 | 99.72 | 5.10 | 19.56 | 94.37 |
| C_AC0124 | Aichi Cancer Center | Kansai | 365,463,403 | 364,025,990 | 54,819,510,450 | 54,712,724,555 | 99.61 | 4.29 | 18.24 | 92.63 |
| C_AC0132 | Aichi Cancer Center | Kansai | 333,052,432 | 331,937,164 | 49,957,864,800 | 49,899,906,307 | 99.67 | 4.56 | 16.63 | 92.45 |
| C_AC0133 | Aichi Cancer Center | Kansai | 335,545,285 | 334,243,419 | 50,331,792,750 | 50,236,057,855 | 99.61 | 3.82 | 16.75 | 89.93 |
| C_AC0141 | Aichi Cancer Center | Kansai | 359,013,003 | 357,597,662 | 53,851,950,450 | 53,722,364,132 | 99.61 | 4.31 | 17.91 | 91.64 |
| C_AC0142 | Aichi Cancer Center | Kansai | 461,681,878 | 459,827,474 | 69,252,281,700 | 69,118,575,689 | 99.60 | 10.38 | 23.04 | 96.18 |
| C_AC0145 | Aichi Cancer Center | Kansai | 350,058,456 | 349,119,854 | 52,508,768,400 | 52,478,723,145 | 99.73 | 4.54 | 17.49 | 94.60 |
| C_AC0146 | Aichi Cancer Center | Kansai | 329,932,241 | 328,676,433 | 49,489,836,150 | 49,395,362,279 | 99.62 | 3.79 | 16.47 | 88.91 |

| | | | | | | | | | | |
|---|---|---|---|---|---|---|---|---|---|---|
| C_AC0147 | Aichi Cancer Center | Kansai | 344,507,962 | 342,887,242 | 51,676,194,300 | 51,557,755,006 | 99.53 | 3.43 | 17.19 | 94.42 |
| C_AC0149 | Aichi Cancer Center | Kansai | 368,751,284 | 367,735,045 | 55,312,692,600 | 55,285,174,600 | 99.72 | 4.88 | 18.43 | 93.13 |
| C_AC0152 | Aichi Cancer Center | Kansai | 307,307,515 | 306,220,211 | 46,096,127,250 | 46,036,662,118 | 99.65 | 4.57 | 15.35 | 85.39 |
| C_AC0166 | Aichi Cancer Center | Kansai | 354,520,609 | 353,110,359 | 53,178,091,350 | 53,066,556,696 | 99.60 | 3.79 | 17.69 | 94.95 |
| C_AC0176 | Aichi Cancer Center | Kansai | 394,468,186 | 393,250,629 | 59,170,227,900 | 59,135,640,727 | 99.69 | 4.10 | 19.71 | 94.74 |
| C_AC0179 | Aichi Cancer Center | Kansai | 332,688,592 | 331,281,123 | 49,903,288,800 | 49,789,080,522 | 99.58 | 4.12 | 16.60 | 89.41 |
| C_AC0180 | Aichi Cancer Center | Kansai | 363,419,228 | 362,161,426 | 54,512,884,200 | 54,425,575,765 | 99.65 | 4.96 | 18.14 | 95.59 |
| C_AC0181 | Aichi Cancer Center | Kansai | 355,659,598 | 354,375,626 | 53,348,939,700 | 53,267,236,912 | 99.64 | 4.90 | 17.76 | 94.99 |
| C_AC0182 | Aichi Cancer Center | Kansai | 409,631,194 | 408,058,683 | 61,444,679,100 | 61,343,066,805 | 99.62 | 4.93 | 20.45 | 97.88 |
| C_AC0185 | Aichi Cancer Center | Kansai | 369,029,629 | 368,074,981 | 55,354,444,350 | 55,320,419,742 | 99.74 | 6.09 | 18.44 | 95.82 |
| C_AC0188 | Aichi Cancer Center | Kansai | 362,093,996 | 360,832,664 | 54,314,099,400 | 54,215,724,350 | 99.65 | 5.74 | 18.07 | 91.78 |
| C_AC0190 | Aichi Cancer Center | Kansai | 333,091,693 | 331,939,476 | 49,963,753,950 | 49,885,677,826 | 99.65 | 4.54 | 16.63 | 89.32 |
| C_AC0207 | Aichi Cancer Center | Kansai | 368,612,184 | 367,153,208 | 55,291,827,600 | 55,182,082,763 | 99.60 | 5.03 | 18.39 | 95.61 |
| C_AC0214 | Aichi Cancer Center | Kansai | 325,259,955 | 323,937,933 | 48,788,993,250 | 48,698,674,592 | 99.59 | 4.30 | 16.23 | 88.59 |
| C_AC0216 | Aichi Cancer Center | Kansai | 346,955,505 | 345,666,752 | 52,043,325,750 | 51,970,204,094 | 99.63 | 4.94 | 17.32 | 94.08 |
| C_AC0218 | Aichi Cancer Center | Kansai | 417,353,922 | 415,644,296 | 62,603,088,300 | 62,485,157,388 | 99.59 | 5.27 | 20.83 | 95.20 |
| C_AC0219 | Aichi Cancer Center | Kansai | 353,928,170 | 352,434,393 | 53,089,225,500 | 52,979,954,468 | 99.58 | 4.88 | 17.66 | 94.30 |
| C_AC0222 | Aichi Cancer Center | Kansai | 373,493,054 | 372,440,130 | 56,023,958,100 | 55,982,003,883 | 99.72 | 5.63 | 18.66 | 93.23 |
| C_AC0223 | Aichi Cancer Center | Kansai | 336,646,279 | 335,291,508 | 50,496,941,850 | 50,397,406,543 | 99.60 | 4.88 | 16.80 | 89.58 |
| C_AC0224 | Aichi Cancer Center | Kansai | 360,547,137 | 358,947,822 | 54,082,070,550 | 53,956,937,935 | 99.56 | 5.23 | 17.99 | 95.01 |
| C_AC0229 | Aichi Cancer Center | Kansai | 353,108,876 | 351,695,449 | 52,966,331,400 | 52,860,048,413 | 99.60 | 5.15 | 17.62 | 91.41 |
| C_AC0234 | Aichi Cancer Center | Kansai | 380,582,068 | 379,145,733 | 57,087,310,200 | 56,952,535,758 | 99.62 | 4.61 | 18.98 | 93.39 |
| C_AC0248 | Aichi Cancer Center | Kansai | 347,699,277 | 346,207,767 | 52,154,891,550 | 52,044,121,604 | 99.57 | 4.82 | 17.35 | 90.95 |
| C_AC0255 | Aichi Cancer Center | Kansai | 370,221,899 | 368,618,031 | 55,533,284,850 | 55,398,805,725 | 99.57 | 5.04 | 18.47 | 92.70 |
| C_AC0256 | Aichi Cancer Center | Kansai | 383,213,279 | 381,822,741 | 57,481,991,850 | 57,404,669,621 | 99.64 | 4.71 | 19.13 | 96.85 |
| C_AC0261 | Aichi Cancer Center | Kansai | 368,082,283 | 366,738,772 | 55,212,342,450 | 55,139,297,966 | 99.63 | 5.08 | 18.38 | 92.87 |
| C_AC0265 | Aichi Cancer Center | Kansai | 389,190,923 | 387,907,207 | 58,378,638,450 | 58,314,790,133 | 99.67 | 6.14 | 19.44 | 96.71 |
| C_AC0271 | Aichi Cancer Center | Kansai | 347,002,497 | 345,677,687 | 52,050,374,550 | 51,959,376,580 | 99.62 | 5.35 | 17.32 | 90.68 |
| C_AC0276 | Aichi Cancer Center | Kansai | 325,073,295 | 323,740,134 | 48,760,994,250 | 48,670,745,477 | 99.59 | 5.16 | 16.22 | 91.02 |
| C_AC0282 | Aichi Cancer Center | Kansai | 331,764,281 | 330,156,167 | 49,764,642,150 | 49,633,871,474 | 99.52 | 4.98 | 16.54 | 88.92 |
| C_AC0285 | Aichi Cancer Center | Kansai | 399,639,876 | 397,906,483 | 59,945,981,400 | 59,779,364,577 | 99.57 | 5.74 | 19.93 | 94.47 |
| C_AC0286 | Aichi Cancer Center | Kansai | 360,415,140 | 358,592,662 | 54,062,271,000 | 53,912,255,044 | 99.49 | 5.13 | 17.97 | 91.75 |
| C_AC0292 | Aichi Cancer Center | Kansai | 358,709,128 | 357,507,293 | 53,806,369,200 | 53,750,143,134 | 99.66 | 5.64 | 17.92 | 94.95 |
| C_AC0299 | Aichi Cancer Center | Kansai | 359,942,627 | 358,940,234 | 53,991,394,050 | 53,948,620,458 | 99.72 | 6.16 | 17.98 | 91.93 |
| C_AC0311 | Aichi Cancer Center | Kansai | 364,565,080 | 363,516,749 | 54,684,762,000 | 54,654,454,367 | 99.71 | 6.18 | 18.22 | 92.42 |
| C_AC0320 | Aichi Cancer Center | Kansai | 326,722,629 | 325,368,838 | 49,008,394,350 | 48,898,383,772 | 99.59 | 5.06 | 16.30 | 88.21 |
| C_AC0321 | Aichi Cancer Center | Kansai | 336,138,705 | 334,589,450 | 50,420,805,750 | 50,265,689,467 | 99.54 | 5.24 | 16.76 | 89.12 |
| C_AC0322 | Aichi Cancer Center | Kansai | 396,688,949 | 394,263,882 | 59,503,342,350 | 59,257,343,402 | 99.39 | 3.79 | 19.75 | 97.69 |
| C_AC0332 | Aichi Cancer Center | Kansai | 378,617,366 | 377,249,800 | 56,792,604,900 | 56,699,683,949 | 99.64 | 5.35 | 18.90 | 93.39 |
| C_AC0334 | Aichi Cancer Center | Kansai | 327,294,944 | 325,935,534 | 49,094,241,600 | 48,998,172,945 | 99.58 | 5.29 | 16.33 | 88.01 |
| C_AC0340 | Aichi Cancer Center | Kansai | 348,589,250 | 347,386,975 | 52,288,387,500 | 52,231,789,705 | 99.66 | 5.04 | 17.41 | 93.19 |
| C_AC0343 | Aichi Cancer Center | Kansai | 393,900,817 | 392,634,773 | 59,085,122,550 | 59,034,922,918 | 99.68 | 6.16 | 19.68 | 94.30 |
| C_AC0346 | Aichi Cancer Center | Kansai | 382,426,726 | 381,096,076 | 57,364,008,900 | 57,293,704,925 | 99.65 | 5.86 | 19.10 | 93.37 |
| C_AC0349 | Aichi Cancer Center | Kansai | 357,037,068 | 355,895,810 | 53,555,560,200 | 53,467,775,949 | 99.68 | 5.23 | 17.82 | 91.59 |
| C_AC0351 | Aichi Cancer Center | Kansai | 333,577,699 | 332,137,022 | 50,036,654,850 | 49,887,054,649 | 99.57 | 4.77 | 16.63 | 92.07 |
| C_AC0359 | Aichi Cancer Center | Kansai | 359,658,809 | 358,145,895 | 53,948,821,350 | 53,840,205,928 | 99.58 | 5.80 | 17.95 | 91.65 |
| C_AC0361 | Aichi Cancer Center | Kansai | 340,917,796 | 339,241,008 | 51,137,669,400 | 51,012,084,071 | 99.51 | 4.10 | 17.00 | 90.36 |
| C_AC0365 | Aichi Cancer Center | Kansai | 364,292,602 | 362,581,996 | 54,643,890,300 | 54,501,997,285 | 99.53 | 4.38 | 18.17 | 95.17 |
| C_AC0376 | Aichi Cancer Center | Kansai | 325,102,149 | 323,580,329 | 48,765,322,350 | 48,646,994,030 | 99.53 | 4.20 | 16.22 | 90.69 |
| C_AC0379 | Aichi Cancer Center | Kansai | 392,764,466 | 391,689,811 | 58,914,669,900 | 58,874,849,139 | 99.73 | 5.04 | 19.62 | 97.27 |
| C_AC0384 | Aichi Cancer Center | Kansai | 325,367,796 | 324,094,581 | 48,805,169,400 | 48,700,180,286 | 99.61 | 5.09 | 16.23 | 87.89 |
| C_AC0386 | Aichi Cancer Center | Kansai | 315,049,547 | 313,745,725 | 47,257,432,050 | 47,170,021,791 | 99.59 | 4.21 | 15.72 | 89.98 |
| C_AC0387 | Aichi Cancer Center | Kansai | 403,251,972 | 401,138,929 | 60,487,795,800 | 60,289,745,740 | 99.48 | 4.41 | 20.10 | 94.55 |
| C_AC0389 | Aichi Cancer Center | Kansai | 356,790,617 | 354,671,052 | 53,518,592,550 | 53,322,518,354 | 99.41 | 4.47 | 17.77 | 91.59 |
| C_AC0401 | Aichi Cancer Center | Kansai | 355,456,112 | 354,208,788 | 53,318,416,800 | 53,267,126,907 | 99.65 | 4.71 | 17.76 | 95.01 |
| C_AC0409 | Aichi Cancer Center | Kansai | 341,960,623 | 340,198,818 | 51,294,093,450 | 51,123,765,060 | 99.48 | 4.68 | 17.04 | 92.45 |
| C_AC0412 | Aichi Cancer Center | Kansai | 331,421,549 | 330,151,792 | 49,713,232,350 | 49,632,924,403 | 99.62 | 4.63 | 16.54 | 91.70 |
| C_AC0414 | Aichi Cancer Center | Kansai | 320,758,675 | 319,459,552 | 48,113,801,250 | 48,026,003,540 | 99.59 | 4.64 | 16.01 | 87.49 |
| C_AC0421 | Aichi Cancer Center | Kansai | 351,772,203 | 350,233,681 | 52,765,830,450 | 52,646,675,170 | 99.56 | 4.56 | 17.55 | 91.14 |
| C_AC0424 | Aichi Cancer Center | Kansai | 373,268,903 | 371,770,606 | 55,990,335,450 | 55,874,142,643 | 99.60 | 4.66 | 18.62 | 93.07 |
| C_AC0429 | Aichi Cancer Center | Kansai | 373,598,901 | 372,228,923 | 56,039,835,150 | 55,933,115,901 | 99.63 | 4.31 | 18.64 | 93.27 |
| C_AC0433 | Aichi Cancer Center | Kansai | 331,857,000 | 330,208,814 | 49,778,550,000 | 49,659,676,560 | 99.50 | 3.99 | 16.55 | 92.10 |
| C_AC0434 | Aichi Cancer Center | Kansai | 369,540,693 | 368,147,338 | 55,431,103,950 | 55,357,384,088 | 99.62 | 4.39 | 18.45 | 96.11 |
| C_AC0437 | Aichi Cancer Center | Kansai | 376,144,180 | 374,415,185 | 56,421,627,000 | 56,281,756,185 | 99.54 | 4.57 | 18.76 | 95.95 |
| C_AC0439 | Aichi Cancer Center | Kansai | 382,303,917 | 380,546,693 | 57,345,587,550 | 57,220,837,222 | 99.54 | 5.16 | 19.07 | 93.57 |
| C_AC0442 | Aichi Cancer Center | Kansai | 353,380,943 | 352,411,955 | 53,007,141,450 | 52,967,626,036 | 99.73 | 5.38 | 17.66 | 94.60 |
| C_AC0443 | Aichi Cancer Center | Kansai | 402,725,245 | 400,453,376 | 60,408,786,750 | 60,185,484,879 | 99.44 | 4.20 | 20.06 | 94.80 |
| C_AC0450 | Aichi Cancer Center | Kansai | 438,362,156 | 437,102,754 | 65,754,323,400 | 65,694,134,971 | 99.71 | 4.99 | 21.90 | 98.59 |
| C_AC0452 | Aichi Cancer Center | Kansai | 437,505,349 | 436,054,246 | 65,625,802,350 | 65,523,092,518 | 99.67 | 4.23 | 21.84 | 98.49 |
| C_AC0456 | Aichi Cancer Center | Kansai | 398,370,814 | 396,477,759 | 59,755,622,100 | 59,610,139,786 | 99.52 | 4.33 | 19.87 | 97.49 |
| C_AC0462 | Aichi Cancer Center | Kansai | 420,380,972 | 418,445,435 | 63,057,145,800 | 62,888,973,821 | 99.54 | 4.35 | 20.96 | 98.13 |
| C_AC0468 | Aichi Cancer Center | Kansai | 436,549,300 | 434,601,024 | 65,482,395,000 | 65,313,007,982 | 99.55 | 4.19 | 21.77 | 95.72 |
| C_AC0469 | Aichi Cancer Center | Kansai | 412,116,203 | 410,512,104 | 61,817,430,450 | 61,713,108,904 | 99.61 | 4.41 | 20.57 | 98.01 |
| C_AC0470 | Aichi Cancer Center | Kansai | 411,556,806 | 410,273,574 | 61,733,520,900 | 61,683,322,466 | 99.69 | 4.62 | 20.56 | 95.39 |
| C_AC0472 | Aichi Cancer Center | Kansai | 484,694,469 | 482,693,178 | 72,704,170,350 | 72,551,382,611 | 99.59 | 5.12 | 24.18 | 97.33 |
| C_AC0473 | Aichi Cancer Center | Kansai | 461,364,231 | 459,384,432 | 69,204,634,650 | 69,036,305,089 | 99.57 | 5.26 | 23.01 | 96.81 |
| C_AC0479 | Aichi Cancer Center | Kansai | 447,169,001 | 445,379,169 | 67,075,350,150 | 66,958,042,491 | 99.60 | 4.28 | 22.32 | 98.58 |
| C_AC0483 | Aichi Cancer Center | Kansai | 433,634,811 | 431,045,206 | 65,045,221,650 | 64,778,851,009 | 99.40 | 4.33 | 21.59 | 96.00 |
| C_AC0484 | Aichi Cancer Center | Kansai | 443,898,952 | 442,306,390 | 66,584,842,800 | 66,492,033,471 | 99.64 | 4.98 | 22.16 | 98.63 |
| C_AC0490 | Aichi Cancer Center | Kansai | 386,207,648 | 384,266,060 | 57,931,147,200 | 57,789,048,289 | 99.50 | 5.24 | 19.26 | 96.57 |
| C_AC0492 | Aichi Cancer Center | Kansai | 401,763,929 | 399,407,886 | 60,264,589,350 | 60,044,327,059 | 99.41 | 5.44 | 20.01 | 96.88 |
| C_AC0495 | Aichi Cancer Center | Kansai | 393,767,471 | 392,389,655 | 59,065,120,650 | 58,992,634,275 | 99.65 | 6.30 | 19.66 | 94.04 |
| C_AC0497 | Aichi Cancer Center | Kansai | 501,710,855 | 499,851,085 | 75,256,628,250 | 75,139,557,869 | 99.63 | 6.56 | 25.05 | 99.13 |
| C_AC0498 | Aichi Cancer Center | Kansai | 461,594,166 | 459,890,940 | 69,239,124,900 | 69,139,611,041 | 99.63 | 6.45 | 23.05 | 98.69 |
| C_AC0499 | Aichi Cancer Center | Kansai | 448,451,833 | 446,769,203 | 67,267,774,950 | 67,140,676,223 | 99.62 | 5.64 | 22.38 | 96.47 |
| C_AC0504 | Aichi Cancer Center | Kansai | 347,397,704 | 345,430,852 | 52,109,655,600 | 51,943,413,199 | 99.43 | 5.04 | 17.31 | 93.31 |
| C_AC0507 | Aichi Cancer Center | Kansai | 470,082,145 | 468,405,742 | 70,512,321,750 | 70,411,851,870 | 99.64 | 6.03 | 23.47 | 98.98 |
| C_AC0511 | Aichi Cancer Center | Kansai | 443,301,687 | 441,586,920 | 66,495,253,050 | 66,380,370,272 | 99.61 | 5.82 | 22.13 | 98.56 |
| C_AC0513 | Aichi Cancer Center | Kansai | 413,366,110 | 411,482,988 | 62,004,916,500 | 61,850,300,164 | 99.54 | 6.30 | 20.62 | 97.58 |
| C_AC0517 | Aichi Cancer Center | Kansai | 483,257,389 | 481,323,813 | 72,488,608,350 | 72,343,571,975 | 99.60 | 9.69 | 24.11 | 96.90 |
| C_AC0519 | Aichi Cancer Center | Kansai | 464,619,011 | 462,857,892 | 69,692,851,650 | 69,583,654,633 | 99.62 | 9.16 | 23.19 | 96.55 |
| C_AC0528 | Aichi Cancer Center | Kansai | 347,945,259 | 346,673,091 | 52,191,788,850 | 52,116,581,723 | 99.63 | 7.80 | 17.37 | 92.68 |
| C_AC0531 | Aichi Cancer Center | Kansai | 426,493,018 | 423,965,417 | 63,973,952,700 | 63,721,099,232 | 99.41 | 8.38 | 21.24 | 94.65 |
| C_AC0538 | Aichi Cancer Center | Kansai | 376,963,766 | 374,896,548 | 56,544,564,900 | 56,344,092,865 | 99.45 | 9.26 | 18.78 | 91.64 |
| C_AC0542 | Aichi Cancer Center | Kansai | 388,970,184 | 387,567,659 | 58,345,527,600 | 58,249,095,595 | 99.64 | 7.69 | 19.42 | 93.24 |
| C_AC0544 | Aichi Cancer Center | Kansai | 392,512,261 | 390,501,087 | 58,876,839,150 | 58,674,993,816 | 99.49 | 8.15 | 19.56 | 96.32 |
| C_AC0547 | Aichi Cancer Center | Kansai | 406,206,183 | 404,325,833 | 60,930,927,450 | 60,764,936,419 | 99.54 | 8.29 | 20.25 | 97.01 |
| C_AC0550 | Aichi Cancer Center | Kansai | 368,102,046 | 366,629,136 | 55,215,306,900 | 55,113,406,018 | 99.60 | 7.52 | 18.37 | 91.60 |
| C_AC0551 | Aichi Cancer Center | Kansai | 422,880,436 | 421,385,494 | 63,432,065,400 | 63,325,637,176 | 99.65 | 8.95 | 21.11 | 97.78 |
| C_AC0560 | Aichi Cancer Center | Kansai | 404,700,096 | 402,931,400 | 60,705,014,400 | 60,571,659,317 | 99.56 | 9.93 | 20.19 | 93.67 |

| | | | | | | | | | | |
|---|---|---|---|---|---|---|---|---|---|---|
| C_AC0562 | Aichi Cancer Center | Kansai | 389,715,763 | 387,805,218 | 58,457,364,450 | 58,299,624,577 | 99.51 | 8.34 | 19.43 | 93.16 |
| C_AC0567 | Aichi Cancer Center | Kansai | 379,926,666 | 378,010,660 | 56,988,999,900 | 56,817,325,314 | 99.50 | 8.38 | 18.94 | 95.41 |
| C_AC0568 | Aichi Cancer Center | Kansai | 371,061,606 | 369,759,117 | 55,659,240,900 | 55,580,756,934 | 99.65 | 8.30 | 18.53 | 94.95 |
| C_AC0572 | Aichi Cancer Center | Kansai | 405,380,885 | 403,493,476 | 60,807,132,750 | 60,634,146,113 | 99.53 | 8.59 | 20.21 | 93.93 |
| C_AC0573 | Aichi Cancer Center | Kansai | 393,673,242 | 392,075,988 | 59,050,986,300 | 58,938,271,890 | 99.59 | 9.47 | 19.65 | 93.03 |
| C_AC0587 | Aichi Cancer Center | Kansai | 419,986,138 | 418,589,755 | 62,997,920,700 | 62,891,752,772 | 99.67 | 8.21 | 20.96 | 97.60 |
| C_AC0596 | Aichi Cancer Center | Kansai | 380,724,296 | 379,447,970 | 57,108,644,400 | 57,032,008,070 | 99.66 | 9.06 | 19.01 | 92.18 |
| C_AC0605 | Aichi Cancer Center | Kansai | 377,506,800 | 376,215,124 | 56,626,020,000 | 56,536,987,736 | 99.66 | 9.81 | 18.85 | 94.73 |
| C_AC0608 | Aichi Cancer Center | Kansai | 373,559,860 | 372,243,877 | 56,033,979,000 | 55,963,020,254 | 99.65 | 7.97 | 18.65 | 95.11 |
| C_AC0611 | Aichi Cancer Center | Kansai | 394,360,961 | 389,037,034 | 59,154,144,150 | 58,476,800,401 | 98.65 | 7.68 | 19.49 | 96.17 |
| C_AC0613 | Aichi Cancer Center | Kansai | 419,774,870 | 418,399,915 | 62,966,230,500 | 62,886,958,223 | 99.67 | 8.58 | 20.96 | 97.61 |
| C_AC0617 | Aichi Cancer Center | Kansai | 438,469,006 | 436,866,241 | 65,770,350,900 | 65,675,416,409 | 99.63 | 9.52 | 21.89 | 98.10 |
| C_AC0620 | Aichi Cancer Center | Kansai | 394,790,295 | 393,079,606 | 59,218,544,250 | 59,067,587,622 | 99.57 | 10.37 | 19.69 | 92.95 |
| C_AC0625 | Aichi Cancer Center | Kansai | 401,725,575 | 400,216,424 | 60,258,836,250 | 60,158,056,308 | 99.62 | 9.67 | 20.05 | 93.71 |
| C_AC0632 | Aichi Cancer Center | Kansai | 410,000,349 | 408,501,902 | 61,500,052,350 | 61,403,552,461 | 99.63 | 10.83 | 20.47 | 93.87 |
| C_AC0640 | Aichi Cancer Center | Kansai | 442,515,678 | 440,896,885 | 66,377,351,700 | 66,236,378,617 | 99.63 | 13.08 | 22.08 | 94.34 |
| C_AC0644 | Aichi Cancer Center | Kansai | 405,592,770 | 404,163,521 | 60,838,915,500 | 60,758,686,535 | 99.65 | 10.26 | 20.25 | 93.80 |
| C_AC0645 | Aichi Cancer Center | Kansai | 403,330,394 | 402,065,076 | 60,499,559,100 | 60,415,832,491 | 99.69 | 10.98 | 20.14 | 93.27 |
| C_AC0648 | Aichi Cancer Center | Kansai | 411,012,007 | 409,575,254 | 61,651,801,050 | 61,553,261,384 | 99.65 | 11.48 | 20.52 | 96.55 |
| C_AC0649 | Aichi Cancer Center | Kansai | 411,667,693 | 410,165,792 | 61,750,153,950 | 61,645,197,935 | 99.64 | 9.34 | 20.55 | 94.29 |
| C_AC0651 | Aichi Cancer Center | Kansai | 384,053,056 | 382,421,310 | 57,607,958,400 | 57,487,985,539 | 99.58 | 10.81 | 19.16 | 95.01 |
| C_AC0655 | Aichi Cancer Center | Kansai | 470,288,672 | 468,647,880 | 70,543,300,800 | 70,446,502,384 | 99.65 | 10.03 | 23.48 | 98.70 |
| C_AC0664 | Aichi Cancer Center | Kansai | 439,447,245 | 438,060,207 | 65,917,086,750 | 65,837,434,011 | 99.68 | 9.39 | 21.95 | 98.06 |
| C_AC0665 | Aichi Cancer Center | Kansai | 374,082,977 | 372,697,493 | 56,112,446,550 | 56,024,507,253 | 99.63 | 9.62 | 18.67 | 94.66 |
| C_AC0666 | Aichi Cancer Center | Kansai | 374,582,344 | 373,182,755 | 56,187,351,600 | 56,071,729,460 | 99.63 | 9.24 | 18.69 | 91.64 |
| C_AC0667 | Aichi Cancer Center | Kansai | 404,980,792 | 403,561,660 | 60,747,118,800 | 60,662,425,584 | 99.65 | 10.30 | 20.22 | 93.75 |
| C_AC0669 | Aichi Cancer Center | Kansai | 424,969,641 | 423,536,351 | 63,745,446,150 | 63,654,971,365 | 99.66 | 10.52 | 21.22 | 97.45 |
| C_AC0685 | Aichi Cancer Center | Kansai | 395,069,868 | 393,681,592 | 59,260,480,200 | 59,178,621,822 | 99.65 | 9.54 | 19.73 | 93.20 |
| C_AC0686 | Aichi Cancer Center | Kansai | 395,048,705 | 393,833,508 | 59,257,305,750 | 59,213,389,886 | 99.69 | 9.97 | 19.74 | 93.28 |
| C_AC0696 | Aichi Cancer Center | Kansai | 415,593,727 | 414,184,748 | 62,339,059,050 | 62,255,805,532 | 99.66 | 10.39 | 20.75 | 94.28 |
| C_AC0701 | Aichi Cancer Center | Kansai | 393,918,293 | 392,631,466 | 59,087,743,950 | 59,000,063,812 | 99.67 | 8.70 | 19.67 | 93.28 |
| C_AC0702 | Aichi Cancer Center | Kansai | 368,526,588 | 367,192,999 | 55,278,988,200 | 55,197,298,580 | 99.64 | 8.83 | 18.40 | 91.61 |
| C_AC0709 | Aichi Cancer Center | Kansai | 419,848,143 | 418,530,304 | 62,977,221,450 | 62,913,679,519 | 99.69 | 8.82 | 20.97 | 95.13 |
| C_AC0713 | Aichi Cancer Center | Kansai | 440,976,952 | 439,726,827 | 66,146,542,800 | 66,081,423,220 | 99.72 | 7.89 | 22.03 | 98.45 |
| C_AC0714 | Aichi Cancer Center | Kansai | 384,990,540 | 383,692,510 | 57,748,581,000 | 57,658,338,241 | 99.66 | 8.94 | 19.22 | 92.86 |
| C_AC0719 | Aichi Cancer Center | Kansai | 396,512,855 | 394,604,954 | 59,476,928,250 | 59,294,818,747 | 99.52 | 7.72 | 19.76 | 96.91 |
| C_AC0722 | Aichi Cancer Center | Kansai | 448,252,628 | 444,521,960 | 67,237,894,200 | 66,803,640,007 | 99.17 | 7.92 | 22.27 | 98.47 |
| C_AC0723 | Aichi Cancer Center | Kansai | 450,123,422 | 448,170,387 | 67,518,513,300 | 67,378,951,602 | 99.57 | 9.13 | 22.46 | 98.49 |
| C_AC0733 | Aichi Cancer Center | Kansai | 427,775,408 | 426,437,844 | 64,166,311,200 | 64,097,018,038 | 99.69 | 8.20 | 21.37 | 98.10 |
| C_AC0736 | Aichi Cancer Center | Kansai | 372,272,935 | 370,839,970 | 55,840,940,250 | 55,770,538,309 | 99.62 | 8.29 | 18.59 | 95.38 |
| C_AC0744 | Aichi Cancer Center | Kansai | 403,335,597 | 401,961,342 | 60,500,339,550 | 60,427,674,751 | 99.66 | 8.78 | 20.14 | 94.20 |
| C_AC0747 | Aichi Cancer Center | Kansai | 382,412,443 | 381,370,111 | 57,361,866,450 | 57,318,770,604 | 99.73 | 7.81 | 19.11 | 93.25 |
| C_AC0753 | Aichi Cancer Center | Kansai | 374,109,820 | 372,975,239 | 56,116,473,000 | 56,074,972,063 | 99.70 | 8.91 | 18.69 | 92.03 |
| C_AC0754 | Aichi Cancer Center | Kansai | 418,029,619 | 416,867,625 | 62,704,442,850 | 62,664,848,667 | 99.72 | 8.84 | 20.89 | 97.72 |
| C_AC0761 | Aichi Cancer Center | Kansai | 396,697,534 | 395,582,805 | 59,504,630,100 | 59,461,756,942 | 99.72 | 8.21 | 19.82 | 93.99 |
| C_AC0766 | Aichi Cancer Center | Kansai | 361,928,057 | 360,760,923 | 54,289,208,550 | 54,236,440,071 | 99.68 | 7.33 | 18.08 | 94.58 |
| C_AC0787 | Aichi Cancer Center | Kansai | 412,148,398 | 410,874,616 | 61,822,259,700 | 61,760,929,325 | 99.69 | 8.41 | 20.59 | 94.57 |
| C_AC0790 | Aichi Cancer Center | Kansai | 388,361,240 | 387,093,029 | 58,254,186,000 | 58,194,004,545 | 99.67 | 9.17 | 19.40 | 96.13 |
| C_AC0795 | Aichi Cancer Center | Kansai | 415,478,351 | 414,233,895 | 62,321,752,650 | 62,267,933,051 | 99.70 | 8.99 | 20.76 | 97.62 |
| C_AC0800 | Aichi Cancer Center | Kansai | 418,534,943 | 417,463,733 | 62,780,241,450 | 62,764,027,177 | 99.74 | 8.35 | 20.92 | 97.83 |
| C_AC0803 | Aichi Cancer Center | Kansai | 394,135,935 | 392,901,275 | 59,120,390,250 | 59,065,028,722 | 99.69 | 8.28 | 19.69 | 93.89 |
| C_AC0807 | Aichi Cancer Center | Kansai | 463,514,840 | 462,225,606 | 69,527,226,000 | 69,445,610,977 | 99.72 | 8.81 | 23.15 | 96.45 |
| C_AC0809 | Aichi Cancer Center | Kansai | 380,355,798 | 379,474,478 | 57,053,369,700 | 57,038,153,091 | 99.77 | 7.18 | 19.01 | 95.92 |
| C_AC0810 | Aichi Cancer Center | Kansai | 386,627,745 | 385,453,805 | 57,994,161,750 | 57,958,444,674 | 99.70 | 8.24 | 19.32 | 93.03 |
| C_AC0814 | Aichi Cancer Center | Kansai | 377,255,571 | 375,315,526 | 56,588,335,650 | 56,425,622,798 | 99.49 | 9.02 | 18.81 | 94.71 |
| C_AC0815 | Aichi Cancer Center | Kansai | 344,632,925 | 343,778,944 | 51,694,938,750 | 51,678,076,102 | 99.75 | 7.15 | 17.23 | 93.14 |
| C_AC0817 | Aichi Cancer Center | Kansai | 358,098,677 | 357,094,481 | 53,714,801,550 | 53,640,622,015 | 99.72 | 7.36 | 17.88 | 94.12 |
| C_AC0839 | Aichi Cancer Center | Kansai | 359,449,288 | 358,464,796 | 53,917,393,200 | 53,873,847,237 | 99.73 | 7.53 | 17.96 | 91.29 |
| C_AC0840 | Aichi Cancer Center | Kansai | 367,518,495 | 366,525,128 | 55,127,774,250 | 55,087,466,999 | 99.73 | 8.18 | 18.36 | 94.72 |
| C_AC0841 | Aichi Cancer Center | Kansai | 380,514,025 | 379,510,873 | 57,077,103,750 | 57,067,516,168 | 99.74 | 8.38 | 19.02 | 95.86 |
| C_AC0847 | Aichi Cancer Center | Kansai | 387,739,724 | 386,880,210 | 58,160,958,600 | 58,140,258,793 | 99.78 | 7.66 | 19.38 | 96.41 |
| C_AC0849 | Aichi Cancer Center | Kansai | 494,544,345 | 492,515,554 | 74,181,651,750 | 74,013,839,686 | 99.59 | 9.10 | 24.67 | 99.03 |
| C_AC0853 | Aichi Cancer Center | Kansai | 406,312,959 | 405,358,324 | 60,946,943,850 | 60,920,851,684 | 99.77 | 8.20 | 20.31 | 94.54 |
| C_AC0861 | Aichi Cancer Center | Kansai | 386,331,383 | 385,317,514 | 57,949,707,450 | 57,909,686,899 | 99.74 | 7.45 | 19.30 | 93.52 |
| C_AC0863 | Aichi Cancer Center | Kansai | 378,209,051 | 377,236,842 | 56,731,357,650 | 56,694,042,832 | 99.74 | 8.33 | 18.90 | 95.71 |
| C_AC0871 | Aichi Cancer Center | Kansai | 423,290,970 | 422,213,312 | 63,493,645,500 | 63,444,704,801 | 99.75 | 8.64 | 21.15 | 95.18 |
| C_AC0875 | Aichi Cancer Center | Kansai | 379,868,032 | 378,953,012 | 56,980,204,800 | 56,975,328,778 | 99.76 | 8.33 | 18.99 | 95.88 |
| C_AC0876 | Aichi Cancer Center | Kansai | 380,073,528 | 379,130,474 | 57,011,029,200 | 56,994,132,656 | 99.75 | 8.23 | 19.00 | 95.69 |
| C_AC0878 | Aichi Cancer Center | Kansai | 368,078,989 | 367,178,178 | 55,211,848,350 | 55,194,451,645 | 99.76 | 8.67 | 18.40 | 91.59 |
| C_AC0879 | Aichi Cancer Center | Kansai | 401,144,651 | 400,148,081 | 60,171,697,650 | 60,156,270,486 | 99.75 | 8.69 | 20.05 | 94.09 |
| C_AC0882 | Aichi Cancer Center | Kansai | 408,376,165 | 407,442,241 | 61,256,424,750 | 61,251,504,253 | 99.77 | 9.86 | 20.42 | 97.14 |
| C_AC0886 | Aichi Cancer Center | Kansai | 351,961,843 | 351,110,748 | 52,794,276,450 | 52,783,017,123 | 99.76 | 7.98 | 17.59 | 90.25 |
| C_AC0908 | Aichi Cancer Center | Kansai | 578,960,043 | 575,973,402 | 86,844,006,450 | 86,573,257,476 | 99.48 | 8.70 | 28.86 | 98.48 |
| C_AC0909 | Aichi Cancer Center | Kansai | 382,341,768 | 381,308,187 | 57,351,265,200 | 57,291,397,028 | 99.73 | 8.29 | 19.10 | 92.75 |
| C_AC0912 | Aichi Cancer Center | Kansai | 330,693,559 | 329,702,620 | 49,604,033,850 | 49,570,409,121 | 99.70 | 5.64 | 16.52 | 91.43 |
| C_AC0915 | Aichi Cancer Center | Kansai | 345,109,921 | 343,993,759 | 51,766,488,150 | 51,704,950,355 | 99.68 | 7.76 | 17.23 | 88.80 |
| C_AC0916 | Aichi Cancer Center | Kansai | 368,813,336 | 367,684,913 | 55,322,000,400 | 55,256,163,119 | 99.69 | 6.67 | 18.42 | 91.96 |
| C_AC0918 | Aichi Cancer Center | Kansai | 483,222,953 | 480,590,594 | 72,483,442,950 | 72,231,467,631 | 99.46 | 7.58 | 24.08 | 96.88 |
| C_AC0921 | Aichi Cancer Center | Kansai | 341,319,318 | 340,348,000 | 51,197,897,700 | 51,172,696,498 | 99.72 | 6.72 | 17.06 | 89.18 |
| C_AC0922 | Aichi Cancer Center | Kansai | 323,789,771 | 322,825,378 | 48,568,465,650 | 48,538,447,657 | 99.70 | 6.90 | 16.18 | 89.65 |
| C_AC0924 | Aichi Cancer Center | Kansai | 369,862,630 | 369,036,606 | 55,479,394,500 | 55,468,262,793 | 99.78 | 7.35 | 18.49 | 91.94 |
| C_AC0926 | Aichi Cancer Center | Kansai | 366,336,638 | 365,263,242 | 54,950,495,700 | 54,912,668,952 | 99.71 | 7.12 | 18.30 | 91.73 |
| C_AC0929 | Aichi Cancer Center | Kansai | 357,401,790 | 356,444,854 | 53,610,268,500 | 53,576,278,450 | 99.73 | 7.30 | 17.86 | 94.02 |
| C_AC0931 | Aichi Cancer Center | Kansai | 354,807,752 | 353,939,705 | 53,221,162,800 | 53,212,858,412 | 99.76 | 6.71 | 17.74 | 90.85 |
| C_AC0932 | Aichi Cancer Center | Kansai | 347,483,652 | 346,420,475 | 52,122,547,800 | 52,040,610,179 | 99.69 | 7.13 | 17.35 | 89.36 |
| C_AC0943 | Aichi Cancer Center | Kansai | 426,482,066 | 424,851,707 | 63,972,309,900 | 63,859,418,679 | 99.62 | 10.26 | 21.29 | 94.65 |
| C_AC0946 | Aichi Cancer Center | Kansai | 422,775,815 | 421,252,268 | 63,416,372,250 | 63,294,945,945 | 99.64 | 8.73 | 21.10 | 94.88 |
| C_AC0953 | Aichi Cancer Center | Kansai | 392,506,458 | 391,192,039 | 58,875,968,700 | 58,806,287,407 | 99.67 | 12.12 | 19.60 | 95.24 |
| C_AC0954 | Aichi Cancer Center | Kansai | 353,095,567 | 351,321,978 | 52,964,335,050 | 52,800,980,702 | 99.50 | 7.74 | 17.60 | 89.99 |
| C_AC0956 | Aichi Cancer Center | Kansai | 375,992,314 | 373,916,920 | 56,398,847,100 | 56,209,033,782 | 99.45 | 7.86 | 18.74 | 92.29 |
| C_AC0959 | Aichi Cancer Center | Kansai | 355,990,141 | 354,186,500 | 53,398,521,150 | 53,233,476,515 | 99.49 | 8.47 | 17.74 | 89.97 |
| C_AC0964 | Aichi Cancer Center | Kansai | 359,234,874 | 357,962,675 | 53,885,231,100 | 53,809,479,587 | 99.65 | 7.70 | 17.94 | 94.28 |
| C_AC0967 | Aichi Cancer Center | Kansai | 371,740,950 | 369,101,712 | 55,761,142,500 | 55,493,833,630 | 99.29 | 7.45 | 18.50 | 95.21 |
| C_AC0968 | Aichi Cancer Center | Kansai | 376,778,592 | 374,984,964 | 56,516,788,800 | 56,357,783,524 | 99.52 | 8.34 | 18.79 | 92.21 |
| C_AC0971 | Aichi Cancer Center | Kansai | 366,651,246 | 365,186,291 | 54,997,686,900 | 54,897,193,252 | 99.60 | 7.13 | 18.30 | 95.08 |
| C_AC0994 | Aichi Cancer Center | Kansai | 372,013,520 | 369,756,037 | 55,802,028,000 | 55,579,660,265 | 99.39 | 7.49 | 18.53 | 95.10 |
| C_AC1238 | Aichi Cancer Center | Kansai | 399,679,500 | 398,155,715 | 59,951,925,000 | 59,842,123,722 | 99.62 | 8.04 | 19.95 | 97.05 |
| C_AC1253 | Aichi Cancer Center | Kansai | 412,947,973 | 411,100,747 | 61,942,195,950 | 61,787,339,284 | 99.55 | 7.57 | 20.60 | 97.67 |
| C_AC1498 | Aichi Cancer Center | Kansai | 418,281,175 | 416,388,994 | 62,742,176,250 | 62,588,564,654 | 99.55 | 10.26 | 20.86 | 97.32 |
| C_AC1500 | Aichi Cancer Center | Kansai | 375,887,084 | 374,104,925 | 56,383,062,600 | 56,246,532,080 | 99.53 | 9.65 | 18.75 | 94.85 |

| | | | | | | | | | | |
|---|---|---|---|---|---|---|---|---|---|---|
| C_AC1512 | Aichi Cancer Center | Kansai | 385,945,724 | 383,735,632 | 57,891,858,600 | 57,696,497,103 | 99.43 | 10.40 | 19.23 | 95.29 |
| C_AC1517 | Aichi Cancer Center | Kansai | 420,641,994 | 418,521,123 | 63,096,299,100 | 62,915,657,539 | 99.50 | 11.59 | 20.97 | 97.07 |
| C_AC1521 | Aichi Cancer Center | Kansai | 393,934,263 | 392,067,931 | 59,090,139,450 | 58,936,033,660 | 99.53 | 10.14 | 19.65 | 92.83 |
| C_AC1522 | Aichi Cancer Center | Kansai | 395,764,038 | 393,159,842 | 59,364,605,700 | 59,081,593,451 | 99.34 | 10.36 | 19.69 | 95.60 |
| C_AC1526 | Aichi Cancer Center | Kansai | 407,514,075 | 405,718,667 | 61,127,111,250 | 61,003,635,948 | 99.56 | 9.49 | 20.33 | 97.10 |
| C_AC1531 | Aichi Cancer Center | Kansai | 408,484,446 | 406,500,363 | 61,272,666,900 | 61,112,428,565 | 99.51 | 8.63 | 20.37 | 97.19 |
| C_AC1533 | Aichi Cancer Center | Kansai | 378,860,614 | 376,649,036 | 56,829,092,100 | 56,631,405,401 | 99.42 | 9.23 | 18.88 | 95.23 |
| C_AC1537 | Aichi Cancer Center | Kansai | 411,242,793 | 409,144,773 | 61,686,418,950 | 61,515,208,440 | 99.49 | 9.19 | 20.51 | 97.18 |
| C_AC1539 | Aichi Cancer Center | Kansai | 401,896,910 | 400,217,961 | 60,284,536,500 | 60,182,313,415 | 99.58 | 8.86 | 20.06 | 96.97 |
| C_AC1548 | Aichi Cancer Center | Kansai | 367,598,506 | 365,647,738 | 55,139,775,900 | 54,961,509,659 | 99.47 | 9.02 | 18.32 | 94.18 |
| C_AC1549 | Aichi Cancer Center | Kansai | 400,032,255 | 397,429,333 | 60,004,838,250 | 59,736,700,588 | 99.35 | 8.70 | 19.91 | 96.66 |
| C_AC1564 | Aichi Cancer Center | Kansai | 408,663,588 | 405,684,811 | 61,299,538,200 | 61,005,055,135 | 99.27 | 9.15 | 20.34 | 97.08 |
| C_AC1566 | Aichi Cancer Center | Kansai | 430,016,856 | 426,880,227 | 64,502,528,400 | 64,179,924,292 | 99.27 | 10.25 | 21.39 | 97.72 |
| C_AC1577 | Aichi Cancer Center | Kansai | 449,168,100 | 447,097,800 | 67,375,215,000 | 67,236,078,213 | 99.54 | 9.82 | 22.41 | 98.42 |
| C_AC1579 | Aichi Cancer Center | Kansai | 398,944,771 | 395,369,587 | 59,841,715,650 | 59,435,000,122 | 99.10 | 7.35 | 19.81 | 93.16 |
| C_AC1586 | Aichi Cancer Center | Kansai | 454,578,386 | 451,929,507 | 68,186,757,900 | 67,932,184,624 | 99.42 | 8.82 | 22.64 | 98.46 |
| C_AC1587 | Aichi Cancer Center | Kansai | 429,119,485 | 426,801,236 | 64,367,922,750 | 64,141,406,008 | 99.46 | 7.96 | 21.38 | 97.94 |
| C_AC1590 | Aichi Cancer Center | Kansai | 434,185,872 | 431,477,633 | 65,127,880,800 | 64,838,868,532 | 99.38 | 8.54 | 21.61 | 97.95 |
| C_AC1595 | Aichi Cancer Center | Kansai | 423,526,696 | 420,868,532 | 63,529,004,400 | 63,237,176,097 | 99.37 | 7.51 | 21.08 | 97.52 |
| C_AC1604 | Aichi Cancer Center | Kansai | 477,978,306 | 475,582,810 | 71,696,745,900 | 71,500,807,785 | 99.50 | 9.14 | 23.83 | 98.85 |
| C_AC1606 | Aichi Cancer Center | Kansai | 402,360,184 | 399,365,975 | 60,354,027,600 | 60,034,798,119 | 99.26 | 7.67 | 20.01 | 96.70 |
| C_AC1612 | Aichi Cancer Center | Kansai | 390,059,555 | 387,976,850 | 58,508,933,250 | 58,310,530,498 | 99.47 | 6.58 | 19.44 | 96.18 |
| C_AC1613 | Aichi Cancer Center | Kansai | 391,377,568 | 389,072,476 | 58,706,635,200 | 58,500,045,329 | 99.41 | 7.09 | 19.50 | 96.26 |
| C_AC1614 | Aichi Cancer Center | Kansai | 424,118,056 | 422,145,221 | 63,617,708,400 | 63,458,017,978 | 99.53 | 8.90 | 21.15 | 97.61 |
| C_AC1623 | Aichi Cancer Center | Kansai | 416,341,094 | 414,873,931 | 62,451,164,100 | 62,394,252,560 | 99.65 | 8.31 | 20.80 | 97.75 |
| C_AC1634 | Aichi Cancer Center | Kansai | 382,167,725 | 380,374,562 | 57,325,158,750 | 57,173,296,012 | 99.53 | 8.44 | 19.06 | 95.57 |
| C_AC1657 | Aichi Cancer Center | Kansai | 423,671,009 | 421,995,679 | 63,550,651,350 | 63,429,870,548 | 99.60 | 8.85 | 21.14 | 97.80 |
| C_AC1661 | Aichi Cancer Center | Kansai | 405,460,640 | 404,003,138 | 60,819,096,000 | 60,740,795,713 | 99.64 | 7.87 | 20.25 | 97.38 |
| C_AC1665 | Aichi Cancer Center | Kansai | 393,696,186 | 392,262,184 | 59,054,427,900 | 58,959,344,581 | 99.64 | 8.73 | 19.65 | 96.49 |
| C_AC1671 | Aichi Cancer Center | Kansai | 472,600,311 | 470,813,367 | 70,890,046,650 | 70,781,787,885 | 99.62 | 8.51 | 23.59 | 98.93 |
| C_AC1675 | Aichi Cancer Center | Kansai | 424,512,269 | 423,129,227 | 63,676,840,350 | 63,609,268,776 | 99.67 | 8.18 | 21.20 | 98.02 |
| C_AC1679 | Aichi Cancer Center | Kansai | 390,883,919 | 389,325,865 | 58,632,587,850 | 58,520,656,331 | 99.60 | 8.68 | 19.51 | 96.25 |
| C_AC1681 | Aichi Cancer Center | Kansai | 398,126,049 | 396,102,615 | 59,718,907,350 | 59,528,892,013 | 99.49 | 8.15 | 19.84 | 96.63 |
| C_AC1682 | Aichi Cancer Center | Kansai | 402,985,930 | 400,993,983 | 60,447,889,500 | 60,274,319,756 | 99.51 | 8.89 | 20.09 | 96.87 |
| C_AC1685 | Aichi Cancer Center | Kansai | 672,378,188 | 668,305,322 | 100,856,728,200 | 100,452,751,025 | 99.39 | 9.71 | 33.48 | 99.66 |
| C_AC1690 | Aichi Cancer Center | Kansai | 641,319,494 | 637,854,741 | 96,197,924,100 | 95,873,273,365 | 99.46 | 8.86 | 31.96 | 99.62 |
| C_AC1692 | Aichi Cancer Center | Kansai | 626,371,587 | 620,696,227 | 93,955,738,050 | 93,304,964,828 | 99.09 | 8.69 | 31.10 | 99.59 |
| C_AC1702 | Aichi Cancer Center | Kansai | 723,547,980 | 719,503,448 | 108,532,197,000 | 108,152,725,406 | 99.44 | 9.41 | 36.05 | 99.67 |
| C_AC1703 | Aichi Cancer Center | Kansai | 637,259,181 | 633,594,718 | 95,588,877,150 | 95,249,303,475 | 99.42 | 8.73 | 31.75 | 99.60 |
| C_AC1704 | Aichi Cancer Center | Kansai | 649,616,049 | 645,340,639 | 97,442,407,350 | 97,000,031,217 | 99.34 | 8.72 | 32.33 | 99.61 |
| C_AC1706 | Aichi Cancer Center | Kansai | 713,445,389 | 709,064,911 | 107,016,808,350 | 106,586,379,717 | 99.39 | 8.62 | 35.53 | 99.70 |
| LCAC0002 | Aichi Cancer Center | Kansai | 367,243,296 | 365,485,623 | 55,086,494,400 | 54,960,310,890 | 99.52 | 7.42 | 18.32 | 91.98 |
| LCAC0004 | Aichi Cancer Center | Kansai | 376,802,702 | 374,898,968 | 56,520,405,300 | 56,364,851,933 | 99.49 | 8.08 | 18.79 | 92.52 |
| LCAC0006 | Aichi Cancer Center | Kansai | 399,791,951 | 397,785,369 | 59,968,792,650 | 59,815,826,297 | 99.50 | 7.81 | 19.94 | 94.27 |
| LCAC0008 | Aichi Cancer Center | Kansai | 392,017,352 | 390,317,915 | 58,802,602,800 | 58,692,187,600 | 99.57 | 6.23 | 19.56 | 94.09 |
| LCAC0017 | Aichi Cancer Center | Kansai | 388,939,549 | 387,328,515 | 58,340,932,350 | 58,237,515,361 | 99.59 | 6.29 | 19.41 | 93.93 |
| LCAC0023 | Aichi Cancer Center | Kansai | 368,454,542 | 366,692,798 | 55,268,181,300 | 55,152,608,450 | 99.52 | 5.75 | 18.38 | 92.75 |
| LCAC0027 | Aichi Cancer Center | Kansai | 383,361,740 | 381,460,109 | 57,504,261,000 | 57,356,582,213 | 99.50 | 6.96 | 19.12 | 93.32 |
| LCAC0034 | Aichi Cancer Center | Kansai | 388,289,008 | 386,148,412 | 58,243,351,200 | 58,069,410,877 | 99.45 | 6.89 | 19.36 | 93.76 |
| LCAC0035 | Aichi Cancer Center | Kansai | 367,564,696 | 365,953,338 | 55,134,704,400 | 55,015,705,098 | 99.56 | 6.03 | 18.34 | 92.57 |
| LCAC0040 | Aichi Cancer Center | Kansai | 376,715,077 | 374,786,120 | 56,507,261,550 | 56,364,742,938 | 99.49 | 6.01 | 18.79 | 93.05 |
| LCAC0044 | Aichi Cancer Center | Kansai | 443,747,495 | 441,530,566 | 66,562,124,250 | 66,389,412,665 | 99.50 | 6.60 | 22.13 | 96.23 |
| LCAC0055 | Aichi Cancer Center | Kansai | 353,849,877 | 352,000,701 | 53,077,481,550 | 52,935,885,114 | 99.48 | 5.45 | 17.65 | 91.48 |
| LCAC0060 | Aichi Cancer Center | Kansai | 374,585,313 | 372,744,189 | 56,187,796,950 | 56,048,069,709 | 99.51 | 6.28 | 18.68 | 92.97 |
| LCAC0062 | Aichi Cancer Center | Kansai | 424,719,929 | 422,592,561 | 63,707,989,350 | 63,538,553,244 | 99.50 | 6.61 | 21.18 | 95.55 |
| LCAC0076 | Aichi Cancer Center | Kansai | 417,506,381 | 415,603,015 | 62,625,957,150 | 62,498,773,838 | 99.54 | 6.31 | 20.83 | 98.08 |
| LCAC0081 | Aichi Cancer Center | Kansai | 385,074,056 | 382,905,850 | 57,761,108,400 | 57,583,718,315 | 99.44 | 6.30 | 19.19 | 93.66 |
| LCAC0093 | Aichi Cancer Center | Kansai | 468,903,821 | 465,737,281 | 70,335,573,150 | 70,022,228,501 | 99.32 | 6.35 | 23.34 | 98.83 |
| LCAC0094 | Aichi Cancer Center | Kansai | 351,552,313 | 349,617,767 | 52,732,846,950 | 52,561,887,167 | 99.45 | 5.45 | 17.52 | 91.08 |
| LCAC0096 | Aichi Cancer Center | Kansai | 430,121,665 | 428,332,887 | 64,518,249,750 | 64,386,885,197 | 99.58 | 6.76 | 21.46 | 95.19 |
| LCAC0102 | Aichi Cancer Center | Kansai | 390,421,433 | 388,454,346 | 58,563,214,950 | 58,393,481,637 | 99.50 | 6.52 | 19.46 | 93.86 |
| LCAC0105 | Aichi Cancer Center | Kansai | 394,852,840 | 392,852,028 | 59,227,926,000 | 59,065,122,268 | 99.49 | 6.26 | 19.69 | 97.15 |
| LCAC0106 | Aichi Cancer Center | Kansai | 372,884,043 | 370,975,238 | 55,932,606,450 | 55,763,750,803 | 99.49 | 5.72 | 18.59 | 92.78 |
| LCAC0109 | Aichi Cancer Center | Kansai | 368,373,059 | 366,543,312 | 55,255,958,850 | 55,110,313,455 | 99.50 | 6.83 | 18.37 | 92.16 |
| LCAC0110 | Aichi Cancer Center | Kansai | 406,627,796 | 404,719,502 | 60,994,169,400 | 60,861,263,601 | 99.53 | 7.12 | 20.29 | 97.61 |
| LCAC0111 | Aichi Cancer Center | Kansai | 371,051,783 | 369,205,788 | 55,657,767,450 | 55,512,411,434 | 99.50 | 6.18 | 18.50 | 92.59 |
| LCAC0119 | Aichi Cancer Center | Kansai | 341,016,729 | 339,200,575 | 51,152,509,350 | 51,000,595,527 | 99.47 | 5.76 | 17.00 | 92.70 |
| LCAC0130 | Aichi Cancer Center | Kansai | 358,003,382 | 355,963,317 | 53,700,507,300 | 53,531,417,723 | 99.43 | 5.65 | 17.84 | 94.59 |
| LCAC0134 | Aichi Cancer Center | Kansai | 334,519,284 | 332,644,990 | 50,177,892,600 | 50,014,868,780 | 99.44 | 5.44 | 16.67 | 88.79 |
| LCAC0139 | Aichi Cancer Center | Kansai | 321,515,215 | 319,454,991 | 48,227,282,250 | 48,028,397,967 | 99.36 | 4.83 | 16.01 | 87.01 |
| LCAC0149 | Aichi Cancer Center | Kansai | 320,847,907 | 318,916,449 | 48,127,186,050 | 47,942,251,738 | 99.40 | 4.70 | 15.98 | 86.88 |
| LCAC0156 | Aichi Cancer Center | Kansai | 349,213,668 | 347,342,260 | 52,382,050,200 | 52,224,281,600 | 99.46 | 6.28 | 17.41 | 93.44 |
| LCAC0158 | Aichi Cancer Center | Kansai | 468,155,482 | 465,540,137 | 70,223,322,300 | 69,987,647,612 | 99.44 | 8.48 | 23.33 | 96.28 |
| LCAC0160 | Aichi Cancer Center | Kansai | 488,941,642 | 486,717,935 | 73,341,246,300 | 73,163,891,340 | 99.55 | 8.98 | 24.39 | 96.97 |
| LCAC0164 | Aichi Cancer Center | Kansai | 348,971,922 | 348,079,443 | 52,345,788,300 | 52,349,321,265 | 99.74 | 5.70 | 17.45 | 91.31 |
| LCAC0178 | Aichi Cancer Center | Kansai | 346,320,833 | 345,269,557 | 51,948,124,950 | 51,905,065,186 | 99.70 | 5.93 | 17.30 | 90.79 |
| LCAC0179 | Aichi Cancer Center | Kansai | 364,973,864 | 364,052,814 | 54,746,079,600 | 54,738,226,347 | 99.75 | 6.16 | 18.25 | 95.64 |
| LCAC0183 | Aichi Cancer Center | Kansai | 334,029,732 | 333,014,375 | 50,104,459,800 | 50,057,333,910 | 99.70 | 5.32 | 16.69 | 89.38 |
| LCAC0188 | Aichi Cancer Center | Kansai | 368,323,223 | 367,051,344 | 55,248,483,450 | 55,171,469,783 | 99.65 | 5.09 | 18.39 | 96.07 |
| LCAC0189 | Aichi Cancer Center | Kansai | 379,939,034 | 378,997,869 | 56,990,855,100 | 56,968,315,333 | 99.75 | 6.24 | 18.99 | 93.63 |
| LCAC0194 | Aichi Cancer Center | Kansai | 339,616,436 | 338,701,884 | 50,942,465,400 | 50,934,386,752 | 99.73 | 5.53 | 16.98 | 90.27 |
| LCAC0196 | Aichi Cancer Center | Kansai | 356,649,995 | 355,582,264 | 53,497,499,250 | 53,453,440,826 | 99.70 | 5.62 | 17.82 | 95.00 |
| LCAC0197 | Aichi Cancer Center | Kansai | 403,111,948 | 402,056,628 | 60,466,792,200 | 60,452,895,087 | 99.74 | 6.70 | 20.15 | 94.67 |
| LCAC0207 | Aichi Cancer Center | Kansai | 374,987,326 | 374,055,472 | 56,248,098,900 | 56,217,677,456 | 99.75 | 5.85 | 18.74 | 96.35 |
| LCAC0220 | Aichi Cancer Center | Kansai | 383,346,070 | 382,332,670 | 57,501,910,500 | 57,477,504,981 | 99.74 | 6.39 | 19.16 | 96.67 |
| LCAC0236 | Aichi Cancer Center | Kansai | 392,702,804 | 391,698,652 | 58,905,420,600 | 58,899,593,357 | 99.74 | 5.82 | 19.63 | 97.34 |
| LCAC0250 | Aichi Cancer Center | Kansai | 387,366,889 | 386,205,711 | 58,105,033,350 | 58,059,011,198 | 99.70 | 6.40 | 19.35 | 94.01 |
| LCAC0254 | Aichi Cancer Center | Kansai | 416,099,925 | 414,996,902 | 62,414,988,750 | 62,346,773,059 | 99.73 | 7.01 | 20.78 | 95.17 |
| LCAC0256 | Aichi Cancer Center | Kansai | 314,236,662 | 313,342,712 | 47,135,499,300 | 47,083,292,396 | 99.72 | 5.45 | 15.69 | 89.09 |
| LCAC0266 | Aichi Cancer Center | Kansai | 388,928,206 | 387,723,029 | 58,339,230,900 | 58,285,357,518 | 99.69 | 6.10 | 19.43 | 94.21 |
| LCAC0276 | Aichi Cancer Center | Kansai | 402,319,083 | 401,314,908 | 60,347,862,450 | 60,335,741,505 | 99.75 | 7.29 | 20.11 | 97.34 |
| LCAC0281 | Aichi Cancer Center | Kansai | 366,446,159 | 365,416,881 | 54,966,923,850 | 54,944,957,318 | 99.72 | 5.87 | 18.31 | 92.74 |
| LCAC0282 | Aichi Cancer Center | Kansai | 360,836,113 | 359,754,749 | 54,125,416,950 | 54,086,807,550 | 99.70 | 6.07 | 18.03 | 92.11 |
| LCAC0295 | Aichi Cancer Center | Kansai | 375,611,285 | 374,509,740 | 56,341,692,750 | 56,295,373,771 | 99.71 | 5.93 | 18.77 | 96.34 |
| LCAC0299 | Aichi Cancer Center | Kansai | 422,917,895 | 421,370,151 | 63,437,684,250 | 63,328,955,239 | 99.63 | 7.10 | 21.11 | 94.87 |
| LCAC0306 | Aichi Cancer Center | Kansai | 421,495,218 | 420,284,930 | 63,224,282,700 | 63,176,850,423 | 99.71 | 6.58 | 21.06 | 95.68 |
| LCAC0308 | Aichi Cancer Center | Kansai | 424,348,408 | 423,142,392 | 63,652,261,200 | 63,611,598,794 | 99.72 | 6.61 | 21.20 | 95.72 |
| LCAC0311 | Aichi Cancer Center | Kansai | 365,168,061 | 363,992,024 | 54,775,209,150 | 54,727,383,790 | 99.68 | 5.83 | 18.24 | 92.64 |
| LCAC0314 | Aichi Cancer Center | Kansai | 410,623,381 | 409,169,245 | 61,593,507,150 | 61,529,308,533 | 99.65 | 6.71 | 20.51 | 94.99 |
| LCAC0321 | Aichi Cancer Center | Kansai | 373,871,801 | 372,830,320 | 56,080,770,150 | 56,044,289,261 | 99.72 | 6.62 | 18.68 | 96.00 |

| | | | | | | | | | | |
|---|---|---|---|---|---|---|---|---|---|---|
| LCAC0323 | Aichi Cancer Center | Kansai | 354,237,340 | 352,902,221 | 53,135,601,000 | 53,045,984,648 | 99.62 | 7.83 | 17.68 | 90.73 |
| LCAC0325 | Aichi Cancer Center | Kansai | 335,242,281 | 333,951,590 | 50,286,342,150 | 50,192,210,040 | 99.61 | 6.13 | 16.73 | 88.97 |
| LCAC0327 | Aichi Cancer Center | Kansai | 339,226,418 | 337,607,212 | 50,883,962,700 | 50,742,106,467 | 99.52 | 6.02 | 16.91 | 89.59 |
| LCAC0343 | Aichi Cancer Center | Kansai | 328,242,929 | 326,816,141 | 49,236,439,350 | 49,137,580,027 | 99.57 | 5.97 | 16.38 | 91.18 |
| LCAC0345 | Aichi Cancer Center | Kansai | 323,472,948 | 322,391,983 | 48,520,942,200 | 48,466,565,646 | 99.67 | 5.59 | 16.16 | 87.73 |
| LCAC0346 | Aichi Cancer Center | Kansai | 338,010,469 | 336,621,070 | 50,701,570,350 | 50,609,501,936 | 99.59 | 6.16 | 16.87 | 92.63 |
| LCAC0347 | Aichi Cancer Center | Kansai | 345,138,452 | 343,849,286 | 51,770,767,800 | 51,721,395,693 | 99.63 | 6.00 | 17.24 | 90.81 |
| LCAC0355 | Aichi Cancer Center | Kansai | 343,071,004 | 341,875,889 | 51,460,650,600 | 51,395,979,963 | 99.65 | 6.43 | 17.13 | 90.10 |
| LCAC0359 | Aichi Cancer Center | Kansai | 386,189,893 | 384,990,615 | 57,928,483,950 | 57,871,886,919 | 99.69 | 6.32 | 19.29 | 93.80 |
| LCAC0360 | Aichi Cancer Center | Kansai | 358,433,696 | 356,907,099 | 53,765,054,400 | 53,641,837,566 | 99.57 | 5.78 | 17.88 | 91.71 |
| LCAC0362 | Aichi Cancer Center | Kansai | 376,679,600 | 375,071,233 | 56,501,940,000 | 56,380,372,502 | 99.57 | 6.31 | 18.79 | 93.06 |
| LCAC0367 | Aichi Cancer Center | Kansai | 361,474,876 | 359,915,339 | 54,221,231,400 | 54,120,633,930 | 99.57 | 6.01 | 18.04 | 92.06 |
| LCAC0372 | Aichi Cancer Center | Kansai | 322,800,685 | 321,346,920 | 48,420,102,750 | 48,307,481,502 | 99.55 | 5.33 | 16.10 | 90.44 |
| LCAC0374 | Aichi Cancer Center | Kansai | 409,238,469 | 407,504,366 | 61,385,770,350 | 61,257,662,107 | 99.58 | 6.60 | 20.42 | 94.96 |
| LCAC0389 | Aichi Cancer Center | Kansai | 380,102,245 | 378,464,095 | 57,015,336,750 | 56,879,813,404 | 99.57 | 6.38 | 18.96 | 93.16 |
| LCAC0390 | Aichi Cancer Center | Kansai | 377,269,333 | 375,734,900 | 56,590,399,950 | 56,481,750,649 | 99.59 | 6.11 | 18.83 | 93.22 |
| LCAC0392 | Aichi Cancer Center | Kansai | 434,344,033 | 433,114,108 | 65,151,604,950 | 65,117,119,573 | 99.72 | 11.27 | 21.71 | 94.09 |
| LCAC0399 | Aichi Cancer Center | Kansai | 475,385,227 | 473,806,798 | 71,307,784,050 | 71,231,263,265 | 99.67 | 6.85 | 23.74 | 97.21 |
| LCAC0410 | Aichi Cancer Center | Kansai | 435,155,768 | 433,429,292 | 65,273,365,200 | 65,170,621,999 | 99.60 | 5.31 | 21.72 | 96.15 |
| LCAC0411 | Aichi Cancer Center | Kansai | 401,014,787 | 399,548,373 | 60,152,218,050 | 60,070,375,870 | 99.63 | 5.46 | 20.02 | 97.59 |
| LCAC0419 | Aichi Cancer Center | Kansai | 382,046,544 | 380,467,757 | 57,306,981,600 | 57,196,659,051 | 99.59 | 5.08 | 19.07 | 93.73 |
| LCAC0420 | Aichi Cancer Center | Kansai | 478,246,518 | 476,589,935 | 71,736,977,700 | 71,638,226,396 | 99.65 | 7.76 | 23.88 | 96.82 |
| LCAC0422 | Aichi Cancer Center | Kansai | 457,207,993 | 455,556,466 | 68,581,198,950 | 68,480,587,551 | 99.64 | 7.38 | 22.83 | 96.50 |
| LCAC0429 | Aichi Cancer Center | Kansai | 444,232,495 | 442,420,270 | 66,634,874,250 | 66,501,319,697 | 99.59 | 6.19 | 22.17 | 96.29 |
| LCAC0441 | Aichi Cancer Center | Kansai | 364,579,851 | 363,125,344 | 54,686,977,650 | 54,567,407,161 | 99.60 | 6.85 | 18.19 | 90.98 |
| LCAC0443 | Aichi Cancer Center | Kansai | 409,677,429 | 407,746,557 | 61,451,614,350 | 61,281,444,046 | 99.53 | 8.49 | 20.43 | 94.56 |
| LCAC0446 | Aichi Cancer Center | Kansai | 410,193,735 | 408,529,937 | 61,529,060,250 | 61,404,805,869 | 99.59 | 8.68 | 20.47 | 94.63 |
| LCAC0451 | Aichi Cancer Center | Kansai | 392,435,679 | 391,194,311 | 58,865,351,850 | 58,799,116,542 | 99.68 | 8.11 | 19.60 | 96.82 |
| LCAC0453 | Aichi Cancer Center | Kansai | 393,659,353 | 390,192,589 | 59,048,902,950 | 58,648,825,988 | 99.12 | 7.49 | 19.55 | 93.74 |
| LCAC0454 | Aichi Cancer Center | Kansai | 403,316,376 | 401,716,214 | 60,497,456,400 | 60,382,179,558 | 99.60 | 8.59 | 20.13 | 94.34 |
| LCAC0459 | Aichi Cancer Center | Kansai | 406,968,236 | 405,236,254 | 61,045,235,400 | 60,925,761,658 | 99.57 | 7.84 | 20.31 | 94.73 |
| LCAC0467 | Aichi Cancer Center | Kansai | 387,132,572 | 385,517,111 | 58,069,885,800 | 57,940,704,411 | 99.58 | 7.75 | 19.31 | 96.56 |
| LCAC0471 | Aichi Cancer Center | Kansai | 392,418,692 | 390,510,353 | 58,862,803,800 | 58,674,414,065 | 99.51 | 9.35 | 19.56 | 93.11 |
| LCAC0472 | Aichi Cancer Center | Kansai | 368,834,829 | 367,173,035 | 55,325,224,350 | 55,214,808,867 | 99.55 | 8.20 | 18.40 | 95.23 |
| LCAC0474 | Aichi Cancer Center | Kansai | 366,961,257 | 365,485,756 | 55,044,188,550 | 54,910,730,448 | 99.60 | 8.91 | 18.30 | 94.54 |
| LCAC0477 | Aichi Cancer Center | Kansai | 369,768,366 | 367,901,017 | 55,465,254,900 | 55,294,297,007 | 99.49 | 8.32 | 18.43 | 91.85 |
| LCAC0487 | Aichi Cancer Center | Kansai | 408,298,585 | 405,154,078 | 61,244,787,750 | 60,891,722,412 | 99.23 | 8.75 | 20.30 | 94.25 |
| LCAC0489 | Aichi Cancer Center | Kansai | 385,242,668 | 382,834,323 | 57,786,400,200 | 57,549,453,328 | 99.37 | 9.58 | 19.18 | 92.80 |
| LCAC0493 | Aichi Cancer Center | Kansai | 402,426,088 | 400,391,139 | 60,363,913,200 | 60,121,448,115 | 99.49 | 9.52 | 20.04 | 93.68 |
| LCAC0498 | Aichi Cancer Center | Kansai | 383,406,522 | 378,422,770 | 57,510,978,300 | 56,887,121,060 | 98.70 | 8.15 | 18.96 | 92.59 |
| LCAC0499 | Aichi Cancer Center | Kansai | 421,971,096 | 418,828,622 | 63,295,664,400 | 62,910,580,315 | 99.26 | 11.19 | 20.97 | 93.98 |
| LCAC0506 | Aichi Cancer Center | Kansai | 368,077,175 | 366,267,733 | 55,211,576,250 | 55,088,845,970 | 99.51 | 7.83 | 18.36 | 95.20 |
| LCAC0511 | Aichi Cancer Center | Kansai | 367,728,374 | 365,819,426 | 55,159,256,100 | 54,988,313,352 | 99.48 | 8.46 | 18.33 | 94.80 |
| LCAC0514 | Aichi Cancer Center | Kansai | 399,496,486 | 397,471,290 | 59,924,472,900 | 59,751,216,268 | 99.49 | 9.00 | 19.92 | 93.92 |
| LCAC0521 | Aichi Cancer Center | Kansai | 378,099,227 | 376,410,159 | 56,714,884,050 | 56,603,256,779 | 99.55 | 4.95 | 18.87 | 96.56 |
| LCAC0522 | Aichi Cancer Center | Kansai | 402,548,604 | 400,692,134 | 60,382,290,600 | 60,219,689,937 | 99.54 | 5.34 | 20.07 | 97.59 |
| LCAC0523 | Aichi Cancer Center | Kansai | 361,587,534 | 360,379,446 | 54,238,130,100 | 54,176,208,850 | 99.67 | 5.48 | 18.06 | 92.19 |
| LCAC0526 | Aichi Cancer Center | Kansai | 349,333,998 | 347,830,689 | 52,400,099,700 | 52,289,768,509 | 99.57 | 4.99 | 17.43 | 91.02 |
| LCAC0527 | Aichi Cancer Center | Kansai | 367,139,869 | 365,884,867 | 55,070,980,350 | 54,988,875,316 | 99.66 | 5.61 | 18.33 | 92.53 |
| LCAC0530 | Aichi Cancer Center | Kansai | 330,745,768 | 329,619,600 | 49,611,865,200 | 49,558,027,721 | 99.66 | 4.98 | 16.52 | 92.22 |
| LCAC0536 | Aichi Cancer Center | Kansai | 307,614,615 | 306,196,335 | 46,142,192,250 | 46,032,989,284 | 99.54 | 4.96 | 15.34 | 84.88 |
| LCAC0537 | Aichi Cancer Center | Kansai | 372,900,555 | 371,855,090 | 55,935,083,250 | 55,909,711,460 | 99.72 | 5.83 | 18.64 | 96.23 |
| LCAC0541 | Aichi Cancer Center | Kansai | 339,014,121 | 337,625,377 | 50,852,118,150 | 50,768,366,934 | 99.59 | 4.64 | 16.92 | 90.16 |
| LCAC0549 | Aichi Cancer Center | Kansai | 355,399,739 | 353,979,765 | 53,309,960,850 | 53,218,630,286 | 99.60 | 5.41 | 17.74 | 91.64 |
| LCAC0550 | Aichi Cancer Center | Kansai | 411,370,876 | 409,946,012 | 61,705,631,400 | 61,615,154,880 | 99.65 | 5.55 | 20.54 | 95.28 |
| LCAC0552 | Aichi Cancer Center | Kansai | 373,943,437 | 372,508,364 | 56,091,515,550 | 55,981,781,379 | 99.62 | 4.51 | 18.66 | 93.36 |
| LCAC0553 | Aichi Cancer Center | Kansai | 325,772,790 | 324,707,044 | 48,865,918,500 | 48,820,933,848 | 99.67 | 4.98 | 16.27 | 91.63 |
| LCAC0566 | Aichi Cancer Center | Kansai | 332,836,996 | 331,239,828 | 49,925,549,400 | 49,802,463,103 | 99.52 | 5.06 | 16.60 | 92.30 |
| LCAC0571 | Aichi Cancer Center | Kansai | 391,939,771 | 389,835,615 | 58,790,965,650 | 58,600,180,942 | 99.46 | 5.23 | 19.53 | 94.35 |
| LCAC0575 | Aichi Cancer Center | Kansai | 340,956,707 | 339,456,197 | 51,143,506,050 | 51,034,153,866 | 99.56 | 5.06 | 17.01 | 90.14 |
| LCAC0577 | Aichi Cancer Center | Kansai | 325,796,693 | 324,381,529 | 48,869,503,950 | 48,772,260,178 | 99.57 | 4.49 | 16.26 | 88.28 |
| LCAC0581 | Aichi Cancer Center | Kansai | 383,939,569 | 381,408,667 | 57,590,935,350 | 57,327,687,078 | 99.34 | 5.47 | 19.11 | 93.71 |
| LCAC0586 | Aichi Cancer Center | Kansai | 372,415,649 | 370,829,562 | 55,862,347,350 | 55,739,242,959 | 99.57 | 5.05 | 18.58 | 93.10 |
| LCAC0588 | Aichi Cancer Center | Kansai | 342,473,991 | 341,046,251 | 51,371,098,650 | 51,275,484,421 | 99.58 | 5.32 | 17.09 | 93.48 |
| LCAC0612 | Aichi Cancer Center | Kansai | 361,132,492 | 359,776,784 | 54,169,873,800 | 54,066,558,946 | 99.62 | 4.94 | 18.02 | 92.30 |
| LCAC0614 | Aichi Cancer Center | Kansai | 414,260,774 | 412,678,080 | 62,139,116,100 | 62,037,632,652 | 99.62 | 4.95 | 20.68 | 98.08 |
| LCAC0616 | Aichi Cancer Center | Kansai | 378,206,946 | 376,835,268 | 56,731,041,900 | 56,651,746,689 | 99.64 | 4.90 | 18.88 | 96.70 |
| LCAC0623 | Aichi Cancer Center | Kansai | 351,754,751 | 350,129,995 | 52,763,212,650 | 52,619,761,516 | 99.54 | 4.60 | 17.54 | 91.64 |
| LCAC0629 | Aichi Cancer Center | Kansai | 340,697,760 | 338,541,334 | 51,104,664,000 | 50,886,505,352 | 99.37 | 3.70 | 16.96 | 90.38 |
| LCAC0630 | Aichi Cancer Center | Kansai | 430,757,854 | 428,653,579 | 64,613,678,100 | 64,388,607,367 | 99.51 | 4.62 | 21.46 | 98.45 |
| LCAC0639 | Aichi Cancer Center | Kansai | 390,062,765 | 387,595,005 | 58,509,414,750 | 58,256,075,937 | 99.37 | 4.70 | 19.42 | 94.21 |
| LCAC0645 | Aichi Cancer Center | Kansai | 378,303,337 | 376,938,411 | 56,745,500,550 | 56,662,120,580 | 99.64 | 4.67 | 18.89 | 93.73 |
| LCAC0650 | Aichi Cancer Center | Kansai | 383,858,588 | 381,308,204 | 57,578,788,200 | 57,321,403,731 | 99.34 | 4.50 | 19.11 | 96.94 |
| LCAC0651 | Aichi Cancer Center | Kansai | 381,070,317 | 379,028,797 | 57,160,547,550 | 56,968,684,731 | 99.46 | 4.67 | 18.99 | 93.71 |
| LCAC0653 | Aichi Cancer Center | Kansai | 382,449,902 | 380,225,748 | 57,367,485,300 | 57,115,796,169 | 99.42 | 5.08 | 19.04 | 93.01 |
| LCAC0655 | Aichi Cancer Center | Kansai | 353,509,377 | 351,420,108 | 53,026,406,550 | 52,821,896,081 | 99.41 | 4.83 | 17.61 | 94.61 |
| LCAC0668 | Aichi Cancer Center | Kansai | 446,123,029 | 444,229,348 | 66,918,454,350 | 66,773,888,806 | 99.58 | 4.57 | 22.26 | 96.53 |
| LCAC0669 | Aichi Cancer Center | Kansai | 394,287,009 | 392,480,053 | 59,143,051,350 | 59,005,426,845 | 99.54 | 4.85 | 19.67 | 94.49 |
| LCAC0672 | Aichi Cancer Center | Kansai | 417,535,381 | 415,550,033 | 62,630,307,150 | 62,443,641,126 | 99.52 | 4.84 | 20.81 | 95.58 |
| LCAC0674 | Aichi Cancer Center | Kansai | 348,439,484 | 347,298,567 | 52,265,922,600 | 52,213,566,231 | 99.67 | 4.92 | 17.40 | 91.12 |
| LCAC0690 | Aichi Cancer Center | Kansai | 390,524,008 | 389,307,534 | 58,578,601,200 | 58,537,232,656 | 99.69 | 5.54 | 19.51 | 97.19 |
| LCAC0691 | Aichi Cancer Center | Kansai | 353,338,638 | 352,026,462 | 53,000,795,700 | 52,918,545,382 | 99.63 | 5.16 | 17.64 | 91.48 |
| LCAC0700 | Aichi Cancer Center | Kansai | 348,534,787 | 347,226,319 | 52,280,218,050 | 52,192,631,101 | 99.62 | 4.76 | 17.40 | 91.12 |
| LCAC0702 | Aichi Cancer Center | Kansai | 335,932,598 | 334,685,447 | 50,389,889,700 | 50,311,843,867 | 99.63 | 5.03 | 16.77 | 89.65 |
| LCAC0705 | Aichi Cancer Center | Kansai | 358,488,140 | 357,359,195 | 53,773,221,000 | 53,734,650,643 | 99.69 | 4.25 | 17.91 | 92.39 |
| LCAC0706 | Aichi Cancer Center | Kansai | 337,663,575 | 336,407,566 | 50,649,536,250 | 50,573,503,198 | 99.63 | 3.95 | 16.86 | 90.07 |
| LCAC0708 | Aichi Cancer Center | Kansai | 348,377,980 | 346,826,599 | 52,256,697,000 | 52,138,153,239 | 99.55 | 3.75 | 17.38 | 91.23 |
| LCAC0711 | Aichi Cancer Center | Kansai | 377,706,548 | 376,294,829 | 56,655,982,200 | 56,575,309,862 | 99.63 | 4.43 | 18.86 | 96.71 |
| LCAC0713 | Aichi Cancer Center | Kansai | 368,195,454 | 367,046,140 | 55,229,318,100 | 55,161,690,175 | 99.69 | 4.81 | 18.39 | 92.75 |
| LCAC0718 | Aichi Cancer Center | Kansai | 320,423,021 | 319,419,657 | 48,063,453,150 | 48,016,377,516 | 99.69 | 3.90 | 16.01 | 91.06 |
| LCAC0719 | Aichi Cancer Center | Kansai | 329,518,133 | 327,836,457 | 49,427,719,950 | 49,301,804,977 | 99.49 | 3.84 | 16.43 | 92.40 |
| LCAC0726 | Aichi Cancer Center | Kansai | 631,774,825 | 626,893,028 | 94,766,223,750 | 94,234,480,636 | 99.23 | 7.73 | 31.41 | 98.97 |
| LCAC0727 | Aichi Cancer Center | Kansai | 355,698,811 | 353,601,570 | 53,354,821,650 | 53,171,703,773 | 99.41 | 4.20 | 17.72 | 95.01 |
| LCAC0732 | Aichi Cancer Center | Kansai | 357,787,163 | 355,863,795 | 53,668,074,450 | 53,516,439,062 | 99.46 | 4.12 | 17.84 | 95.09 |
| LCAC0736 | Aichi Cancer Center | Kansai | 360,864,120 | 359,595,765 | 54,129,618,000 | 54,068,229,276 | 99.65 | 4.29 | 18.02 | 92.50 |
| LCAC0738 | Aichi Cancer Center | Kansai | 343,450,377 | 341,869,109 | 51,517,556,550 | 51,394,870,811 | 99.54 | 4.84 | 17.13 | 90.44 |
| LCAC0771 | Aichi Cancer Center | Kansai | 347,197,379 | 345,619,086 | 52,079,606,850 | 51,945,424,917 | 99.55 | 4.28 | 17.32 | 90.90 |
| LCAC0775 | Aichi Cancer Center | Kansai | 309,675,484 | 308,398,331 | 46,451,322,600 | 46,371,783,931 | 99.59 | 4.76 | 15.46 | 85.49 |
| LCAC0779 | Aichi Cancer Center | Kansai | 651,830,087 | 647,399,169 | 97,774,513,050 | 97,303,231,562 | 99.32 | 7.37 | 32.43 | 99.05 |
| LCAC0782 | Aichi Cancer Center | Kansai | 339,366,034 | 337,847,666 | 50,904,905,100 | 50,799,620,521 | 99.55 | 4.29 | 16.93 | 90.31 |

| | | | | | | | | | | |
|---|---|---|---|---|---|---|---|---|---|---|
| LCAC0786 | Aichi Cancer Center | Kansai | 358,773,906 | 357,400,434 | 53,816,085,900 | 53,739,119,696 | 99.62 | 4.38 | 17.91 | 95.25 |
| LCAC0795 | Aichi Cancer Center | Kansai | 345,125,351 | 343,583,332 | 51,768,802,650 | 51,665,376,029 | 99.55 | 4.27 | 17.22 | 93.89 |
| LCAC0802 | Aichi Cancer Center | Kansai | 351,130,728 | 349,800,095 | 52,669,609,200 | 52,576,445,340 | 99.62 | 4.39 | 17.53 | 91.57 |
| LCAC0805 | Aichi Cancer Center | Kansai | 350,158,814 | 348,328,034 | 52,523,822,100 | 52,385,349,604 | 99.48 | 3.88 | 17.46 | 94.74 |
| LCAC0809 | Aichi Cancer Center | Kansai | 403,667,605 | 402,017,855 | 60,550,140,750 | 60,403,259,819 | 99.59 | 7.60 | 20.13 | 93.51 |
| LCAC0820 | Aichi Cancer Center | Kansai | 400,533,967 | 398,517,930 | 60,080,095,050 | 59,917,527,428 | 99.50 | 4.29 | 19.97 | 97.58 |
| LCAC0826 | Aichi Cancer Center | Kansai | 357,312,440 | 354,951,886 | 53,596,866,000 | 53,359,383,811 | 99.34 | 4.24 | 17.79 | 91.91 |
| LCAC0827 | Aichi Cancer Center | Kansai | 327,631,291 | 325,981,042 | 49,144,693,650 | 49,012,328,956 | 99.50 | 5.66 | 16.34 | 91.02 |
| LCAC0828 | Aichi Cancer Center | Kansai | 342,199,765 | 340,481,534 | 51,329,964,750 | 51,181,563,140 | 99.50 | 5.23 | 17.06 | 93.31 |
| LCAC0835 | Aichi Cancer Center | Kansai | 370,836,353 | 369,049,082 | 55,625,452,950 | 55,484,409,217 | 99.52 | 5.99 | 18.49 | 92.74 |
| LCAC0838 | Aichi Cancer Center | Kansai | 354,872,846 | 352,975,345 | 53,230,926,900 | 53,061,757,796 | 99.47 | 4.94 | 17.69 | 91.74 |
| LCAC0845 | Aichi Cancer Center | Kansai | 375,867,698 | 373,594,673 | 56,380,154,700 | 56,147,742,024 | 99.40 | 4.79 | 18.72 | 93.15 |
| LCAC0848 | Aichi Cancer Center | Kansai | 594,513,123 | 590,445,467 | 89,176,968,450 | 88,768,019,642 | 99.32 | 8.03 | 29.59 | 99.52 |
| LCAC0853 | Aichi Cancer Center | Kansai | 698,028,888 | 693,044,464 | 104,704,333,200 | 104,162,733,542 | 99.29 | 8.56 | 34.72 | 99.32 |
| LCAC0864 | Aichi Cancer Center | Kansai | 570,743,642 | 567,330,488 | 85,611,546,300 | 85,302,749,127 | 99.40 | 7.84 | 28.43 | 98.40 |
| LCAC0866 | Aichi Cancer Center | Kansai | 618,133,631 | 613,221,974 | 92,720,044,650 | 92,182,236,579 | 99.21 | 7.14 | 30.73 | 98.78 |
| LCAC0867 | Aichi Cancer Center | Kansai | 653,899,246 | 649,484,113 | 98,084,886,900 | 97,620,442,031 | 99.32 | 8.15 | 32.54 | 99.04 |
| LCAC0869 | Aichi Cancer Center | Kansai | 629,665,317 | 625,592,041 | 94,449,797,550 | 94,059,217,606 | 99.35 | 7.09 | 31.35 | 98.99 |
| LCAC0874 | Aichi Cancer Center | Kansai | 675,391,166 | 670,810,853 | 101,308,674,900 | 100,858,519,991 | 99.32 | 7.90 | 33.62 | 99.68 |
| LCAC0880 | Aichi Cancer Center | Kansai | 669,190,399 | 664,851,634 | 100,378,559,850 | 99,958,294,799 | 99.35 | 7.62 | 33.32 | 99.65 |
| LCAC0881 | Aichi Cancer Center | Kansai | 419,295,520 | 417,143,624 | 62,894,328,000 | 62,723,740,468 | 99.49 | 5.85 | 20.91 | 95.45 |
| LCAC0882 | Aichi Cancer Center | Kansai | 412,447,264 | 409,907,806 | 61,867,089,600 | 61,594,940,874 | 99.38 | 8.69 | 20.53 | 94.23 |
| LCAC0891 | Aichi Cancer Center | Kansai | 396,928,269 | 394,700,292 | 59,539,240,350 | 59,333,564,652 | 99.44 | 7.24 | 19.78 | 96.81 |
| LCAC0895 | Aichi Cancer Center | Kansai | 411,485,201 | 409,129,879 | 61,722,780,150 | 61,506,001,038 | 99.43 | 7.91 | 20.50 | 97.32 |
| LCAC0901 | Aichi Cancer Center | Kansai | 411,380,906 | 409,338,206 | 61,707,135,900 | 61,531,078,695 | 99.50 | 8.95 | 20.51 | 97.24 |
| LCAC0902 | Aichi Cancer Center | Kansai | 360,996,549 | 359,328,105 | 54,149,482,350 | 54,019,454,799 | 99.54 | 8.35 | 18.01 | 93.74 |
| LCAC0903 | Aichi Cancer Center | Kansai | 377,085,359 | 374,233,863 | 56,562,803,850 | 56,278,533,779 | 99.24 | 8.23 | 18.76 | 92.02 |
| LCAC0910 | Aichi Cancer Center | Kansai | 382,253,637 | 380,286,504 | 57,338,045,550 | 57,151,557,591 | 99.49 | 8.61 | 19.05 | 95.44 |
| LCAC0912 | Aichi Cancer Center | Kansai | 372,400,914 | 370,809,731 | 55,860,137,100 | 55,738,355,790 | 99.57 | 7.83 | 18.58 | 92.03 |
| LCAC0921 | Aichi Cancer Center | Kansai | 422,906,195 | 420,681,641 | 63,435,929,250 | 63,234,431,543 | 99.47 | 8.67 | 21.08 | 94.97 |
| NAG10004 | Kyoto Univ. | Kansai | 307,450,563 | 306,363,952 | 46,117,584,450 | 46,012,749,023 | 99.65 | 4.72 | 15.34 | 84.53 |
| NAG10005 | Kyoto Univ. | Kansai | 369,138,912 | 367,010,958 | 55,370,836,800 | 55,145,896,741 | 99.42 | 6.91 | 18.38 | 94.21 |
| NAG10019 | Kyoto Univ. | Kansai | 346,815,099 | 345,070,562 | 52,022,264,850 | 51,823,163,821 | 99.50 | 6.63 | 17.27 | 92.31 |
| NAG10022 | Kyoto Univ. | Kansai | 417,912,181 | 415,491,079 | 62,686,827,150 | 62,340,030,633 | 99.42 | 4.84 | 20.78 | 97.74 |
| NAG10024 | Kyoto Univ. | Kansai | 338,584,235 | 336,603,692 | 50,787,635,250 | 50,538,958,717 | 99.42 | 5.43 | 16.85 | 91.58 |
| NAG10028 | Kyoto Univ. | Kansai | 348,063,858 | 346,465,401 | 52,209,578,700 | 52,057,993,347 | 99.54 | 7.52 | 17.35 | 92.16 |
| NAG10030 | Kyoto Univ. | Kansai | 432,278,366 | 430,569,207 | 64,841,754,900 | 64,644,029,186 | 99.60 | 7.39 | 21.55 | 97.51 |
| NAG10032 | Kyoto Univ. | Kansai | 397,150,713 | 394,018,188 | 59,572,606,950 | 59,180,514,763 | 99.21 | 5.17 | 19.73 | 97.14 |
| NAG10033 | Kyoto Univ. | Kansai | 330,608,493 | 328,702,222 | 49,591,273,950 | 49,357,785,284 | 99.42 | 5.25 | 16.45 | 90.21 |
| NAG10049 | Kyoto Univ. | Kansai | 406,267,515 | 404,211,056 | 60,940,127,250 | 60,678,008,486 | 99.49 | 5.63 | 20.23 | 97.20 |
| NAG10050 | Kyoto Univ. | Kansai | 401,229,338 | 399,809,588 | 60,184,400,700 | 60,033,260,652 | 99.65 | 5.83 | 20.01 | 94.10 |
| NAG10053 | Kyoto Univ. | Kansai | 388,538,750 | 385,485,310 | 58,280,812,500 | 57,836,635,991 | 99.21 | 5.94 | 19.28 | 92.85 |
| NAG10055 | Kyoto Univ. | Kansai | 304,123,768 | 302,445,612 | 45,618,565,200 | 45,398,124,144 | 99.45 | 3.90 | 15.13 | 86.69 |
| NAG10061 | Kyoto Univ. | Kansai | 332,643,068 | 331,313,478 | 49,896,460,200 | 49,783,262,662 | 99.60 | 6.40 | 16.59 | 90.91 |
| NAG10066 | Kyoto Univ. | Kansai | 380,027,901 | 376,907,878 | 57,004,185,150 | 56,635,171,453 | 99.18 | 5.87 | 18.88 | 95.98 |
| NAG10067 | Kyoto Univ. | Kansai | 384,661,369 | 382,927,079 | 57,699,205,350 | 57,486,668,205 | 99.55 | 5.04 | 19.16 | 96.47 |
| NAG10094 | Kyoto Univ. | Kansai | 332,440,746 | 330,895,460 | 49,866,111,900 | 49,688,652,548 | 99.54 | 6.51 | 16.56 | 90.19 |
| NAG10099 | Kyoto Univ. | Kansai | 355,492,381 | 354,253,157 | 53,323,857,150 | 53,222,098,455 | 99.65 | 5.82 | 17.74 | 91.14 |
| NAG10102 | Kyoto Univ. | Kansai | 369,515,491 | 367,523,075 | 55,427,323,650 | 55,160,196,733 | 99.46 | 4.83 | 18.39 | 95.08 |
| NAG10104 | Kyoto Univ. | Kansai | 335,104,465 | 332,254,946 | 50,265,669,750 | 49,892,200,577 | 99.15 | 5.58 | 16.63 | 91.24 |
| NAG10106 | Kyoto Univ. | Kansai | 382,982,431 | 379,130,967 | 57,447,364,650 | 56,891,257,156 | 98.99 | 4.62 | 18.96 | 95.92 |
| NAG10110 | Kyoto Univ. | Kansai | 522,418,535 | 519,719,193 | 78,362,780,250 | 78,028,513,241 | 99.48 | 9.88 | 26.01 | 97.29 |
| NAG10123 | Kyoto Univ. | Kansai | 336,879,503 | 335,430,778 | 50,531,925,450 | 50,355,478,033 | 99.57 | 4.05 | 16.79 | 89.58 |
| NAG10136 | Kyoto Univ. | Kansai | 432,731,377 | 427,632,102 | 64,909,706,550 | 64,158,985,273 | 98.82 | 5.76 | 21.39 | 95.17 |
| NAG10137 | Kyoto Univ. | Kansai | 356,770,771 | 355,276,535 | 53,515,615,650 | 53,382,276,117 | 99.58 | 6.51 | 17.79 | 93.97 |
| NAG10138 | Kyoto Univ. | Kansai | 370,350,865 | 367,900,764 | 55,552,629,750 | 55,236,369,846 | 99.34 | 5.94 | 18.41 | 95.14 |
| NAG10161 | Kyoto Univ. | Kansai | 350,456,181 | 349,276,586 | 52,568,427,150 | 52,491,348,415 | 99.66 | 5.17 | 17.50 | 94.37 |
| NAG10164 | Kyoto Univ. | Kansai | 648,278,273 | 641,581,349 | 97,241,740,950 | 96,329,121,038 | 98.97 | 7.21 | 32.11 | 99.49 |
| NAG10169 | Kyoto Univ. | Kansai | 377,300,730 | 374,860,081 | 56,595,109,500 | 56,299,394,611 | 99.35 | 6.58 | 18.77 | 91.47 |
| NAG10172 | Kyoto Univ. | Kansai | 341,046,067 | 339,855,492 | 51,156,910,050 | 51,082,456,679 | 99.65 | 7.25 | 17.03 | 89.44 |
| NAG10188 | Kyoto Univ. | Kansai | 394,872,765 | 391,682,635 | 59,230,914,750 | 58,871,088,426 | 99.19 | 7.85 | 19.62 | 95.79 |
| NAG10200 | Kyoto Univ. | Kansai | 437,573,606 | 435,917,325 | 65,636,040,900 | 65,499,242,527 | 99.62 | 5.00 | 21.83 | 98.36 |
| NAG10213 | Kyoto Univ. | Kansai | 340,988,078 | 339,586,296 | 51,148,211,700 | 51,004,213,084 | 99.59 | 4.44 | 17.00 | 93.53 |
| NAG10219 | Kyoto Univ. | Kansai | 342,816,393 | 340,556,782 | 51,422,458,950 | 51,045,363,234 | 99.34 | 3.09 | 17.02 | 92.18 |
| NAG10227 | Kyoto Univ. | Kansai | 344,600,782 | 343,243,094 | 51,690,117,300 | 51,583,140,591 | 99.61 | 6.15 | 17.19 | 92.90 |
| NAG10233 | Kyoto Univ. | Kansai | 326,805,616 | 322,163,830 | 49,020,842,400 | 48,360,356,713 | 98.58 | 4.49 | 16.12 | 89.01 |
| NAG10238 | Kyoto Univ. | Kansai | 376,329,412 | 373,683,014 | 56,449,411,800 | 56,129,023,452 | 99.30 | 2.90 | 18.71 | 96.18 |
| NAG10242 | Kyoto Univ. | Kansai | 386,659,111 | 385,232,626 | 57,998,866,650 | 57,885,610,547 | 99.63 | 4.13 | 19.30 | 97.24 |
| NAG10244 | Kyoto Univ. | Kansai | 437,765,458 | 435,298,555 | 65,664,818,700 | 65,393,921,449 | 99.44 | 8.07 | 21.80 | 97.75 |
| NAG10252 | Kyoto Univ. | Kansai | 361,394,065 | 359,773,932 | 54,209,109,750 | 53,965,376,809 | 99.55 | 7.14 | 17.99 | 93.09 |
| NAG10258 | Kyoto Univ. | Kansai | 338,719,002 | 337,313,701 | 50,807,850,300 | 50,661,568,848 | 99.59 | 4.81 | 16.89 | 92.92 |
| NAG10262 | Kyoto Univ. | Kansai | 352,405,224 | 349,273,097 | 52,860,783,600 | 52,430,516,372 | 99.11 | 4.59 | 17.48 | 93.82 |
| NAG10267 | Kyoto Univ. | Kansai | 338,722,067 | 337,445,054 | 50,808,310,050 | 50,684,692,735 | 99.62 | 7.56 | 16.89 | 91.24 |
| NAG10276 | Kyoto Univ. | Kansai | 367,752,232 | 366,651,893 | 55,162,834,800 | 55,070,330,601 | 99.70 | 7.35 | 18.36 | 95.01 |
| NAG10278 | Kyoto Univ. | Kansai | 320,880,539 | 318,928,684 | 48,132,080,850 | 47,914,951,716 | 99.39 | 3.19 | 15.97 | 87.29 |
| NAG10293 | Kyoto Univ. | Kansai | 380,496,569 | 379,217,110 | 57,074,485,350 | 56,978,576,459 | 99.66 | 7.54 | 18.99 | 92.80 |
| NAG10294 | Kyoto Univ. | Kansai | 317,826,032 | 315,950,812 | 47,673,904,800 | 47,431,408,525 | 99.41 | 3.45 | 15.81 | 89.06 |
| NAG10300 | Kyoto Univ. | Kansai | 313,561,383 | 312,180,915 | 47,034,207,450 | 46,846,054,916 | 99.56 | 5.18 | 15.62 | 87.01 |
| NAG10305 | Kyoto Univ. | Kansai | 347,677,625 | 346,425,471 | 52,151,643,750 | 52,047,088,236 | 99.64 | 4.07 | 17.35 | 94.57 |
| NAG10307 | Kyoto Univ. | Kansai | 342,477,678 | 340,611,584 | 51,371,651,700 | 51,169,355,518 | 99.46 | 5.22 | 17.06 | 89.02 |
| NAG10318 | Kyoto Univ. | Kansai | 411,991,272 | 410,575,184 | 61,798,690,800 | 61,641,753,211 | 99.66 | 8.17 | 20.55 | 94.31 |
| NAG10320 | Kyoto Univ. | Kansai | 356,953,987 | 355,310,733 | 53,543,098,050 | 53,341,691,491 | 99.54 | 5.34 | 17.78 | 93.77 |
| NAG10323 | Kyoto Univ. | Kansai | 373,006,284 | 371,505,528 | 55,950,942,600 | 55,803,796,999 | 99.60 | 3.89 | 18.60 | 96.43 |
| NAG10326 | Kyoto Univ. | Kansai | 356,536,686 | 354,351,626 | 53,480,502,900 | 53,166,995,810 | 99.39 | 4.30 | 17.72 | 93.97 |
| NAG10363 | Kyoto Univ. | Kansai | 333,761,368 | 332,444,840 | 50,064,205,200 | 49,887,762,079 | 99.61 | 6.69 | 16.63 | 89.49 |
| NAG10366 | Kyoto Univ. | Kansai | 338,856,193 | 337,077,196 | 50,828,428,950 | 50,585,111,636 | 99.47 | 4.13 | 16.86 | 92.40 |
| NAG10367 | Kyoto Univ. | Kansai | 337,136,119 | 335,273,806 | 50,570,417,850 | 50,359,394,495 | 99.45 | 4.56 | 16.79 | 92.30 |
| NAG10375 | Kyoto Univ. | Kansai | 319,607,189 | 317,718,219 | 47,941,078,350 | 47,715,037,091 | 99.41 | 4.68 | 15.91 | 89.02 |
| NAG10378 | Kyoto Univ. | Kansai | 320,605,138 | 318,917,530 | 48,090,770,700 | 47,960,784,679 | 99.47 | 5.67 | 15.99 | 89.53 |
| NAG10381 | Kyoto Univ. | Kansai | 429,705,443 | 427,547,539 | 64,455,816,450 | 64,198,783,236 | 99.50 | 6.96 | 21.40 | 97.27 |
| NAG10417 | Kyoto Univ. | Kansai | 350,514,957 | 349,089,568 | 52,577,243,550 | 52,407,883,167 | 99.59 | 5.93 | 17.47 | 92.73 |
| NAG10418 | Kyoto Univ. | Kansai | 374,917,273 | 373,697,854 | 56,237,590,950 | 56,172,351,655 | 99.67 | 7.05 | 18.72 | 95.91 |
| NAG10428 | Kyoto Univ. | Kansai | 327,612,406 | 326,127,932 | 49,141,860,900 | 48,973,723,371 | 99.55 | 5.30 | 16.32 | 90.13 |
| NAG10431 | Kyoto Univ. | Kansai | 377,646,469 | 375,222,129 | 56,646,970,350 | 56,326,760,875 | 99.36 | 5.19 | 18.78 | 95.77 |
| NAG10438 | Kyoto Univ. | Kansai | 343,749,712 | 341,881,132 | 51,562,456,800 | 51,271,775,745 | 99.46 | 3.89 | 17.09 | 92.92 |
| NAG10441 | Kyoto Univ. | Kansai | 348,490,196 | 347,234,403 | 52,273,529,400 | 52,181,939,053 | 99.64 | 5.17 | 17.39 | 94.24 |
| NAG10464 | Kyoto Univ. | Kansai | 347,066,180 | 345,836,390 | 52,059,927,000 | 51,962,554,091 | 99.65 | 4.39 | 17.32 | 94.31 |
| NAG10488 | Kyoto Univ. | Kansai | 349,996,441 | 348,917,238 | 52,499,466,150 | 52,425,104,971 | 99.69 | 7.50 | 17.48 | 93.22 |
| NAG10501 | Kyoto Univ. | Kansai | 347,614,960 | 345,942,068 | 52,142,244,000 | 51,990,167,960 | 99.52 | 7.86 | 17.33 | 91.22 |
| NAG10504 | Kyoto Univ. | Kansai | 334,274,787 | 332,712,927 | 50,141,218,050 | 49,965,271,108 | 99.53 | 4.19 | 16.66 | 92.67 |

| | | | | | | | | | | |
|---|---|---|---|---|---|---|---|---|---|---|
| NAG10524 | Kyoto Univ. | Kansai | 361,365,486 | 358,522,417 | 54,204,822,900 | 53,835,278,721 | 99.21 | 5.08 | 17.95 | 94.46 |
| NAG10529 | Kyoto Univ. | Kansai | 372,532,780 | 371,139,776 | 55,879,917,000 | 55,732,087,056 | 99.63 | 6.93 | 18.58 | 92.28 |
| NAG10530 | Kyoto Univ. | Kansai | 381,148,863 | 379,561,069 | 57,172,329,450 | 57,011,188,806 | 99.58 | 7.71 | 19.00 | 92.58 |
| NAG10534 | Kyoto Univ. | Kansai | 427,040,927 | 424,466,626 | 64,056,139,050 | 63,726,202,234 | 99.40 | 4.95 | 21.24 | 97.95 |
| NAG10536 | Kyoto Univ. | Kansai | 401,894,188 | 400,279,584 | 60,284,128,200 | 60,118,365,195 | 99.60 | 8.39 | 20.04 | 93.87 |
| NAG10544 | Kyoto Univ. | Kansai | 398,116,433 | 396,068,934 | 59,717,464,950 | 59,381,808,936 | 99.49 | 5.55 | 19.79 | 92.88 |
| NAG10569 | Kyoto Univ. | Kansai | 383,652,286 | 381,306,856 | 57,547,842,900 | 57,236,566,541 | 99.39 | 5.46 | 19.08 | 95.94 |
| NAG10571 | Kyoto Univ. | Kansai | 422,112,141 | 420,309,816 | 63,316,821,150 | 63,147,928,772 | 99.57 | 4.66 | 21.05 | 98.16 |
| NAG10572 | Kyoto Univ. | Kansai | 353,095,062 | 351,337,428 | 52,964,259,300 | 52,795,582,918 | 99.50 | 8.09 | 17.60 | 89.85 |
| NAG10579 | Kyoto Univ. | Kansai | 424,882,479 | 422,701,430 | 63,732,371,850 | 63,489,348,320 | 99.49 | 5.11 | 21.16 | 95.51 |
| NAG10588 | Kyoto Univ. | Kansai | 377,053,825 | 373,720,914 | 56,558,073,750 | 56,071,185,338 | 99.12 | 3.82 | 18.69 | 95.89 |
| NAG10589 | Kyoto Univ. | Kansai | 334,123,641 | 332,074,454 | 50,118,546,150 | 49,868,055,728 | 99.39 | 4.83 | 16.62 | 87.49 |
| NAG10595 | Kyoto Univ. | Kansai | 353,540,203 | 350,629,689 | 53,031,030,450 | 52,675,360,616 | 99.18 | 4.60 | 17.56 | 94.51 |
| NAG10598 | Kyoto Univ. | Kansai | 350,544,445 | 347,587,462 | 52,581,666,750 | 52,218,453,226 | 99.16 | 4.47 | 17.41 | 94.21 |
| NAG10602 | Kyoto Univ. | Kansai | 344,058,361 | 340,523,122 | 51,608,754,150 | 51,128,140,757 | 98.97 | 2.98 | 17.04 | 89.21 |
| NAG10615 | Kyoto Univ. | Kansai | 370,444,621 | 366,375,979 | 55,566,693,150 | 55,082,773,996 | 98.90 | 4.25 | 18.36 | 95.46 |
| NAG10623 | Kyoto Univ. | Kansai | 386,212,419 | 383,678,282 | 57,931,862,850 | 57,615,820,538 | 99.34 | 4.24 | 19.21 | 96.97 |
| NAG10632 | Kyoto Univ. | Kansai | 310,004,030 | 308,427,763 | 46,500,604,500 | 46,278,271,573 | 99.49 | 3.75 | 15.43 | 87.27 |
| NAG10650 | Kyoto Univ. | Kansai | 362,661,933 | 360,413,764 | 54,399,289,950 | 54,164,883,666 | 99.38 | 4.46 | 18.05 | 95.64 |
| NAG10656 | Kyoto Univ. | Kansai | 377,839,609 | 371,590,000 | 56,675,941,350 | 55,793,478,012 | 98.35 | 4.41 | 18.60 | 93.71 |
| NAG10660 | Kyoto Univ. | Kansai | 338,722,193 | 337,115,412 | 50,808,328,950 | 50,635,771,488 | 99.53 | 4.37 | 16.88 | 90.01 |
| NAG10670 | Kyoto Univ. | Kansai | 371,817,154 | 369,150,708 | 55,772,573,100 | 55,429,267,585 | 99.28 | 3.50 | 18.48 | 95.89 |
| NAG10685 | Kyoto Univ. | Kansai | 379,516,795 | 377,733,402 | 56,927,519,250 | 56,766,427,831 | 99.53 | 5.49 | 18.92 | 96.30 |
| NAG10696 | Kyoto Univ. | Kansai | 381,732,374 | 379,694,838 | 57,259,856,100 | 56,973,093,294 | 99.47 | 4.26 | 18.99 | 96.26 |
| NAG10699 | Kyoto Univ. | Kansai | 334,614,472 | 332,577,220 | 50,192,170,800 | 49,893,342,416 | 99.39 | 4.60 | 16.63 | 86.95 |
| NAG10710 | Kyoto Univ. | Kansai | 366,120,251 | 364,204,024 | 54,918,037,650 | 54,699,745,322 | 99.48 | 4.42 | 18.23 | 95.40 |
| NAG10722 | Kyoto Univ. | Kansai | 326,567,910 | 324,576,992 | 48,985,186,500 | 48,722,815,277 | 99.39 | 3.97 | 16.24 | 87.32 |
| NAG10728 | Kyoto Univ. | Kansai | 345,106,432 | 342,237,228 | 51,765,964,800 | 51,430,606,413 | 99.17 | 6.57 | 17.14 | 91.40 |
| NAG10734 | Kyoto Univ. | Kansai | 340,955,654 | 337,685,427 | 51,143,348,100 | 50,654,139,812 | 99.04 | 5.06 | 16.88 | 85.20 |
| NAG10741 | Kyoto Univ. | Kansai | 402,830,820 | 400,330,128 | 60,424,623,000 | 60,071,807,739 | 99.38 | 5.38 | 20.02 | 94.06 |
| NAG10745 | Kyoto Univ. | Kansai | 350,136,005 | 348,025,588 | 52,520,400,750 | 52,292,225,241 | 99.40 | 4.08 | 17.43 | 93.66 |
| NAG10747 | Kyoto Univ. | Kansai | 363,129,543 | 360,422,698 | 54,469,431,450 | 54,127,593,493 | 99.25 | 3.77 | 18.04 | 95.66 |
| NAG10758 | Kyoto Univ. | Kansai | 359,892,439 | 357,764,619 | 53,983,865,850 | 53,732,707,223 | 99.41 | 4.57 | 17.91 | 95.12 |
| NAG10774 | Kyoto Univ. | Kansai | 395,517,551 | 393,088,310 | 59,327,632,650 | 59,001,237,322 | 99.39 | 4.56 | 19.67 | 96.83 |
| NAG10775 | Kyoto Univ. | Kansai | 391,929,324 | 389,705,494 | 58,789,398,600 | 58,466,062,355 | 99.43 | 5.59 | 19.49 | 93.40 |
| NAG10790 | Kyoto Univ. | Kansai | 377,068,605 | 373,312,019 | 56,560,290,750 | 56,067,411,700 | 99.00 | 4.35 | 18.69 | 96.12 |
| NAG10810 | Kyoto Univ. | Kansai | 333,003,623 | 331,123,272 | 49,950,543,450 | 49,757,002,255 | 99.44 | 5.06 | 16.59 | 88.12 |
| NAG10811 | Kyoto Univ. | Kansai | 313,121,059 | 311,373,714 | 46,968,158,850 | 46,794,472,747 | 99.44 | 4.60 | 15.60 | 85.62 |
| NAG10817 | Kyoto Univ. | Kansai | 361,445,861 | 359,432,957 | 54,216,879,150 | 54,014,797,834 | 99.44 | 4.99 | 18.00 | 91.65 |
| NAG10833 | Kyoto Univ. | Kansai | 409,039,814 | 406,770,789 | 61,355,972,100 | 61,087,593,572 | 99.45 | 4.01 | 20.36 | 98.03 |
| NAG10835 | Kyoto Univ. | Kansai | 349,414,346 | 347,466,813 | 52,412,151,900 | 52,201,144,885 | 99.44 | 4.67 | 17.40 | 93.97 |
| NAG10846 | Kyoto Univ. | Kansai | 356,223,062 | 354,184,293 | 53,433,459,300 | 53,233,924,040 | 99.43 | 7.41 | 17.74 | 89.54 |
| NAG10854 | Kyoto Univ. | Kansai | 365,927,420 | 364,244,522 | 54,889,113,000 | 54,743,610,163 | 99.54 | 5.57 | 18.25 | 92.18 |
| NAG10855 | Kyoto Univ. | Kansai | 327,437,595 | 325,544,900 | 49,115,639,250 | 48,918,122,714 | 99.42 | 6.95 | 16.31 | 88.56 |
| NAG10860 | Kyoto Univ. | Kansai | 360,275,638 | 358,102,993 | 54,041,345,700 | 53,783,841,243 | 99.40 | 5.10 | 17.93 | 91.05 |
| NAG10866 | Kyoto Univ. | Kansai | 338,867,750 | 337,209,490 | 50,830,162,500 | 50,671,000,061 | 99.51 | 5.45 | 16.89 | 89.31 |
| NAG10877 | Kyoto Univ. | Kansai | 353,426,320 | 351,710,157 | 53,013,948,000 | 52,819,608,250 | 99.51 | 5.35 | 17.61 | 90.54 |
| NAG10885 | Kyoto Univ. | Kansai | 366,810,658 | 365,160,776 | 55,021,598,700 | 54,807,129,494 | 99.55 | 6.09 | 18.27 | 94.37 |
| NAG10889 | Kyoto Univ. | Kansai | 344,982,310 | 343,065,557 | 51,747,346,500 | 51,600,080,256 | 99.44 | 6.47 | 17.20 | 89.77 |
| NAG10897 | Kyoto Univ. | Kansai | 359,368,009 | 357,018,067 | 53,905,201,350 | 53,591,196,272 | 99.35 | 5.15 | 17.86 | 91.34 |
| NAG10903 | Kyoto Univ. | Kansai | 378,336,468 | 374,907,341 | 56,750,470,200 | 56,298,385,432 | 99.09 | 3.05 | 18.77 | 93.29 |
| NAG10908 | Kyoto Univ. | Kansai | 356,600,717 | 355,235,325 | 53,490,107,550 | 53,360,007,052 | 99.62 | 6.06 | 17.79 | 94.03 |
| NAG10915 | Kyoto Univ. | Kansai | 361,373,937 | 358,988,087 | 54,206,090,550 | 53,947,204,750 | 99.34 | 5.03 | 17.98 | 95.08 |
| NAG10918 | Kyoto Univ. | Kansai | 353,078,890 | 350,348,583 | 52,961,833,500 | 52,678,344,400 | 99.23 | 4.53 | 17.56 | 94.82 |
| NAG10920 | Kyoto Univ. | Kansai | 370,384,386 | 368,568,755 | 55,557,657,900 | 55,380,089,378 | 99.51 | 5.70 | 18.46 | 95.52 |
| NAG10927 | Kyoto Univ. | Kansai | 407,163,923 | 402,039,770 | 61,074,588,450 | 60,376,072,946 | 98.74 | 4.89 | 20.13 | 92.08 |
| NAG10928 | Kyoto Univ. | Kansai | 378,852,672 | 375,437,385 | 56,827,900,800 | 56,313,653,231 | 99.10 | 5.93 | 18.77 | 92.71 |
| NAG10943 | Kyoto Univ. | Kansai | 380,134,518 | 377,896,879 | 57,020,177,700 | 56,804,658,118 | 99.41 | 5.09 | 18.93 | 96.69 |
| NAG10965 | Kyoto Univ. | Kansai | 377,824,631 | 374,646,083 | 56,673,694,650 | 56,285,859,078 | 99.16 | 6.79 | 18.76 | 94.44 |
| NAG10968 | Kyoto Univ. | Kansai | 379,800,944 | 376,548,212 | 56,970,141,600 | 56,553,123,230 | 99.14 | 4.73 | 18.85 | 96.04 |
| NAG10975 | Kyoto Univ. | Kansai | 332,723,105 | 330,832,027 | 49,908,465,750 | 49,668,587,571 | 99.43 | 4.73 | 16.56 | 90.52 |
| NAG10977 | Kyoto Univ. | Kansai | 343,742,214 | 339,436,735 | 51,561,332,100 | 51,000,999,638 | 98.75 | 3.87 | 17.00 | 93.15 |
| NAG10997 | Kyoto Univ. | Kansai | 370,568,653 | 368,977,047 | 55,585,297,950 | 55,456,566,636 | 99.57 | 5.81 | 18.49 | 95.52 |
| NAG11002 | Kyoto Univ. | Kansai | 339,380,648 | 337,920,936 | 50,907,097,200 | 50,768,478,046 | 99.57 | 5.47 | 16.92 | 92.15 |
| NAG11005 | Kyoto Univ. | Kansai | 345,272,143 | 343,178,098 | 51,790,821,450 | 51,492,235,603 | 99.39 | 6.10 | 17.16 | 91.23 |
| NAG11009 | Kyoto Univ. | Kansai | 385,720,418 | 383,989,585 | 57,858,062,700 | 57,689,902,748 | 99.55 | 4.82 | 19.23 | 93.87 |
| NAG11015 | Kyoto Univ. | Kansai | 338,523,347 | 336,018,201 | 50,778,502,050 | 50,434,285,650 | 99.26 | 4.39 | 16.81 | 91.97 |
| NAG11026 | Kyoto Univ. | Kansai | 462,015,517 | 460,121,933 | 69,302,327,550 | 68,956,134,814 | 99.59 | 6.73 | 22.99 | 95.47 |
| NAG11029 | Kyoto Univ. | Kansai | 383,023,401 | 381,351,651 | 57,453,510,150 | 57,301,237,399 | 99.56 | 4.38 | 19.10 | 96.87 |
| NAG11035 | Kyoto Univ. | Kansai | 389,669,698 | 387,335,791 | 58,450,454,700 | 58,178,073,593 | 99.40 | 5.29 | 19.39 | 96.77 |
| NAG11065 | Kyoto Univ. | Kansai | 437,728,983 | 435,994,474 | 65,659,347,450 | 65,311,735,954 | 99.60 | 5.55 | 21.77 | 94.86 |
| NAG11072 | Kyoto Univ. | Kansai | 394,412,987 | 392,965,681 | 59,161,948,050 | 59,033,884,173 | 99.63 | 5.56 | 19.68 | 97.08 |
| NAG11075 | Kyoto Univ. | Kansai | 348,402,435 | 346,341,832 | 52,260,365,250 | 51,976,095,820 | 99.41 | 3.25 | 17.33 | 90.56 |
| NAG11077 | Kyoto Univ. | Kansai | 337,535,561 | 335,140,935 | 50,630,334,150 | 50,340,191,941 | 99.29 | 4.14 | 16.78 | 92.80 |
| NAG11096 | Kyoto Univ. | Kansai | 376,827,910 | 374,468,184 | 56,524,186,500 | 56,274,985,815 | 99.37 | 4.86 | 18.76 | 96.41 |
| NAG11103 | Kyoto Univ. | Kansai | 383,483,606 | 380,929,866 | 57,522,540,900 | 57,234,233,847 | 99.33 | 4.49 | 19.08 | 96.85 |
| NAG11116 | Kyoto Univ. | Kansai | 327,354,787 | 325,266,167 | 49,103,218,050 | 48,802,276,864 | 99.36 | 5.27 | 16.27 | 85.81 |
| NAG11125 | Kyoto Univ. | Kansai | 369,183,339 | 365,849,944 | 55,377,500,850 | 54,979,220,213 | 99.10 | 4.74 | 18.33 | 95.65 |
| NAG11131 | Kyoto Univ. | Kansai | 349,365,845 | 347,622,472 | 52,404,876,750 | 52,190,977,018 | 99.50 | 5.96 | 17.40 | 92.49 |
| NAG11137 | Kyoto Univ. | Kansai | 343,832,595 | 342,326,948 | 51,574,889,250 | 51,414,142,439 | 99.56 | 5.20 | 17.14 | 93.16 |
| NAG11148 | Kyoto Univ. | Kansai | 346,628,468 | 344,831,629 | 51,994,270,200 | 51,772,439,436 | 99.48 | 5.65 | 17.26 | 92.31 |
| NAG11153 | Kyoto Univ. | Kansai | 332,517,105 | 329,698,983 | 49,877,565,750 | 49,543,810,914 | 99.15 | 5.28 | 16.51 | 90.91 |
| NAG11158 | Kyoto Univ. | Kansai | 353,604,805 | 349,649,908 | 53,040,720,750 | 52,568,160,922 | 98.88 | 4.11 | 17.52 | 94.56 |
| NAG11177 | Kyoto Univ. | Kansai | 344,391,817 | 342,604,291 | 51,658,772,550 | 51,480,291,125 | 99.48 | 4.48 | 17.16 | 93.72 |
| NAG11182 | Kyoto Univ. | Kansai | 393,006,639 | 391,159,660 | 58,950,995,850 | 58,699,118,018 | 99.53 | 5.24 | 19.57 | 96.53 |
| NAG11192 | Kyoto Univ. | Kansai | 365,848,636 | 363,977,044 | 54,877,295,400 | 54,688,350,106 | 99.49 | 6.39 | 18.23 | 91.31 |
| NAG11201 | Kyoto Univ. | Kansai | 360,956,353 | 359,037,981 | 54,143,452,950 | 53,945,627,416 | 99.47 | 4.80 | 17.98 | 95.32 |
| NAG11202 | Kyoto Univ. | Kansai | 388,655,572 | 386,410,128 | 58,298,335,800 | 58,048,080,430 | 99.42 | 5.01 | 19.35 | 96.62 |
| NAG11204 | Kyoto Univ. | Kansai | 359,935,836 | 358,145,599 | 53,990,375,400 | 53,754,485,623 | 99.50 | 3.77 | 17.92 | 94.94 |
| NAG11207 | Kyoto Univ. | Kansai | 341,505,116 | 339,345,483 | 51,225,767,400 | 50,962,578,709 | 99.37 | 4.22 | 16.99 | 93.16 |
| NAG11231 | Kyoto Univ. | Kansai | 346,995,523 | 345,084,420 | 52,049,328,450 | 51,850,194,425 | 99.45 | 6.05 | 17.28 | 89.41 |
| NAG11236 | Kyoto Univ. | Kansai | 363,992,181 | 362,298,195 | 54,598,827,150 | 54,415,563,409 | 99.53 | 5.80 | 18.14 | 94.54 |
| NAG11243 | Kyoto Univ. | Kansai | 348,490,364 | 345,822,403 | 52,273,554,600 | 51,990,223,256 | 99.23 | 5.01 | 17.33 | 90.72 |
| NAG11248 | Kyoto Univ. | Kansai | 362,772,268 | 360,179,859 | 54,415,840,200 | 53,986,469,949 | 99.29 | 5.88 | 18.00 | 86.36 |
| NAG11262 | Kyoto Univ. | Kansai | 355,026,103 | 352,957,922 | 53,253,915,450 | 53,039,716,498 | 99.42 | 4.61 | 17.68 | 94.87 |
| NAG11263 | Kyoto Univ. | Kansai | 365,867,614 | 363,291,656 | 54,880,142,100 | 54,561,332,502 | 99.30 | 5.41 | 18.19 | 95.27 |
| NAG11266 | Kyoto Univ. | Kansai | 357,762,485 | 355,601,948 | 53,664,372,750 | 53,406,125,833 | 99.40 | 4.47 | 17.80 | 95.00 |
| NAG11267 | Kyoto Univ. | Kansai | 422,347,718 | 419,987,403 | 63,352,157,700 | 63,068,102,060 | 99.44 | 5.59 | 21.02 | 95.37 |
| NAG11277 | Kyoto Univ. | Kansai | 440,787,986 | 438,341,715 | 66,118,197,900 | 65,891,173,955 | 99.45 | 6.37 | 21.96 | 95.86 |
| NAG11283 | Kyoto Univ. | Kansai | 356,392,704 | 354,455,087 | 53,458,905,600 | 53,264,284,221 | 99.46 | 4.07 | 17.75 | 95.15 |

| | | | | | | | | | | |
|---|---|---|---|---|---|---|---|---|---|---|
| NAG11296 | Kyoto Univ. | Kansai | 372,981,252 | 371,176,593 | 55,947,187,800 | 55,670,657,205 | 99.52 | 6.49 | 18.56 | 94.09 |
| NAG11299 | Kyoto Univ. | Kansai | 360,900,876 | 358,897,004 | 54,135,131,400 | 53,900,819,765 | 99.44 | 4.79 | 17.97 | 94.79 |
| NAG11310 | Kyoto Univ. | Kansai | 355,988,460 | 352,795,050 | 53,398,269,000 | 52,996,784,639 | 99.10 | 3.98 | 17.67 | 91.63 |
| NAG11315 | Kyoto Univ. | Kansai | 390,884,885 | 387,823,292 | 58,632,732,750 | 58,207,549,661 | 99.22 | 6.27 | 19.40 | 94.53 |
| NAG11318 | Kyoto Univ. | Kansai | 315,610,620 | 313,661,497 | 47,341,593,000 | 47,095,102,835 | 99.38 | 4.59 | 15.70 | 88.97 |
| NAG11319 | Kyoto Univ. | Kansai | 406,669,047 | 404,352,021 | 61,000,357,050 | 60,765,789,592 | 99.43 | 7.98 | 20.26 | 96.69 |
| NAG11322 | Kyoto Univ. | Kansai | 325,270,035 | 323,543,316 | 48,790,505,250 | 48,613,767,483 | 99.47 | 3.92 | 16.20 | 91.67 |
| NAG11326 | Kyoto Univ. | Kansai | 370,042,893 | 367,003,367 | 55,506,433,950 | 55,072,781,248 | 99.18 | 6.21 | 18.36 | 90.25 |
| NAG11332 | Kyoto Univ. | Kansai | 339,777,643 | 337,985,156 | 50,966,646,450 | 50,784,390,847 | 99.47 | 5.14 | 16.93 | 92.12 |
| NAG11336 | Kyoto Univ. | Kansai | 340,979,516 | 338,639,603 | 51,146,927,400 | 50,853,699,415 | 99.31 | 4.76 | 16.95 | 92.40 |
| NAG11339 | Kyoto Univ. | Kansai | 340,026,058 | 338,307,785 | 51,003,908,700 | 50,794,614,887 | 99.49 | 5.88 | 16.93 | 91.30 |
| NAG11341 | Kyoto Univ. | Kansai | 348,349,379 | 346,182,875 | 52,252,406,850 | 51,978,522,369 | 99.38 | 4.03 | 17.33 | 94.10 |
| NAG11347 | Kyoto Univ. | Kansai | 353,238,100 | 351,366,173 | 52,985,715,000 | 52,745,201,914 | 99.47 | 5.25 | 17.58 | 91.16 |
| NAG11357 | Kyoto Univ. | Kansai | 390,727,694 | 388,269,654 | 58,609,154,100 | 58,313,105,878 | 99.37 | 5.34 | 19.44 | 96.90 |
| NAG11358 | Kyoto Univ. | Kansai | 313,611,285 | 311,300,769 | 47,041,692,750 | 46,743,623,969 | 99.26 | 4.45 | 15.58 | 87.31 |
| NAG11365 | Kyoto Univ. | Kansai | 325,905,733 | 322,595,370 | 48,885,859,950 | 48,458,963,086 | 98.98 | 4.90 | 16.15 | 88.90 |
| NAG11373 | Kyoto Univ. | Kansai | 385,802,121 | 383,719,819 | 57,870,318,150 | 57,584,718,040 | 99.46 | 6.20 | 19.19 | 95.42 |
| NAG11377 | Kyoto Univ. | Kansai | 314,175,560 | 312,803,969 | 47,126,334,000 | 46,996,846,076 | 99.56 | 4.71 | 15.67 | 89.28 |
| NAG11392 | Kyoto Univ. | Kansai | 365,910,958 | 364,057,916 | 54,886,643,700 | 54,682,806,937 | 99.49 | 5.51 | 18.23 | 94.74 |
| NAG11415 | Kyoto Univ. | Kansai | 363,403,993 | 361,681,358 | 54,510,598,950 | 54,316,866,689 | 99.53 | 5.89 | 18.11 | 94.47 |
| NAG11416 | Kyoto Univ. | Kansai | 333,641,506 | 332,281,677 | 50,046,225,900 | 49,941,116,711 | 99.59 | 4.56 | 16.65 | 92.40 |
| NAG11443 | Kyoto Univ. | Kansai | 389,071,675 | 386,738,861 | 58,360,751,250 | 58,051,919,967 | 99.40 | 7.80 | 19.35 | 94.90 |
| NAG11456 | Kyoto Univ. | Kansai | 332,575,974 | 331,419,074 | 49,886,396,100 | 49,779,586,687 | 99.65 | 4.42 | 16.59 | 91.87 |
| NAG11458 | Kyoto Univ. | Kansai | 334,247,114 | 330,641,509 | 50,137,067,100 | 49,635,553,026 | 98.92 | 5.07 | 16.55 | 88.24 |
| NAG11475 | Kyoto Univ. | Kansai | 347,170,107 | 344,276,775 | 52,075,516,050 | 51,691,975,706 | 99.17 | 5.45 | 17.23 | 90.55 |
| NAG11476 | Kyoto Univ. | Kansai | 365,010,798 | 362,566,682 | 54,751,619,700 | 54,493,475,342 | 99.33 | 4.89 | 18.16 | 95.66 |
| NAG11483 | Kyoto Univ. | Kansai | 323,417,663 | 321,083,089 | 48,512,649,450 | 48,255,903,586 | 99.28 | 4.69 | 16.09 | 89.10 |
| NAG11493 | Kyoto Univ. | Kansai | 365,678,591 | 362,733,497 | 54,851,788,650 | 54,537,942,379 | 99.19 | 4.23 | 18.18 | 91.68 |
| NAG11505 | Kyoto Univ. | Kansai | 447,074,393 | 444,319,092 | 67,061,158,950 | 66,622,078,376 | 99.38 | 6.69 | 22.21 | 94.19 |
| NAG11524 | Kyoto Univ. | Kansai | 360,249,404 | 358,816,533 | 54,037,410,600 | 53,903,767,272 | 99.60 | 5.38 | 17.97 | 91.34 |
| NAG11528 | Kyoto Univ. | Kansai | 384,378,653 | 382,587,913 | 57,656,797,950 | 57,500,347,442 | 99.53 | 7.07 | 19.17 | 96.26 |
| NAG11534 | Kyoto Univ. | Kansai | 322,226,202 | 319,905,644 | 48,333,930,300 | 48,050,802,895 | 99.28 | 3.83 | 16.02 | 86.91 |
| NAG11537 | Kyoto Univ. | Kansai | 396,899,860 | 395,628,305 | 59,534,979,000 | 59,384,862,795 | 99.68 | 5.93 | 19.79 | 96.77 |
| NAG11538 | Kyoto Univ. | Kansai | 360,342,115 | 358,324,027 | 54,051,317,250 | 53,846,639,789 | 99.44 | 5.00 | 17.95 | 95.21 |
| NAG11544 | Kyoto Univ. | Kansai | 345,796,573 | 344,623,460 | 51,869,485,950 | 51,774,002,039 | 99.66 | 4.92 | 17.26 | 93.47 |
| NAG11551 | Kyoto Univ. | Kansai | 450,674,120 | 447,919,988 | 67,601,118,000 | 67,205,520,489 | 99.39 | 4.93 | 22.40 | 98.51 |
| NAG11556 | Kyoto Univ. | Kansai | 339,183,440 | 336,975,053 | 50,877,516,000 | 50,589,132,179 | 99.35 | 5.08 | 16.86 | 89.33 |
| NAG11564 | Kyoto Univ. | Kansai | 352,792,828 | 351,649,895 | 52,918,924,200 | 52,815,785,425 | 99.68 | 5.53 | 17.61 | 94.04 |
| NAG11569 | Kyoto Univ. | Kansai | 387,289,295 | 384,814,775 | 58,093,394,250 | 57,768,468,914 | 99.36 | 4.90 | 19.26 | 96.44 |
| NAG11576 | Kyoto Univ. | Kansai | 369,378,206 | 367,622,372 | 55,406,730,900 | 55,242,469,670 | 99.52 | 4.64 | 18.41 | 96.06 |
| NAG11577 | Kyoto Univ. | Kansai | 358,231,801 | 355,793,919 | 53,734,770,150 | 53,471,761,731 | 99.32 | 4.59 | 17.82 | 90.78 |
| NAG11579 | Kyoto Univ. | Kansai | 360,850,140 | 359,584,148 | 54,127,521,000 | 54,016,888,764 | 99.65 | 5.52 | 18.01 | 94.84 |
| NAG11582 | Kyoto Univ. | Kansai | 347,720,349 | 346,513,644 | 52,158,052,350 | 52,021,134,703 | 99.65 | 4.56 | 17.34 | 93.38 |
| NAG11589 | Kyoto Univ. | Kansai | 337,000,164 | 335,901,202 | 50,550,024,600 | 50,464,552,209 | 99.67 | 5.25 | 16.82 | 92.40 |
| NAG11603 | Kyoto Univ. | Kansai | 355,803,350 | 354,368,046 | 53,370,502,500 | 53,251,019,427 | 99.60 | 5.08 | 17.75 | 95.05 |
| NAG11605 | Kyoto Univ. | Kansai | 308,772,738 | 306,955,932 | 46,315,910,700 | 46,092,721,422 | 99.41 | 4.23 | 15.36 | 88.15 |
| NAG11615 | Kyoto Univ. | Kansai | 384,057,996 | 381,359,798 | 57,608,699,400 | 57,302,557,270 | 99.30 | 7.49 | 19.10 | 95.17 |
| NAG11623 | Kyoto Univ. | Kansai | 355,143,864 | 353,755,841 | 53,271,579,600 | 53,205,374,578 | 99.61 | 5.77 | 17.74 | 94.44 |
| NAG11632 | Kyoto Univ. | Kansai | 441,024,935 | 439,507,459 | 66,153,740,250 | 65,987,178,919 | 99.66 | 7.72 | 22.00 | 95.55 |
| NAG11645 | Kyoto Univ. | Kansai | 469,430,754 | 466,250,306 | 70,414,613,100 | 70,002,635,526 | 99.32 | 7.53 | 23.33 | 98.16 |
| NAG11647 | Kyoto Univ. | Kansai | 332,405,734 | 329,582,261 | 49,860,860,100 | 49,533,189,821 | 99.15 | 4.69 | 16.51 | 87.82 |
| NAG11651 | Kyoto Univ. | Kansai | 411,803,980 | 409,365,447 | 61,770,597,000 | 61,469,859,873 | 99.41 | 4.43 | 20.49 | 97.84 |
| NAG11652 | Kyoto Univ. | Kansai | 400,694,097 | 399,315,081 | 60,104,114,550 | 60,005,703,802 | 99.66 | 5.19 | 20.00 | 97.49 |
| NAG11657 | Kyoto Univ. | Kansai | 351,305,995 | 349,401,891 | 52,695,899,250 | 52,491,448,825 | 99.46 | 4.87 | 17.50 | 94.35 |
| NAG11661 | Kyoto Univ. | Kansai | 340,231,224 | 338,334,537 | 51,034,683,600 | 50,810,283,425 | 99.44 | 4.18 | 16.94 | 93.21 |
| NAG11665 | Kyoto Univ. | Kansai | 344,618,936 | 342,540,714 | 51,692,840,400 | 51,417,553,649 | 99.40 | 4.92 | 17.14 | 92.25 |
| NAG11669 | Kyoto Univ. | Kansai | 316,185,696 | 314,177,673 | 47,427,854,400 | 47,185,230,557 | 99.36 | 4.73 | 15.73 | 88.75 |
| NAG11671 | Kyoto Univ. | Kansai | 380,892,036 | 379,400,001 | 57,133,805,400 | 56,954,139,329 | 99.61 | 6.35 | 18.98 | 95.32 |
| NAG11673 | Kyoto Univ. | Kansai | 411,921,990 | 410,512,662 | 61,788,298,500 | 61,733,615,260 | 99.66 | 6.74 | 20.58 | 97.75 |
| NAG11674 | Kyoto Univ. | Kansai | 338,764,646 | 336,472,409 | 50,814,696,900 | 50,507,115,372 | 99.32 | 4.81 | 16.84 | 91.64 |
| NAG11677 | Kyoto Univ. | Kansai | 338,911,770 | 336,696,589 | 50,836,765,500 | 50,532,602,041 | 99.35 | 5.34 | 16.84 | 91.84 |
| NAG11679 | Kyoto Univ. | Kansai | 311,179,518 | 309,943,333 | 46,676,927,700 | 46,564,055,785 | 99.60 | 4.32 | 15.52 | 89.13 |
| NAG11682 | Kyoto Univ. | Kansai | 340,340,627 | 337,803,253 | 51,051,094,050 | 50,781,080,364 | 99.25 | 5.31 | 16.93 | 88.87 |
| NAG11694 | Kyoto Univ. | Kansai | 345,050,384 | 342,039,106 | 51,757,557,600 | 51,390,208,998 | 99.13 | 6.42 | 17.13 | 91.01 |
| NAG11707 | Kyoto Univ. | Kansai | 351,774,744 | 349,819,927 | 52,766,211,600 | 52,536,736,732 | 99.44 | 5.61 | 17.51 | 93.92 |
| NAG11725 | Kyoto Univ. | Kansai | 354,893,411 | 353,456,139 | 53,234,011,650 | 53,124,400,541 | 99.60 | 4.44 | 17.71 | 95.15 |
| NAG11726 | Kyoto Univ. | Kansai | 382,978,469 | 381,626,699 | 57,446,770,350 | 57,310,544,949 | 99.65 | 5.14 | 19.10 | 93.49 |
| NAG11733 | Kyoto Univ. | Kansai | 397,137,884 | 393,253,111 | 59,570,682,600 | 59,071,835,589 | 99.02 | 6.91 | 19.69 | 95.92 |
| NAG11734 | Kyoto Univ. | Kansai | 342,158,009 | 340,137,635 | 51,323,701,350 | 51,065,193,422 | 99.41 | 5.27 | 17.02 | 92.16 |
| NAG11738 | Kyoto Univ. | Kansai | 368,757,574 | 366,968,259 | 55,313,636,100 | 55,170,362,862 | 99.51 | 4.47 | 18.39 | 92.98 |
| NAG11744 | Kyoto Univ. | Kansai | 326,984,309 | 325,562,342 | 49,047,646,350 | 48,905,323,120 | 99.57 | 4.53 | 16.30 | 91.58 |
| NAG11746 | Kyoto Univ. | Kansai | 381,756,008 | 379,637,389 | 57,263,401,200 | 56,845,943,933 | 99.45 | 6.34 | 18.95 | 90.71 |
| NAG11777 | Kyoto Univ. | Kansai | 354,979,477 | 352,691,682 | 53,246,921,550 | 52,952,254,948 | 99.36 | 4.67 | 17.65 | 93.78 |
| NAG11780 | Kyoto Univ. | Kansai | 376,148,457 | 374,390,123 | 56,422,268,550 | 56,234,640,004 | 99.53 | 4.61 | 18.74 | 96.43 |
| NAG11786 | Kyoto Univ. | Kansai | 383,682,078 | 380,681,287 | 57,552,311,700 | 57,162,483,399 | 99.22 | 5.04 | 19.05 | 92.42 |
| NAG11787 | Kyoto Univ. | Kansai | 328,596,778 | 325,732,730 | 49,289,516,700 | 48,917,902,494 | 99.13 | 5.55 | 16.31 | 86.73 |
| NAG11801 | Kyoto Univ. | Kansai | 372,215,108 | 370,819,709 | 55,832,266,200 | 55,693,981,742 | 99.63 | 5.56 | 18.56 | 92.32 |
| NAG11806 | Kyoto Univ. | Kansai | 359,472,236 | 357,480,807 | 53,920,835,400 | 53,683,149,061 | 99.45 | 4.77 | 17.89 | 95.10 |
| NAG11809 | Kyoto Univ. | Kansai | 353,440,638 | 350,470,742 | 53,016,095,700 | 52,643,278,003 | 99.16 | 4.74 | 17.55 | 92.55 |
| NAG11811 | Kyoto Univ. | Kansai | 338,578,122 | 337,326,106 | 50,786,718,300 | 50,721,576,694 | 99.63 | 5.07 | 16.91 | 93.28 |
| NAG11830 | Kyoto Univ. | Kansai | 379,786,044 | 377,980,107 | 56,967,906,600 | 56,797,607,292 | 99.52 | 5.68 | 18.93 | 96.44 |
| NAG11848 | Kyoto Univ. | Kansai | 352,734,430 | 348,022,945 | 52,910,164,500 | 52,234,844,926 | 98.66 | 3.86 | 17.41 | 93.91 |
| NAG11851 | Kyoto Univ. | Kansai | 325,605,108 | 323,794,428 | 48,840,766,200 | 48,612,590,180 | 99.44 | 5.39 | 16.20 | 90.26 |
| NAG11857 | Kyoto Univ. | Kansai | 321,530,663 | 319,924,767 | 48,229,599,450 | 48,062,584,116 | 99.50 | 4.67 | 16.02 | 90.22 |
| NAG11858 | Kyoto Univ. | Kansai | 361,014,240 | 359,565,663 | 54,152,136,000 | 54,019,462,359 | 99.60 | 5.41 | 18.01 | 95.29 |
| NAG11869 | Kyoto Univ. | Kansai | 344,962,557 | 343,456,749 | 51,744,383,550 | 51,605,934,695 | 99.56 | 4.75 | 17.20 | 94.01 |
| NAG11873 | Kyoto Univ. | Kansai | 391,478,524 | 389,749,658 | 58,721,778,600 | 58,527,414,094 | 99.56 | 4.68 | 19.51 | 96.93 |
| NAG11884 | Kyoto Univ. | Kansai | 329,919,694 | 328,082,735 | 49,487,954,100 | 49,236,513,043 | 99.44 | 4.39 | 16.41 | 90.90 |
| NAG11892 | Kyoto Univ. | Kansai | 385,004,777 | 382,248,981 | 57,750,716,550 | 57,445,755,937 | 99.28 | 4.39 | 19.15 | 96.96 |
| NAG11899 | Kyoto Univ. | Kansai | 357,347,674 | 354,965,300 | 53,602,151,100 | 53,344,021,431 | 99.33 | 4.98 | 17.78 | 88.48 |
| NAG11908 | Kyoto Univ. | Kansai | 356,652,346 | 354,630,598 | 53,497,851,900 | 53,210,698,628 | 99.43 | 4.51 | 17.74 | 90.78 |
| NAG11921 | Kyoto Univ. | Kansai | 396,980,005 | 394,688,869 | 59,547,000,750 | 59,263,136,167 | 99.42 | 5.31 | 19.75 | 88.10 |
| NAG11935 | Kyoto Univ. | Kansai | 328,421,865 | 327,143,236 | 49,263,279,750 | 49,127,787,469 | 99.61 | 4.49 | 16.38 | 90.72 |
| NAG11936 | Kyoto Univ. | Kansai | 394,746,305 | 391,796,777 | 59,211,945,750 | 58,849,806,428 | 99.25 | 6.22 | 19.62 | 92.78 |
| NAG11940 | Kyoto Univ. | Kansai | 418,621,182 | 417,067,931 | 62,793,177,300 | 62,644,987,830 | 99.63 | 6.33 | 20.88 | 97.36 |
| NAG11943 | Kyoto Univ. | Kansai | 329,509,323 | 328,301,746 | 49,426,398,450 | 49,295,094,298 | 99.63 | 5.22 | 16.43 | 90.47 |
| NAG11944 | Kyoto Univ. | Kansai | 356,924,144 | 355,066,017 | 53,538,621,600 | 53,351,439,592 | 99.48 | 4.98 | 17.78 | 94.81 |
| NAG11955 | Kyoto Univ. | Kansai | 359,224,954 | 357,588,438 | 53,883,743,100 | 53,733,628,979 | 99.54 | 8.56 | 17.91 | 93.82 |
| NAG11960 | Kyoto Univ. | Kansai | 354,113,157 | 352,866,593 | 53,116,973,550 | 52,984,500,079 | 99.65 | 5.80 | 17.66 | 93.47 |
| NAG11980 | Kyoto Univ. | Kansai | 358,901,990 | 357,353,671 | 53,835,298,500 | 53,634,946,829 | 99.57 | 4.51 | 17.88 | 94.56 |

| | | | | | | | | | | |
|---|---|---|---|---|---|---|---|---|---|---|
| NAG11985 | Kyoto Univ. | Kansai | 397,126,814 | 395,038,390 | 59,569,022,100 | 59,345,406,651 | 99.47 | 5.56 | 19.78 | 97.35 |
| NAG11988 | Kyoto Univ. | Kansai | 355,223,522 | 351,804,873 | 53,283,528,300 | 52,864,158,592 | 99.04 | 7.38 | 17.62 | 91.78 |
| NAG11995 | Kyoto Univ. | Kansai | 415,188,467 | 412,381,861 | 62,278,270,050 | 61,968,302,858 | 99.32 | 6.06 | 20.66 | 97.90 |
| NAG12013 | Kyoto Univ. | Kansai | 401,425,806 | 399,880,321 | 60,213,870,900 | 60,070,187,998 | 99.62 | 8.82 | 20.02 | 96.80 |
| NAG12014 | Kyoto Univ. | Kansai | 362,042,419 | 360,196,515 | 54,306,362,850 | 54,061,224,008 | 99.49 | 5.00 | 18.02 | 94.82 |
| NAG12028 | Kyoto Univ. | Kansai | 342,028,989 | 339,871,951 | 51,304,348,350 | 51,058,048,497 | 99.37 | 4.81 | 17.02 | 88.55 |
| NAG12030 | Kyoto Univ. | Kansai | 391,290,128 | 389,197,220 | 58,693,519,200 | 58,459,270,178 | 99.47 | 5.48 | 19.49 | 94.21 |
| NAG12033 | Kyoto Univ. | Kansai | 542,282,748 | 539,057,653 | 81,342,412,200 | 80,824,107,124 | 99.41 | 7.31 | 26.94 | 98.84 |
| NAG12038 | Kyoto Univ. | Kansai | 374,326,382 | 371,252,084 | 56,148,957,300 | 55,714,127,817 | 99.18 | 4.92 | 18.57 | 95.32 |
| NAG12051 | Kyoto Univ. | Kansai | 338,158,384 | 336,326,432 | 50,723,757,600 | 50,468,051,420 | 99.46 | 4.82 | 16.82 | 92.18 |
| NAG12061 | Kyoto Univ. | Kansai | 320,887,765 | 319,287,659 | 48,133,164,750 | 47,990,106,506 | 99.50 | 4.71 | 16.00 | 90.71 |
| NAG12092 | Kyoto Univ. | Kansai | 364,704,509 | 362,652,821 | 54,705,676,350 | 54,464,707,948 | 99.44 | 7.22 | 18.15 | 93.36 |
| NAG12093 | Kyoto Univ. | Kansai | 336,402,235 | 334,270,921 | 50,460,335,250 | 50,167,688,927 | 99.37 | 4.42 | 16.72 | 92.24 |
| NAG12106 | Kyoto Univ. | Kansai | 360,732,978 | 354,843,194 | 54,109,946,700 | 53,315,766,459 | 98.37 | 5.85 | 17.77 | 92.64 |
| NAG12128 | Kyoto Univ. | Kansai | 359,884,329 | 358,648,648 | 53,982,649,350 | 53,837,051,466 | 99.66 | 5.15 | 17.95 | 94.06 |
| NAG12130 | Kyoto Univ. | Kansai | 336,079,339 | 334,584,449 | 50,411,900,850 | 50,275,673,548 | 99.56 | 5.16 | 16.76 | 89.47 |
| NAG12131 | Kyoto Univ. | Kansai | 358,802,921 | 357,317,665 | 53,820,438,150 | 53,691,608,101 | 99.59 | 7.44 | 17.90 | 94.13 |
| NAG12137 | Kyoto Univ. | Kansai | 347,545,943 | 346,412,580 | 52,131,891,450 | 52,037,481,608 | 99.67 | 5.22 | 17.35 | 93.38 |
| NAG12141 | Kyoto Univ. | Kansai | 347,295,773 | 345,494,997 | 52,094,365,950 | 51,828,632,881 | 99.48 | 4.97 | 17.28 | 92.33 |
| NAG12142 | Kyoto Univ. | Kansai | 391,855,845 | 388,414,656 | 58,778,376,750 | 58,326,789,574 | 99.12 | 4.79 | 19.44 | 92.91 |
| NAG12147 | Kyoto Univ. | Kansai | 367,910,169 | 366,590,045 | 55,186,525,350 | 55,045,773,488 | 99.64 | 5.46 | 18.35 | 95.05 |
| NAG12157 | Kyoto Univ. | Kansai | 380,634,807 | 377,767,722 | 57,095,221,050 | 56,795,039,452 | 99.25 | 5.54 | 18.93 | 95.76 |
| NAG12159 | Kyoto Univ. | Kansai | 306,236,833 | 304,550,067 | 45,935,524,950 | 45,765,677,578 | 99.45 | 4.97 | 15.26 | 87.41 |
| NAG12160 | Kyoto Univ. | Kansai | 328,785,159 | 326,556,650 | 49,317,773,850 | 49,089,191,135 | 99.32 | 4.98 | 16.36 | 90.74 |
| NAG12161 | Kyoto Univ. | Kansai | 375,818,908 | 373,436,841 | 56,372,836,200 | 56,069,779,985 | 99.37 | 5.83 | 18.69 | 93.98 |
| NAG12163 | Kyoto Univ. | Kansai | 362,895,965 | 360,383,876 | 54,434,394,750 | 54,160,628,718 | 99.31 | 4.77 | 18.05 | 95.54 |
| NAG12165 | Kyoto Univ. | Kansai | 378,129,179 | 376,828,564 | 56,719,376,850 | 56,585,290,507 | 99.66 | 5.67 | 18.86 | 95.83 |
| NAG12172 | Kyoto Univ. | Kansai | 381,202,415 | 379,381,808 | 57,180,362,250 | 56,969,681,438 | 99.52 | 5.16 | 18.99 | 93.60 |
| NAG12179 | Kyoto Univ. | Kansai | 355,262,442 | 353,678,219 | 53,289,366,300 | 53,136,958,711 | 99.55 | 4.72 | 17.71 | 94.94 |
| NAG12195 | Kyoto Univ. | Kansai | 382,723,814 | 381,169,749 | 57,408,572,100 | 57,220,991,912 | 99.59 | 4.91 | 19.07 | 96.39 |
| NAG12196 | Kyoto Univ. | Kansai | 362,498,850 | 360,004,551 | 54,374,827,500 | 54,069,321,842 | 99.31 | 4.57 | 18.02 | 92.27 |
| NAG12198 | Kyoto Univ. | Kansai | 378,826,337 | 377,182,795 | 56,823,950,550 | 56,643,450,577 | 99.57 | 5.18 | 18.88 | 96.17 |
| NAG12200 | Kyoto Univ. | Kansai | 411,440,369 | 410,048,037 | 61,716,055,350 | 61,537,371,536 | 99.66 | 8.34 | 20.51 | 94.16 |
| NAG12217 | Kyoto Univ. | Kansai | 336,491,317 | 334,195,515 | 50,473,697,550 | 50,199,331,383 | 99.32 | 4.84 | 16.73 | 87.14 |
| NAG12222 | Kyoto Univ. | Kansai | 340,190,908 | 337,713,960 | 51,028,636,200 | 50,726,231,886 | 99.27 | 4.88 | 16.91 | 91.98 |
| NAG12224 | Kyoto Univ. | Kansai | 363,083,689 | 361,799,187 | 54,462,553,350 | 54,357,093,639 | 99.65 | 5.20 | 18.12 | 95.13 |
| NAG12226 | Kyoto Univ. | Kansai | 361,010,793 | 359,821,192 | 54,151,618,950 | 53,938,185,133 | 99.67 | 7.76 | 17.98 | 92.19 |
| NAG12228 | Kyoto Univ. | Kansai | 406,446,508 | 404,971,258 | 60,966,976,200 | 60,830,487,853 | 99.64 | 5.16 | 20.28 | 97.62 |
| NAG12236 | Kyoto Univ. | Kansai | 337,409,379 | 336,059,960 | 50,611,406,850 | 50,492,499,538 | 99.60 | 5.20 | 16.83 | 92.59 |
| NAG12237 | Kyoto Univ. | Kansai | 360,911,933 | 359,004,505 | 54,136,789,950 | 53,891,695,421 | 99.47 | 5.24 | 17.96 | 94.82 |
| NAG12239 | Kyoto Univ. | Kansai | 374,469,448 | 372,474,818 | 56,170,417,200 | 55,934,397,462 | 99.47 | 5.69 | 18.64 | 95.82 |
| NAG12244 | Kyoto Univ. | Kansai | 462,409,830 | 459,620,199 | 69,361,474,500 | 69,047,702,499 | 99.40 | 4.27 | 23.02 | 98.93 |
| NAG12245 | Kyoto Univ. | Kansai | 370,341,490 | 367,727,338 | 55,551,223,500 | 55,235,166,853 | 99.29 | 5.59 | 18.41 | 89.56 |
| NAG12248 | Kyoto Univ. | Kansai | 399,073,560 | 396,451,015 | 59,861,034,000 | 59,573,910,171 | 99.34 | 9.07 | 19.86 | 95.71 |
| NAG12259 | Kyoto Univ. | Kansai | 362,358,221 | 360,001,646 | 54,353,733,150 | 54,093,284,805 | 99.35 | 4.95 | 18.03 | 90.05 |
| NAG12260 | Kyoto Univ. | Kansai | 344,213,434 | 341,538,554 | 51,632,015,100 | 51,314,043,591 | 99.22 | 4.83 | 17.10 | 92.97 |
| NAG12273 | Kyoto Univ. | Kansai | 387,962,306 | 386,761,029 | 58,194,345,900 | 58,084,059,789 | 99.69 | 6.01 | 19.36 | 96.64 |
| NAG12281 | Kyoto Univ. | Kansai | 376,218,718 | 375,034,513 | 56,432,807,700 | 56,297,441,097 | 99.69 | 6.08 | 18.77 | 95.59 |
| NAG12283 | Kyoto Univ. | Kansai | 416,392,022 | 414,437,255 | 62,458,803,300 | 62,234,372,941 | 99.53 | 6.53 | 20.74 | 97.32 |
| NAG12299 | Kyoto Univ. | Kansai | 335,454,474 | 333,713,016 | 50,318,171,100 | 50,169,069,434 | 99.48 | 5.36 | 16.72 | 92.58 |
| NAG12307 | Kyoto Univ. | Kansai | 355,289,176 | 352,962,603 | 53,293,376,400 | 53,039,928,328 | 99.35 | 5.40 | 17.68 | 94.15 |
| NAG12313 | Kyoto Univ. | Kansai | 328,040,676 | 325,383,311 | 49,206,101,400 | 48,846,858,485 | 99.19 | 4.88 | 16.28 | 90.64 |
| NAG12317 | Kyoto Univ. | Kansai | 416,084,333 | 414,309,497 | 62,412,649,950 | 62,202,415,749 | 99.57 | 9.49 | 20.73 | 96.87 |
| NAG12319 | Kyoto Univ. | Kansai | 330,133,950 | 328,234,570 | 49,520,092,500 | 49,319,049,140 | 99.42 | 4.85 | 16.44 | 91.35 |
| NAG12330 | Kyoto Univ. | Kansai | 401,512,894 | 399,124,615 | 60,226,934,100 | 59,942,307,719 | 99.41 | 5.09 | 19.98 | 97.28 |
| NAG12331 | Kyoto Univ. | Kansai | 336,044,030 | 334,569,930 | 50,406,604,500 | 50,242,108,660 | 99.56 | 4.61 | 16.75 | 92.44 |
| NAG12334 | Kyoto Univ. | Kansai | 356,496,757 | 353,120,488 | 53,474,513,550 | 53,048,636,312 | 99.05 | 4.45 | 17.68 | 94.31 |
| NAG12336 | Kyoto Univ. | Kansai | 621,647,144 | 618,559,218 | 93,247,071,600 | 92,829,034,605 | 99.50 | 8.51 | 30.94 | 98.73 |
| NAG12337 | Kyoto Univ. | Kansai | 391,268,665 | 390,086,520 | 58,690,299,750 | 58,590,024,269 | 99.70 | 5.86 | 19.53 | 96.84 |
| NAG12338 | Kyoto Univ. | Kansai | 365,737,464 | 363,849,038 | 54,860,619,600 | 54,626,559,720 | 99.48 | 4.97 | 18.21 | 95.23 |
| NAG12339 | Kyoto Univ. | Kansai | 318,562,645 | 315,252,496 | 47,784,396,750 | 47,313,005,491 | 98.96 | 3.89 | 15.77 | 88.81 |
| NAG12340 | Kyoto Univ. | Kansai | 364,175,563 | 362,882,185 | 54,626,334,450 | 54,512,590,996 | 99.64 | 8.15 | 18.17 | 94.17 |
| NAG12342 | Kyoto Univ. | Kansai | 422,826,938 | 421,510,186 | 63,424,040,700 | 63,311,416,076 | 99.69 | 6.42 | 21.10 | 98.05 |
| NAG12352 | Kyoto Univ. | Kansai | 389,739,164 | 386,866,005 | 58,460,874,600 | 58,064,462,541 | 99.26 | 5.26 | 19.35 | 95.93 |
| NAG12383 | Kyoto Univ. | Kansai | 380,965,602 | 378,526,201 | 57,144,840,300 | 56,857,368,179 | 99.36 | 7.78 | 18.95 | 91.37 |
| NAG12385 | Kyoto Univ. | Kansai | 359,320,632 | 356,883,482 | 53,898,094,800 | 53,567,058,520 | 99.32 | 4.56 | 17.86 | 94.03 |
| NAG12395 | Kyoto Univ. | Kansai | 375,014,890 | 372,633,527 | 56,252,233,500 | 56,009,327,102 | 99.36 | 5.33 | 18.67 | 96.01 |
| NAG12413 | Kyoto Univ. | Kansai | 347,224,737 | 343,158,896 | 52,083,710,550 | 51,540,437,136 | 98.83 | 4.51 | 17.18 | 93.17 |
| NAG12417 | Kyoto Univ. | Kansai | 380,730,627 | 377,666,992 | 57,109,594,050 | 56,742,682,405 | 99.20 | 6.89 | 18.91 | 95.04 |
| NAG12420 | Kyoto Univ. | Kansai | 400,796,924 | 399,502,881 | 60,119,538,600 | 60,028,115,197 | 99.68 | 8.17 | 20.01 | 94.13 |
| NAG12423 | Kyoto Univ. | Kansai | 439,157,261 | 437,779,182 | 65,873,589,150 | 65,702,475,470 | 99.69 | 8.11 | 21.90 | 98.11 |
| NAG12438 | Kyoto Univ. | Kansai | 333,120,387 | 331,227,137 | 49,968,058,050 | 49,740,968,263 | 99.43 | 8.96 | 16.58 | 85.15 |
| NAG12440 | Kyoto Univ. | Kansai | 345,319,051 | 341,977,463 | 51,797,857,650 | 51,330,883,751 | 99.03 | 4.32 | 17.11 | 92.87 |
| NAG12480 | Kyoto Univ. | Kansai | 386,495,121 | 384,416,097 | 57,974,268,150 | 57,727,237,180 | 99.46 | 4.94 | 19.24 | 96.76 |
| NAG12492 | Kyoto Univ. | Kansai | 403,287,458 | 401,987,132 | 60,493,118,700 | 60,416,444,019 | 99.68 | 6.65 | 20.14 | 94.57 |
| NAG12496 | Kyoto Univ. | Kansai | 376,908,217 | 374,477,427 | 56,536,232,550 | 56,252,946,437 | 99.36 | 5.85 | 18.75 | 91.78 |
| NAG12500 | Kyoto Univ. | Kansai | 338,986,241 | 336,889,505 | 50,847,936,150 | 50,643,376,495 | 99.38 | 4.63 | 16.88 | 93.11 |
| NAG12508 | Kyoto Univ. | Kansai | 384,443,292 | 383,055,759 | 57,666,493,800 | 57,522,087,594 | 99.64 | 7.79 | 19.17 | 92.59 |
| NAG12519 | Kyoto Univ. | Kansai | 348,130,307 | 345,633,857 | 52,219,546,050 | 51,930,794,850 | 99.28 | 4.08 | 17.31 | 93.98 |
| NAG12520 | Kyoto Univ. | Kansai | 337,968,673 | 335,024,270 | 50,695,300,950 | 50,337,104,391 | 99.13 | 4.41 | 16.78 | 92.17 |
| NAG12536 | Kyoto Univ. | Kansai | 416,193,870 | 413,341,351 | 62,429,080,500 | 62,071,748,156 | 99.31 | 5.04 | 20.69 | 96.69 |
| NAG12541 | Kyoto Univ. | Kansai | 394,734,819 | 393,286,366 | 59,210,222,850 | 59,083,690,217 | 99.63 | 8.04 | 19.69 | 96.23 |
| NAG12558 | Kyoto Univ. | Kansai | 382,777,757 | 381,464,543 | 57,416,663,550 | 57,266,852,006 | 99.66 | 6.11 | 19.09 | 92.91 |
| NAG12567 | Kyoto Univ. | Kansai | 336,060,883 | 334,094,280 | 50,409,132,450 | 50,137,451,203 | 99.41 | 4.50 | 16.71 | 87.71 |
| NAG12573 | Kyoto Univ. | Kansai | 377,980,952 | 375,444,671 | 56,697,142,800 | 56,422,651,158 | 99.33 | 4.40 | 18.81 | 96.27 |
| NAG12575 | Kyoto Univ. | Kansai | 412,045,788 | 409,670,071 | 61,806,868,200 | 61,568,477,120 | 99.42 | 5.54 | 20.52 | 96.75 |
| NAG12577 | Kyoto Univ. | Kansai | 327,960,061 | 326,522,858 | 49,194,009,150 | 49,084,306,460 | 99.56 | 4.42 | 16.36 | 88.79 |
| NAG12583 | Kyoto Univ. | Kansai | 393,978,844 | 391,676,347 | 59,096,826,600 | 58,784,552,047 | 99.42 | 5.23 | 19.59 | 96.66 |
| NAG12596 | Kyoto Univ. | Kansai | 399,552,317 | 394,642,918 | 59,932,847,550 | 59,204,198,881 | 98.77 | 5.55 | 19.73 | 96.65 |
| NAG12604 | Kyoto Univ. | Kansai | 357,068,807 | 354,540,931 | 53,560,321,050 | 53,253,790,085 | 99.29 | 5.29 | 17.75 | 94.10 |
| NAG12626 | Kyoto Univ. | Kansai | 352,789,101 | 350,345,363 | 52,918,365,150 | 52,579,970,110 | 99.31 | 5.82 | 17.53 | 88.30 |
| NAG12629 | Kyoto Univ. | Kansai | 373,115,513 | 370,639,342 | 55,967,326,950 | 55,672,539,730 | 99.34 | 5.61 | 18.56 | 95.05 |
| NAG12634 | Kyoto Univ. | Kansai | 333,808,028 | 331,536,839 | 50,071,204,200 | 49,823,139,497 | 99.32 | 4.63 | 16.61 | 91.03 |
| NAG12646 | Kyoto Univ. | Kansai | 376,278,241 | 374,804,799 | 56,441,736,150 | 56,283,650,486 | 99.61 | 7.90 | 18.76 | 94.81 |
| NAG12647 | Kyoto Univ. | Kansai | 319,845,823 | 317,501,390 | 47,976,873,450 | 47,696,964,127 | 99.27 | 4.44 | 15.90 | 89.50 |
| NAG12652 | Kyoto Univ. | Kansai | 389,028,743 | 387,086,654 | 58,354,311,450 | 58,184,927,287 | 99.50 | 5.52 | 19.39 | 96.99 |
| NAG12661 | Kyoto Univ. | Kansai | 355,496,561 | 352,542,168 | 53,324,484,150 | 52,953,139,038 | 99.17 | 4.94 | 17.65 | 93.38 |
| NAG12667 | Kyoto Univ. | Kansai | 373,854,010 | 372,015,151 | 56,078,101,500 | 55,902,709,351 | 99.51 | 4.34 | 18.63 | 96.22 |
| NAG12668 | Kyoto Univ. | Kansai | 441,760,804 | 435,436,754 | 66,264,120,600 | 65,400,958,366 | 98.57 | 4.17 | 21.80 | 97.26 |
| NAG12675 | Kyoto Univ. | Kansai | 395,312,452 | 392,712,486 | 59,296,867,800 | 58,993,184,663 | 99.34 | 7.49 | 19.66 | 95.74 |

| | | | | | | | | | | |
|---|---|---|---|---|---|---|---|---|---|---|
| NAG12687 | Kyoto Univ. | Kansai | 373,477,207 | 372,041,564 | 56,021,581,050 | 55,898,659,652 | 99.62 | 5.98 | 18.63 | 92.78 |
| NAG12689 | Kyoto Univ. | Kansai | 359,094,380 | 358,077,106 | 53,864,157,000 | 53,768,944,521 | 99.72 | 6.15 | 17.92 | 94.44 |
| NAG12702 | Kyoto Univ. | Kansai | 325,451,439 | 323,611,154 | 48,817,715,850 | 48,656,690,237 | 99.43 | 4.50 | 16.22 | 88.36 |
| NAG12720 | Kyoto Univ. | Kansai | 372,866,601 | 371,765,213 | 55,929,990,150 | 55,820,337,382 | 99.70 | 7.23 | 18.61 | 94.91 |
| NAG12731 | Kyoto Univ. | Kansai | 339,787,068 | 338,712,548 | 50,968,060,200 | 50,860,212,996 | 99.68 | 6.05 | 16.95 | 88.37 |
| NAG12749 | Kyoto Univ. | Kansai | 347,137,903 | 345,253,449 | 52,070,685,450 | 51,772,342,782 | 99.46 | 5.05 | 17.26 | 91.23 |
| NAG12750 | Kyoto Univ. | Kansai | 319,037,790 | 317,456,896 | 47,855,668,500 | 47,684,140,778 | 99.50 | 4.76 | 15.89 | 89.38 |
| NAG12754 | Kyoto Univ. | Kansai | 338,891,826 | 336,709,592 | 50,833,773,900 | 50,578,926,281 | 99.36 | 4.38 | 16.86 | 92.53 |
| NAG12758 | Kyoto Univ. | Kansai | 380,816,330 | 378,687,837 | 57,122,449,500 | 56,908,386,760 | 99.44 | 6.08 | 18.97 | 95.72 |
| NAG12762 | Kyoto Univ. | Kansai | 513,884,465 | 510,627,558 | 77,082,669,750 | 76,671,902,752 | 99.37 | 10.55 | 25.56 | 98.81 |
| NAG12763 | Kyoto Univ. | Kansai | 359,229,665 | 357,051,506 | 53,884,449,750 | 53,612,564,117 | 99.39 | 4.62 | 17.87 | 94.47 |
| NAG12765 | Kyoto Univ. | Kansai | 368,512,085 | 366,052,288 | 55,276,812,750 | 55,008,438,534 | 99.33 | 5.16 | 18.34 | 91.60 |
| NAG12767 | Kyoto Univ. | Kansai | 357,782,196 | 355,668,770 | 53,667,329,400 | 53,431,907,945 | 99.41 | 4.52 | 17.81 | 93.37 |
| NAG12770 | Kyoto Univ. | Kansai | 356,349,151 | 354,416,291 | 53,452,372,650 | 53,265,066,631 | 99.46 | 5.26 | 17.76 | 94.43 |
| NAG12773 | Kyoto Univ. | Kansai | 338,336,258 | 335,885,353 | 50,750,438,700 | 50,413,705,073 | 99.28 | 3.29 | 16.80 | 89.05 |
| NAG12790 | Kyoto Univ. | Kansai | 319,194,455 | 316,460,722 | 47,879,168,250 | 47,524,801,139 | 99.14 | 4.89 | 15.84 | 88.70 |
| NAG12793 | Kyoto Univ. | Kansai | 377,161,749 | 375,466,225 | 56,574,262,350 | 56,402,041,417 | 99.55 | 7.15 | 18.80 | 95.61 |
| NAG12796 | Kyoto Univ. | Kansai | 331,512,071 | 329,474,397 | 49,726,810,650 | 49,509,085,750 | 99.39 | 5.10 | 16.50 | 91.17 |
| NAG12864 | Kyoto Univ. | Kansai | 304,409,675 | 302,896,987 | 45,661,451,250 | 45,481,292,441 | 99.50 | 4.96 | 15.16 | 85.62 |
| NAG12869 | Kyoto Univ. | Kansai | 378,048,679 | 375,825,161 | 56,707,301,850 | 56,486,370,389 | 99.41 | 5.44 | 18.83 | 92.27 |
| NAG12873 | Kyoto Univ. | Kansai | 374,689,984 | 372,739,877 | 56,203,497,600 | 56,007,732,897 | 99.48 | 5.72 | 18.67 | 94.24 |
| NAG12882 | Kyoto Univ. | Kansai | 388,202,785 | 384,842,761 | 58,230,417,750 | 57,783,071,729 | 99.13 | 5.06 | 19.26 | 93.29 |
| NAG12890 | Kyoto Univ. | Kansai | 364,192,374 | 362,823,384 | 54,628,856,100 | 54,537,926,168 | 99.62 | 6.14 | 18.18 | 95.36 |
| NAG12919 | Kyoto Univ. | Kansai | 365,596,215 | 363,489,032 | 54,839,432,250 | 54,642,032,488 | 99.42 | 5.03 | 18.21 | 95.66 |
| NAG12934 | Kyoto Univ. | Kansai | 386,340,172 | 383,743,349 | 57,951,025,800 | 57,595,388,703 | 99.33 | 5.44 | 19.20 | 92.16 |
| NAG12945 | Kyoto Univ. | Kansai | 347,515,975 | 346,098,126 | 52,127,396,250 | 51,959,361,307 | 99.59 | 7.87 | 17.32 | 89.07 |
| NAG12949 | Kyoto Univ. | Kansai | 361,335,628 | 359,040,912 | 54,200,344,200 | 53,923,474,006 | 99.36 | 4.98 | 17.97 | 90.63 |
| NAG12954 | Kyoto Univ. | Kansai | 334,729,809 | 333,648,958 | 50,209,471,350 | 50,058,327,865 | 99.68 | 6.33 | 16.69 | 89.66 |
| NAG12960 | Kyoto Univ. | Kansai | 384,170,285 | 382,490,088 | 57,625,542,750 | 57,481,865,487 | 99.56 | 6.39 | 19.16 | 93.21 |
| NAG12963 | Kyoto Univ. | Kansai | 356,873,141 | 354,750,797 | 53,530,971,150 | 53,275,222,747 | 99.41 | 5.76 | 17.76 | 91.11 |
| NAG12965 | Kyoto Univ. | Kansai | 307,691,149 | 304,571,402 | 46,153,672,350 | 45,765,286,148 | 98.99 | 4.08 | 15.26 | 87.12 |
| NAG12972 | Kyoto Univ. | Kansai | 387,320,925 | 384,640,005 | 58,098,138,750 | 57,797,394,197 | 99.31 | 6.32 | 19.27 | 96.01 |
| NAG12980 | Kyoto Univ. | Kansai | 359,795,183 | 358,352,594 | 53,969,277,450 | 53,802,860,615 | 99.60 | 5.68 | 17.93 | 94.24 |
| NAG12982 | Kyoto Univ. | Kansai | 376,068,379 | 374,098,729 | 56,410,256,850 | 56,171,878,592 | 99.48 | 5.78 | 18.72 | 95.42 |
| NAG12998 | Kyoto Univ. | Kansai | 355,592,116 | 354,164,505 | 53,338,817,400 | 53,230,526,348 | 99.60 | 6.00 | 17.74 | 93.92 |
| NAG13007 | Kyoto Univ. | Kansai | 344,684,617 | 343,329,731 | 51,702,692,550 | 51,603,294,083 | 99.61 | 5.93 | 17.20 | 93.35 |
| NAG13014 | Kyoto Univ. | Kansai | 397,679,604 | 395,989,269 | 59,651,940,600 | 59,509,429,326 | 99.57 | 6.20 | 19.84 | 96.50 |
| NAG13037 | Kyoto Univ. | Kansai | 359,174,891 | 356,841,842 | 53,876,233,650 | 53,610,116,292 | 99.35 | 5.76 | 17.87 | 93.28 |
| NAG13043 | Kyoto Univ. | Kansai | 412,698,928 | 409,165,746 | 61,904,839,200 | 61,427,752,937 | 99.14 | 5.62 | 20.48 | 97.47 |
| NAG13083 | Kyoto Univ. | Kansai | 357,043,407 | 354,712,346 | 53,556,511,050 | 53,234,439,057 | 99.35 | 5.88 | 17.74 | 93.63 |
| NAG13095 | Kyoto Univ. | Kansai | 471,608,335 | 468,239,116 | 70,741,250,250 | 70,307,085,652 | 99.29 | 7.47 | 23.44 | 98.72 |
| NAG13098 | Kyoto Univ. | Kansai | 370,955,837 | 368,629,358 | 55,643,375,550 | 55,393,328,522 | 99.37 | 5.21 | 18.46 | 93.60 |
| NAG13105 | Kyoto Univ. | Kansai | 417,362,230 | 415,492,551 | 62,604,334,500 | 62,378,548,078 | 99.55 | 6.27 | 20.79 | 88.30 |
| NAG13110 | Kyoto Univ. | Kansai | 386,499,717 | 384,819,992 | 57,974,957,550 | 57,784,765,114 | 99.57 | 6.14 | 19.26 | 93.43 |
| NAG13112 | Kyoto Univ. | Kansai | 332,479,042 | 330,635,571 | 49,871,856,300 | 49,674,252,933 | 99.45 | 6.42 | 16.56 | 87.21 |
| NAG13130 | Kyoto Univ. | Kansai | 390,464,297 | 388,721,512 | 58,569,644,550 | 58,420,491,275 | 99.55 | 6.43 | 19.47 | 96.51 |
| NAG13134 | Kyoto Univ. | Kansai | 365,214,990 | 363,395,422 | 54,782,248,500 | 54,547,715,031 | 99.50 | 4.87 | 18.18 | 95.14 |
| NAG13136 | Kyoto Univ. | Kansai | 360,551,518 | 358,933,461 | 54,082,727,700 | 53,931,040,323 | 99.55 | 5.57 | 17.98 | 94.63 |
| NAG13154 | Kyoto Univ. | Kansai | 350,650,589 | 348,678,869 | 52,597,588,350 | 52,381,663,201 | 99.44 | 5.28 | 17.46 | 88.89 |
| NAG13163 | Kyoto Univ. | Kansai | 350,746,613 | 346,753,722 | 52,611,991,950 | 52,095,450,643 | 98.86 | 4.47 | 17.37 | 93.41 |
| NAG13167 | Kyoto Univ. | Kansai | 354,440,948 | 352,242,074 | 53,166,142,200 | 52,911,665,454 | 99.38 | 5.13 | 17.64 | 92.57 |
| NAG13188 | Kyoto Univ. | Kansai | 369,021,719 | 367,720,985 | 55,353,257,850 | 55,271,603,129 | 99.65 | 6.32 | 18.42 | 95.02 |
| NAG13208 | Kyoto Univ. | Kansai | 367,840,786 | 365,954,447 | 55,176,117,900 | 55,004,782,385 | 99.49 | 6.56 | 18.33 | 92.18 |
| NAG13209 | Kyoto Univ. | Kansai | 335,309,257 | 332,366,836 | 50,296,388,550 | 49,901,360,975 | 99.12 | 4.70 | 16.63 | 88.72 |
| NAG13217 | Kyoto Univ. | Kansai | 401,109,653 | 399,730,513 | 60,166,447,950 | 60,102,867,852 | 99.66 | 6.77 | 20.03 | 97.38 |
| NAG13220 | Kyoto Univ. | Kansai | 381,083,217 | 379,210,134 | 57,162,482,550 | 56,985,435,501 | 99.51 | 6.34 | 19.00 | 93.36 |
| NAG13231 | Kyoto Univ. | Kansai | 339,418,426 | 337,556,494 | 50,912,763,900 | 50,547,103,627 | 99.45 | 5.15 | 16.85 | 86.02 |
| NAG13232 | Kyoto Univ. | Kansai | 370,784,102 | 368,917,160 | 55,617,615,300 | 55,369,136,478 | 99.50 | 7.15 | 18.46 | 90.95 |
| NAG13241 | Kyoto Univ. | Kansai | 357,005,971 | 353,501,206 | 53,550,895,650 | 53,119,151,958 | 99.02 | 4.53 | 17.71 | 94.43 |
| NAG13245 | Kyoto Univ. | Kansai | 331,834,024 | 329,329,421 | 49,775,103,600 | 49,472,461,353 | 99.25 | 4.85 | 16.49 | 91.63 |
| NAG13253 | Kyoto Univ. | Kansai | 362,837,435 | 360,829,424 | 54,425,615,250 | 54,230,185,670 | 99.45 | 4.99 | 18.08 | 91.54 |
| NAG13260 | Kyoto Univ. | Kansai | 337,548,124 | 334,714,380 | 50,632,218,600 | 50,278,124,037 | 99.16 | 4.68 | 16.76 | 92.12 |
| NAG13261 | Kyoto Univ. | Kansai | 363,124,096 | 360,656,515 | 54,468,614,400 | 54,116,280,197 | 99.32 | 5.06 | 18.04 | 91.21 |
| NAG13271 | Kyoto Univ. | Kansai | 338,261,436 | 336,612,232 | 50,739,215,400 | 50,595,988,718 | 99.51 | 5.96 | 16.87 | 91.77 |
| NAG13275 | Kyoto Univ. | Kansai | 380,458,741 | 378,761,831 | 57,068,811,150 | 56,896,287,912 | 99.55 | 5.18 | 18.97 | 93.51 |
| NAG13279 | Kyoto Univ. | Kansai | 384,507,027 | 383,018,093 | 57,676,054,050 | 57,556,262,120 | 99.61 | 5.25 | 19.19 | 96.61 |
| NAG13292 | Kyoto Univ. | Kansai | 352,550,561 | 351,241,125 | 52,882,584,150 | 52,770,402,488 | 99.63 | 5.25 | 17.59 | 93.97 |
| NAG13303 | Kyoto Univ. | Kansai | 338,060,899 | 336,081,451 | 50,709,134,850 | 50,513,979,380 | 99.41 | 5.52 | 16.84 | 91.51 |
| NAG13315 | Kyoto Univ. | Kansai | 344,438,946 | 342,469,621 | 51,665,841,900 | 51,444,338,994 | 99.43 | 4.78 | 17.15 | 90.98 |
| NAG13328 | Kyoto Univ. | Kansai | 347,535,956 | 346,223,471 | 52,130,393,400 | 52,013,826,813 | 99.62 | 5.35 | 17.34 | 90.10 |
| NAG13333 | Kyoto Univ. | Kansai | 379,247,013 | 370,172,653 | 56,887,051,950 | 55,605,761,738 | 97.61 | 3.17 | 18.54 | 92.67 |
| NAG13336 | Kyoto Univ. | Kansai | 414,310,336 | 412,463,514 | 62,146,550,400 | 61,866,555,863 | 99.55 | 5.74 | 20.62 | 96.99 |
| NAG13339 | Kyoto Univ. | Kansai | 360,038,673 | 358,418,863 | 54,005,800,950 | 53,876,155,260 | 99.55 | 5.89 | 17.96 | 95.01 |
| NAG13347 | Kyoto Univ. | Kansai | 346,578,696 | 344,837,499 | 51,986,804,400 | 51,841,184,017 | 99.50 | 6.11 | 17.28 | 90.32 |
| NAG13350 | Kyoto Univ. | Kansai | 377,472,820 | 375,138,438 | 56,620,923,000 | 56,382,311,665 | 99.38 | 5.17 | 18.79 | 92.60 |
| NAG13364 | Kyoto Univ. | Kansai | 340,274,500 | 338,071,589 | 51,041,175,000 | 50,769,271,981 | 99.35 | 3.65 | 16.92 | 88.76 |
| NAG13365 | Kyoto Univ. | Kansai | 330,577,444 | 329,251,124 | 49,586,616,600 | 49,462,958,108 | 99.60 | 5.41 | 16.49 | 91.00 |
| NAG13373 | Kyoto Univ. | Kansai | 352,514,586 | 351,214,790 | 52,877,187,900 | 52,721,899,666 | 99.63 | 5.79 | 17.57 | 93.06 |
| NAG13379 | Kyoto Univ. | Kansai | 386,528,840 | 384,009,636 | 57,979,326,000 | 57,641,281,541 | 99.35 | 5.39 | 19.21 | 95.45 |
| NAG13380 | Kyoto Univ. | Kansai | 346,216,663 | 343,471,663 | 51,932,499,450 | 51,579,289,698 | 99.21 | 5.14 | 17.19 | 92.63 |
| NAG13381 | Kyoto Univ. | Kansai | 337,166,144 | 335,024,686 | 50,574,921,600 | 50,345,004,274 | 99.36 | 4.75 | 16.78 | 92.50 |
| NAG13390 | Kyoto Univ. | Kansai | 361,298,802 | 359,862,967 | 54,194,820,300 | 54,065,285,924 | 99.60 | 6.02 | 18.02 | 94.19 |
| NAG13391 | Kyoto Univ. | Kansai | 339,641,703 | 337,259,189 | 50,946,255,450 | 50,678,724,329 | 99.30 | 5.11 | 16.89 | 92.54 |
| NAG13402 | Kyoto Univ. | Kansai | 332,380,394 | 330,428,112 | 49,857,059,100 | 49,531,394,868 | 99.41 | 4.48 | 16.51 | 89.08 |
| NAG13405 | Kyoto Univ. | Kansai | 376,132,844 | 374,727,517 | 56,419,926,600 | 56,260,161,700 | 99.63 | 5.61 | 18.75 | 95.41 |
| NAG13410 | Kyoto Univ. | Kansai | 362,254,890 | 359,610,124 | 54,338,233,500 | 54,017,800,815 | 99.27 | 4.84 | 18.01 | 91.17 |
| NAG13426 | Kyoto Univ. | Kansai | 414,399,264 | 412,491,309 | 62,159,889,600 | 61,971,231,881 | 99.54 | 8.30 | 20.66 | 97.46 |
| NAG13430 | Kyoto Univ. | Kansai | 353,291,302 | 351,885,050 | 52,993,695,300 | 52,798,687,607 | 99.60 | 5.73 | 17.60 | 92.90 |
| NAG13433 | Kyoto Univ. | Kansai | 314,995,613 | 312,599,958 | 47,249,341,950 | 46,961,364,960 | 99.24 | 3.08 | 15.65 | 88.92 |
| NAG13434 | Kyoto Univ. | Kansai | 363,872,038 | 361,637,049 | 54,580,805,700 | 54,349,108,252 | 99.39 | 5.06 | 18.12 | 92.05 |
| NAG13440 | Kyoto Univ. | Kansai | 308,162,036 | 305,674,921 | 46,224,305,400 | 45,940,420,142 | 99.19 | 4.08 | 15.31 | 87.53 |
| NAG13447 | Kyoto Univ. | Kansai | 374,532,516 | 373,084,834 | 56,179,877,400 | 56,053,103,472 | 99.61 | 5.00 | 18.68 | 95.92 |
| NAG13453 | Kyoto Univ. | Kansai | 316,981,412 | 315,439,846 | 47,547,211,800 | 47,353,070,133 | 99.51 | 5.22 | 15.78 | 83.52 |
| NAG13456 | Kyoto Univ. | Kansai | 346,515,579 | 343,707,180 | 51,977,336,850 | 51,628,651,067 | 99.19 | 3.71 | 17.21 | 90.09 |
| NAG13458 | Kyoto Univ. | Kansai | 371,706,459 | 368,802,588 | 55,755,968,850 | 55,422,611,311 | 99.22 | 5.66 | 18.47 | 95.59 |
| NAG13459 | Kyoto Univ. | Kansai | 382,586,235 | 378,910,682 | 57,387,935,250 | 56,814,334,978 | 99.04 | 6.27 | 18.94 | 91.17 |
| NAG13460 | Kyoto Univ. | Kansai | 367,110,089 | 365,557,347 | 55,066,513,350 | 54,952,656,324 | 99.58 | 5.72 | 18.32 | 94.96 |
| NAG13500 | Kyoto Univ. | Kansai | 430,089,960 | 428,385,347 | 64,513,494,000 | 64,349,984,878 | 99.60 | 8.90 | 21.45 | 95.29 |
| NAG13503 | Kyoto Univ. | Kansai | 395,707,808 | 393,923,602 | 59,356,171,200 | 59,173,927,608 | 99.55 | 8.36 | 19.72 | 93.54 |
| NAG13528 | Kyoto Univ. | Kansai | 392,968,858 | 391,640,068 | 58,945,328,700 | 58,767,537,141 | 99.66 | 8.49 | 19.59 | 92.68 |

| | | | | | | | | | | |
|---|---|---|---|---|---|---|---|---|---|---|
| NAG13540 | Kyoto Univ. | Kansai | 384,952,438 | 381,803,757 | 57,742,865,700 | 57,355,414,752 | 99.18 | 5.21 | 19.12 | 93.42 |
| NAG13544 | Kyoto Univ. | Kansai | 357,829,863 | 355,798,532 | 53,674,479,450 | 53,445,143,467 | 99.43 | 5.57 | 17.82 | 94.14 |
| NAG13549 | Kyoto Univ. | Kansai | 314,471,086 | 311,824,352 | 47,170,662,900 | 46,826,462,361 | 99.16 | 3.01 | 15.61 | 85.47 |
| NAG13554 | Kyoto Univ. | Kansai | 402,679,949 | 400,377,124 | 60,401,992,350 | 60,184,276,642 | 99.43 | 6.06 | 20.06 | 97.46 |
| NAG13563 | Kyoto Univ. | Kansai | 337,922,039 | 335,639,595 | 50,688,305,850 | 50,393,081,797 | 99.32 | 3.91 | 16.80 | 89.12 |
| NAG13574 | Kyoto Univ. | Kansai | 368,896,386 | 367,250,738 | 55,334,457,900 | 55,168,123,152 | 99.55 | 8.02 | 18.39 | 94.67 |
| NAG13576 | Kyoto Univ. | Kansai | 337,950,596 | 336,341,980 | 50,692,589,400 | 50,558,819,866 | 99.52 | 5.02 | 16.85 | 92.02 |
| NAG13580 | Kyoto Univ. | Kansai | 333,064,976 | 330,258,383 | 49,959,746,400 | 49,632,954,434 | 99.16 | 5.13 | 16.54 | 91.42 |
| NAG13581 | Kyoto Univ. | Kansai | 346,168,238 | 342,545,131 | 51,925,235,700 | 51,446,607,225 | 98.95 | 4.48 | 17.15 | 92.88 |
| NAG13582 | Kyoto Univ. | Kansai | 366,932,793 | 364,334,246 | 55,039,918,950 | 54,628,252,983 | 99.29 | 5.53 | 18.21 | 94.05 |
| NAG13594 | Kyoto Univ. | Kansai | 493,853,914 | 490,016,787 | 74,078,087,100 | 73,483,447,332 | 99.22 | 8.66 | 24.49 | 98.47 |
| NAG13622 | Kyoto Univ. | Kansai | 374,043,054 | 371,096,836 | 56,106,458,100 | 55,768,516,816 | 99.21 | 5.21 | 18.59 | 95.80 |
| NAG13630 | Kyoto Univ. | Kansai | 344,519,581 | 342,226,796 | 51,677,937,150 | 51,414,984,028 | 99.33 | 6.10 | 17.14 | 92.77 |
| NAG13634 | Kyoto Univ. | Kansai | 373,240,644 | 370,812,519 | 55,986,096,600 | 55,722,123,156 | 99.35 | 4.80 | 18.57 | 95.95 |
| NAG13636 | Kyoto Univ. | Kansai | 374,317,075 | 372,787,501 | 56,147,561,250 | 56,037,536,685 | 99.59 | 6.62 | 18.68 | 95.22 |
| NAG13638 | Kyoto Univ. | Kansai | 382,963,314 | 380,966,753 | 57,444,497,100 | 57,217,888,911 | 99.48 | 4.80 | 19.07 | 92.62 |
| NAG13651 | Kyoto Univ. | Kansai | 389,394,001 | 386,202,174 | 58,409,100,150 | 57,964,269,986 | 99.18 | 4.43 | 19.32 | 92.86 |
| NAG13653 | Kyoto Univ. | Kansai | 335,132,473 | 333,212,171 | 50,269,870,950 | 50,053,813,786 | 99.43 | 4.30 | 16.68 | 88.78 |
| NAG13656 | Kyoto Univ. | Kansai | 348,041,984 | 345,553,819 | 52,206,297,600 | 51,923,882,675 | 99.29 | 4.21 | 17.31 | 93.78 |
| NAG13657 | Kyoto Univ. | Kansai | 369,324,474 | 366,905,192 | 55,398,671,100 | 55,100,147,536 | 99.34 | 6.13 | 18.37 | 91.63 |
| NAG13662 | Kyoto Univ. | Kansai | 394,262,135 | 391,894,229 | 59,139,320,250 | 58,878,318,693 | 99.40 | 4.66 | 19.63 | 95.52 |
| NAG13673 | Kyoto Univ. | Kansai | 486,991,946 | 485,597,657 | 73,048,791,900 | 72,964,651,029 | 99.71 | 9.14 | 24.32 | 99.04 |
| NAG13674 | Kyoto Univ. | Kansai | 350,968,811 | 349,482,760 | 52,645,321,650 | 52,540,880,611 | 99.58 | 6.95 | 17.51 | 93.74 |
| NAG13678 | Kyoto Univ. | Kansai | 345,031,004 | 342,453,863 | 51,754,650,600 | 51,444,303,619 | 99.25 | 5.60 | 17.15 | 92.44 |
| NAG13686 | Kyoto Univ. | Kansai | 406,214,500 | 405,084,141 | 60,932,175,000 | 60,887,752,594 | 99.72 | 8.29 | 20.30 | 97.33 |
| NAG13693 | Kyoto Univ. | Kansai | 353,736,686 | 352,240,059 | 53,060,502,900 | 52,935,588,083 | 99.58 | 5.03 | 17.65 | 90.29 |
| NAG13698 | Kyoto Univ. | Kansai | 386,524,227 | 385,396,376 | 57,978,634,050 | 57,870,192,145 | 99.71 | 8.92 | 19.29 | 95.44 |
| NAG13700 | Kyoto Univ. | Kansai | 372,229,701 | 370,252,049 | 55,834,455,150 | 55,647,862,724 | 99.47 | 5.29 | 18.55 | 94.99 |
| NAG13708 | Kyoto Univ. | Kansai | 372,720,295 | 371,608,374 | 55,908,044,250 | 55,825,572,594 | 99.70 | 8.07 | 18.61 | 91.96 |
| NAG13709 | Kyoto Univ. | Kansai | 332,689,483 | 330,231,450 | 49,903,422,450 | 49,577,151,219 | 99.26 | 4.65 | 16.53 | 87.75 |
| NAG13710 | Kyoto Univ. | Kansai | 392,582,458 | 391,226,365 | 58,887,368,700 | 58,778,485,809 | 99.65 | 6.60 | 19.59 | 93.96 |
| NAG13723 | Kyoto Univ. | Kansai | 391,214,869 | 390,040,944 | 58,682,230,350 | 58,609,346,176 | 99.70 | 8.51 | 19.54 | 96.40 |
| NAG13727 | Kyoto Univ. | Kansai | 386,003,645 | 383,306,231 | 57,900,546,750 | 57,596,420,959 | 99.30 | 5.86 | 19.20 | 96.60 |
| NAG13736 | Kyoto Univ. | Kansai | 341,093,122 | 339,339,112 | 51,163,968,300 | 50,993,473,341 | 99.49 | 4.94 | 17.00 | 92.24 |
| NAG13737 | Kyoto Univ. | Kansai | 376,305,901 | 374,039,626 | 56,445,885,150 | 56,189,162,020 | 99.40 | 5.72 | 18.73 | 92.59 |
| NAG13751 | Kyoto Univ. | Kansai | 392,084,004 | 390,800,293 | 58,812,600,600 | 58,705,471,963 | 99.67 | 9.01 | 19.57 | 96.17 |
| NAG13764 | Kyoto Univ. | Kansai | 365,945,709 | 363,341,117 | 54,891,856,350 | 54,608,919,998 | 99.29 | 5.58 | 18.20 | 91.51 |
| NAG13767 | Kyoto Univ. | Kansai | 450,578,154 | 448,129,272 | 67,586,723,100 | 67,229,145,266 | 99.46 | 10.44 | 22.41 | 97.67 |
| NAG13769 | Kyoto Univ. | Kansai | 383,761,530 | 382,045,186 | 57,564,229,500 | 57,415,250,208 | 99.55 | 5.82 | 19.14 | 92.88 |
| NAG13785 | Kyoto Univ. | Kansai | 385,961,214 | 383,733,057 | 57,894,182,100 | 57,690,793,885 | 99.42 | 5.86 | 19.23 | 96.58 |
| NAG13786 | Kyoto Univ. | Kansai | 373,134,802 | 369,829,566 | 55,970,220,300 | 55,547,264,170 | 99.11 | 6.04 | 18.52 | 95.42 |
| NAG13791 | Kyoto Univ. | Kansai | 339,367,265 | 337,893,106 | 50,905,089,750 | 50,796,106,298 | 99.57 | 4.95 | 16.93 | 93.17 |
| NAG13803 | Kyoto Univ. | Kansai | 371,643,104 | 368,903,174 | 55,746,465,600 | 55,408,092,486 | 99.26 | 4.35 | 18.47 | 94.89 |
| NAG13812 | Kyoto Univ. | Kansai | 362,216,637 | 361,143,670 | 54,332,495,550 | 54,258,600,471 | 99.70 | 8.96 | 18.09 | 90.61 |
| NAG13819 | Kyoto Univ. | Kansai | 342,504,996 | 340,103,291 | 51,375,749,400 | 51,103,501,411 | 99.30 | 4.45 | 17.03 | 93.10 |
| NAG13828 | Kyoto Univ. | Kansai | 371,763,402 | 369,016,497 | 55,764,510,300 | 55,433,870,234 | 99.26 | 5.95 | 18.48 | 91.99 |
| NAG13829 | Kyoto Univ. | Kansai | 361,296,016 | 359,251,621 | 54,194,402,400 | 53,904,964,541 | 99.43 | 5.70 | 17.97 | 91.18 |
| NAG13839 | Kyoto Univ. | Kansai | 361,060,231 | 358,409,042 | 54,159,034,650 | 53,824,994,785 | 99.27 | 4.06 | 17.94 | 94.84 |
| NAG13852 | Kyoto Univ. | Kansai | 372,590,014 | 370,553,565 | 55,888,502,100 | 55,703,469,105 | 99.45 | 5.53 | 18.57 | 95.87 |
| NAG13860 | Kyoto Univ. | Kansai | 404,270,336 | 401,712,792 | 60,640,550,400 | 60,251,857,041 | 99.37 | 6.75 | 20.08 | 96.28 |
| NAG13880 | Kyoto Univ. | Kansai | 308,924,107 | 307,170,313 | 46,338,616,050 | 46,149,741,105 | 99.43 | 4.37 | 15.38 | 87.93 |
| NAG13898 | Kyoto Univ. | Kansai | 362,611,817 | 360,920,454 | 54,391,772,550 | 54,161,315,452 | 99.53 | 5.78 | 18.05 | 91.07 |
| NAG13900 | Kyoto Univ. | Kansai | 373,128,829 | 371,606,316 | 55,969,324,350 | 55,813,859,695 | 99.59 | 6.17 | 18.60 | 91.74 |
| NAG13905 | Kyoto Univ. | Kansai | 396,597,662 | 394,576,053 | 59,489,649,300 | 59,197,466,116 | 99.49 | 6.84 | 19.73 | 96.51 |
| NAG13911 | Kyoto Univ. | Kansai | 381,332,130 | 378,845,921 | 57,199,819,500 | 56,920,971,754 | 99.35 | 5.95 | 18.97 | 96.21 |
| NAG13927 | Kyoto Univ. | Kansai | 400,476,650 | 398,865,425 | 60,071,497,500 | 59,914,477,801 | 99.60 | 7.16 | 19.97 | 97.10 |
| NAG13933 | Kyoto Univ. | Kansai | 353,634,097 | 351,301,707 | 53,045,114,550 | 52,807,573,380 | 99.34 | 4.23 | 17.60 | 94.34 |
| NAG13937 | Kyoto Univ. | Kansai | 360,976,803 | 357,620,138 | 54,146,520,450 | 53,674,987,494 | 99.07 | 4.89 | 17.89 | 93.95 |
| NAG13961 | Kyoto Univ. | Kansai | 340,304,580 | 337,805,525 | 51,045,687,000 | 50,752,536,960 | 99.27 | 4.79 | 16.92 | 91.88 |
| NAG13969 | Kyoto Univ. | Kansai | 321,857,512 | 319,885,179 | 48,278,626,800 | 48,080,526,756 | 99.39 | 4.89 | 16.03 | 90.02 |
| NAG13978 | Kyoto Univ. | Kansai | 347,054,790 | 345,470,986 | 52,058,218,500 | 51,909,060,379 | 99.54 | 4.89 | 17.30 | 90.50 |
| NAG13984 | Kyoto Univ. | Kansai | 388,304,039 | 386,764,721 | 58,245,605,850 | 58,094,358,644 | 99.60 | 5.75 | 19.36 | 92.82 |
| NAG13994 | Kyoto Univ. | Kansai | 345,265,547 | 344,305,157 | 51,789,832,050 | 51,654,420,727 | 99.72 | 8.81 | 17.22 | 89.88 |
| NAG13995 | Kyoto Univ. | Kansai | 348,036,592 | 346,003,915 | 52,205,488,800 | 51,986,673,545 | 99.42 | 5.07 | 17.33 | 93.82 |
| NAG13997 | Kyoto Univ. | Kansai | 327,911,820 | 326,101,745 | 49,186,773,000 | 48,985,935,657 | 99.45 | 4.77 | 16.33 | 91.01 |
| NAG14002 | Kyoto Univ. | Kansai | 363,210,013 | 361,809,809 | 54,481,501,950 | 54,351,331,386 | 99.61 | 6.18 | 18.12 | 94.34 |
| NAG14017 | Kyoto Univ. | Kansai | 319,147,701 | 317,390,206 | 47,872,155,150 | 47,696,609,348 | 99.45 | 4.75 | 15.90 | 89.76 |
| NAG14027 | Kyoto Univ. | Kansai | 365,154,845 | 364,069,446 | 54,773,226,750 | 54,691,534,916 | 99.70 | 8.51 | 18.23 | 94.28 |
| NAG14030 | Kyoto Univ. | Kansai | 348,071,929 | 345,706,316 | 52,210,789,350 | 51,885,911,516 | 99.32 | 5.85 | 17.30 | 91.99 |
| NAG14048 | Kyoto Univ. | Kansai | 339,146,579 | 337,705,654 | 50,871,986,850 | 50,734,919,429 | 99.58 | 4.86 | 16.91 | 91.67 |
| NAG14054 | Kyoto Univ. | Kansai | 310,798,063 | 308,884,191 | 46,619,709,450 | 46,408,199,956 | 99.38 | 4.13 | 15.47 | 88.31 |
| NAG14055 | Kyoto Univ. | Kansai | 394,061,119 | 392,817,564 | 59,109,167,850 | 58,986,406,844 | 99.68 | 7.81 | 19.66 | 96.26 |
| NAG14060 | Kyoto Univ. | Kansai | 371,481,846 | 370,231,792 | 55,722,276,900 | 55,602,124,429 | 99.66 | 7.79 | 18.53 | 94.93 |
| NAG14070 | Kyoto Univ. | Kansai | 341,811,245 | 340,430,964 | 51,271,686,750 | 51,162,868,566 | 99.60 | 5.48 | 17.05 | 92.19 |
| NAG14089 | Kyoto Univ. | Kansai | 396,884,689 | 395,149,454 | 59,532,703,350 | 59,323,075,178 | 99.56 | 5.81 | 19.77 | 93.54 |
| NAG14091 | Kyoto Univ. | Kansai | 407,481,120 | 405,731,971 | 61,122,168,000 | 60,939,799,223 | 99.57 | 6.79 | 20.31 | 97.57 |
| NAG14098 | Kyoto Univ. | Kansai | 371,828,322 | 369,643,669 | 55,774,248,300 | 55,533,912,357 | 99.41 | 5.06 | 18.51 | 92.69 |
| NAG14103 | Kyoto Univ. | Kansai | 364,457,588 | 361,822,672 | 54,668,638,200 | 54,317,587,061 | 99.28 | 6.31 | 18.11 | 90.94 |
| NAG14125 | Kyoto Univ. | Kansai | 401,703,737 | 399,742,276 | 60,255,560,550 | 60,025,323,253 | 99.51 | 5.75 | 20.01 | 96.43 |
| NAG14176 | Kyoto Univ. | Kansai | 376,409,510 | 374,437,642 | 56,461,426,500 | 56,254,277,143 | 99.48 | 5.31 | 18.75 | 95.94 |
| NAG14182 | Kyoto Univ. | Kansai | 488,587,180 | 486,107,056 | 73,288,077,000 | 72,977,998,071 | 99.49 | 6.99 | 24.33 | 98.94 |
| NAG14186 | Kyoto Univ. | Kansai | 392,869,426 | 391,488,614 | 58,930,413,900 | 58,809,053,219 | 99.65 | 6.43 | 19.60 | 96.68 |
| NAG14214 | Kyoto Univ. | Kansai | 338,426,754 | 336,158,871 | 50,764,013,100 | 50,502,598,498 | 99.33 | 5.92 | 16.83 | 91.80 |
| NAG14221 | Kyoto Univ. | Kansai | 340,769,994 | 339,140,436 | 51,115,499,100 | 50,919,727,150 | 99.52 | 4.22 | 16.97 | 92.95 |
| NAG14225 | Kyoto Univ. | Kansai | 335,427,748 | 333,689,627 | 50,314,162,200 | 50,141,857,257 | 99.48 | 4.31 | 16.71 | 89.27 |
| NAG14226 | Kyoto Univ. | Kansai | 368,635,109 | 366,788,801 | 55,295,266,350 | 55,095,177,126 | 99.50 | 6.74 | 18.37 | 94.86 |
| NAG14227 | Kyoto Univ. | Kansai | 346,756,597 | 343,821,478 | 52,013,489,550 | 51,691,954,098 | 99.15 | 5.70 | 17.23 | 93.20 |
| NAG14231 | Kyoto Univ. | Kansai | 425,880,373 | 423,607,922 | 63,882,055,950 | 63,603,486,047 | 99.47 | 4.70 | 21.20 | 94.95 |
| NAG14246 | Kyoto Univ. | Kansai | 376,419,394 | 374,480,777 | 56,462,909,100 | 56,257,789,273 | 99.48 | 4.95 | 18.75 | 92.74 |
| NAG14248 | Kyoto Univ. | Kansai | 354,127,269 | 352,038,882 | 53,119,090,350 | 52,851,020,792 | 99.41 | 7.53 | 17.62 | 92.30 |
| NAG14261 | Kyoto Univ. | Kansai | 345,938,125 | 343,945,528 | 51,890,718,750 | 51,672,978,237 | 99.42 | 6.12 | 17.22 | 92.89 |
| NAG14262 | Kyoto Univ. | Kansai | 409,821,017 | 408,551,373 | 61,473,152,550 | 61,353,666,448 | 99.69 | 8.21 | 20.45 | 97.37 |
| NAG14263 | Kyoto Univ. | Kansai | 345,662,733 | 344,184,758 | 51,849,409,950 | 51,719,017,536 | 99.57 | 4.90 | 17.24 | 93.61 |
| NAG14277 | Kyoto Univ. | Kansai | 342,951,658 | 340,544,392 | 51,442,748,700 | 51,104,138,320 | 99.30 | 4.28 | 17.03 | 89.40 |
| NAG14279 | Kyoto Univ. | Kansai | 395,145,405 | 393,450,082 | 59,271,810,750 | 59,040,053,775 | 99.57 | 7.16 | 19.68 | 95.71 |
| NAG14288 | Kyoto Univ. | Kansai | 332,233,663 | 329,472,818 | 49,835,049,450 | 49,510,504,233 | 99.17 | 6.33 | 16.50 | 90.99 |
| NAG14290 | Kyoto Univ. | Kansai | 483,016,189 | 481,123,972 | 72,452,428,350 | 72,272,028,018 | 99.61 | 9.38 | 24.09 | 98.96 |
| NAG14299 | Kyoto Univ. | Kansai | 311,459,538 | 309,910,555 | 46,718,930,700 | 46,562,426,489 | 99.50 | 4.59 | 15.52 | 88.45 |
| NAG14302 | Kyoto Univ. | Kansai | 404,250,936 | 402,843,636 | 60,637,640,400 | 60,535,088,071 | 99.65 | 7.69 | 20.18 | 94.34 |
| NAG14320 | Kyoto Univ. | Kansai | 381,015,704 | 379,393,499 | 57,152,355,600 | 56,999,749,578 | 99.57 | 8.41 | 19.00 | 95.66 |

| | | | | | | | | | | |
|---|---|---|---|---|---|---|---|---|---|---|
| NAG14321 | Kyoto Univ. | Kansai | 324,237,552 | 322,105,021 | 48,635,632,800 | 48,342,911,463 | 99.34 | 4.35 | 16.11 | 86.36 |
| NAG14324 | Kyoto Univ. | Kansai | 392,860,858 | 390,979,408 | 58,929,128,700 | 58,702,042,649 | 99.52 | 4.82 | 19.57 | 97.11 |
| NAG14331 | Kyoto Univ. | Kansai | 404,849,942 | 403,228,619 | 60,727,491,300 | 60,573,886,641 | 99.60 | 6.67 | 20.19 | 97.43 |
| NAG14332 | Kyoto Univ. | Kansai | 400,741,597 | 399,450,175 | 60,111,239,550 | 59,960,756,758 | 99.68 | 8.37 | 19.99 | 96.36 |
| NAG14335 | Kyoto Univ. | Kansai | 423,297,616 | 422,051,422 | 63,494,642,400 | 63,361,503,035 | 99.71 | 7.88 | 21.12 | 97.81 |
| NAG14348 | Kyoto Univ. | Kansai | 358,071,251 | 356,427,281 | 53,710,687,650 | 53,521,943,278 | 99.54 | 8.01 | 17.84 | 93.26 |
| NAG14353 | Kyoto Univ. | Kansai | 358,948,910 | 356,944,338 | 53,842,336,500 | 53,624,318,468 | 99.44 | 5.09 | 17.87 | 94.65 |
| NAG14355 | Kyoto Univ. | Kansai | 367,097,692 | 364,821,265 | 55,064,653,800 | 54,797,781,695 | 99.38 | 6.69 | 18.27 | 94.43 |
| NAG14363 | Kyoto Univ. | Kansai | 332,952,837 | 331,327,640 | 49,942,925,550 | 49,797,284,858 | 99.51 | 5.21 | 16.60 | 91.89 |
| NAG14381 | Kyoto Univ. | Kansai | 360,780,189 | 359,196,724 | 54,117,028,350 | 53,981,649,397 | 99.56 | 6.41 | 17.99 | 93.92 |
| NAG14388 | Kyoto Univ. | Kansai | 361,325,228 | 359,267,458 | 54,198,784,200 | 53,984,161,741 | 99.43 | 4.59 | 17.99 | 91.99 |
| NAG14391 | Kyoto Univ. | Kansai | 365,674,605 | 363,950,467 | 54,851,190,750 | 54,693,194,803 | 99.53 | 5.86 | 18.23 | 94.43 |
| NAG14392 | Kyoto Univ. | Kansai | 389,291,312 | 387,001,776 | 58,393,696,800 | 58,137,290,058 | 99.41 | 5.65 | 19.38 | 93.03 |
| NAG14396 | Kyoto Univ. | Kansai | 491,724,140 | 488,774,617 | 73,758,621,000 | 73,372,912,621 | 99.40 | 5.09 | 24.46 | 98.85 |
| NAG14403 | Kyoto Univ. | Kansai | 319,374,239 | 317,891,015 | 47,906,135,850 | 47,767,197,029 | 99.54 | 5.06 | 15.92 | 89.77 |
| NAG14410 | Kyoto Univ. | Kansai | 322,311,919 | 319,875,352 | 48,346,787,850 | 48,045,509,244 | 99.24 | 5.12 | 16.02 | 89.52 |
| NAG14416 | Kyoto Univ. | Kansai | 352,646,845 | 350,447,134 | 52,897,026,750 | 52,582,988,654 | 99.38 | 4.67 | 17.53 | 90.14 |
| NAG14418 | Kyoto Univ. | Kansai | 351,604,702 | 348,919,144 | 52,740,705,300 | 52,454,768,320 | 99.24 | 4.71 | 17.48 | 94.10 |
| NAG14425 | Kyoto Univ. | Kansai | 331,265,805 | 329,344,171 | 49,689,870,750 | 49,509,001,245 | 99.42 | 5.85 | 16.50 | 91.48 |
| NAG14427 | Kyoto Univ. | Kansai | 341,234,612 | 338,679,131 | 51,185,191,800 | 50,802,538,421 | 99.25 | 6.11 | 16.93 | 89.57 |
| NAG14430 | Kyoto Univ. | Kansai | 328,806,624 | 326,206,295 | 49,320,993,600 | 48,983,324,221 | 99.21 | 4.47 | 16.33 | 90.36 |
| NAG14433 | Kyoto Univ. | Kansai | 412,636,744 | 411,104,632 | 61,895,511,600 | 61,711,388,066 | 99.63 | 8.01 | 20.57 | 94.37 |
| NAG14434 | Kyoto Univ. | Kansai | 365,413,485 | 364,108,981 | 54,812,022,750 | 54,686,765,234 | 99.64 | 8.42 | 18.23 | 90.87 |
| NAG14441 | Kyoto Univ. | Kansai | 367,519,551 | 365,693,040 | 55,127,932,650 | 54,897,853,414 | 99.50 | 6.48 | 18.30 | 94.28 |
| NAG14453 | Kyoto Univ. | Kansai | 405,954,067 | 402,991,817 | 60,893,110,050 | 60,548,064,448 | 99.27 | 4.88 | 20.18 | 94.78 |
| NAG14458 | Kyoto Univ. | Kansai | 418,726,272 | 415,816,436 | 62,808,940,800 | 62,470,783,499 | 99.31 | 6.12 | 20.82 | 97.75 |
| NAG14461 | Kyoto Univ. | Kansai | 383,627,969 | 380,566,725 | 57,544,195,350 | 57,165,342,058 | 99.20 | 4.98 | 19.06 | 93.13 |
| NAG14476 | Kyoto Univ. | Kansai | 337,317,874 | 335,753,602 | 50,597,681,100 | 50,469,322,852 | 99.54 | 5.74 | 16.82 | 92.09 |
| NAG14478 | Kyoto Univ. | Kansai | 329,920,466 | 328,479,965 | 49,488,069,900 | 49,354,426,581 | 99.56 | 5.44 | 16.45 | 90.14 |
| NAG14486 | Kyoto Univ. | Kansai | 392,195,038 | 389,639,767 | 58,829,255,700 | 58,556,239,967 | 99.35 | 5.23 | 19.52 | 96.95 |
| NAG14488 | Kyoto Univ. | Kansai | 324,602,214 | 322,367,304 | 48,690,332,100 | 48,483,033,724 | 99.31 | 4.88 | 16.16 | 90.96 |
| NAG14489 | Kyoto Univ. | Kansai | 378,545,076 | 377,275,836 | 56,781,761,400 | 56,655,714,157 | 99.66 | 7.77 | 18.89 | 95.39 |
| NAG14503 | Kyoto Univ. | Kansai | 383,397,157 | 381,530,315 | 57,509,573,550 | 57,314,238,107 | 99.51 | 6.45 | 19.10 | 93.27 |
| NAG14545 | Kyoto Univ. | Kansai | 390,539,430 | 387,591,234 | 58,580,914,500 | 58,229,037,538 | 99.25 | 5.58 | 19.41 | 96.47 |
| NAG14546 | Kyoto Univ. | Kansai | 325,945,274 | 324,285,078 | 48,891,791,100 | 48,730,818,525 | 99.49 | 5.23 | 16.24 | 90.80 |
| NAG14572 | Kyoto Univ. | Kansai | 346,934,809 | 344,918,613 | 52,040,221,350 | 51,828,134,673 | 99.42 | 5.21 | 17.28 | 93.45 |
| NAG14573 | Kyoto Univ. | Kansai | 388,709,886 | 386,134,380 | 58,306,482,900 | 57,952,487,689 | 99.34 | 5.44 | 19.32 | 96.23 |
| NAG14583 | Kyoto Univ. | Kansai | 404,541,567 | 402,424,017 | 60,681,235,050 | 60,377,479,750 | 99.48 | 6.61 | 20.13 | 93.74 |
| NAG14586 | Kyoto Univ. | Kansai | 319,721,207 | 317,990,656 | 47,958,181,050 | 47,787,641,889 | 99.46 | 4.50 | 15.93 | 90.07 |
| NAG14591 | Kyoto Univ. | Kansai | 345,664,717 | 343,870,591 | 51,849,707,550 | 51,673,970,881 | 99.48 | 6.04 | 17.22 | 89.81 |
| NAG14595 | Kyoto Univ. | Kansai | 353,870,460 | 352,662,440 | 53,080,569,000 | 52,974,850,693 | 99.66 | 5.72 | 17.66 | 94.22 |
| NAG14608 | Kyoto Univ. | Kansai | 352,295,095 | 350,429,096 | 52,844,264,250 | 52,682,990,978 | 99.47 | 4.65 | 17.56 | 94.61 |
| NAG14613 | Kyoto Univ. | Kansai | 361,392,635 | 358,812,553 | 54,208,895,250 | 53,924,791,666 | 99.29 | 4.04 | 17.97 | 95.31 |
| NAG14616 | Kyoto Univ. | Kansai | 347,623,910 | 346,042,622 | 52,143,586,500 | 52,011,944,861 | 99.55 | 5.32 | 17.34 | 93.12 |
| NAG14644 | Kyoto Univ. | Kansai | 415,872,942 | 413,790,383 | 62,380,941,300 | 62,126,525,303 | 99.50 | 6.67 | 20.71 | 94.01 |
| NAG14647 | Kyoto Univ. | Kansai | 358,628,823 | 355,566,614 | 53,794,323,450 | 53,372,995,786 | 99.15 | 6.04 | 17.79 | 94.00 |
| NAG14669 | Kyoto Univ. | Kansai | 340,574,610 | 338,900,552 | 51,086,191,500 | 50,872,322,253 | 99.51 | 4.25 | 16.96 | 93.18 |
| NAG14685 | Kyoto Univ. | Kansai | 366,058,528 | 364,459,381 | 54,908,779,200 | 54,762,142,353 | 99.56 | 6.05 | 18.25 | 94.47 |
| NAG14686 | Kyoto Univ. | Kansai | 344,262,845 | 341,603,005 | 51,639,426,750 | 51,305,655,436 | 99.23 | 5.31 | 17.10 | 92.33 |
| NAG14687 | Kyoto Univ. | Kansai | 398,647,439 | 397,361,148 | 59,797,115,850 | 59,635,962,394 | 99.68 | 7.66 | 19.88 | 96.76 |
| NAG14703 | Kyoto Univ. | Kansai | 379,947,260 | 378,801,996 | 56,992,089,000 | 56,907,035,615 | 99.70 | 6.37 | 18.97 | 96.11 |
| NAG14708 | Kyoto Univ. | Kansai | 382,828,486 | 381,206,762 | 57,424,272,900 | 57,252,823,742 | 99.58 | 5.30 | 19.08 | 95.86 |
| NAG14709 | Kyoto Univ. | Kansai | 370,071,011 | 367,490,510 | 55,510,651,650 | 55,155,541,832 | 99.30 | 6.39 | 18.39 | 94.60 |
| NAG14741 | Kyoto Univ. | Kansai | 408,426,543 | 405,789,674 | 61,263,981,450 | 60,986,172,316 | 99.35 | 7.95 | 20.33 | 96.55 |
| NAG14742 | Kyoto Univ. | Kansai | 343,822,079 | 341,929,600 | 51,573,311,850 | 51,392,174,303 | 99.45 | 5.32 | 17.13 | 89.91 |
| NAG14754 | Kyoto Univ. | Kansai | 385,167,930 | 382,989,444 | 57,775,189,500 | 57,531,677,429 | 99.43 | 4.28 | 19.18 | 96.81 |
| NAG14761 | Kyoto Univ. | Kansai | 269,205,472 | 267,920,225 | 40,380,820,800 | 40,248,474,555 | 99.52 | 5.90 | 13.42 | 75.90 |
| NAG14769 | Kyoto Univ. | Kansai | 367,259,377 | 365,773,452 | 55,088,906,550 | 54,832,813,834 | 99.60 | 3.97 | 18.28 | 94.96 |
| NAG14793 | Kyoto Univ. | Kansai | 400,117,633 | 397,681,515 | 60,017,644,950 | 59,741,054,794 | 99.39 | 6.52 | 19.91 | 97.07 |
| NAG14802 | Kyoto Univ. | Kansai | 381,222,130 | 379,477,301 | 57,183,319,500 | 56,976,982,595 | 99.54 | 4.84 | 18.99 | 93.54 |
| NAG14803 | Kyoto Univ. | Kansai | 362,041,606 | 359,509,612 | 54,306,240,900 | 53,990,400,617 | 99.30 | 4.63 | 18.00 | 94.56 |
| NAG14805 | Kyoto Univ. | Kansai | 463,424,830 | 460,002,210 | 69,513,724,500 | 69,079,056,732 | 99.26 | 5.07 | 23.03 | 96.71 |
| NAG14814 | Kyoto Univ. | Kansai | 351,059,246 | 349,477,256 | 52,658,886,900 | 52,484,641,301 | 99.55 | 5.04 | 17.49 | 90.79 |
| NAG14817 | Kyoto Univ. | Kansai | 340,652,329 | 338,549,580 | 51,097,849,350 | 50,874,788,881 | 99.38 | 3.81 | 16.96 | 93.35 |
| NAG14822 | Kyoto Univ. | Kansai | 427,147,870 | 425,431,793 | 64,072,180,500 | 63,944,273,892 | 99.60 | 8.36 | 21.31 | 97.62 |
| NAG14823 | Kyoto Univ. | Kansai | 349,714,370 | 347,585,043 | 52,457,155,500 | 52,193,492,614 | 99.39 | 5.32 | 17.40 | 93.35 |
| NAG14826 | Kyoto Univ. | Kansai | 480,196,324 | 479,033,504 | 72,029,448,600 | 71,891,044,209 | 99.76 | 11.27 | 23.96 | 98.61 |
| NAG14827 | Kyoto Univ. | Kansai | 362,519,786 | 359,756,420 | 54,377,967,900 | 54,026,374,585 | 99.24 | 6.15 | 18.01 | 90.99 |
| NAG14834 | Kyoto Univ. | Kansai | 388,051,977 | 385,896,789 | 58,207,796,550 | 57,956,895,052 | 99.44 | 6.36 | 19.32 | 96.27 |
| NAG14841 | Kyoto Univ. | Kansai | 350,371,745 | 348,489,686 | 52,555,761,750 | 52,370,439,202 | 99.46 | 4.05 | 17.46 | 94.37 |
| NAG14845 | Kyoto Univ. | Kansai | 346,581,780 | 345,191,915 | 51,987,267,000 | 51,859,848,853 | 99.60 | 5.92 | 17.29 | 89.88 |
| NAG14850 | Kyoto Univ. | Kansai | 411,726,952 | 410,504,607 | 61,759,042,800 | 61,640,117,181 | 99.70 | 7.12 | 20.55 | 97.39 |
| NAG14853 | Kyoto Univ. | Kansai | 324,003,751 | 322,855,970 | 48,600,562,650 | 48,523,581,980 | 99.65 | 5.79 | 16.17 | 90.38 |
| NAG14868 | Kyoto Univ. | Kansai | 347,369,883 | 344,642,473 | 52,105,482,450 | 51,740,494,402 | 99.21 | 5.40 | 17.25 | 92.20 |
| NAG14872 | Kyoto Univ. | Kansai | 317,116,954 | 316,019,980 | 47,567,543,100 | 47,466,488,351 | 99.65 | 5.62 | 15.82 | 88.80 |
| NAG14873 | Kyoto Univ. | Kansai | 372,995,819 | 371,051,725 | 55,949,372,850 | 55,763,037,588 | 99.48 | 5.83 | 18.59 | 92.70 |
| NAG14881 | Kyoto Univ. | Kansai | 360,684,975 | 358,914,821 | 54,102,746,250 | 53,770,048,683 | 99.51 | 6.19 | 17.92 | 88.46 |
| NAG14885 | Kyoto Univ. | Kansai | 350,561,788 | 348,316,441 | 52,584,268,200 | 52,322,307,671 | 99.36 | 4.50 | 17.44 | 93.79 |
| NAG14890 | Kyoto Univ. | Kansai | 365,984,198 | 363,086,937 | 54,897,629,700 | 54,509,628,913 | 99.21 | 5.75 | 18.17 | 94.37 |
| NAG14895 | Kyoto Univ. | Kansai | 373,703,432 | 372,571,200 | 56,055,514,800 | 55,885,301,416 | 99.70 | 6.10 | 18.63 | 95.02 |
| NAG14896 | Kyoto Univ. | Kansai | 383,290,545 | 382,135,709 | 57,493,581,750 | 57,380,309,474 | 99.70 | 6.27 | 19.13 | 96.29 |
| NAG14899 | Kyoto Univ. | Kansai | 334,688,237 | 331,843,323 | 50,203,235,550 | 49,842,205,854 | 99.15 | 5.16 | 16.61 | 87.76 |
| NAG14900 | Kyoto Univ. | Kansai | 341,368,647 | 337,896,915 | 51,205,297,050 | 50,767,463,161 | 98.98 | 4.93 | 16.92 | 91.84 |
| NAG14901 | Kyoto Univ. | Kansai | 362,779,767 | 359,817,146 | 54,416,965,050 | 54,041,479,015 | 99.18 | 5.76 | 18.01 | 91.02 |
| NAG14903 | Kyoto Univ. | Kansai | 366,626,360 | 363,922,683 | 54,993,954,000 | 54,633,162,522 | 99.26 | 6.54 | 18.21 | 93.96 |
| NAG14913 | Kyoto Univ. | Kansai | 338,444,653 | 335,822,469 | 50,766,697,950 | 50,436,616,739 | 99.23 | 5.19 | 16.81 | 88.31 |
| NAG14927 | Kyoto Univ. | Kansai | 384,372,091 | 382,575,357 | 57,655,813,650 | 57,431,282,748 | 99.53 | 6.50 | 19.14 | 96.19 |
| NAG14935 | Kyoto Univ. | Kansai | 447,136,185 | 444,841,857 | 67,070,427,750 | 66,812,964,817 | 99.49 | 4.80 | 22.27 | 98.63 |
| NAG14943 | Kyoto Univ. | Kansai | 357,054,695 | 354,220,368 | 53,558,204,250 | 53,181,360,122 | 99.21 | 5.33 | 17.73 | 90.30 |
| NAG14944 | Kyoto Univ. | Kansai | 326,327,771 | 324,117,851 | 48,949,165,650 | 48,725,590,340 | 99.32 | 4.11 | 16.24 | 91.21 |
| NAG14951 | Kyoto Univ. | Kansai | 351,783,562 | 349,588,004 | 52,767,534,300 | 52,509,690,530 | 99.38 | 4.50 | 17.50 | 91.01 |
| NAG14958 | Kyoto Univ. | Kansai | 346,736,297 | 344,448,327 | 52,010,444,550 | 51,717,449,963 | 99.34 | 4.77 | 17.24 | 89.53 |
| NAG14971 | Kyoto Univ. | Kansai | 327,468,974 | 325,277,222 | 49,120,346,100 | 48,821,935,806 | 99.33 | 6.41 | 16.27 | 89.05 |
| NAG14975 | Kyoto Univ. | Kansai | 377,591,000 | 375,096,862 | 56,638,650,000 | 56,384,207,626 | 99.34 | 5.65 | 18.79 | 96.04 |
| NAG14985 | Kyoto Univ. | Kansai | 354,049,054 | 351,895,238 | 53,107,358,100 | 52,863,289,184 | 99.39 | 6.93 | 17.62 | 93.41 |
| NAG14986 | Kyoto Univ. | Kansai | 328,972,186 | 326,347,004 | 49,345,827,900 | 49,008,698,248 | 99.20 | 5.21 | 16.34 | 86.30 |
| NAG15001 | Kyoto Univ. | Kansai | 356,184,169 | 354,695,046 | 53,427,625,350 | 53,257,812,313 | 99.58 | 3.94 | 17.75 | 94.80 |
| NAG15006 | Kyoto Univ. | Kansai | 316,964,956 | 315,718,827 | 47,544,743,400 | 47,438,453,780 | 99.61 | 4.78 | 15.81 | 89.51 |
| NAG15007 | Kyoto Univ. | Kansai | 343,401,945 | 339,473,088 | 51,510,291,750 | 50,993,988,572 | 98.86 | 4.71 | 17.00 | 89.21 |
| NAG15016 | Kyoto Univ. | Kansai | 376,401,686 | 374,782,147 | 56,460,252,900 | 56,330,059,986 | 99.57 | 7.68 | 18.78 | 94.91 |

| | | | | | | | | | | |
|---|---|---|---|---|---|---|---|---|---|---|
| NAG15018 | Kyoto Univ. | Kansai | 332,734,202 | 329,473,694 | 49,910,130,300 | 49,490,094,973 | 99.02 | 4.39 | 16.50 | 90.76 |
| NAG15020 | Kyoto Univ. | Kansai | 319,094,937 | 316,253,875 | 47,864,240,550 | 47,429,180,806 | 99.11 | 2.80 | 15.81 | 88.65 |
| NAG15023 | Kyoto Univ. | Kansai | 389,950,827 | 387,599,345 | 58,492,624,050 | 58,215,738,456 | 99.40 | 5.93 | 19.41 | 96.37 |
| NAG15050 | Kyoto Univ. | Kansai | 339,584,830 | 336,427,890 | 50,937,724,500 | 50,515,208,432 | 99.07 | 4.59 | 16.84 | 91.42 |
| NAG15059 | Kyoto Univ. | Kansai | 357,163,645 | 354,275,587 | 53,574,546,750 | 53,203,890,142 | 99.19 | 7.71 | 17.73 | 92.89 |
| NAG15062 | Kyoto Univ. | Kansai | 336,950,662 | 335,068,504 | 50,542,599,300 | 50,269,879,174 | 99.44 | 6.67 | 16.76 | 90.22 |
| NAG15064 | Kyoto Univ. | Kansai | 379,476,807 | 377,023,688 | 56,921,521,050 | 56,562,136,215 | 99.35 | 7.86 | 18.85 | 94.71 |
| NAG15067 | Kyoto Univ. | Kansai | 350,449,722 | 348,480,530 | 52,567,458,300 | 52,309,989,806 | 99.44 | 7.51 | 17.44 | 90.61 |
| NAG15068 | Kyoto Univ. | Kansai | 387,621,836 | 384,583,648 | 58,143,275,400 | 57,684,844,703 | 99.22 | 5.16 | 19.23 | 95.79 |
| NAG15077 | Kyoto Univ. | Kansai | 385,973,396 | 383,835,649 | 57,896,009,400 | 57,630,075,987 | 99.45 | 5.63 | 19.21 | 96.27 |
| NAG15078 | Kyoto Univ. | Kansai | 353,589,151 | 351,152,253 | 53,038,372,650 | 52,701,598,935 | 99.31 | 4.81 | 17.57 | 93.24 |
| NAG15089 | Kyoto Univ. | Kansai | 367,399,501 | 364,152,820 | 55,109,925,150 | 54,740,743,452 | 99.12 | 5.20 | 18.25 | 92.05 |
| NAG15094 | Kyoto Univ. | Kansai | 389,615,591 | 388,341,962 | 58,442,338,650 | 58,249,466,322 | 99.67 | 7.51 | 19.42 | 92.41 |
| NAG15095 | Kyoto Univ. | Kansai | 352,132,853 | 351,028,429 | 52,819,927,950 | 52,755,383,011 | 99.69 | 6.02 | 17.59 | 93.91 |
| NAG15118 | Kyoto Univ. | Kansai | 345,879,335 | 343,931,798 | 51,881,900,250 | 51,707,102,076 | 99.44 | 5.84 | 17.24 | 93.56 |
| NAG15134 | Kyoto Univ. | Kansai | 375,389,082 | 373,214,761 | 56,308,362,300 | 56,053,741,886 | 99.42 | 6.50 | 18.68 | 95.41 |
| NAG15135 | Kyoto Univ. | Kansai | 344,198,100 | 342,415,727 | 51,629,715,000 | 51,424,954,066 | 99.48 | 6.70 | 17.14 | 92.17 |
| NAG15139 | Kyoto Univ. | Kansai | 389,180,562 | 387,564,144 | 58,377,084,300 | 58,142,226,129 | 99.58 | 4.88 | 19.38 | 96.29 |
| NAG15140 | Kyoto Univ. | Kansai | 395,040,903 | 393,874,240 | 59,256,135,450 | 59,130,979,448 | 99.70 | 6.22 | 19.71 | 96.85 |
| NAG15144 | Kyoto Univ. | Kansai | 328,961,893 | 326,474,940 | 49,344,283,950 | 49,010,971,685 | 99.24 | 5.06 | 16.34 | 90.19 |
| NAG15145 | Kyoto Univ. | Kansai | 365,371,051 | 364,282,391 | 54,805,657,650 | 54,689,499,035 | 99.70 | 6.13 | 18.23 | 94.60 |
| NAG15147 | Kyoto Univ. | Kansai | 312,896,125 | 310,635,373 | 46,934,418,750 | 46,660,219,996 | 99.28 | 4.78 | 15.55 | 84.81 |
| NAG15155 | Kyoto Univ. | Kansai | 376,463,001 | 374,995,390 | 56,469,450,150 | 56,347,788,276 | 99.61 | 7.87 | 18.78 | 92.34 |
| NAG15166 | Kyoto Univ. | Kansai | 347,038,690 | 344,370,427 | 52,055,803,500 | 51,775,109,066 | 99.23 | 5.10 | 17.26 | 93.29 |
| NAG15170 | Kyoto Univ. | Kansai | 353,915,066 | 351,905,759 | 53,087,259,900 | 52,868,934,766 | 99.43 | 5.66 | 17.62 | 91.00 |
| NAG15179 | Kyoto Univ. | Kansai | 329,879,976 | 328,310,438 | 49,481,996,400 | 49,331,816,511 | 99.52 | 7.58 | 16.44 | 88.54 |
| NAG15190 | Kyoto Univ. | Kansai | 359,803,751 | 357,672,506 | 53,970,562,650 | 53,752,182,092 | 99.41 | 7.25 | 17.92 | 93.10 |
| NAG15193 | Kyoto Univ. | Kansai | 371,972,657 | 370,146,369 | 55,795,898,550 | 55,614,044,296 | 99.51 | 8.20 | 18.54 | 94.14 |
| NAG15210 | Kyoto Univ. | Kansai | 337,947,540 | 335,079,479 | 50,692,131,000 | 50,352,840,123 | 99.15 | 4.97 | 16.78 | 91.67 |
| NAG15222 | Kyoto Univ. | Kansai | 411,568,535 | 409,912,822 | 61,735,280,250 | 61,613,300,156 | 99.60 | 9.03 | 20.54 | 96.48 |
| NAG15229 | Kyoto Univ. | Kansai | 422,742,253 | 418,968,418 | 63,411,337,950 | 62,941,847,347 | 99.11 | 4.43 | 20.98 | 95.55 |
| NAG15233 | Kyoto Univ. | Kansai | 366,552,469 | 364,513,882 | 54,982,870,350 | 54,686,568,410 | 99.44 | 6.80 | 18.23 | 93.65 |
| NAG15244 | Kyoto Univ. | Kansai | 409,345,421 | 407,826,484 | 61,401,813,150 | 61,245,817,414 | 99.63 | 8.69 | 20.42 | 96.85 |
| NAG15248 | Kyoto Univ. | Kansai | 343,052,322 | 339,653,435 | 51,457,848,300 | 50,969,507,499 | 99.01 | 6.25 | 16.99 | 91.55 |
| NAG15251 | Kyoto Univ. | Kansai | 348,267,922 | 346,176,738 | 52,240,188,300 | 51,995,777,407 | 99.40 | 5.33 | 17.33 | 89.44 |
| NAG15259 | Kyoto Univ. | Kansai | 387,610,774 | 385,477,235 | 58,141,616,100 | 57,957,283,829 | 99.45 | 5.96 | 19.32 | 96.70 |
| NAG15265 | Kyoto Univ. | Kansai | 342,681,298 | 340,601,816 | 51,402,194,700 | 51,089,664,201 | 99.39 | 6.78 | 17.03 | 87.52 |
| NAG15266 | Kyoto Univ. | Kansai | 385,881,951 | 384,023,935 | 57,882,292,650 | 57,706,779,331 | 99.52 | 7.69 | 19.24 | 96.20 |
| NAG15274 | Kyoto Univ. | Kansai | 380,538,586 | 378,748,565 | 57,080,787,900 | 56,903,638,777 | 99.53 | 6.65 | 18.97 | 95.95 |
| NAG15295 | Kyoto Univ. | Kansai | 380,873,777 | 379,153,645 | 57,131,066,550 | 56,943,604,641 | 99.55 | 6.52 | 18.98 | 93.14 |
| NAG15313 | Kyoto Univ. | Kansai | 343,539,820 | 341,837,753 | 51,530,973,000 | 51,360,792,194 | 99.50 | 6.13 | 17.12 | 89.35 |
| NAG15320 | Kyoto Univ. | Kansai | 353,354,841 | 351,495,596 | 53,003,226,150 | 52,792,417,217 | 99.47 | 7.76 | 17.60 | 92.01 |
| NAG15323 | Kyoto Univ. | Kansai | 349,205,587 | 347,235,282 | 52,380,838,050 | 52,138,414,802 | 99.44 | 5.35 | 17.38 | 89.74 |
| NAG15325 | Kyoto Univ. | Kansai | 335,923,845 | 333,854,205 | 50,388,576,750 | 50,124,928,391 | 99.38 | 7.31 | 16.71 | 87.35 |
| NAG15326 | Kyoto Univ. | Kansai | 373,496,771 | 371,919,840 | 56,024,515,650 | 55,874,937,307 | 99.58 | 7.90 | 18.62 | 94.48 |
| NAG15353 | Kyoto Univ. | Kansai | 310,269,161 | 309,257,859 | 46,540,374,150 | 46,444,278,344 | 99.67 | 4.87 | 15.48 | 87.87 |
| NAG15360 | Kyoto Univ. | Kansai | 366,253,140 | 364,593,173 | 54,937,971,000 | 54,767,107,015 | 99.55 | 5.44 | 18.26 | 91.82 |
| NAG15363 | Kyoto Univ. | Kansai | 363,561,923 | 361,306,299 | 54,534,288,450 | 54,233,205,642 | 99.38 | 6.46 | 18.08 | 94.20 |
| NAG15369 | Kyoto Univ. | Kansai | 316,503,764 | 314,198,658 | 47,475,564,600 | 47,217,652,104 | 99.27 | 4.83 | 15.74 | 88.51 |
| NAG15375 | Kyoto Univ. | Kansai | 366,135,828 | 364,634,160 | 54,920,374,200 | 54,808,757,933 | 99.59 | 7.82 | 18.27 | 93.81 |
| NAG15393 | Kyoto Univ. | Kansai | 329,241,288 | 326,839,879 | 49,386,193,200 | 49,045,106,031 | 99.27 | 5.19 | 16.35 | 85.64 |
| NAG15396 | Kyoto Univ. | Kansai | 355,749,077 | 353,696,360 | 53,362,361,550 | 53,140,184,223 | 99.42 | 5.92 | 17.71 | 90.59 |
| NAG15400 | Kyoto Univ. | Kansai | 389,134,507 | 387,139,034 | 58,370,176,050 | 58,106,628,681 | 99.49 | 5.34 | 19.37 | 93.40 |
| NAG15414 | Kyoto Univ. | Kansai | 324,231,490 | 323,121,048 | 48,634,723,500 | 48,557,069,984 | 99.66 | 5.16 | 16.19 | 90.74 |
| NAG15425 | Kyoto Univ. | Kansai | 369,608,929 | 368,199,457 | 55,441,339,350 | 55,347,887,950 | 99.62 | 5.22 | 18.45 | 95.87 |
| NAG15430 | Kyoto Univ. | Kansai | 339,680,346 | 338,566,095 | 50,952,051,900 | 50,874,900,673 | 99.67 | 5.92 | 16.96 | 92.76 |
| NAG15436 | Kyoto Univ. | Kansai | 330,324,764 | 328,226,936 | 49,548,714,600 | 49,291,703,742 | 99.36 | 4.71 | 16.43 | 86.96 |
| NAG15440 | Kyoto Univ. | Kansai | 336,723,291 | 335,519,866 | 50,508,493,650 | 50,398,598,271 | 99.64 | 6.42 | 16.80 | 88.69 |
| NAG15445 | Kyoto Univ. | Kansai | 338,602,252 | 337,515,229 | 50,790,337,800 | 50,684,315,658 | 99.68 | 7.06 | 16.89 | 91.66 |
| NAG15446 | Kyoto Univ. | Kansai | 372,102,035 | 370,329,716 | 55,815,305,250 | 55,664,026,601 | 99.52 | 4.50 | 18.55 | 96.26 |
| NAG15460 | Kyoto Univ. | Kansai | 365,247,340 | 363,202,971 | 54,787,101,000 | 54,504,101,829 | 99.44 | 4.83 | 18.17 | 94.60 |
| NAG15467 | Kyoto Univ. | Kansai | 416,477,158 | 413,968,464 | 62,471,573,700 | 62,161,923,366 | 99.40 | 7.41 | 20.72 | 94.77 |
| NAG15501 | Kyoto Univ. | Kansai | 346,924,936 | 345,743,774 | 52,038,740,400 | 51,936,739,713 | 99.66 | 5.98 | 17.31 | 90.05 |
| NAG15504 | Kyoto Univ. | Kansai | 319,049,490 | 317,105,875 | 47,857,423,500 | 47,647,915,611 | 99.39 | 5.26 | 15.88 | 88.78 |
| NAG15506 | Kyoto Univ. | Kansai | 357,858,379 | 356,454,657 | 53,678,756,850 | 53,528,402,398 | 99.61 | 6.22 | 17.84 | 91.20 |
| NAG15515 | Kyoto Univ. | Kansai | 365,921,957 | 364,575,199 | 54,888,293,550 | 54,720,040,241 | 99.63 | 6.35 | 18.24 | 91.55 |
| NAG15537 | Kyoto Univ. | Kansai | 388,516,478 | 386,154,075 | 58,277,471,700 | 57,991,877,621 | 99.39 | 7.29 | 19.33 | 95.94 |
| NAG15571 | Kyoto Univ. | Kansai | 393,849,549 | 391,678,103 | 59,077,432,350 | 58,797,878,985 | 99.45 | 6.56 | 19.60 | 96.26 |
| NAG15576 | Kyoto Univ. | Kansai | 354,824,314 | 352,570,461 | 53,223,647,100 | 52,895,207,895 | 99.36 | 5.76 | 17.63 | 93.71 |
| NAG15584 | Kyoto Univ. | Kansai | 344,670,833 | 343,503,587 | 51,700,624,950 | 51,551,332,208 | 99.66 | 6.63 | 17.18 | 91.61 |
| NAG15589 | Kyoto Univ. | Kansai | 388,024,491 | 385,705,478 | 58,203,673,650 | 57,977,261,279 | 99.40 | 5.63 | 19.33 | 96.66 |
| NAG15599 | Kyoto Univ. | Kansai | 329,913,129 | 328,777,847 | 49,486,969,350 | 49,376,866,804 | 99.66 | 6.62 | 16.46 | 90.45 |
| NAG15611 | Kyoto Univ. | Kansai | 330,700,911 | 329,433,733 | 49,605,136,650 | 49,501,984,201 | 99.62 | 5.13 | 16.50 | 91.41 |
| NAG15614 | Kyoto Univ. | Kansai | 392,872,461 | 390,618,921 | 58,930,869,150 | 58,653,154,815 | 99.43 | 6.32 | 19.55 | 96.36 |
| NAG15617 | Kyoto Univ. | Kansai | 325,555,911 | 322,379,684 | 48,833,386,650 | 48,376,712,078 | 99.02 | 6.49 | 16.13 | 88.42 |
| NAG15629 | Kyoto Univ. | Kansai | 414,473,320 | 413,053,464 | 62,170,998,000 | 62,065,482,050 | 99.66 | 6.77 | 20.69 | 97.76 |
| NAG15645 | Kyoto Univ. | Kansai | 365,574,218 | 363,250,975 | 54,836,132,700 | 54,550,033,976 | 99.36 | 5.89 | 18.18 | 91.13 |
| NAG15646 | Kyoto Univ. | Kansai | 302,696,055 | 300,446,526 | 45,404,408,250 | 45,089,540,787 | 99.26 | 6.19 | 15.03 | 83.16 |
| NAG15650 | Kyoto Univ. | Kansai | 386,369,869 | 384,004,404 | 57,955,480,350 | 57,590,186,068 | 99.39 | 5.55 | 19.20 | 92.88 |
| NAG15655 | Kyoto Univ. | Kansai | 337,669,067 | 335,402,951 | 50,650,360,050 | 50,415,549,124 | 99.33 | 6.22 | 16.81 | 88.37 |
| NAG15663 | Kyoto Univ. | Kansai | 343,414,581 | 341,558,851 | 51,512,187,150 | 51,339,254,161 | 99.46 | 7.19 | 17.11 | 90.84 |
| NAG15665 | Kyoto Univ. | Kansai | 382,946,593 | 381,524,515 | 57,441,988,950 | 57,327,430,615 | 99.63 | 6.02 | 19.11 | 96.60 |
| NAG15685 | Kyoto Univ. | Kansai | 371,712,277 | 369,600,246 | 55,756,841,550 | 55,532,192,253 | 99.43 | 6.35 | 18.51 | 95.17 |
| NAG15688 | Kyoto Univ. | Kansai | 375,428,557 | 371,518,454 | 56,314,283,550 | 55,737,753,094 | 98.96 | 5.86 | 18.58 | 91.67 |
| NAG15694 | Kyoto Univ. | Kansai | 477,554,393 | 475,923,510 | 71,633,158,950 | 71,525,763,450 | 99.66 | 6.47 | 23.84 | 99.07 |
| NAG15705 | Kyoto Univ. | Kansai | 382,919,970 | 378,990,484 | 57,437,995,500 | 56,906,468,471 | 98.97 | 6.53 | 18.97 | 95.80 |
| NAG15710 | Kyoto Univ. | Kansai | 347,643,764 | 345,475,238 | 52,146,564,600 | 51,824,001,983 | 99.38 | 4.71 | 17.27 | 92.32 |
| NAG15719 | Kyoto Univ. | Kansai | 336,416,579 | 334,227,245 | 50,462,486,850 | 50,183,854,042 | 99.35 | 6.19 | 16.73 | 87.50 |
| NAG15723 | Kyoto Univ. | Kansai | 350,368,155 | 348,752,827 | 52,555,223,250 | 52,383,067,832 | 99.54 | 5.15 | 17.46 | 90.63 |
| NAG15736 | Kyoto Univ. | Kansai | 382,910,374 | 381,285,207 | 57,436,556,100 | 57,265,681,328 | 99.58 | 8.52 | 19.09 | 95.57 |
| NAG15737 | Kyoto Univ. | Kansai | 354,118,003 | 352,685,047 | 53,117,700,450 | 53,000,542,882 | 99.60 | 5.63 | 17.67 | 94.08 |
| NAG15763 | Kyoto Univ. | Kansai | 368,063,239 | 365,830,406 | 55,209,485,850 | 54,997,199,683 | 99.39 | 6.34 | 18.33 | 95.06 |
| NAG15770 | Kyoto Univ. | Kansai | 404,851,434 | 402,827,162 | 60,727,715,100 | 60,474,553,897 | 99.50 | 4.99 | 20.16 | 94.61 |
| NAG15785 | Kyoto Univ. | Kansai | 353,583,698 | 350,633,675 | 53,037,554,700 | 52,626,099,835 | 99.17 | 6.85 | 17.54 | 91.62 |
| NAG15799 | Kyoto Univ. | Kansai | 306,530,154 | 305,181,928 | 45,979,523,100 | 45,862,973,697 | 99.56 | 5.36 | 15.29 | 87.16 |
| NAG15803 | Kyoto Univ. | Kansai | 426,605,812 | 422,419,874 | 63,990,871,800 | 63,371,840,976 | 99.02 | 5.12 | 21.12 | 97.72 |
| NAG15811 | Kyoto Univ. | Kansai | 342,659,328 | 340,583,125 | 51,398,899,200 | 51,148,487,470 | 99.39 | 6.06 | 17.05 | 88.46 |
| NAG15818 | Kyoto Univ. | Kansai | 355,419,523 | 353,311,888 | 53,312,928,450 | 53,031,458,067 | 99.41 | 5.63 | 17.68 | 93.91 |
| NAG15823 | Kyoto Univ. | Kansai | 399,888,561 | 398,272,550 | 59,983,284,150 | 59,822,526,419 | 99.60 | 9.58 | 19.94 | 96.47 |
| NAG15827 | Kyoto Univ. | Kansai | 334,446,855 | 332,930,581 | 50,167,028,250 | 50,033,140,755 | 99.55 | 4.86 | 16.68 | 88.96 |

| | | | | | | | | | | |
|---|---|---|---|---|---|---|---|---|---|---|
| NAG15828 | Kyoto Univ. | Kansai | 374,881,783 | 373,026,924 | 56,232,267,450 | 56,030,559,098 | 99.51 | 7.53 | 18.68 | 94.21 |
| NAG15834 | Kyoto Univ. | Kansai | 314,657,635 | 313,649,564 | 47,198,645,250 | 47,145,657,987 | 99.68 | 5.06 | 15.72 | 89.70 |
| NAG15846 | Kyoto Univ. | Kansai | 333,991,230 | 331,342,250 | 50,098,684,500 | 49,738,252,406 | 99.21 | 4.46 | 16.58 | 90.80 |
| NAG15850 | Kyoto Univ. | Kansai | 373,165,662 | 369,208,008 | 55,974,849,300 | 55,419,185,934 | 98.94 | 6.47 | 18.47 | 94.68 |
| NAG15878 | Kyoto Univ. | Kansai | 370,373,844 | 367,976,795 | 55,556,076,600 | 55,274,717,882 | 99.35 | 6.21 | 18.42 | 95.46 |
| NAG15880 | Kyoto Univ. | Kansai | 373,096,330 | 371,660,284 | 55,964,449,500 | 55,750,368,166 | 99.62 | 5.47 | 18.58 | 92.06 |
| NAG15887 | Kyoto Univ. | Kansai | 358,202,919 | 357,027,491 | 53,730,437,850 | 53,612,296,933 | 99.67 | 6.05 | 17.87 | 94.47 |
| NAG15890 | Kyoto Univ. | Kansai | 362,114,514 | 360,666,346 | 54,317,177,100 | 54,128,904,116 | 99.60 | 7.09 | 18.04 | 94.04 |
| NAG15893 | Kyoto Univ. | Kansai | 375,723,695 | 373,095,389 | 56,358,554,250 | 55,986,366,865 | 99.30 | 6.70 | 18.66 | 94.39 |
| NAG15906 | Kyoto Univ. | Kansai | 379,958,149 | 378,108,122 | 56,993,722,350 | 56,791,197,184 | 99.51 | 7.61 | 18.93 | 91.98 |
| NAG15929 | Kyoto Univ. | Kansai | 325,319,458 | 323,885,767 | 48,797,918,700 | 48,663,680,550 | 99.56 | 5.21 | 16.22 | 90.21 |
| NAG15957 | Kyoto Univ. | Kansai | 356,447,234 | 354,801,008 | 53,467,085,100 | 53,319,332,292 | 99.54 | 7.33 | 17.77 | 92.66 |
| NAG15960 | Kyoto Univ. | Kansai | 359,297,613 | 357,404,125 | 53,894,641,950 | 53,628,916,487 | 99.47 | 6.31 | 17.88 | 90.69 |
| NAG15969 | Kyoto Univ. | Kansai | 358,804,546 | 356,323,404 | 53,820,681,900 | 53,501,328,561 | 99.31 | 6.42 | 17.83 | 90.41 |
| NAG15973 | Kyoto Univ. | Kansai | 332,984,418 | 331,997,024 | 49,947,662,700 | 49,838,089,235 | 99.70 | 5.63 | 16.61 | 90.69 |
| NAG15977 | Kyoto Univ. | Kansai | 349,839,610 | 347,675,305 | 52,475,941,500 | 52,248,254,223 | 99.38 | 6.28 | 17.42 | 93.06 |
| NAG15978 | Kyoto Univ. | Kansai | 344,495,709 | 342,004,360 | 51,674,356,350 | 51,352,120,091 | 99.28 | 5.40 | 17.12 | 89.53 |
| NAG15982 | Kyoto Univ. | Kansai | 365,287,530 | 363,806,778 | 54,793,129,500 | 54,653,192,741 | 99.59 | 5.89 | 18.22 | 94.86 |
| NAG15983 | Kyoto Univ. | Kansai | 356,805,411 | 352,386,452 | 53,520,811,650 | 52,894,830,434 | 98.76 | 5.45 | 17.63 | 93.21 |
| NAG15985 | Kyoto Univ. | Kansai | 398,676,764 | 396,227,765 | 59,801,514,600 | 59,524,808,128 | 99.39 | 6.98 | 19.84 | 96.76 |
| NAG15991 | Kyoto Univ. | Kansai | 325,870,812 | 322,936,371 | 48,880,621,800 | 48,496,931,453 | 99.10 | 5.79 | 16.17 | 88.98 |
| NAG15994 | Kyoto Univ. | Kansai | 409,474,381 | 407,606,439 | 61,421,157,150 | 61,263,877,702 | 99.54 | 4.64 | 20.42 | 97.94 |
| NAG16000 | Kyoto Univ. | Kansai | 340,110,627 | 338,024,360 | 51,016,594,050 | 50,735,002,118 | 99.39 | 4.61 | 16.91 | 89.03 |
| NAG16004 | Kyoto Univ. | Kansai | 361,529,485 | 360,354,102 | 54,229,422,750 | 54,111,552,142 | 99.67 | 5.38 | 18.04 | 94.80 |
| NAG16025 | Kyoto Univ. | Kansai | 361,716,966 | 360,325,222 | 54,257,544,900 | 54,130,193,699 | 99.62 | 5.49 | 18.04 | 94.97 |
| NAG16040 | Kyoto Univ. | Kansai | 354,190,799 | 353,071,584 | 53,128,619,850 | 53,058,378,086 | 99.68 | 4.65 | 17.69 | 94.76 |
| NAG16051 | Kyoto Univ. | Kansai | 326,077,815 | 324,993,479 | 48,911,672,250 | 48,799,255,500 | 99.67 | 4.94 | 16.27 | 87.42 |
| NAG16053 | Kyoto Univ. | Kansai | 348,899,042 | 347,326,341 | 52,334,856,300 | 52,187,815,431 | 99.55 | 5.31 | 17.40 | 93.66 |
| NAG16055 | Kyoto Univ. | Kansai | 341,687,054 | 339,409,481 | 51,253,058,100 | 50,978,934,754 | 99.33 | 6.57 | 16.99 | 91.49 |
| NAG16060 | Kyoto Univ. | Kansai | 361,862,132 | 360,044,333 | 54,279,319,800 | 54,107,587,883 | 99.50 | 5.96 | 18.04 | 91.44 |
| NAG16064 | Kyoto Univ. | Kansai | 383,159,783 | 381,715,660 | 57,473,967,450 | 57,368,495,337 | 99.62 | 4.61 | 19.12 | 96.84 |
| NAG16073 | Kyoto Univ. | Kansai | 441,124,852 | 439,700,634 | 66,168,727,800 | 66,043,822,333 | 99.68 | 5.91 | 22.01 | 96.18 |
| NAG16085 | Kyoto Univ. | Kansai | 351,360,974 | 350,052,550 | 52,704,146,100 | 52,598,278,036 | 99.63 | 4.98 | 17.53 | 94.28 |
| NAG16089 | Kyoto Univ. | Kansai | 310,516,918 | 308,535,230 | 46,577,537,700 | 46,318,212,550 | 99.36 | 5.17 | 15.44 | 83.31 |
| NAG16106 | Kyoto Univ. | Kansai | 314,127,734 | 311,393,559 | 47,119,160,100 | 46,699,584,806 | 99.13 | 3.32 | 15.57 | 87.39 |
| NAG16113 | Kyoto Univ. | Kansai | 372,395,493 | 370,244,288 | 55,859,323,950 | 55,613,637,038 | 99.42 | 6.42 | 18.54 | 91.98 |
| NAG16119 | Kyoto Univ. | Kansai | 355,205,448 | 353,788,218 | 53,280,817,200 | 53,121,534,337 | 99.60 | 5.67 | 17.71 | 90.55 |
| NAG16120 | Kyoto Univ. | Kansai | 386,996,743 | 383,639,210 | 58,049,511,450 | 57,531,972,346 | 99.13 | 6.48 | 19.18 | 92.57 |
| NAG16134 | Kyoto Univ. | Kansai | 393,006,162 | 390,523,676 | 58,950,924,300 | 58,695,519,961 | 99.37 | 6.75 | 19.57 | 93.72 |
| NAG16151 | Kyoto Univ. | Kansai | 332,780,757 | 330,597,951 | 49,917,113,550 | 49,641,230,652 | 99.34 | 5.98 | 16.55 | 90.53 |
| NAG16154 | Kyoto Univ. | Kansai | 341,787,774 | 339,692,664 | 51,268,166,100 | 51,049,302,848 | 99.39 | 6.17 | 17.02 | 92.05 |
| NAG16160 | Kyoto Univ. | Kansai | 343,349,597 | 340,963,877 | 51,502,439,550 | 51,260,191,004 | 99.31 | 5.65 | 17.09 | 92.76 |
| NAG16174 | Kyoto Univ. | Kansai | 338,489,438 | 336,279,268 | 50,773,415,700 | 50,532,891,707 | 99.35 | 6.18 | 16.84 | 91.81 |
| NAG16176 | Kyoto Univ. | Kansai | 338,296,485 | 337,092,378 | 50,744,472,750 | 50,634,848,976 | 99.64 | 4.69 | 16.88 | 92.96 |
| NAG16181 | Kyoto Univ. | Kansai | 366,916,355 | 365,497,451 | 55,037,453,250 | 54,933,509,271 | 99.61 | 5.30 | 18.31 | 95.80 |
| NAG16183 | Kyoto Univ. | Kansai | 339,396,170 | 337,580,497 | 50,909,425,500 | 50,689,594,371 | 99.47 | 6.33 | 16.90 | 91.89 |
| NAG16186 | Kyoto Univ. | Kansai | 349,313,378 | 347,411,786 | 52,397,006,700 | 52,205,754,010 | 99.46 | 5.43 | 17.40 | 93.46 |
| NAG16198 | Kyoto Univ. | Kansai | 349,896,073 | 347,551,127 | 52,484,410,950 | 52,229,286,230 | 99.33 | 6.45 | 17.41 | 92.75 |
| NAG16217 | Kyoto Univ. | Kansai | 369,632,738 | 367,391,338 | 55,444,910,700 | 55,215,069,810 | 99.39 | 6.75 | 18.41 | 94.85 |
| NAG16222 | Kyoto Univ. | Kansai | 381,057,867 | 378,680,023 | 57,158,680,050 | 56,848,112,191 | 99.38 | 5.27 | 18.95 | 93.13 |
| NAG16224 | Kyoto Univ. | Kansai | 349,359,126 | 348,229,456 | 52,403,868,900 | 52,283,415,041 | 99.68 | 5.05 | 17.43 | 93.63 |
| NAG16230 | Kyoto Univ. | Kansai | 336,153,044 | 334,891,591 | 50,422,956,600 | 50,325,539,137 | 99.62 | 4.93 | 16.78 | 92.47 |
| NAG16231 | Kyoto Univ. | Kansai | 362,894,212 | 361,087,491 | 54,434,131,800 | 54,250,746,513 | 99.50 | 8.06 | 18.08 | 89.76 |
| NAG16237 | Kyoto Univ. | Kansai | 376,775,990 | 374,975,427 | 56,516,398,500 | 56,342,470,446 | 99.52 | 6.61 | 18.78 | 91.80 |
| NAG16239 | Kyoto Univ. | Kansai | 372,196,599 | 368,439,730 | 55,829,489,850 | 55,325,703,421 | 98.99 | 6.33 | 18.44 | 94.78 |
| NAG16243 | Kyoto Univ. | Kansai | 338,532,137 | 337,242,616 | 50,779,820,550 | 50,637,635,933 | 99.62 | 4.69 | 16.88 | 89.33 |
| NAG16244 | Kyoto Univ. | Kansai | 348,645,859 | 346,894,355 | 52,296,878,850 | 52,098,730,057 | 99.50 | 4.84 | 17.37 | 93.66 |
| NAG16260 | Kyoto Univ. | Kansai | 343,855,930 | 342,003,712 | 51,578,389,500 | 51,401,466,880 | 99.46 | 6.17 | 17.13 | 92.85 |
| NAG16264 | Kyoto Univ. | Kansai | 349,273,983 | 345,465,656 | 52,391,097,450 | 51,869,230,096 | 98.91 | 5.59 | 17.29 | 92.56 |
| NAG16266 | Kyoto Univ. | Kansai | 328,129,721 | 326,076,542 | 49,219,458,150 | 49,000,115,539 | 99.37 | 5.43 | 16.33 | 90.62 |
| NAG16276 | Kyoto Univ. | Kansai | 387,948,924 | 385,548,634 | 58,192,338,600 | 57,868,816,574 | 99.38 | 5.91 | 19.29 | 94.90 |
| NAG16286 | Kyoto Univ. | Kansai | 416,122,180 | 414,019,888 | 62,418,327,000 | 62,158,541,180 | 99.49 | 6.74 | 20.72 | 96.84 |
| NAG16297 | Kyoto Univ. | Kansai | 376,478,676 | 375,056,206 | 56,471,801,400 | 56,311,447,448 | 99.62 | 6.83 | 18.77 | 94.88 |
| NAG16298 | Kyoto Univ. | Kansai | 360,297,083 | 358,311,086 | 54,044,562,450 | 53,863,378,324 | 99.45 | 5.69 | 17.95 | 94.84 |
| NAG16303 | Kyoto Univ. | Kansai | 366,370,776 | 364,324,052 | 54,955,616,400 | 54,700,912,189 | 99.44 | 6.45 | 18.23 | 94.61 |
| NAG16308 | Kyoto Univ. | Kansai | 415,500,156 | 413,672,296 | 62,325,023,400 | 62,142,860,692 | 99.56 | 7.81 | 20.71 | 96.92 |
| NAG16318 | Kyoto Univ. | Kansai | 364,344,096 | 362,351,119 | 54,651,614,400 | 54,443,642,140 | 99.45 | 6.50 | 18.15 | 94.38 |
| NAG16320 | Kyoto Univ. | Kansai | 361,029,174 | 359,665,649 | 54,154,376,100 | 54,049,776,970 | 99.62 | 5.14 | 18.02 | 94.65 |
| NAG16324 | Kyoto Univ. | Kansai | 335,368,302 | 333,666,842 | 50,305,245,300 | 50,113,802,032 | 99.49 | 6.08 | 16.70 | 90.92 |
| NAG16326 | Kyoto Univ. | Kansai | 335,641,440 | 333,691,344 | 50,346,216,000 | 50,096,847,468 | 99.42 | 6.88 | 16.70 | 90.37 |
| NAG16337 | Kyoto Univ. | Kansai | 357,289,605 | 354,004,153 | 53,593,440,750 | 53,107,061,531 | 99.08 | 5.62 | 17.70 | 91.87 |
| NAG16367 | Kyoto Univ. | Kansai | 379,598,674 | 378,066,303 | 56,939,801,100 | 56,645,192,736 | 99.60 | 5.87 | 18.88 | 93.99 |
| NAG16385 | Kyoto Univ. | Kansai | 352,676,222 | 351,060,631 | 52,901,433,300 | 52,772,656,553 | 99.54 | 4.97 | 17.59 | 94.69 |
| NAG16390 | Kyoto Univ. | Kansai | 340,261,651 | 339,070,175 | 51,039,247,650 | 50,948,352,489 | 99.65 | 4.82 | 16.98 | 93.27 |
| NAG16398 | Kyoto Univ. | Kansai | 414,495,923 | 412,787,705 | 62,174,388,450 | 62,033,114,884 | 99.59 | 7.46 | 20.68 | 97.79 |
| NAG16399 | Kyoto Univ. | Kansai | 348,956,571 | 346,580,050 | 52,343,485,650 | 52,077,809,838 | 99.32 | 6.36 | 17.36 | 93.39 |
| NAG16412 | Kyoto Univ. | Kansai | 365,284,487 | 363,437,719 | 54,792,673,050 | 54,627,595,532 | 99.49 | 4.23 | 18.21 | 95.89 |
| NAG16413 | Kyoto Univ. | Kansai | 380,111,897 | 378,016,578 | 57,016,784,550 | 56,799,047,340 | 99.45 | 5.87 | 18.93 | 96.17 |
| NAG16420 | Kyoto Univ. | Kansai | 365,342,085 | 364,064,106 | 54,801,312,750 | 54,613,142,498 | 99.65 | 6.27 | 18.20 | 94.08 |
| NAG16437 | Kyoto Univ. | Kansai | 354,277,119 | 352,404,365 | 53,141,567,850 | 52,891,511,747 | 99.47 | 5.32 | 17.63 | 94.12 |
| NAG16441 | Kyoto Univ. | Kansai | 442,784,692 | 441,219,136 | 66,417,703,800 | 66,089,967,587 | 99.65 | 6.37 | 22.03 | 94.42 |
| NAG16455 | Kyoto Univ. | Kansai | 379,091,120 | 377,793,330 | 56,863,668,000 | 56,740,029,413 | 99.66 | 5.01 | 18.91 | 93.56 |
| NAG16456 | Kyoto Univ. | Kansai | 384,516,961 | 381,068,652 | 57,677,544,150 | 57,224,071,297 | 99.10 | 5.60 | 19.07 | 95.33 |
| NAG16492 | Kyoto Univ. | Kansai | 339,713,651 | 337,999,687 | 50,957,047,650 | 50,793,668,334 | 99.50 | 5.86 | 16.93 | 89.16 |
| NAG16497 | Kyoto Univ. | Kansai | 352,205,050 | 350,250,204 | 52,830,757,500 | 52,654,981,433 | 99.44 | 5.73 | 17.55 | 90.48 |
| NAG16498 | Kyoto Univ. | Kansai | 351,653,361 | 350,334,501 | 52,748,004,150 | 52,624,239,230 | 99.62 | 5.25 | 17.54 | 94.37 |
| NAG16510 | Kyoto Univ. | Kansai | 408,161,632 | 406,362,128 | 61,224,244,800 | 61,063,890,658 | 99.56 | 8.77 | 20.35 | 96.47 |
| NAG16515 | Kyoto Univ. | Kansai | 362,327,144 | 360,102,556 | 54,349,071,600 | 53,980,404,099 | 99.39 | 6.94 | 17.99 | 92.81 |
| NAG16517 | Kyoto Univ. | Kansai | 389,366,581 | 387,698,692 | 58,404,987,150 | 58,256,146,575 | 99.57 | 9.16 | 19.42 | 95.16 |
| NAG16528 | Kyoto Univ. | Kansai | 307,962,175 | 306,036,339 | 46,194,326,250 | 45,917,974,985 | 99.37 | 5.07 | 15.31 | 86.08 |
| NAG16534 | Kyoto Univ. | Kansai | 370,258,045 | 367,645,633 | 55,538,706,750 | 55,200,407,420 | 99.29 | 6.48 | 18.40 | 94.87 |
| NAG16539 | Kyoto Univ. | Kansai | 341,862,213 | 339,073,562 | 51,279,331,950 | 50,881,779,545 | 99.18 | 5.10 | 16.96 | 91.11 |
| NAG16542 | Kyoto Univ. | Kansai | 372,161,111 | 370,707,822 | 55,824,166,650 | 55,622,266,695 | 99.61 | 6.25 | 18.54 | 94.65 |
| NAG16556 | Kyoto Univ. | Kansai | 354,956,149 | 352,347,503 | 53,243,422,350 | 52,910,649,186 | 99.27 | 5.01 | 17.64 | 93.65 |
| NAG16557 | Kyoto Univ. | Kansai | 335,225,689 | 334,017,274 | 50,283,853,350 | 50,140,204,150 | 99.64 | 5.56 | 16.71 | 90.78 |
| NAG16559 | Kyoto Univ. | Kansai | 362,593,162 | 360,148,337 | 54,388,974,300 | 54,120,033,657 | 99.33 | 5.60 | 18.04 | 91.10 |
| NAG16560 | Kyoto Univ. | Kansai | 340,936,453 | 339,071,702 | 51,140,467,950 | 50,945,473,059 | 99.45 | 5.44 | 16.98 | 92.35 |
| NAG16564 | Kyoto Univ. | Kansai | 318,295,289 | 316,022,817 | 47,744,293,350 | 47,433,077,698 | 99.29 | 5.30 | 15.81 | 87.91 |
| NAG16572 | Kyoto Univ. | Kansai | 374,707,950 | 372,412,930 | 56,206,192,500 | 55,979,193,728 | 99.39 | 6.34 | 18.66 | 95.37 |
| NAG16574 | Kyoto Univ. | Kansai | 387,088,483 | 385,557,966 | 58,063,272,450 | 57,824,303,627 | 99.60 | 8.97 | 19.27 | 94.58 |

| | | | | | | | | | | |
|---|---|---|---|---|---|---|---|---|---|---|
| NAG16581 | Kyoto Univ. | Kansai | 323,974,554 | 321,157,029 | 48,596,183,100 | 48,229,137,235 | 99.13 | 5.51 | 16.08 | 87.41 |
| NAG16622 | Kyoto Univ. | Kansai | 320,596,363 | 319,348,699 | 48,089,454,450 | 47,975,818,164 | 99.61 | 7.08 | 15.99 | 88.13 |
| NAG16627 | Kyoto Univ. | Kansai | 347,715,562 | 345,445,730 | 52,157,334,300 | 51,853,207,646 | 99.35 | 7.35 | 17.28 | 91.47 |
| NAG16629 | Kyoto Univ. | Kansai | 360,580,077 | 359,137,455 | 54,087,011,550 | 53,974,957,287 | 99.60 | 8.05 | 17.99 | 92.94 |
| NAG16643 | Kyoto Univ. | Kansai | 342,529,686 | 341,019,720 | 51,379,452,900 | 51,260,312,507 | 99.56 | 4.66 | 17.09 | 90.56 |
| NAG16644 | Kyoto Univ. | Kansai | 359,202,135 | 355,973,193 | 53,880,320,250 | 53,490,840,287 | 99.10 | 5.14 | 17.83 | 93.66 |
| NAG16645 | Kyoto Univ. | Kansai | 378,197,172 | 374,866,114 | 56,729,575,800 | 56,292,925,621 | 99.12 | 5.65 | 18.76 | 94.92 |
| NAG16661 | Kyoto Univ. | Kansai | 354,044,136 | 352,031,251 | 53,106,620,400 | 52,911,877,295 | 99.43 | 5.80 | 17.64 | 93.81 |
| NAG16678 | Kyoto Univ. | Kansai | 359,662,447 | 357,298,841 | 53,949,367,050 | 53,604,652,993 | 99.34 | 4.86 | 17.87 | 90.93 |
| NAG16689 | Kyoto Univ. | Kansai | 321,015,166 | 319,628,491 | 48,152,274,900 | 48,000,171,270 | 99.57 | 4.43 | 16.00 | 89.92 |
| NAG16692 | Kyoto Univ. | Kansai | 337,743,476 | 336,592,141 | 50,661,521,400 | 50,526,683,300 | 99.66 | 4.88 | 16.84 | 92.49 |
| NAG16714 | Kyoto Univ. | Kansai | 328,194,850 | 326,027,496 | 49,229,227,500 | 48,975,124,143 | 99.34 | 5.43 | 16.33 | 90.11 |
| NAG16715 | Kyoto Univ. | Kansai | 337,983,976 | 336,167,062 | 50,697,596,400 | 50,517,323,949 | 99.46 | 6.64 | 16.84 | 90.54 |
| NAG16731 | Kyoto Univ. | Kansai | 350,657,977 | 348,840,033 | 52,598,696,550 | 52,366,371,354 | 99.48 | 5.86 | 17.46 | 92.16 |
| NAG16754 | Kyoto Univ. | Kansai | 328,759,390 | 326,881,828 | 49,313,908,500 | 49,083,010,029 | 99.43 | 5.80 | 16.36 | 89.92 |
| NAG16756 | Kyoto Univ. | Kansai | 330,110,928 | 328,525,117 | 49,516,639,200 | 49,327,767,802 | 99.52 | 4.47 | 16.44 | 88.11 |
| NAG16759 | Kyoto Univ. | Kansai | 339,273,463 | 337,392,320 | 50,891,019,450 | 50,628,508,769 | 99.45 | 4.90 | 16.88 | 88.54 |
| NAG16764 | Kyoto Univ. | Kansai | 371,758,146 | 368,257,602 | 55,763,721,900 | 55,281,712,375 | 99.06 | 5.29 | 18.43 | 91.15 |
| NAG16769 | Kyoto Univ. | Kansai | 346,606,562 | 344,736,839 | 51,990,984,300 | 51,799,043,401 | 99.46 | 5.01 | 17.27 | 90.29 |
| NAG16770 | Kyoto Univ. | Kansai | 362,726,110 | 360,909,914 | 54,408,916,500 | 54,216,177,089 | 99.50 | 4.47 | 18.07 | 95.25 |
| NAG16772 | Kyoto Univ. | Kansai | 308,117,822 | 305,910,740 | 46,217,673,300 | 45,973,123,593 | 99.28 | 4.42 | 15.32 | 87.08 |
| NAG16786 | Kyoto Univ. | Kansai | 369,117,416 | 367,725,928 | 55,367,612,400 | 55,268,458,899 | 99.62 | 5.38 | 18.42 | 95.83 |
| NAG16788 | Kyoto Univ. | Kansai | 371,327,353 | 368,298,671 | 55,699,102,950 | 55,344,379,064 | 99.18 | 5.20 | 18.45 | 95.40 |
| NAG16802 | Kyoto Univ. | Kansai | 354,852,448 | 353,546,931 | 53,227,867,200 | 53,112,515,242 | 99.63 | 5.43 | 17.70 | 94.46 |
| NAG16845 | Kyoto Univ. | Kansai | 356,220,765 | 354,608,046 | 53,433,114,750 | 53,274,297,128 | 99.55 | 6.09 | 17.76 | 93.98 |
| NAG16859 | Kyoto Univ. | Kansai | 410,255,792 | 408,387,025 | 61,538,368,800 | 61,341,775,458 | 99.54 | 4.82 | 20.45 | 97.81 |
| NAG16860 | Kyoto Univ. | Kansai | 400,482,660 | 399,102,101 | 60,072,399,000 | 59,915,853,240 | 99.66 | 6.07 | 19.97 | 96.93 |
| NAG16863 | Kyoto Univ. | Kansai | 366,244,364 | 363,591,694 | 54,936,654,600 | 54,600,508,224 | 99.28 | 6.03 | 18.20 | 90.84 |
| NAG16878 | Kyoto Univ. | Kansai | 324,768,052 | 323,503,122 | 48,715,207,800 | 48,613,741,944 | 99.61 | 6.09 | 16.20 | 90.28 |
| NAG16882 | Kyoto Univ. | Kansai | 367,802,440 | 366,515,980 | 55,170,366,000 | 55,046,733,970 | 99.65 | 5.92 | 18.35 | 92.16 |
| NAG16884 | Kyoto Univ. | Kansai | 355,837,838 | 354,038,977 | 53,375,675,700 | 53,185,557,759 | 99.49 | 7.34 | 17.73 | 92.37 |
| NAG16897 | Kyoto Univ. | Kansai | 331,677,817 | 329,664,675 | 49,751,672,550 | 49,545,956,796 | 99.39 | 4.98 | 16.52 | 91.37 |
| NAG16901 | Kyoto Univ. | Kansai | 323,857,208 | 321,444,967 | 48,578,581,200 | 48,295,601,785 | 99.26 | 4.24 | 16.10 | 87.09 |
| NAG16908 | Kyoto Univ. | Kansai | 345,158,728 | 343,447,634 | 51,773,809,200 | 51,594,550,671 | 99.50 | 6.30 | 17.20 | 91.55 |
| NAG16922 | Kyoto Univ. | Kansai | 360,186,820 | 358,480,162 | 54,028,023,000 | 53,853,434,959 | 99.53 | 5.11 | 17.95 | 95.14 |
| NAG16924 | Kyoto Univ. | Kansai | 371,457,239 | 369,972,760 | 55,718,585,850 | 55,499,730,442 | 99.60 | 8.49 | 18.50 | 90.93 |
| NAG16929 | Kyoto Univ. | Kansai | 387,740,022 | 386,570,442 | 58,161,003,300 | 58,080,865,416 | 99.70 | 6.13 | 19.36 | 96.92 |
| NAG16931 | Kyoto Univ. | Kansai | 371,738,852 | 369,547,825 | 55,760,827,800 | 55,473,851,148 | 99.41 | 5.74 | 18.49 | 92.12 |
| NAG16941 | Kyoto Univ. | Kansai | 334,413,809 | 332,272,604 | 50,162,071,350 | 49,891,418,133 | 99.36 | 6.78 | 16.63 | 89.24 |
| NAG16942 | Kyoto Univ. | Kansai | 346,802,917 | 344,610,793 | 52,020,437,550 | 51,756,908,475 | 99.37 | 5.91 | 17.25 | 92.40 |
| NAG16954 | Kyoto Univ. | Kansai | 350,812,552 | 349,102,539 | 52,621,882,800 | 52,435,613,637 | 99.51 | 4.64 | 17.48 | 90.97 |
| NAG16957 | Kyoto Univ. | Kansai | 357,419,475 | 356,045,211 | 53,612,921,250 | 53,486,598,138 | 99.62 | 4.77 | 17.83 | 94.83 |
| NAG16965 | Kyoto Univ. | Kansai | 341,052,426 | 339,683,728 | 51,157,863,900 | 51,021,173,685 | 99.60 | 3.91 | 17.01 | 93.33 |
| NAG16969 | Kyoto Univ. | Kansai | 441,456,776 | 438,841,204 | 66,218,516,400 | 65,908,550,111 | 99.41 | 7.06 | 21.97 | 98.02 |
| NAG16984 | Kyoto Univ. | Kansai | 358,755,860 | 357,533,126 | 53,813,379,000 | 53,714,602,554 | 99.66 | 4.98 | 17.90 | 95.33 |
| NAG16988 | Kyoto Univ. | Kansai | 328,278,429 | 326,184,734 | 49,241,764,350 | 48,908,051,001 | 99.36 | 5.74 | 16.30 | 88.54 |
| NAG16991 | Kyoto Univ. | Kansai | 424,560,393 | 422,950,917 | 63,684,058,950 | 63,320,389,050 | 99.62 | 6.64 | 21.11 | 92.63 |
| NAG16993 | Kyoto Univ. | Kansai | 335,719,808 | 333,522,681 | 50,357,971,200 | 50,075,965,180 | 99.35 | 6.12 | 16.69 | 90.80 |
| NAG17004 | Kyoto Univ. | Kansai | 314,118,056 | 311,967,091 | 47,117,708,400 | 46,827,870,970 | 99.32 | 4.37 | 15.61 | 84.07 |
| NAG17008 | Kyoto Univ. | Kansai | 356,784,039 | 354,660,792 | 53,517,605,850 | 53,251,944,398 | 99.40 | 6.19 | 17.75 | 93.50 |
| NAG17009 | Kyoto Univ. | Kansai | 397,332,098 | 395,482,541 | 59,599,814,700 | 59,363,363,577 | 99.53 | 6.88 | 19.79 | 96.28 |
| NAG17012 | Kyoto Univ. | Kansai | 303,638,840 | 301,364,027 | 45,545,826,000 | 45,291,732,871 | 99.25 | 3.34 | 15.10 | 86.79 |
| NAG17016 | Kyoto Univ. | Kansai | 342,680,143 | 340,566,858 | 51,402,021,450 | 51,139,893,014 | 99.38 | 5.56 | 17.05 | 91.87 |
| NAG17017 | Kyoto Univ. | Kansai | 340,922,659 | 339,568,679 | 51,138,398,850 | 51,011,942,594 | 99.60 | 4.96 | 17.00 | 93.13 |
| NAG17018 | Kyoto Univ. | Kansai | 418,497,533 | 417,106,846 | 62,774,629,950 | 62,613,765,781 | 99.67 | 6.09 | 20.87 | 97.69 |
| NAG17019 | Kyoto Univ. | Kansai | 405,287,274 | 403,775,901 | 60,793,091,100 | 60,649,917,913 | 99.63 | 5.18 | 20.22 | 97.66 |
| NAG17022 | Kyoto Univ. | Kansai | 369,554,088 | 367,930,436 | 55,433,113,200 | 55,273,229,225 | 99.56 | 5.04 | 18.42 | 92.74 |
| NAG17024 | Kyoto Univ. | Kansai | 383,543,422 | 382,053,359 | 57,531,513,300 | 57,406,482,797 | 99.61 | 8.62 | 19.14 | 95.71 |
| NAG17033 | Kyoto Univ. | Kansai | 370,261,066 | 368,572,591 | 55,539,159,900 | 55,317,436,564 | 99.54 | 4.89 | 18.44 | 92.30 |
| NAG17038 | Kyoto Univ. | Kansai | 416,580,652 | 415,020,442 | 62,487,097,800 | 62,371,910,878 | 99.63 | 4.67 | 20.79 | 98.17 |
| NAG17039 | Kyoto Univ. | Kansai | 367,640,691 | 366,216,671 | 55,146,103,650 | 55,031,444,528 | 99.61 | 5.18 | 18.34 | 92.68 |
| NAG17041 | Kyoto Univ. | Kansai | 388,547,909 | 385,866,389 | 58,282,186,350 | 57,790,846,366 | 99.31 | 5.40 | 19.26 | 91.88 |
| NAG17042 | Kyoto Univ. | Kansai | 333,852,785 | 332,496,235 | 50,077,917,750 | 49,924,893,461 | 99.59 | 4.59 | 16.64 | 91.61 |
| NAG17043 | Kyoto Univ. | Kansai | 351,258,571 | 349,747,102 | 52,688,785,650 | 52,497,525,333 | 99.57 | 6.60 | 17.50 | 93.06 |
| NAG17046 | Kyoto Univ. | Kansai | 358,608,461 | 354,860,447 | 53,791,269,150 | 53,268,869,866 | 98.95 | 2.93 | 17.76 | 94.12 |
| NAG17050 | Kyoto Univ. | Kansai | 335,298,717 | 333,680,768 | 50,294,807,550 | 50,115,934,988 | 99.52 | 5.45 | 16.71 | 91.48 |
| NAG17058 | Kyoto Univ. | Kansai | 321,456,314 | 319,582,942 | 48,218,447,100 | 47,895,135,770 | 99.42 | 4.77 | 15.97 | 87.84 |
| NAG17059 | Kyoto Univ. | Kansai | 317,499,525 | 314,826,639 | 47,624,928,750 | 47,265,302,522 | 99.16 | 4.99 | 15.76 | 85.01 |
| NAG17060 | Kyoto Univ. | Kansai | 349,285,799 | 347,875,458 | 52,392,869,850 | 52,251,412,784 | 99.60 | 4.70 | 17.42 | 91.15 |
| NAG17064 | Kyoto Univ. | Kansai | 385,248,208 | 383,577,157 | 57,787,231,200 | 57,620,521,505 | 99.57 | 6.24 | 19.21 | 96.49 |
| NAG17070 | Kyoto Univ. | Kansai | 341,588,558 | 339,755,862 | 51,238,283,700 | 50,996,355,695 | 99.46 | 5.49 | 17.00 | 88.89 |
| NAG17074 | Kyoto Univ. | Kansai | 389,326,445 | 387,399,571 | 58,398,966,750 | 58,147,944,012 | 99.51 | 6.78 | 19.38 | 96.38 |
| NAG17077 | Kyoto Univ. | Kansai | 315,575,689 | 314,161,612 | 47,336,353,350 | 47,214,624,722 | 99.55 | 5.04 | 15.74 | 89.19 |
| NAG17081 | Kyoto Univ. | Kansai | 355,101,249 | 353,381,381 | 53,265,187,350 | 53,069,204,247 | 99.52 | 4.12 | 17.69 | 91.69 |
| NAG17082 | Kyoto Univ. | Kansai | 366,542,911 | 364,812,977 | 54,981,436,650 | 54,737,000,886 | 99.53 | 5.94 | 18.25 | 94.79 |
| NAG17088 | Kyoto Univ. | Kansai | 356,775,004 | 355,545,179 | 53,516,250,600 | 53,427,753,740 | 99.66 | 4.82 | 17.81 | 94.95 |
| NAG17092 | Kyoto Univ. | Kansai | 392,820,224 | 391,481,045 | 58,923,033,600 | 58,802,113,695 | 99.66 | 5.75 | 19.60 | 97.14 |
| NAG17098 | Kyoto Univ. | Kansai | 357,690,847 | 355,366,477 | 53,653,627,050 | 53,368,248,535 | 99.35 | 6.05 | 17.79 | 90.32 |
| NAG17104 | Kyoto Univ. | Kansai | 336,817,235 | 335,181,313 | 50,522,585,250 | 50,334,199,956 | 99.51 | 5.67 | 16.78 | 91.27 |
| NAG17105 | Kyoto Univ. | Kansai | 318,231,919 | 316,788,912 | 47,734,787,850 | 47,622,682,940 | 99.55 | 4.81 | 15.87 | 89.75 |
| NAG17106 | Kyoto Univ. | Kansai | 378,832,571 | 377,528,408 | 56,824,885,650 | 56,674,581,349 | 99.66 | 5.74 | 18.89 | 95.94 |
| NAG17110 | Kyoto Univ. | Kansai | 326,613,101 | 325,273,737 | 48,991,965,150 | 48,874,048,866 | 99.59 | 4.66 | 16.29 | 91.39 |
| NAG17113 | Kyoto Univ. | Kansai | 321,497,551 | 320,412,561 | 48,224,632,650 | 48,132,811,174 | 99.66 | 4.36 | 16.04 | 90.66 |
| NAG17121 | Kyoto Univ. | Kansai | 335,962,388 | 334,247,408 | 50,394,358,200 | 50,192,924,576 | 99.49 | 5.07 | 16.73 | 91.96 |
| NAG17125 | Kyoto Univ. | Kansai | 352,220,243 | 351,077,595 | 52,833,036,450 | 52,743,801,748 | 99.68 | 5.00 | 17.58 | 94.47 |
| NAG17150 | Kyoto Univ. | Kansai | 348,175,000 | 346,655,728 | 52,226,250,000 | 52,032,069,058 | 99.56 | 4.90 | 17.34 | 90.08 |
| NAG17151 | Kyoto Univ. | Kansai | 391,216,999 | 388,566,438 | 58,682,549,850 | 58,338,365,902 | 99.32 | 5.78 | 19.45 | 93.76 |
| NAG17158 | Kyoto Univ. | Kansai | 364,430,800 | 362,704,923 | 54,664,620,000 | 54,491,789,165 | 99.53 | 7.13 | 18.16 | 93.71 |
| NAG17162 | Kyoto Univ. | Kansai | 348,157,253 | 346,940,910 | 52,223,587,950 | 52,134,503,060 | 99.65 | 4.80 | 17.38 | 94.16 |
| NAG17169 | Kyoto Univ. | Kansai | 333,519,915 | 332,179,000 | 50,027,987,250 | 49,906,763,277 | 99.60 | 3.95 | 16.64 | 89.21 |
| NAG17177 | Kyoto Univ. | Kansai | 318,144,561 | 316,161,399 | 47,721,684,150 | 47,498,093,635 | 99.38 | 4.42 | 15.83 | 86.54 |
| NAG17186 | Kyoto Univ. | Kansai | 323,655,145 | 322,301,257 | 48,548,271,750 | 48,430,696,185 | 99.58 | 3.95 | 16.14 | 91.21 |
| NAG17194 | Kyoto Univ. | Kansai | 369,434,541 | 367,565,045 | 55,415,181,150 | 55,208,660,331 | 99.49 | 5.42 | 18.40 | 95.36 |
| NAG17195 | Kyoto Univ. | Kansai | 416,227,150 | 414,235,573 | 62,434,072,500 | 62,260,925,874 | 99.52 | 7.55 | 20.75 | 97.06 |
| NAG17205 | Kyoto Univ. | Kansai | 373,454,934 | 371,459,377 | 56,018,240,100 | 55,788,299,828 | 99.47 | 6.28 | 18.60 | 95.48 |
| NAG17208 | Kyoto Univ. | Kansai | 375,199,561 | 373,916,131 | 56,279,934,150 | 56,154,211,064 | 99.66 | 5.34 | 18.72 | 96.14 |
| NAG17210 | Kyoto Univ. | Kansai | 416,621,858 | 414,837,941 | 62,493,278,700 | 62,357,447,925 | 99.57 | 7.75 | 20.79 | 94.42 |
| NAG17226 | Kyoto Univ. | Kansai | 322,593,246 | 319,324,932 | 48,388,986,900 | 47,977,655,695 | 98.99 | 4.60 | 15.99 | 85.42 |
| NAG17232 | Kyoto Univ. | Kansai | 385,406,612 | 383,865,730 | 57,810,991,800 | 57,620,684,986 | 99.60 | 6.25 | 19.21 | 96.33 |
| NAG17243 | Kyoto Univ. | Kansai | 358,089,708 | 356,768,844 | 53,713,456,200 | 53,556,763,269 | 99.63 | 5.50 | 17.85 | 94.14 |

| | | | | | | | | | | |
|---|---|---|---|---|---|---|---|---|---|---|
| NAG17252 | Kyoto Univ. | Kansai | 382,248,600 | 380,239,313 | 57,337,290,000 | 57,068,869,454 | 99.47 | 7.16 | 19.02 | 95.46 |
| NAG17270 | Kyoto Univ. | Kansai | 332,240,667 | 330,745,240 | 49,836,100,050 | 49,696,156,063 | 99.55 | 4.14 | 16.57 | 92.33 |
| NAG17278 | Kyoto Univ. | Kansai | 337,886,707 | 336,545,116 | 50,683,006,050 | 50,547,123,352 | 99.60 | 3.95 | 16.85 | 92.43 |
| NAG17282 | Kyoto Univ. | Kansai | 369,945,758 | 367,924,198 | 55,491,863,700 | 55,290,878,784 | 99.45 | 7.63 | 18.43 | 94.08 |
| NAG17290 | Kyoto Univ. | Kansai | 333,847,793 | 331,712,936 | 50,077,168,950 | 49,861,412,308 | 99.36 | 4.58 | 16.62 | 88.58 |
| NAG17298 | Kyoto Univ. | Kansai | 354,775,866 | 353,269,519 | 53,216,379,900 | 53,044,832,118 | 99.58 | 4.66 | 17.68 | 94.49 |
| NAG17318 | Kyoto Univ. | Kansai | 342,723,420 | 341,261,108 | 51,408,513,000 | 51,294,243,448 | 99.57 | 4.27 | 17.10 | 93.85 |
| NAG17333 | Kyoto Univ. | Kansai | 368,453,210 | 366,219,392 | 55,267,981,500 | 54,856,560,354 | 99.39 | 5.05 | 18.29 | 90.30 |
| NAG17349 | Kyoto Univ. | Kansai | 354,401,984 | 352,868,436 | 53,160,297,600 | 53,029,823,279 | 99.57 | 6.30 | 17.68 | 93.26 |
| NAG17351 | Kyoto Univ. | Kansai | 414,309,551 | 412,407,323 | 62,146,432,650 | 61,912,088,282 | 99.54 | 6.68 | 20.64 | 97.54 |
| NAG17355 | Kyoto Univ. | Kansai | 390,054,567 | 387,688,698 | 58,508,185,050 | 58,223,460,752 | 99.39 | 6.62 | 19.41 | 95.46 |
| NAG17362 | Kyoto Univ. | Kansai | 337,318,225 | 335,540,221 | 50,597,733,750 | 50,364,811,362 | 99.47 | 7.15 | 16.79 | 87.78 |
| NAG17367 | Kyoto Univ. | Kansai | 340,654,052 | 339,009,138 | 51,098,107,800 | 50,909,100,891 | 99.52 | 5.74 | 16.97 | 88.46 |
| NAG17372 | Kyoto Univ. | Kansai | 361,984,364 | 360,463,036 | 54,297,654,600 | 54,150,327,725 | 99.58 | 5.96 | 18.05 | 94.48 |
| NAG17380 | Kyoto Univ. | Kansai | 322,001,515 | 319,968,434 | 48,300,227,250 | 47,998,437,267 | 99.37 | 5.44 | 16.00 | 84.47 |
| NAG17400 | Kyoto Univ. | Kansai | 344,779,127 | 342,385,884 | 51,716,869,050 | 51,367,330,984 | 99.31 | 5.15 | 17.12 | 92.09 |
| NAG17403 | Kyoto Univ. | Kansai | 351,862,605 | 350,098,162 | 52,779,390,750 | 52,571,908,170 | 99.50 | 7.19 | 17.52 | 92.27 |
| NAG17404 | Kyoto Univ. | Kansai | 326,996,445 | 324,533,859 | 49,049,466,750 | 48,759,663,087 | 99.25 | 5.11 | 16.25 | 90.41 |
| NAG17406 | Kyoto Univ. | Kansai | 362,543,282 | 360,872,197 | 54,381,492,300 | 54,246,363,429 | 99.54 | 5.57 | 18.08 | 95.04 |
| NAG17420 | Kyoto Univ. | Kansai | 346,842,656 | 345,076,044 | 52,026,398,400 | 51,846,460,115 | 99.49 | 6.46 | 17.28 | 88.64 |
| NAG17435 | Kyoto Univ. | Kansai | 320,671,939 | 317,683,385 | 48,100,790,850 | 47,707,187,507 | 99.07 | 5.31 | 15.90 | 84.56 |
| NAG17437 | Kyoto Univ. | Kansai | 327,826,394 | 324,200,529 | 49,173,959,100 | 48,638,719,711 | 98.89 | 4.01 | 16.21 | 89.33 |
| NAG17443 | Kyoto Univ. | Kansai | 338,509,826 | 335,828,274 | 50,776,473,900 | 50,472,818,701 | 99.21 | 4.00 | 16.82 | 92.81 |
| NAG17444 | Kyoto Univ. | Kansai | 357,219,279 | 355,486,106 | 53,582,891,850 | 53,388,285,842 | 99.51 | 5.86 | 17.80 | 94.28 |
| NAG17455 | Kyoto Univ. | Kansai | 327,934,943 | 325,938,722 | 49,190,241,450 | 48,933,160,414 | 99.39 | 3.94 | 16.31 | 90.47 |
| NAG17465 | Kyoto Univ. | Kansai | 343,024,901 | 340,506,916 | 51,453,735,150 | 51,012,660,031 | 99.27 | 4.84 | 17.00 | 87.92 |
| NAG17466 | Kyoto Univ. | Kansai | 380,754,732 | 378,738,063 | 57,113,209,800 | 56,868,395,223 | 99.47 | 5.05 | 18.96 | 96.02 |
| NAG17496 | Kyoto Univ. | Kansai | 342,299,442 | 339,872,079 | 51,344,916,300 | 51,031,272,498 | 99.29 | 5.11 | 17.01 | 92.21 |
| NAG17498 | Kyoto Univ. | Kansai | 349,035,191 | 342,999,938 | 52,355,278,650 | 51,492,853,400 | 98.27 | 4.22 | 17.16 | 89.83 |
| NAG17505 | Kyoto Univ. | Kansai | 347,477,205 | 346,074,596 | 52,121,580,750 | 51,925,079,445 | 99.60 | 5.30 | 17.31 | 92.21 |
| NAG17507 | Kyoto Univ. | Kansai | 327,407,146 | 323,512,157 | 49,111,071,900 | 48,565,447,813 | 98.81 | 3.81 | 16.19 | 90.09 |
| NAG17521 | Kyoto Univ. | Kansai | 323,448,853 | 322,038,743 | 48,517,327,950 | 48,359,092,256 | 99.56 | 4.18 | 16.12 | 90.34 |
| NAG17523 | Kyoto Univ. | Kansai | 352,308,944 | 350,282,915 | 52,846,341,600 | 52,589,166,552 | 99.42 | 5.16 | 17.53 | 90.57 |
| NAG17530 | Kyoto Univ. | Kansai | 388,104,403 | 386,580,901 | 58,215,660,450 | 58,074,405,928 | 99.61 | 5.27 | 19.36 | 96.89 |
| NAG17532 | Kyoto Univ. | Kansai | 334,952,547 | 332,280,471 | 50,242,882,050 | 49,915,206,515 | 99.20 | 3.70 | 16.64 | 91.04 |
| NAG17541 | Kyoto Univ. | Kansai | 329,646,354 | 327,950,609 | 49,446,953,100 | 49,209,022,992 | 99.49 | 4.31 | 16.40 | 90.80 |
| NAG17561 | Kyoto Univ. | Kansai | 413,413,862 | 411,524,430 | 62,012,079,300 | 61,849,756,177 | 99.54 | 4.35 | 20.62 | 95.42 |
| NAG17567 | Kyoto Univ. | Kansai | 382,029,766 | 380,754,197 | 57,304,464,900 | 57,198,436,162 | 99.67 | 7.65 | 19.07 | 95.57 |
| NAG17569 | Kyoto Univ. | Kansai | 338,963,143 | 335,128,821 | 50,844,471,450 | 50,348,298,962 | 98.87 | 5.30 | 16.78 | 87.36 |
| NAG17570 | Kyoto Univ. | Kansai | 316,090,380 | 314,089,080 | 47,413,557,000 | 47,133,589,122 | 99.37 | 4.39 | 15.71 | 88.16 |
| NAG17589 | Kyoto Univ. | Kansai | 361,348,514 | 359,084,958 | 54,202,277,100 | 53,902,415,566 | 99.37 | 4.84 | 17.97 | 91.09 |
| NAG17592 | Kyoto Univ. | Kansai | 314,445,525 | 311,965,812 | 47,166,828,750 | 46,841,376,688 | 99.21 | 4.13 | 15.61 | 84.86 |
| NAG17610 | Kyoto Univ. | Kansai | 374,511,074 | 372,336,926 | 56,176,661,100 | 55,939,237,510 | 99.42 | 6.17 | 18.65 | 95.67 |
| NAG17614 | Kyoto Univ. | Kansai | 396,335,255 | 394,937,275 | 59,450,288,250 | 59,334,330,022 | 99.65 | 4.87 | 19.78 | 97.50 |
| NAG17621 | Kyoto Univ. | Kansai | 355,111,981 | 353,675,243 | 53,266,797,150 | 53,146,019,220 | 99.60 | 4.02 | 17.72 | 94.76 |
| NAG17623 | Kyoto Univ. | Kansai | 359,227,068 | 356,752,337 | 53,884,060,200 | 53,566,625,859 | 99.31 | 5.07 | 17.86 | 91.16 |
| NAG17626 | Kyoto Univ. | Kansai | 325,464,218 | 323,538,912 | 48,819,632,700 | 48,556,744,857 | 99.41 | 4.26 | 16.19 | 89.98 |
| NAG17634 | Kyoto Univ. | Kansai | 335,804,376 | 333,605,311 | 50,370,656,400 | 50,086,562,387 | 99.35 | 6.39 | 16.70 | 89.66 |
| NAG17637 | Kyoto Univ. | Kansai | 324,654,214 | 323,394,655 | 48,698,132,100 | 48,592,534,269 | 99.61 | 4.24 | 16.20 | 91.26 |
| NAG17648 | Kyoto Univ. | Kansai | 338,633,030 | 337,004,807 | 50,794,954,500 | 50,560,154,568 | 99.52 | 4.83 | 16.85 | 90.20 |
| NAG17650 | Kyoto Univ. | Kansai | 323,430,478 | 320,805,647 | 48,514,571,700 | 48,162,664,492 | 99.19 | 3.94 | 16.05 | 86.96 |
| NAG17676 | Kyoto Univ. | Kansai | 313,624,447 | 311,643,872 | 47,043,667,050 | 46,789,286,418 | 99.37 | 5.34 | 15.60 | 87.31 |
| NAG17686 | Kyoto Univ. | Kansai | 374,230,993 | 367,833,150 | 56,134,648,950 | 55,226,583,969 | 98.29 | 3.94 | 18.41 | 93.94 |
| NAG17695 | Kyoto Univ. | Kansai | 372,449,364 | 370,134,255 | 55,867,404,600 | 55,547,854,467 | 99.38 | 5.11 | 18.52 | 94.91 |
| NAG17702 | Kyoto Univ. | Kansai | 359,749,338 | 356,066,979 | 53,962,400,700 | 53,507,982,118 | 98.98 | 5.58 | 17.84 | 93.39 |
| NAG17704 | Kyoto Univ. | Kansai | 325,165,855 | 323,118,606 | 48,774,878,250 | 48,534,653,279 | 99.37 | 5.65 | 16.18 | 89.85 |
| NAG17708 | Kyoto Univ. | Kansai | 303,102,603 | 299,767,246 | 45,465,390,450 | 44,970,833,046 | 98.90 | 4.12 | 14.99 | 84.05 |
| NAG17709 | Kyoto Univ. | Kansai | 337,711,196 | 335,721,339 | 50,656,679,400 | 50,424,060,486 | 99.41 | 5.02 | 16.81 | 92.05 |
| NAG17713 | Kyoto Univ. | Kansai | 379,538,567 | 377,149,283 | 56,930,785,050 | 56,668,729,463 | 99.37 | 6.99 | 18.89 | 95.46 |
| NAG17733 | Kyoto Univ. | Kansai | 371,831,571 | 368,224,182 | 55,774,735,650 | 55,292,796,186 | 99.03 | 5.03 | 18.43 | 93.78 |
| NAG17797 | Kyoto Univ. | Kansai | 323,430,935 | 321,640,132 | 48,514,640,250 | 48,231,478,939 | 99.45 | 3.57 | 16.08 | 86.32 |
| NAG17814 | Kyoto Univ. | Kansai | 401,373,498 | 399,519,753 | 60,206,024,700 | 60,022,052,255 | 99.54 | 7.05 | 20.01 | 97.24 |
| NAG17815 | Kyoto Univ. | Kansai | 321,005,787 | 318,398,962 | 48,150,868,050 | 47,756,900,232 | 99.19 | 3.72 | 15.92 | 84.59 |
| NAG17816 | Kyoto Univ. | Kansai | 401,230,350 | 399,358,485 | 60,184,552,500 | 59,963,489,822 | 99.53 | 7.25 | 19.99 | 97.12 |
| NAG17824 | Kyoto Univ. | Kansai | 341,861,490 | 338,735,069 | 51,279,223,500 | 50,874,257,348 | 99.09 | 4.39 | 16.96 | 91.28 |
| NAG17849 | Kyoto Univ. | Kansai | 336,342,110 | 335,202,579 | 50,451,316,500 | 50,354,308,842 | 99.66 | 4.82 | 16.78 | 89.63 |
| NAG17856 | Kyoto Univ. | Kansai | 410,035,393 | 408,167,068 | 61,505,308,950 | 61,329,714,647 | 99.54 | 7.25 | 20.44 | 97.57 |
| NAG17858 | Kyoto Univ. | Kansai | 393,391,634 | 391,571,571 | 59,008,745,100 | 58,794,447,849 | 99.54 | 7.74 | 19.60 | 96.28 |
| NAG17863 | Kyoto Univ. | Kansai | 390,333,871 | 387,569,306 | 58,550,080,650 | 58,225,275,134 | 99.29 | 5.37 | 19.41 | 96.70 |
| NAG17880 | Kyoto Univ. | Kansai | 356,782,434 | 353,174,441 | 53,517,365,100 | 53,016,940,399 | 98.99 | 5.17 | 17.67 | 88.18 |
| NAG17884 | Kyoto Univ. | Kansai | 346,484,920 | 345,300,223 | 51,972,738,000 | 51,871,333,520 | 99.66 | 4.69 | 17.29 | 94.00 |
| NAG17888 | Kyoto Univ. | Kansai | 355,568,533 | 353,721,326 | 53,335,279,950 | 53,128,741,427 | 99.48 | 4.74 | 17.71 | 91.09 |
| NAG17903 | Kyoto Univ. | Kansai | 361,785,710 | 355,840,368 | 54,267,856,500 | 53,385,894,947 | 98.36 | 4.45 | 17.80 | 89.60 |
| NAG17917 | Kyoto Univ. | Kansai | 395,189,473 | 393,667,553 | 59,278,420,950 | 59,192,754,973 | 99.61 | 6.37 | 19.73 | 97.11 |
| NAG17918 | Kyoto Univ. | Kansai | 402,166,272 | 400,370,098 | 60,324,940,800 | 60,058,916,659 | 99.55 | 5.66 | 20.02 | 96.85 |
| NAG17919 | Kyoto Univ. | Kansai | 394,249,432 | 392,265,077 | 59,137,414,800 | 58,893,566,807 | 99.50 | 6.75 | 19.63 | 96.30 |
| NAG17935 | Kyoto Univ. | Kansai | 336,751,165 | 335,041,774 | 50,512,674,750 | 50,321,872,438 | 99.49 | 4.60 | 16.77 | 91.82 |
| NAG17943 | Kyoto Univ. | Kansai | 363,367,507 | 362,154,668 | 54,505,126,050 | 54,413,078,453 | 99.67 | 5.54 | 18.14 | 95.14 |
| NAG17952 | Kyoto Univ. | Kansai | 314,723,267 | 313,129,801 | 47,208,490,050 | 46,929,110,274 | 99.49 | 3.80 | 15.64 | 87.11 |
| NAG17963 | Kyoto Univ. | Kansai | 378,506,203 | 375,107,231 | 56,775,930,450 | 56,244,274,263 | 99.10 | 5.66 | 18.75 | 92.54 |
| NAG17971 | Kyoto Univ. | Kansai | 311,785,497 | 309,842,392 | 46,767,824,550 | 46,444,713,337 | 99.38 | 4.13 | 15.48 | 86.02 |
| NAG17982 | Kyoto Univ. | Kansai | 376,889,211 | 375,401,159 | 56,533,381,650 | 56,330,014,292 | 99.61 | 6.57 | 18.78 | 95.27 |
| NAG17994 | Kyoto Univ. | Kansai | 328,213,109 | 326,629,469 | 49,231,966,350 | 49,060,732,445 | 99.52 | 5.08 | 16.35 | 90.86 |
| NAG17995 | Kyoto Univ. | Kansai | 341,237,243 | 337,527,658 | 51,185,586,450 | 50,689,076,409 | 98.91 | 4.99 | 16.90 | 90.97 |
| NAG18011 | Kyoto Univ. | Kansai | 341,039,756 | 339,169,669 | 51,155,963,400 | 50,949,945,714 | 99.45 | 4.83 | 16.98 | 92.67 |
| NAG18012 | Kyoto Univ. | Kansai | 351,811,125 | 349,821,760 | 52,771,668,750 | 52,447,974,426 | 99.43 | 4.70 | 17.48 | 89.37 |
| NAG18026 | Kyoto Univ. | Kansai | 330,357,690 | 328,480,883 | 49,553,653,500 | 49,094,107,915 | 99.43 | 4.05 | 16.36 | 84.13 |
| NAG18035 | Kyoto Univ. | Kansai | 320,116,938 | 318,168,678 | 48,017,540,700 | 47,724,681,931 | 99.39 | 4.06 | 15.91 | 85.24 |
| NAG18036 | Kyoto Univ. | Kansai | 343,841,984 | 342,171,621 | 51,576,297,600 | 51,394,944,243 | 99.51 | 4.09 | 17.13 | 93.17 |
| NAG18064 | Kyoto Univ. | Kansai | 390,490,797 | 387,694,394 | 58,573,619,550 | 58,078,792,457 | 99.28 | 5.41 | 19.36 | 95.51 |
| NAG18067 | Kyoto Univ. | Kansai | 340,067,036 | 338,000,028 | 51,010,055,400 | 50,711,592,708 | 99.39 | 4.25 | 16.90 | 91.77 |
| NAG18072 | Kyoto Univ. | Kansai | 354,160,558 | 352,092,754 | 53,124,083,700 | 52,805,662,867 | 99.42 | 4.40 | 17.60 | 93.20 |
| NAG18084 | Kyoto Univ. | Kansai | 340,406,629 | 338,804,583 | 51,060,994,350 | 50,887,380,886 | 99.53 | 5.01 | 16.96 | 92.21 |
| NAG18086 | Kyoto Univ. | Kansai | 377,410,059 | 374,040,003 | 56,611,508,850 | 56,183,806,186 | 99.11 | 5.33 | 18.73 | 95.18 |
| NAG18106 | Kyoto Univ. | Kansai | 342,021,310 | 338,769,050 | 51,303,196,500 | 50,909,526,569 | 99.05 | 4.52 | 16.97 | 91.96 |
| NAG18131 | Kyoto Univ. | Kansai | 434,793,472 | 432,684,740 | 65,219,020,800 | 64,949,603,391 | 99.52 | 7.62 | 21.65 | 97.74 |
| NAG18144 | Kyoto Univ. | Kansai | 375,767,838 | 373,768,645 | 56,365,175,700 | 56,127,778,491 | 99.47 | 9.14 | 18.71 | 94.65 |
| NAG18150 | Kyoto Univ. | Kansai | 391,716,897 | 390,235,587 | 58,757,534,550 | 58,620,875,545 | 99.62 | 5.77 | 19.54 | 96.86 |
| NAG18151 | Kyoto Univ. | Kansai | 382,072,524 | 380,465,563 | 57,310,878,600 | 57,186,679,765 | 99.58 | 5.76 | 19.06 | 96.15 |
| NAG18167 | Kyoto Univ. | Kansai | 309,244,085 | 305,265,070 | 46,386,612,750 | 45,860,482,218 | 98.71 | 4.05 | 15.29 | 84.29 |

| | | | | | | | | | | |
|---|---|---|---|---|---|---|---|---|---|---|
| NAG18170 | Kyoto Univ. | Kansai | 405,233,121 | 403,724,070 | 60,784,968,150 | 60,717,344,628 | 99.63 | 4.55 | 20.24 | 95.18 |
| NAG18185 | Kyoto Univ. | Kansai | 380,042,213 | 374,348,072 | 57,006,331,950 | 56,161,334,887 | 98.50 | 5.22 | 18.72 | 92.36 |
| NAG18187 | Kyoto Univ. | Kansai | 355,072,726 | 353,396,562 | 53,260,908,900 | 53,011,747,709 | 99.53 | 4.94 | 17.67 | 92.85 |
| NAG18188 | Kyoto Univ. | Kansai | 392,274,833 | 389,779,275 | 58,841,224,950 | 58,563,454,384 | 99.36 | 5.24 | 19.52 | 96.85 |
| NAG18190 | Kyoto Univ. | Kansai | 405,974,021 | 404,123,453 | 60,896,103,150 | 60,576,479,208 | 99.54 | 6.23 | 20.19 | 93.87 |
| NAG18193 | Kyoto Univ. | Kansai | 375,446,992 | 373,951,315 | 56,317,048,800 | 56,160,782,367 | 99.60 | 5.61 | 18.72 | 96.12 |
| NAG18195 | Kyoto Univ. | Kansai | 320,726,217 | 319,412,937 | 48,108,932,550 | 47,928,772,147 | 99.59 | 6.45 | 15.98 | 88.38 |
| NAG18196 | Kyoto Univ. | Kansai | 353,688,608 | 352,166,528 | 53,053,291,200 | 52,897,342,181 | 99.57 | 5.91 | 17.63 | 94.07 |
| NAG18199 | Kyoto Univ. | Kansai | 332,678,182 | 330,626,078 | 49,901,727,300 | 49,634,333,192 | 99.38 | 5.56 | 16.54 | 90.74 |
| NAG18200 | Kyoto Univ. | Kansai | 348,951,040 | 347,712,468 | 52,342,656,000 | 52,209,332,667 | 99.65 | 5.40 | 17.40 | 93.40 |
| NAG18210 | Kyoto Univ. | Kansai | 341,254,389 | 339,391,662 | 51,188,158,350 | 50,944,254,750 | 99.45 | 4.86 | 16.98 | 91.23 |
| NAG18220 | Kyoto Univ. | Kansai | 354,783,802 | 352,194,831 | 53,217,570,300 | 52,866,171,865 | 99.27 | 3.24 | 17.62 | 94.08 |
| NAG18223 | Kyoto Univ. | Kansai | 346,497,015 | 344,263,177 | 51,974,552,250 | 51,728,852,268 | 99.36 | 4.34 | 17.24 | 93.46 |
| NAG18224 | Kyoto Univ. | Kansai | 327,615,312 | 325,820,629 | 49,142,296,800 | 48,919,607,191 | 99.45 | 3.90 | 16.31 | 90.49 |
| NAG18229 | Kyoto Univ. | Kansai | 332,011,720 | 330,142,156 | 49,801,758,000 | 49,606,830,369 | 99.44 | 4.19 | 16.54 | 91.90 |
| NAG18239 | Kyoto Univ. | Kansai | 348,807,325 | 347,284,509 | 52,321,098,750 | 52,081,903,317 | 99.56 | 5.71 | 17.36 | 90.85 |
| NAG18241 | Kyoto Univ. | Kansai | 354,951,007 | 353,462,193 | 53,242,651,050 | 53,098,770,206 | 99.58 | 5.61 | 17.70 | 90.96 |
| NAG18245 | Kyoto Univ. | Kansai | 351,398,427 | 349,772,196 | 52,709,764,050 | 52,491,273,758 | 99.54 | 6.29 | 17.50 | 92.90 |
| NAG18271 | Kyoto Univ. | Kansai | 369,386,510 | 367,336,217 | 55,407,976,500 | 55,102,575,898 | 99.44 | 6.39 | 18.37 | 91.36 |
| NAG18287 | Kyoto Univ. | Kansai | 338,166,847 | 336,020,860 | 50,725,027,050 | 50,454,025,546 | 99.37 | 5.14 | 16.82 | 91.59 |
| NAG18292 | Kyoto Univ. | Kansai | 398,921,539 | 397,489,690 | 59,838,230,850 | 59,716,782,011 | 99.64 | 5.32 | 19.91 | 97.46 |
| NAG18295 | Kyoto Univ. | Kansai | 388,848,979 | 387,546,820 | 58,327,346,850 | 58,233,226,039 | 99.67 | 6.83 | 19.41 | 93.73 |
| NAG18303 | Kyoto Univ. | Kansai | 373,112,504 | 371,476,773 | 55,966,875,600 | 55,738,498,274 | 99.56 | 4.73 | 18.58 | 95.47 |
| NAG18305 | Kyoto Univ. | Kansai | 355,330,322 | 353,942,049 | 53,299,548,300 | 53,173,670,847 | 99.61 | 4.68 | 17.72 | 94.76 |
| NAG18307 | Kyoto Univ. | Kansai | 323,615,011 | 321,720,561 | 48,542,251,650 | 48,296,021,443 | 99.41 | 6.61 | 16.10 | 85.01 |
| NAG18326 | Kyoto Univ. | Kansai | 339,932,433 | 337,967,543 | 50,989,864,950 | 50,763,623,846 | 99.42 | 5.79 | 16.92 | 88.94 |
| NAG18342 | Kyoto Univ. | Kansai | 362,598,673 | 360,378,617 | 54,389,800,950 | 54,082,151,536 | 99.39 | 6.11 | 18.03 | 94.29 |
| NAG18355 | Kyoto Univ. | Kansai | 373,763,976 | 370,910,822 | 56,064,596,400 | 55,649,087,187 | 99.24 | 5.22 | 18.55 | 94.64 |
| NAG18361 | Kyoto Univ. | Kansai | 416,698,387 | 414,242,366 | 62,504,758,050 | 62,206,464,971 | 99.41 | 6.97 | 20.74 | 97.07 |
| NAG18365 | Kyoto Univ. | Kansai | 401,447,293 | 399,551,937 | 60,217,093,950 | 60,062,897,829 | 99.53 | 6.86 | 20.02 | 97.34 |
| NAG18368 | Kyoto Univ. | Kansai | 358,674,453 | 356,917,785 | 53,801,167,950 | 53,608,612,024 | 99.51 | 4.81 | 17.87 | 91.14 |
| NAG18376 | Kyoto Univ. | Kansai | 453,135,703 | 450,549,851 | 67,970,355,450 | 67,696,080,116 | 99.43 | 6.69 | 22.57 | 98.27 |
| NAG18389 | Kyoto Univ. | Kansai | 352,530,702 | 348,745,359 | 52,879,605,300 | 52,378,356,318 | 98.93 | 4.88 | 17.46 | 89.30 |
| NAG18398 | Kyoto Univ. | Kansai | 355,046,825 | 353,389,022 | 53,257,023,750 | 53,010,201,956 | 99.53 | 10.02 | 17.67 | 87.42 |
| NAG18406 | Kyoto Univ. | Kansai | 341,964,697 | 340,169,428 | 51,294,704,550 | 51,123,127,325 | 99.48 | 4.01 | 17.04 | 93.26 |
| NAG18428 | Kyoto Univ. | Kansai | 372,330,274 | 370,634,279 | 55,849,541,100 | 55,733,851,259 | 99.54 | 5.81 | 18.58 | 92.57 |
| NAG18433 | Kyoto Univ. | Kansai | 404,780,728 | 403,020,233 | 60,717,109,200 | 60,532,940,619 | 99.57 | 9.51 | 20.18 | 96.69 |
| NAG18438 | Kyoto Univ. | Kansai | 328,405,038 | 326,788,928 | 49,260,755,700 | 49,081,155,870 | 99.51 | 4.18 | 16.36 | 91.33 |
| NAG18447 | Kyoto Univ. | Kansai | 405,410,882 | 402,971,214 | 60,811,632,300 | 60,540,369,147 | 99.40 | 7.10 | 20.18 | 97.23 |
| NAG18452 | Kyoto Univ. | Kansai | 399,739,460 | 397,585,732 | 59,960,919,000 | 59,586,634,112 | 99.46 | 8.16 | 19.86 | 92.33 |
| NAG18481 | Kyoto Univ. | Kansai | 346,709,176 | 344,382,944 | 52,006,376,400 | 51,715,927,770 | 99.33 | 5.39 | 17.24 | 92.89 |
| NAG18485 | Kyoto Univ. | Kansai | 392,883,506 | 390,690,653 | 58,932,525,900 | 58,555,338,215 | 99.44 | 6.84 | 19.52 | 92.52 |
| NAG18499 | Kyoto Univ. | Kansai | 342,056,717 | 340,475,812 | 51,308,507,550 | 51,058,485,206 | 99.54 | 4.62 | 17.02 | 91.90 |
| NAG18500 | Kyoto Univ. | Kansai | 344,129,870 | 342,406,286 | 51,619,480,500 | 51,395,799,045 | 99.50 | 5.83 | 17.13 | 92.31 |
| NAG18501 | Kyoto Univ. | Kansai | 323,575,672 | 321,778,762 | 48,536,350,800 | 48,262,564,770 | 99.44 | 5.81 | 16.09 | 85.37 |
| NAG18502 | Kyoto Univ. | Kansai | 350,395,195 | 348,866,382 | 52,559,279,250 | 52,326,293,146 | 99.56 | 6.62 | 17.44 | 89.19 |
| NAG18509 | Kyoto Univ. | Kansai | 325,260,025 | 323,636,413 | 48,789,003,750 | 48,613,959,612 | 99.50 | 4.12 | 16.20 | 90.91 |
| NAG18510 | Kyoto Univ. | Kansai | 374,307,785 | 372,296,618 | 56,146,167,750 | 55,913,748,332 | 99.46 | 4.62 | 18.64 | 96.09 |
| NAG18511 | Kyoto Univ. | Kansai | 350,647,814 | 348,647,060 | 52,597,172,100 | 52,335,606,006 | 99.43 | 5.55 | 17.45 | 89.82 |
| NAG18519 | Kyoto Univ. | Kansai | 350,707,303 | 348,321,731 | 52,606,095,450 | 52,310,546,503 | 99.32 | 4.43 | 17.44 | 93.85 |
| NAG18520 | Kyoto Univ. | Kansai | 381,293,772 | 379,763,627 | 57,194,065,800 | 57,000,021,457 | 99.60 | 6.56 | 19.00 | 95.78 |
| NAG18523 | Kyoto Univ. | Kansai | 374,347,383 | 372,678,546 | 56,152,107,450 | 55,895,733,809 | 99.55 | 6.37 | 18.63 | 94.74 |
| NAG18527 | Kyoto Univ. | Kansai | 360,765,909 | 358,177,047 | 54,114,886,350 | 53,750,735,822 | 99.28 | 3.99 | 17.92 | 94.19 |
| NAG18533 | Kyoto Univ. | Kansai | 358,087,444 | 356,944,995 | 53,713,116,600 | 53,614,150,262 | 99.68 | 7.82 | 17.87 | 93.79 |
| NAG18535 | Kyoto Univ. | Kansai | 364,450,290 | 362,931,450 | 54,667,543,500 | 54,532,311,602 | 99.58 | 6.87 | 18.18 | 91.69 |
| NAG18536 | Kyoto Univ. | Kansai | 381,383,363 | 379,567,500 | 57,207,504,450 | 56,973,319,287 | 99.52 | 6.49 | 18.99 | 95.87 |
| NAG18539 | Kyoto Univ. | Kansai | 419,927,886 | 418,299,198 | 62,989,182,900 | 62,861,656,814 | 99.61 | 9.22 | 20.95 | 97.70 |
| NAG18552 | Kyoto Univ. | Kansai | 379,698,774 | 378,070,859 | 56,954,816,100 | 56,804,966,149 | 99.57 | 9.11 | 18.93 | 95.20 |
| NAG18554 | Kyoto Univ. | Kansai | 363,368,063 | 361,661,893 | 54,505,209,450 | 54,353,342,986 | 99.53 | 6.32 | 18.12 | 95.19 |
| NAG18557 | Kyoto Univ. | Kansai | 375,718,231 | 374,077,842 | 56,357,734,650 | 56,123,645,255 | 99.56 | 9.83 | 18.71 | 92.74 |
| NAG18566 | Kyoto Univ. | Kansai | 357,962,196 | 356,225,583 | 53,694,329,400 | 53,482,496,949 | 99.51 | 7.04 | 17.83 | 93.41 |
| NAG18568 | Kyoto Univ. | Kansai | 406,264,093 | 405,033,667 | 60,939,613,950 | 60,834,782,367 | 99.70 | 7.43 | 20.28 | 97.11 |
| NAG18574 | Kyoto Univ. | Kansai | 391,926,859 | 390,054,625 | 58,789,028,850 | 58,533,212,177 | 99.52 | 6.85 | 19.51 | 96.29 |
| NAG18576 | Kyoto Univ. | Kansai | 340,674,500 | 339,023,150 | 51,101,175,000 | 50,907,300,071 | 99.52 | 6.07 | 16.97 | 91.81 |
| NAG18577 | Kyoto Univ. | Kansai | 333,140,074 | 331,194,480 | 49,971,011,100 | 49,580,887,167 | 99.42 | 7.03 | 16.53 | 85.83 |
| NAG18580 | Kyoto Univ. | Kansai | 349,024,617 | 346,537,088 | 52,353,692,550 | 51,976,625,508 | 99.29 | 4.54 | 17.33 | 92.81 |
| NAG18592 | Kyoto Univ. | Kansai | 346,447,864 | 344,692,740 | 51,967,179,600 | 51,715,037,222 | 99.49 | 5.67 | 17.24 | 92.44 |
| NAG18599 | Kyoto Univ. | Kansai | 433,639,677 | 431,886,396 | 65,045,951,550 | 64,912,914,097 | 99.60 | 6.08 | 21.64 | 95.90 |
| NAG18610 | Kyoto Univ. | Kansai | 393,305,805 | 391,920,570 | 58,995,870,750 | 58,855,811,883 | 99.65 | 7.73 | 19.62 | 96.61 |
| NAG18623 | Kyoto Univ. | Kansai | 338,119,873 | 336,259,390 | 50,717,980,950 | 50,457,045,286 | 99.45 | 6.35 | 16.82 | 90.77 |
| NAG18625 | Kyoto Univ. | Kansai | 341,178,404 | 339,204,711 | 51,176,760,600 | 50,943,918,273 | 99.42 | 6.38 | 16.98 | 90.50 |
| NAG18646 | Kyoto Univ. | Kansai | 356,420,079 | 354,830,952 | 53,463,011,850 | 53,334,615,962 | 99.55 | 8.98 | 17.78 | 90.17 |
| NAG18648 | Kyoto Univ. | Kansai | 447,748,452 | 446,439,851 | 67,162,267,800 | 67,069,396,532 | 99.71 | 8.24 | 22.36 | 98.52 |
| NAG18649 | Kyoto Univ. | Kansai | 345,765,725 | 343,517,213 | 51,864,858,750 | 51,602,429,111 | 99.35 | 7.00 | 17.20 | 92.03 |
| NAG18652 | Kyoto Univ. | Kansai | 416,364,863 | 403,929,350 | 62,454,729,450 | 60,604,308,149 | 97.01 | 2.28 | 20.20 | 94.78 |
| NAG18653 | Kyoto Univ. | Kansai | 321,513,317 | 319,447,771 | 48,226,997,550 | 47,971,671,904 | 99.36 | 4.85 | 15.99 | 88.98 |
| NAG18659 | Kyoto Univ. | Kansai | 375,693,200 | 373,625,474 | 56,353,980,000 | 56,125,782,817 | 99.45 | 7.94 | 18.71 | 95.09 |
| NAG18662 | Kyoto Univ. | Kansai | 368,304,412 | 365,310,976 | 55,245,661,800 | 54,861,572,261 | 99.19 | 6.03 | 18.29 | 95.00 |
| NAG18665 | Kyoto Univ. | Kansai | 352,385,459 | 350,560,906 | 52,857,818,850 | 52,632,873,810 | 99.48 | 7.10 | 17.54 | 89.69 |
| NAG18670 | Kyoto Univ. | Kansai | 369,466,610 | 367,880,738 | 55,419,991,500 | 55,277,936,866 | 99.57 | 7.45 | 18.43 | 95.07 |
| NAG18675 | Kyoto Univ. | Kansai | 404,177,402 | 400,818,325 | 60,626,610,300 | 60,121,878,670 | 99.17 | 7.19 | 20.04 | 96.36 |
| NAG18691 | Kyoto Univ. | Kansai | 367,550,562 | 365,697,874 | 55,132,584,300 | 54,930,146,046 | 99.50 | 7.30 | 18.31 | 94.60 |
| NAG18719 | Kyoto Univ. | Kansai | 370,953,541 | 369,785,611 | 55,643,031,150 | 55,537,835,184 | 99.69 | 7.33 | 18.51 | 95.16 |
| NAG18728 | Kyoto Univ. | Kansai | 348,283,361 | 346,583,869 | 52,242,504,150 | 52,012,366,366 | 99.51 | 6.90 | 17.34 | 92.37 |
| NAG18743 | Kyoto Univ. | Kansai | 418,985,541 | 414,409,294 | 62,847,831,150 | 62,237,535,090 | 98.91 | 3.63 | 20.75 | 95.90 |
| NAG18745 | Kyoto Univ. | Kansai | 447,409,158 | 444,183,421 | 67,111,373,700 | 66,649,620,828 | 99.28 | 7.77 | 22.22 | 98.17 |
| NAG18756 | Kyoto Univ. | Kansai | 326,463,038 | 324,944,304 | 48,969,455,700 | 48,797,660,290 | 99.53 | 5.82 | 16.27 | 89.93 |
| NAG18758 | Kyoto Univ. | Kansai | 400,224,439 | 398,721,295 | 60,033,665,850 | 59,880,162,181 | 99.62 | 6.93 | 19.96 | 96.61 |
| NAG18787 | Kyoto Univ. | Kansai | 351,621,957 | 349,482,258 | 52,743,293,550 | 52,362,880,875 | 99.39 | 7.63 | 17.45 | 86.61 |
| NAG18803 | Kyoto Univ. | Kansai | 316,174,981 | 313,780,878 | 47,426,247,150 | 47,116,833,012 | 99.24 | 5.27 | 15.71 | 87.49 |
| NAG18813 | Kyoto Univ. | Kansai | 333,724,606 | 332,030,066 | 50,058,690,900 | 49,843,811,081 | 99.49 | 5.19 | 16.61 | 87.68 |
| NAG18835 | Kyoto Univ. | Kansai | 421,322,232 | 419,328,095 | 63,198,334,800 | 62,887,352,289 | 99.53 | 10.43 | 20.96 | 96.22 |
| NAG18847 | Kyoto Univ. | Kansai | 384,160,756 | 382,486,229 | 57,624,113,400 | 57,464,291,787 | 99.56 | 11.48 | 19.15 | 94.67 |
| NAG18862 | Kyoto Univ. | Kansai | 362,443,501 | 360,373,485 | 54,366,525,150 | 54,167,994,699 | 99.43 | 4.90 | 18.06 | 95.10 |
| NAG18870 | Kyoto Univ. | Kansai | 375,417,429 | 373,672,096 | 56,312,614,350 | 56,121,009,333 | 99.54 | 9.73 | 18.71 | 91.20 |
| NAG18871 | Kyoto Univ. | Kansai | 359,516,654 | 357,710,919 | 53,927,498,100 | 53,670,106,738 | 99.50 | 4.58 | 17.89 | 94.59 |
| NAG18875 | Kyoto Univ. | Kansai | 361,195,047 | 358,922,746 | 54,179,257,050 | 53,890,067,761 | 99.37 | 6.16 | 17.96 | 90.97 |
| NAG18883 | Kyoto Univ. | Kansai | 348,695,429 | 346,858,619 | 52,304,314,350 | 52,059,200,639 | 99.47 | 5.81 | 17.35 | 88.68 |
| NAG18897 | Kyoto Univ. | Kansai | 342,448,922 | 340,527,013 | 51,367,338,300 | 51,167,777,751 | 99.44 | 5.51 | 17.06 | 92.80 |
| NAG18899 | Kyoto Univ. | Kansai | 338,784,791 | 337,413,472 | 50,817,718,650 | 50,682,921,839 | 99.60 | 6.02 | 16.89 | 92.27 |

| | | | | | | | | | | |
|---|---|---|---|---|---|---|---|---|---|---|
| NAG18901 | Kyoto Univ. | Kansai | 381,559,353 | 380,012,536 | 57,233,902,950 | 57,041,978,122 | 99.59 | 9.94 | 19.01 | 94.67 |
| NAG18914 | Kyoto Univ. | Kansai | 363,204,647 | 361,302,275 | 54,480,697,050 | 54,284,903,062 | 99.48 | 3.72 | 18.09 | 95.30 |
| NAG18916 | Kyoto Univ. | Kansai | 324,710,718 | 323,145,808 | 48,706,607,700 | 48,531,302,203 | 99.52 | 3.92 | 16.18 | 90.42 |
| NAG18946 | Kyoto Univ. | Kansai | 334,120,275 | 332,038,529 | 50,118,041,250 | 49,777,272,954 | 99.38 | 4.14 | 16.59 | 87.66 |
| NAG18965 | Kyoto Univ. | Kansai | 339,994,812 | 338,393,013 | 50,999,221,800 | 50,781,193,766 | 99.53 | 4.04 | 16.93 | 91.93 |
| NAG18970 | Kyoto Univ. | Kansai | 331,150,454 | 329,043,423 | 49,672,568,100 | 49,228,847,836 | 99.36 | 4.06 | 16.41 | 84.08 |
| NAG18974 | Kyoto Univ. | Kansai | 356,407,342 | 354,545,939 | 53,461,101,300 | 53,172,581,792 | 99.48 | 4.58 | 17.72 | 93.90 |
| NAG18978 | Kyoto Univ. | Kansai | 334,630,075 | 332,329,150 | 50,194,511,250 | 49,901,239,287 | 99.31 | 4.76 | 16.63 | 91.30 |
| NAG18984 | Kyoto Univ. | Kansai | 422,628,115 | 420,841,114 | 63,394,217,250 | 63,035,145,580 | 99.58 | 10.56 | 21.01 | 95.92 |
| NAG18985 | Kyoto Univ. | Kansai | 372,558,932 | 370,866,978 | 55,883,839,800 | 55,537,593,432 | 99.55 | 6.42 | 18.51 | 88.87 |
| NAG18988 | Kyoto Univ. | Kansai | 341,349,086 | 339,114,615 | 51,202,362,900 | 50,939,922,149 | 99.35 | 5.61 | 16.98 | 92.18 |
| NAG18993 | Kyoto Univ. | Kansai | 374,384,935 | 372,332,506 | 56,157,740,250 | 55,922,328,185 | 99.45 | 4.87 | 18.64 | 95.76 |
| NAG19002 | Kyoto Univ. | Kansai | 320,223,668 | 318,230,686 | 48,033,550,200 | 47,812,691,529 | 99.38 | 5.28 | 15.94 | 89.29 |
| NAG19008 | Kyoto Univ. | Kansai | 337,789,228 | 335,223,250 | 50,668,384,200 | 50,353,011,587 | 99.24 | 4.22 | 16.78 | 89.33 |
| NAG19023 | Kyoto Univ. | Kansai | 382,829,949 | 381,268,034 | 57,424,492,350 | 57,244,061,212 | 99.59 | 7.05 | 19.08 | 95.90 |
| NAG19027 | Kyoto Univ. | Kansai | 347,262,120 | 346,007,548 | 52,089,318,000 | 51,992,449,924 | 99.64 | 8.44 | 17.33 | 92.58 |
| NAG19036 | Kyoto Univ. | Kansai | 332,680,963 | 330,154,027 | 49,902,144,450 | 49,547,206,024 | 99.24 | 4.84 | 16.52 | 90.80 |
| NAG19061 | Kyoto Univ. | Kansai | 399,041,930 | 397,263,753 | 59,856,289,500 | 59,626,286,230 | 99.55 | 5.88 | 19.88 | 96.83 |
| NAG19063 | Kyoto Univ. | Kansai | 391,722,594 | 387,134,321 | 58,758,389,100 | 58,031,469,667 | 98.83 | 4.86 | 19.34 | 93.42 |
| NAG19067 | Kyoto Univ. | Kansai | 385,479,883 | 382,506,160 | 57,821,982,450 | 57,489,915,458 | 99.23 | 5.30 | 19.16 | 96.44 |
| NAG19071 | Kyoto Univ. | Kansai | 338,141,072 | 336,179,093 | 50,721,160,800 | 50,502,088,138 | 99.42 | 5.98 | 16.83 | 91.74 |
| NAG19083 | Kyoto Univ. | Kansai | 341,694,614 | 338,887,395 | 51,254,192,100 | 50,902,202,199 | 99.18 | 5.99 | 16.97 | 91.89 |
| NAG19087 | Kyoto Univ. | Kansai | 381,073,097 | 379,441,019 | 57,160,964,550 | 56,989,271,741 | 99.57 | 5.57 | 19.00 | 96.30 |
| NAG19094 | Kyoto Univ. | Kansai | 381,666,981 | 379,504,924 | 57,250,047,150 | 57,004,018,544 | 99.43 | 5.86 | 19.00 | 96.21 |
| NAG19095 | Kyoto Univ. | Kansai | 309,354,288 | 307,565,262 | 46,403,143,200 | 46,159,993,341 | 99.42 | 4.59 | 15.39 | 86.18 |
| NAG19096 | Kyoto Univ. | Kansai | 362,055,740 | 359,779,122 | 54,308,361,000 | 54,013,774,471 | 99.37 | 5.12 | 18.00 | 94.78 |
| NAG19098 | Kyoto Univ. | Kansai | 351,008,453 | 348,379,018 | 52,651,267,950 | 52,328,094,836 | 99.25 | 5.70 | 17.44 | 90.33 |
| NAG19102 | Kyoto Univ. | Kansai | 337,332,687 | 334,338,933 | 50,599,903,050 | 50,195,098,351 | 99.11 | 5.29 | 16.73 | 88.08 |
| NAG19106 | Kyoto Univ. | Kansai | 348,447,533 | 346,513,595 | 52,267,129,950 | 52,065,420,610 | 99.44 | 4.05 | 17.36 | 90.84 |
| NAG19110 | Kyoto Univ. | Kansai | 367,896,559 | 362,228,627 | 55,184,483,850 | 54,311,390,332 | 98.46 | 4.00 | 18.10 | 92.06 |
| NAG19122 | Kyoto Univ. | Kansai | 355,870,998 | 353,427,943 | 53,380,649,700 | 53,020,730,101 | 99.31 | 3.43 | 17.67 | 93.95 |
| NAG19125 | Kyoto Univ. | Kansai | 331,671,109 | 328,651,520 | 49,750,666,350 | 49,275,577,387 | 99.09 | 4.97 | 16.43 | 88.50 |
| NAG19135 | Kyoto Univ. | Kansai | 362,000,696 | 360,577,854 | 54,300,104,400 | 54,159,349,948 | 99.61 | 3.93 | 18.05 | 95.51 |
| NAG19136 | Kyoto Univ. | Kansai | 350,216,517 | 347,451,216 | 52,532,477,550 | 52,179,114,958 | 99.21 | 5.64 | 17.39 | 92.57 |
| NAG19141 | Kyoto Univ. | Kansai | 344,843,545 | 342,135,432 | 51,726,531,750 | 51,403,042,419 | 99.21 | 3.52 | 17.13 | 90.49 |
| NAG19145 | Kyoto Univ. | Kansai | 320,810,603 | 318,156,872 | 48,121,590,450 | 47,759,974,885 | 99.17 | 5.02 | 15.92 | 88.88 |
| NAG19147 | Kyoto Univ. | Kansai | 334,097,415 | 332,156,526 | 50,114,612,250 | 49,897,895,290 | 99.42 | 3.67 | 16.63 | 92.09 |
| NAG19148 | Kyoto Univ. | Kansai | 326,462,335 | 324,260,040 | 48,969,350,250 | 48,634,525,844 | 99.33 | 5.52 | 16.21 | 88.62 |
| NAG19152 | Kyoto Univ. | Kansai | 368,313,034 | 366,684,631 | 55,246,955,100 | 55,114,291,951 | 99.56 | 4.90 | 18.37 | 95.77 |
| NAG19159 | Kyoto Univ. | Kansai | 374,521,346 | 372,977,241 | 56,178,201,900 | 55,978,591,918 | 99.59 | 9.24 | 18.66 | 93.47 |
| NAG19178 | Kyoto Univ. | Kansai | 361,995,346 | 360,159,124 | 54,299,301,900 | 53,953,133,806 | 99.49 | 4.09 | 17.98 | 94.01 |
| NAG19194 | Kyoto Univ. | Kansai | 382,529,062 | 374,979,077 | 57,379,359,300 | 56,264,187,389 | 98.03 | 2.66 | 18.75 | 94.43 |
| NAG19196 | Kyoto Univ. | Kansai | 349,017,356 | 344,235,259 | 52,352,603,400 | 51,696,418,328 | 98.63 | 4.43 | 17.23 | 89.98 |
| NAG19207 | Kyoto Univ. | Kansai | 368,510,437 | 367,071,367 | 55,276,565,550 | 55,137,138,006 | 99.61 | 5.39 | 18.38 | 95.54 |
| NAG19210 | Kyoto Univ. | Kansai | 357,188,253 | 355,369,510 | 53,578,237,950 | 53,379,522,342 | 99.49 | 3.50 | 17.79 | 91.98 |
| NAG19220 | Kyoto Univ. | Kansai | 383,319,571 | 381,700,614 | 57,497,935,650 | 57,321,575,464 | 99.58 | 8.80 | 19.11 | 92.47 |
| NAG19224 | Kyoto Univ. | Kansai | 365,384,719 | 362,740,416 | 54,807,707,850 | 54,440,470,101 | 99.28 | 4.52 | 18.15 | 94.84 |
| NAG19231 | Kyoto Univ. | Kansai | 368,046,365 | 366,544,246 | 55,206,954,750 | 55,022,958,605 | 99.59 | 6.04 | 18.34 | 94.99 |
| NAG19242 | Kyoto Univ. | Kansai | 320,451,584 | 318,010,922 | 48,067,737,600 | 47,719,206,594 | 99.24 | 4.80 | 15.91 | 85.28 |
| NAG19248 | Kyoto Univ. | Kansai | 339,463,649 | 337,780,689 | 50,919,547,350 | 50,697,663,475 | 99.50 | 4.41 | 16.90 | 92.09 |
| NAG19256 | Kyoto Univ. | Kansai | 366,718,965 | 365,096,425 | 55,007,844,750 | 54,757,871,108 | 99.56 | 6.39 | 18.25 | 93.09 |
| NAG19291 | Kyoto Univ. | Kansai | 460,787,812 | 459,087,335 | 69,118,171,800 | 68,876,856,223 | 99.63 | 7.35 | 22.96 | 98.53 |
| NAG19293 | Kyoto Univ. | Kansai | 429,093,578 | 426,321,484 | 64,364,036,700 | 64,021,373,744 | 99.35 | 5.55 | 21.34 | 98.26 |
| NAG19303 | Kyoto Univ. | Kansai | 323,423,934 | 322,270,772 | 48,513,590,100 | 48,379,212,329 | 99.64 | 4.79 | 16.13 | 89.76 |
| NAG19306 | Kyoto Univ. | Kansai | 364,841,334 | 355,584,610 | 54,726,200,100 | 53,362,880,616 | 97.46 | 2.51 | 17.79 | 91.93 |
| NAG19307 | Kyoto Univ. | Kansai | 376,315,262 | 373,481,344 | 56,447,289,300 | 56,016,960,778 | 99.25 | 3.91 | 18.67 | 96.02 |
| NAG19314 | Kyoto Univ. | Kansai | 359,356,825 | 357,014,150 | 53,903,523,750 | 53,621,365,208 | 99.35 | 6.34 | 17.87 | 89.74 |
| NAG19329 | Kyoto Univ. | Kansai | 403,792,887 | 401,945,800 | 60,568,933,050 | 60,417,992,492 | 99.54 | 8.96 | 20.14 | 96.94 |
| NAG19338 | Kyoto Univ. | Kansai | 325,063,063 | 323,032,460 | 48,759,459,450 | 48,546,826,406 | 99.38 | 3.29 | 16.18 | 88.24 |
| NAG19339 | Kyoto Univ. | Kansai | 393,638,411 | 392,138,245 | 59,045,761,650 | 58,916,297,864 | 99.62 | 8.58 | 19.64 | 96.49 |
| NAG19349 | Kyoto Univ. | Kansai | 355,038,571 | 352,625,701 | 53,255,785,650 | 52,943,679,518 | 99.32 | 5.12 | 17.65 | 93.58 |
| NAG19350 | Kyoto Univ. | Kansai | 479,761,233 | 478,018,436 | 71,964,184,950 | 71,835,575,274 | 99.64 | 6.87 | 23.95 | 98.93 |
| NAG19357 | Kyoto Univ. | Kansai | 385,033,885 | 375,829,122 | 57,755,082,750 | 56,409,517,783 | 97.61 | 3.60 | 18.80 | 94.07 |
| NAG19361 | Kyoto Univ. | Kansai | 374,027,831 | 371,886,945 | 56,104,174,650 | 55,899,649,173 | 99.43 | 4.37 | 18.63 | 96.17 |
| NAG19371 | Kyoto Univ. | Kansai | 382,315,176 | 380,601,664 | 57,347,276,400 | 57,020,746,473 | 99.55 | 4.87 | 19.01 | 95.22 |
| NAG19375 | Kyoto Univ. | Kansai | 372,466,629 | 370,119,454 | 55,869,994,350 | 55,626,205,482 | 99.37 | 5.89 | 18.54 | 95.58 |
| NAG19378 | Kyoto Univ. | Kansai | 395,481,931 | 392,416,536 | 59,322,289,650 | 58,821,036,543 | 99.22 | 4.39 | 19.61 | 93.78 |
| NAG19384 | Kyoto Univ. | Kansai | 354,810,227 | 352,851,115 | 53,221,534,050 | 52,996,845,905 | 99.45 | 5.28 | 17.67 | 90.65 |
| NAG19393 | Kyoto Univ. | Kansai | 342,987,200 | 340,226,102 | 51,448,080,000 | 51,107,093,856 | 99.19 | 4.93 | 17.04 | 92.46 |
| NAG19401 | Kyoto Univ. | Kansai | 330,138,375 | 328,535,141 | 49,520,756,250 | 49,361,105,249 | 99.51 | 4.84 | 16.45 | 88.02 |
| NAG19403 | Kyoto Univ. | Kansai | 364,233,584 | 360,554,281 | 54,635,037,600 | 54,106,542,090 | 98.99 | 4.00 | 18.04 | 92.09 |
| NAG19412 | Kyoto Univ. | Kansai | 395,153,753 | 393,727,173 | 59,273,062,950 | 59,138,257,854 | 99.64 | 9.35 | 19.71 | 96.31 |
| NAG19415 | Kyoto Univ. | Kansai | 385,531,868 | 364,211,713 | 57,829,780,200 | 54,697,112,530 | 94.47 | -0.10 | 18.23 | 91.32 |
| NAG19424 | Kyoto Univ. | Kansai | 453,737,603 | 450,329,339 | 68,060,640,450 | 67,563,100,410 | 99.25 | 4.47 | 22.52 | 96.42 |
| NAG19427 | Kyoto Univ. | Kansai | 430,744,478 | 427,667,633 | 64,611,671,700 | 64,205,151,835 | 99.29 | 4.62 | 21.40 | 98.30 |
| NAG19429 | Kyoto Univ. | Kansai | 349,249,799 | 347,195,202 | 52,387,469,850 | 52,124,669,413 | 99.41 | 5.86 | 17.37 | 89.54 |
| NAG19430 | Kyoto Univ. | Kansai | 356,956,897 | 354,316,287 | 53,543,534,550 | 53,093,035,664 | 99.26 | 4.65 | 17.70 | 90.24 |
| NAG19432 | Kyoto Univ. | Kansai | 342,070,401 | 340,082,604 | 51,310,560,150 | 51,038,100,109 | 99.42 | 5.57 | 17.01 | 88.41 |
| NAG19433 | Kyoto Univ. | Kansai | 474,287,576 | 472,696,892 | 71,143,136,400 | 70,948,794,654 | 99.66 | 7.28 | 23.65 | 96.72 |
| NAG19449 | Kyoto Univ. | Kansai | 353,783,016 | 351,874,551 | 53,067,452,400 | 52,862,233,999 | 99.46 | 4.25 | 17.62 | 91.30 |
| NAG19458 | Kyoto Univ. | Kansai | 359,563,272 | 358,293,712 | 53,934,490,800 | 53,865,152,277 | 99.65 | 8.69 | 17.96 | 93.90 |
| NAG19461 | Kyoto Univ. | Kansai | 340,166,647 | 335,079,014 | 51,024,997,050 | 50,285,897,350 | 98.50 | 3.91 | 16.76 | 91.60 |
| NAG19462 | Kyoto Univ. | Kansai | 359,503,404 | 356,715,486 | 53,925,510,600 | 53,458,799,238 | 99.22 | 5.23 | 17.82 | 93.10 |
| NAG19477 | Kyoto Univ. | Kansai | 381,635,819 | 377,053,964 | 57,245,372,850 | 56,486,236,575 | 98.80 | 4.72 | 18.83 | 91.30 |
| NAG19489 | Kyoto Univ. | Kansai | 373,070,174 | 370,676,705 | 55,960,526,100 | 55,694,304,935 | 99.36 | 3.86 | 18.56 | 92.85 |
| NAG19491 | Kyoto Univ. | Kansai | 327,919,865 | 326,747,993 | 49,187,979,750 | 49,049,912,893 | 99.64 | 5.54 | 16.35 | 86.78 |
| NAG19492 | Kyoto Univ. | Kansai | 481,982,930 | 480,030,193 | 72,297,439,500 | 72,114,319,586 | 99.59 | 5.86 | 24.04 | 97.24 |
| NAG19506 | Kyoto Univ. | Kansai | 326,329,122 | 324,362,075 | 48,949,368,300 | 48,626,352,748 | 99.40 | 3.62 | 16.21 | 86.94 |
| NAG19515 | Kyoto Univ. | Kansai | 317,514,000 | 315,249,934 | 47,627,100,000 | 47,371,954,827 | 99.29 | 4.83 | 15.79 | 88.97 |
| NAG19521 | Kyoto Univ. | Kansai | 402,625,953 | 400,910,199 | 60,393,892,950 | 60,250,487,004 | 99.57 | 9.79 | 20.08 | 96.77 |
| NAG19523 | Kyoto Univ. | Kansai | 403,837,097 | 400,395,412 | 60,575,564,550 | 60,087,336,029 | 99.15 | 5.59 | 20.03 | 96.68 |
| NAG19529 | Kyoto Univ. | Kansai | 464,986,272 | 462,007,680 | 69,747,940,800 | 69,310,767,065 | 99.36 | 4.76 | 23.10 | 96.72 |
| NAG19530 | Kyoto Univ. | Kansai | 401,895,757 | 399,556,082 | 60,284,363,550 | 60,007,910,734 | 99.42 | 4.64 | 20.00 | 97.52 |
| NAG19536 | Kyoto Univ. | Kansai | 404,665,504 | 403,115,897 | 60,699,825,600 | 60,537,293,316 | 99.62 | 4.85 | 20.18 | 97.51 |
| NAG19537 | Kyoto Univ. | Kansai | 404,042,426 | 402,469,078 | 60,606,363,900 | 60,454,923,584 | 99.61 | 10.95 | 20.15 | 96.38 |
| NAG19538 | Kyoto Univ. | Kansai | 377,482,575 | 370,885,500 | 56,622,386,250 | 55,648,466,788 | 98.25 | 3.30 | 18.55 | 93.24 |
| NAG19543 | Kyoto Univ. | Kansai | 362,413,864 | 355,640,831 | 54,362,079,600 | 53,375,595,161 | 98.13 | 3.26 | 17.79 | 91.89 |
| NAG19547 | Kyoto Univ. | Kansai | 384,051,571 | 381,392,064 | 57,607,735,650 | 57,244,599,861 | 99.31 | 5.26 | 19.08 | 95.99 |
| NAG19549 | Kyoto Univ. | Kansai | 373,137,878 | 371,578,181 | 55,970,681,700 | 55,835,939,639 | 99.58 | 8.82 | 18.61 | 91.83 |
| NAG19553 | Kyoto Univ. | Kansai | 345,002,940 | 342,818,499 | 51,750,441,000 | 51,485,288,359 | 99.37 | 3.52 | 17.16 | 90.28 |

| | | | | | | | | | | |
|---|---|---|---|---|---|---|---|---|---|---|
| NAG19555 | Kyoto Univ. | Kansai | 360,427,284 | 358,118,554 | 54,064,092,600 | 53,788,426,577 | 99.36 | 3.68 | 17.93 | 91.83 |
| NAG19564 | Kyoto Univ. | Kansai | 377,429,239 | 376,008,181 | 56,614,385,850 | 56,486,742,843 | 99.62 | 9.25 | 18.83 | 95.21 |
| NAG19568 | Kyoto Univ. | Kansai | 339,688,727 | 337,913,562 | 50,953,309,050 | 50,570,481,859 | 99.48 | 3.84 | 16.86 | 90.41 |
| NAG19570 | Kyoto Univ. | Kansai | 365,677,651 | 364,042,779 | 54,851,647,650 | 54,643,235,237 | 99.55 | 5.09 | 18.21 | 94.94 |
| NAG19572 | Kyoto Univ. | Kansai | 342,715,851 | 339,972,370 | 51,407,377,650 | 51,076,869,155 | 99.20 | 3.79 | 17.03 | 90.15 |
| NAG19576 | Kyoto Univ. | Kansai | 361,724,244 | 360,380,689 | 54,258,636,600 | 54,117,531,871 | 99.63 | 5.35 | 18.04 | 94.87 |
| NAG19587 | Kyoto Univ. | Kansai | 361,894,604 | 360,286,555 | 54,284,190,600 | 54,072,151,668 | 99.56 | 5.61 | 18.02 | 93.62 |
| NAG19607 | Kyoto Univ. | Kansai | 371,239,207 | 369,235,751 | 55,685,881,050 | 55,464,751,670 | 99.46 | 5.38 | 18.49 | 92.20 |
| NAG19608 | Kyoto Univ. | Kansai | 372,574,105 | 370,671,086 | 55,886,115,750 | 55,630,400,201 | 99.49 | 4.29 | 18.54 | 95.82 |
| NAG19626 | Kyoto Univ. | Kansai | 396,314,186 | 394,108,432 | 59,447,127,900 | 59,212,141,212 | 99.44 | 7.15 | 19.74 | 96.68 |
| NAG19627 | Kyoto Univ. | Kansai | 332,000,462 | 330,110,561 | 49,800,069,300 | 49,525,899,149 | 99.43 | 3.99 | 16.51 | 87.38 |
| NAG19630 | Kyoto Univ. | Kansai | 361,384,184 | 360,085,916 | 54,207,627,600 | 54,047,273,647 | 99.64 | 5.36 | 18.02 | 94.60 |
| NAG19632 | Kyoto Univ. | Kansai | 432,802,590 | 430,662,259 | 64,920,388,500 | 64,676,370,418 | 99.51 | 6.93 | 21.56 | 95.32 |
| NAG19642 | Kyoto Univ. | Kansai | 385,271,100 | 383,199,613 | 57,790,665,000 | 57,571,005,336 | 99.46 | 7.02 | 19.19 | 93.00 |
| NAG19648 | Kyoto Univ. | Kansai | 466,314,839 | 464,909,234 | 69,947,225,850 | 69,801,827,371 | 99.70 | 7.08 | 23.27 | 98.82 |
| NAG19664 | Kyoto Univ. | Kansai | 340,049,877 | 338,539,104 | 51,007,481,550 | 50,828,045,144 | 99.56 | 4.43 | 16.94 | 89.35 |
| NAG19666 | Kyoto Univ. | Kansai | 327,562,091 | 325,513,142 | 49,134,313,650 | 48,866,630,463 | 99.37 | 3.78 | 16.29 | 87.42 |
| NAG19675 | Kyoto Univ. | Kansai | 364,691,498 | 360,880,870 | 54,703,724,700 | 54,148,491,441 | 98.96 | 3.66 | 18.05 | 91.29 |
| NAG19679 | Kyoto Univ. | Kansai | 469,840,343 | 467,855,167 | 70,476,051,450 | 70,305,915,339 | 99.58 | 6.34 | 23.44 | 98.90 |
| NAG19685 | Kyoto Univ. | Kansai | 360,535,639 | 359,072,700 | 54,080,345,850 | 53,927,185,198 | 99.59 | 5.18 | 17.98 | 94.70 |
| NAG19688 | Kyoto Univ. | Kansai | 363,547,399 | 362,067,618 | 54,532,109,850 | 54,389,853,730 | 99.59 | 7.78 | 18.13 | 91.06 |
| NAG19691 | Kyoto Univ. | Kansai | 479,529,733 | 477,846,271 | 71,929,459,950 | 71,765,933,046 | 99.65 | 7.42 | 23.92 | 98.94 |
| NAG19706 | Kyoto Univ. | Kansai | 344,099,999 | 342,047,065 | 51,614,999,850 | 51,358,920,353 | 99.40 | 3.96 | 17.12 | 89.69 |
| NAG19707 | Kyoto Univ. | Kansai | 359,236,982 | 356,421,183 | 53,885,547,300 | 53,462,702,592 | 99.22 | 4.09 | 17.82 | 93.47 |
| NAG19720 | Kyoto Univ. | Kansai | 342,368,699 | 340,206,205 | 51,355,304,850 | 51,129,638,807 | 99.37 | 3.41 | 17.04 | 93.54 |
| NAG19729 | Kyoto Univ. | Kansai | 347,230,870 | 345,279,076 | 52,084,630,500 | 51,866,426,086 | 99.44 | 3.36 | 17.29 | 90.94 |
| NAG19736 | Kyoto Univ. | Kansai | 481,282,961 | 478,502,147 | 72,192,444,150 | 71,829,127,600 | 99.42 | 9.66 | 23.94 | 95.98 |
| NAG19744 | Kyoto Univ. | Kansai | 377,098,240 | 375,567,394 | 56,564,736,000 | 56,399,731,831 | 99.59 | 4.96 | 18.80 | 95.99 |
| NAG19745 | Kyoto Univ. | Kansai | 351,618,427 | 350,224,068 | 52,742,764,050 | 52,629,331,180 | 99.60 | 5.51 | 17.54 | 94.00 |
| NAG19753 | Kyoto Univ. | Kansai | 408,167,241 | 405,357,157 | 61,225,086,150 | 60,819,490,973 | 99.31 | 4.66 | 20.27 | 97.41 |
| NAG19757 | Kyoto Univ. | Kansai | 387,370,252 | 385,970,823 | 58,105,537,800 | 57,963,211,609 | 99.64 | 6.01 | 19.32 | 93.54 |
| NAG19769 | Kyoto Univ. | Kansai | 363,092,043 | 356,450,256 | 54,463,806,450 | 53,480,645,830 | 98.17 | 3.55 | 17.83 | 91.18 |
| NAG19775 | Kyoto Univ. | Kansai | 376,885,256 | 375,389,282 | 56,532,788,400 | 56,375,510,559 | 99.60 | 4.89 | 18.79 | 95.98 |
| NAG19786 | Kyoto Univ. | Kansai | 360,931,262 | 358,891,759 | 54,139,689,300 | 53,936,127,674 | 99.43 | 6.28 | 17.98 | 94.41 |
| NAG19789 | Kyoto Univ. | Kansai | 365,120,150 | 362,332,100 | 54,768,022,500 | 54,407,817,194 | 99.24 | 5.27 | 18.14 | 94.43 |
| NAG19797 | Kyoto Univ. | Kansai | 367,527,049 | 365,236,096 | 55,129,057,350 | 54,831,101,850 | 99.38 | 3.84 | 18.28 | 95.31 |
| NAG19800 | Kyoto Univ. | Kansai | 393,167,991 | 390,096,587 | 58,975,198,650 | 58,504,475,049 | 99.22 | 4.63 | 19.50 | 96.48 |
| NAG19817 | Kyoto Univ. | Kansai | 351,075,317 | 350,033,694 | 52,661,297,550 | 52,624,824,738 | 99.70 | 5.52 | 17.54 | 94.37 |
| NAG19819 | Kyoto Univ. | Kansai | 339,395,600 | 337,316,876 | 50,909,340,000 | 50,641,700,753 | 99.39 | 5.45 | 16.88 | 91.57 |
| NAG19834 | Kyoto Univ. | Kansai | 344,949,085 | 343,409,066 | 51,742,362,750 | 51,595,356,913 | 99.55 | 5.94 | 17.20 | 92.52 |
| NAG19838 | Kyoto Univ. | Kansai | 353,479,927 | 351,557,029 | 53,021,989,050 | 52,787,494,929 | 99.46 | 3.81 | 17.60 | 91.11 |
| NAG19846 | Kyoto Univ. | Kansai | 354,851,431 | 352,431,318 | 53,227,714,650 | 52,914,621,444 | 99.32 | 4.79 | 17.64 | 93.74 |
| NAG19849 | Kyoto Univ. | Kansai | 340,980,242 | 339,942,984 | 51,147,036,300 | 51,078,062,737 | 99.70 | 5.50 | 17.03 | 92.96 |
| NAG19861 | Kyoto Univ. | Kansai | 351,011,337 | 349,620,834 | 52,651,700,550 | 52,516,154,955 | 99.60 | 3.71 | 17.51 | 94.59 |
| NAG19895 | Kyoto Univ. | Kansai | 347,931,216 | 346,251,903 | 52,189,682,400 | 51,998,634,906 | 99.52 | 3.76 | 17.33 | 94.14 |
| NAG19908 | Kyoto Univ. | Kansai | 396,811,896 | 394,740,784 | 59,521,784,400 | 59,154,558,370 | 99.48 | 4.58 | 19.72 | 96.53 |
| NAG19920 | Kyoto Univ. | Kansai | 334,103,886 | 331,589,075 | 50,115,582,900 | 49,679,118,680 | 99.25 | 3.84 | 16.56 | 89.34 |
| NAG19921 | Kyoto Univ. | Kansai | 360,883,524 | 358,695,659 | 54,132,528,600 | 53,803,219,639 | 99.39 | 4.66 | 17.93 | 93.96 |
| NAG19922 | Kyoto Univ. | Kansai | 332,477,309 | 330,243,757 | 49,871,596,350 | 49,594,155,564 | 99.33 | 4.03 | 16.53 | 88.44 |
| NAG19927 | Kyoto Univ. | Kansai | 328,314,175 | 326,321,698 | 49,247,126,250 | 49,027,623,207 | 99.39 | 3.45 | 16.34 | 91.54 |
| NAG19929 | Kyoto Univ. | Kansai | 376,288,973 | 374,770,427 | 56,443,345,950 | 56,337,801,389 | 99.60 | 5.98 | 18.78 | 92.97 |
| NAG19943 | Kyoto Univ. | Kansai | 366,040,933 | 360,511,782 | 54,906,139,950 | 54,129,896,957 | 98.49 | 4.40 | 18.04 | 92.82 |
| Total / Average | | | 1,250,620,265,025 | 1,243,698,537,104 | 187,593,039,753,750 | 186,415,856,745,066 | 99.45 | 6.53 | 19.74 | 94.29 |
| Standard deviation | | | | | | | 0.25 | 2.04 | | 3.24 |

Supplementary Table S2. Description of the haplotype reference panels

| Abbreviation | JHRP | 1KGP | JPT | GAsP | HRC | TOPMed |
|---|---|---|---|---|---|---|
| Full name | Japanese Haplotype Reference Panel | 1000 Genome Project (All population) | 1000 Genome Project (Japanese Tokyo) | Genome Asia Pilot | Haplotype Reference Consortium | Trans-Omics for Precision Medicine |
| Description | Japanese diversity panel | 26 cohorts worldwide | Single Japanese cohort | East and Southeast asian population including 152, 3 and 35 Korean, Taiwanese and Japanese respectively | European | Multiethnic |
| Number of haplotypes | 6,270 | 5,008 | 208 | 3,308 | 64,940 | 194,512 |
| | call-rate<99% | Call-rate<95% | Call-rate<95% | VQSR (99% of the true sites) | ad hoc methods in ref. 4 or Ts/Tv < 1.7 | Support vector machine approach |
| SNVs Exclusion criteria | HWE-p<1x10$^{-6}$ | | HWE-p<1x10$^{-6}$ | Michigan imputation server QCs | HWE-p<1x10$^{-10}$ inbreeding coefficient < −0.1 | |
| | MAC<2 | MAC<2 | MAC<2 | | MAC<5 | MAC<5 |
| Number of autosomal SNVs after QC | 21,033,874 | 43,259,822 | 8,184,007 | 21,494,814 | 39,635,008 | 282,841,184 |
| Phasing and Imputation software | SHAPEIT2, Minimac4 | SHAPEIT2, Minimac4 | SHAPEIT2, Minimac4 | EAGLE2.4, Minimac4 | EAGLE2.4, Minimac4 | EAGLE2.4, Minimac4 |
| Imputation pipeline | in-house | in-house | in-house | Michigan imputation server | Michigan imputation server | TOPMed imputation server |
| Number of high quality variant after imputation (rsq>0.8) | 7,880,938 | 5,594,371 | 5,135,969 | 4,519,307 | 5,428,451 | 8,278,173 |
| references | | 1 | 1 | 2, 3 | 2, 4 | 5 |

SNV : Single-nucleotide Variant, QC : Quality control, MAC : Minor Allele Count.

Supplementary Table S3. Summary of mapping statistics

| Institute | Geographical region | Read info. | #Sample | Number of reads | | Number of bases | | Percent | | | Coverage |
|---|---|---|---|---|---|---|---|---|---|---|---|
| | | | | Total | Mapped | Total | Mapped | Mapped reads | PCR duplicates | Target covered† | |
| Kyoto University | Kansai | 2 x 150bp PE | 1,322 | 481,549,887,294 | 478,728,015,330 | 72,232,483,094,100 | 71,892,066,079,209 | 99.41 ± 0.28 | 5.62 ± 1.33 | 92.95 ± 3.10 | 18.13 ± 1.69 |
| Aichi Cancer Center | Kansai | | 458 | 180,522,453,114 | 179,735,183,701 | 27,078,367,967,100 | 27,017,907,314,810 | 99.57 ± 0.14 | 6.73 ± 1.92 | 94.31 ± 2.91 | 19.66 ± 3.15 |
| St. Marianna University | Kyushu | | 361 | 136,488,916,798 | 135,681,391,237 | 20,473,337,519,700 | 20,387,693,381,262 | 99.41 ± 0.22 | 6.62 ± 1.63 | 93.55 ± 3.37 | 18.83 ± 1.87 |
| BBJ | Kanto | | 1,007 | 452,059,007,819 | 449,553,946,836 | 67,808,851,172,850 | 67,118,189,969,785 | 99.45 ± 0.23 | 7.58 ± 2.41 | 96.32 ± 2.38 | 22.22 ± 2.48 |
| | | | 3,148 | 1,250,620,265,025 | 1,243,698,537,104 | 187,593,039,753,750 | 186,415,856,745,066 | 99.45 ± 0.25 | 6.53 ± 2.04 | 94.29 ± 3.24 | 19.74 ± 2.86 |

† Percent of total targeted bases covered at least 10x depths.

Supplementary Table S4. Admixture analysis of the Japanese population

**a**

| Population | Japanese | Korean | Han | Tujia | She | Miao | Ami | Hezhen | Oroqen | Atayal | Xibo | Yi | Naxi | Kinh | Dai | Lahu | Daur | Tu | Thai | Burmese | Cambodian | Uygur |
|---|---|---|---|---|---|---|---|---|---|---|---|---|---|---|---|---|---|---|---|---|---|---|
| Country | Japan | Korea | China | China | China | China | Taiwan | China | China | Taiwan | China | China | China | Vietnam | China | China | China | China | Thailand | Myanmar | Cambodia | China |
| Hondo | 0.219 | 0.217 | 0.215 | 0.214 | 0.214 | 0.213 | 0.212 | 0.212 | 0.211 | 0.211 | 0.211 | 0.211 | 0.210 | 0.209 | 0.209 | 0.207 | 0.207 | 0.205 | 0.202 | 0.199 | 0.197 | 0.172 |
| Ryukyu | 0.217 | 0.212 | 0.211 | 0.210 | 0.210 | 0.209 | 0.209 | 0.208 | 0.207 | 0.208 | 0.207 | 0.207 | 0.206 | 0.205 | 0.206 | 0.204 | 0.203 | 0.201 | 0.199 | 0.196 | 0.194 | 0.170 |
| Korean | 0.217 | 0.219 | 0.218 | 0.217 | 0.217 | 0.215 | 0.214 | 0.214 | 0.213 | 0.212 | 0.213 | 0.213 | 0.213 | 0.211 | 0.211 | 0.209 | 0.209 | 0.208 | 0.204 | 0.202 | 0.198 | 0.173 |
| H/R mix | 0.218 | 0.215 | 0.213 | 0.212 | 0.212 | 0.211 | 0.211 | 0.210 | 0.209 | 0.209 | 0.209 | 0.208 | 0.208 | 0.207 | 0.207 | 0.206 | 0.205 | 0.203 | 0.200 | 0.198 | 0.196 | 0.171 |
| Ainu | 0.216 | 0.211 | 0.209 | 0.209 | 0.208 | 0.207 | 0.207 | 0.208 | 0.207 | 0.206 | 0.206 | 0.205 | 0.204 | 0.204 | 0.204 | 0.203 | 0.202 | 0.200 | 0.197 | 0.195 | 0.193 | 0.169 |

**b**

| Population | Ulchi | Mongola | Yakut | Even | Eskimo Chaplin | Itelman | Eskimo Sireniki | Eskimo Naukan | Altaian | Kyrgyz | Tubalar | Aleut | Mansi | Chukchi | Tlingit |
|---|---|---|---|---|---|---|---|---|---|---|---|---|---|---|---|
| Country | Russia | China | Russia | Russia | Russia | Russia | Russia | Russia | Russia | Kyrgyzystan | Russia | Russia | Russia | Russia | Russia |
| Ainu | 0.211 | 0.207 | 0.199 | 0.200 | 0.198 | 0.197 | 0.196 | 0.196 | 0.190 | 0.184 | 0.179 | 0.175 | 0.171 | 0.170 | 0.163 |
| Ryukyu | 0.209 | 0.208 | 0.199 | 0.199 | 0.197 | 0.196 | 0.195 | 0.194 | 0.189 | 0.184 | 0.179 | 0.174 | 0.171 | 0.170 | 0.162 |
| Hondo | 0.212 | 0.211 | 0.202 | 0.202 | 0.199 | 0.198 | 0.197 | 0.197 | 0.193 | 0.187 | 0.181 | 0.176 | 0.172 | 0.171 | 0.163 |
| Korean | 0.212 | 0.213 | 0.204 | 0.203 | 0.201 | 0.199 | 0.199 | 0.199 | 0.194 | 0.189 | 0.182 | 0.177 | 0.173 | 0.172 | 0.164 |
| H/R mix | 0.211 | 0.210 | 0.201 | 0.201 | 0.198 | 0.197 | 0.196 | 0.196 | 0.191 | 0.186 | 0.180 | 0.175 | 0.172 | 0.171 | 0.162 |

**c**

| Population | Igorot | Dusun | Hawaiian | Bougainville | Maori | Australian | Papuan |
|---|---|---|---|---|---|---|---|
| Country | Philippines | Brunei | USA | Papua New Guinea | New Zealan d | Australia | Papua New Guinea |
| Ainu | 0.205 | 0.202 | 0.196 | 0.167 | 0.158 | 0.159 | 0.156 |
| Ryukyu | 0.207 | 0.203 | 0.198 | 0.167 | 0.159 | 0.158 | 0.157 |
| Hondo | 0.210 | 0.206 | 0.200 | 0.167 | 0.161 | 0.158 | 0.156 |
| Korean | 0.212 | 0.208 | 0.202 | 0.168 | 0.161 | 0.157 | 0.156 |
| H/R mix | 0.209 | 0.205 | 0.199 | 0.167 | 0.160 | 0.158 | 0.156 |

**d**

| Population | Kusunda | Hazara | Khonda Dora | Bengali | Relli | Mala | Kapu | Madiga | Irula | Yadava | Burusho | Punjabi | Brahmin | Sindhi | Pathan | Kalash | Balochi | Brahui | Makrani |
|---|---|---|---|---|---|---|---|---|---|---|---|---|---|---|---|---|---|---|---|
| Country | Nepal | Pakistan | India | Bangladesh | India | India | India | India | India | India | Pakistan | Pakistan | India | Pakistan | Pakistan | Pakistan | Pakistan | Pakistan | Pakistan |
| Ainu | 0.181 | 0.170 | 0.168 | 0.154 | 0.153 | 0.153 | 0.152 | 0.152 | 0.151 | 0.150 | 0.148 | 0.148 | 0.146 | 0.143 | 0.143 | 0.141 | 0.137 | 0.136 | 0.133 |
| Ryukyu | 0.181 | 0.171 | 0.168 | 0.154 | 0.153 | 0.152 | 0.152 | 0.152 | 0.151 | 0.150 | 0.148 | 0.148 | 0.146 | 0.143 | 0.143 | 0.141 | 0.137 | 0.136 | 0.132 |
| Hondo | 0.184 | 0.172 | 0.169 | 0.154 | 0.153 | 0.152 | 0.152 | 0.152 | 0.151 | 0.150 | 0.149 | 0.148 | 0.146 | 0.143 | 0.142 | 0.141 | 0.137 | 0.136 | 0.132 |
| Korean | 0.185 | 0.174 | 0.169 | 0.154 | 0.153 | 0.152 | 0.152 | 0.151 | 0.151 | 0.150 | 0.149 | 0.148 | 0.146 | 0.142 | 0.142 | 0.140 | 0.136 | 0.135 | 0.132 |
| H/R mix | 0.183 | 0.171 | 0.168 | 0.154 | 0.153 | 0.152 | 0.152 | 0.152 | 0.151 | 0.150 | 0.148 | 0.148 | 0.146 | 0.143 | 0.142 | 0.140 | 0.137 | 0.135 | 0.132 |

Pairwise estimates of outgroup f3 [X, Y; YRI] statistics for East Asia (a), Central Asia/Siberia (b), Oceania (c), and South Asia (d) samples from Simons Genome Diversity Project (SGDP) are shown. Dark and light red colors represent high and low values for each statistic.

Supplementary Table S5. Number of pLOF variants

| | Total | Type of variants | | | | Per individual ($n$ = 3,135) | | | |
|---|---|---|---|---|---|---|---|---|---|
| | | Singleton | Stop gained | Frameshift | Splice | Mean | Median | Min | Max |
| pLOF | 22,617 | 14,662 | 7,664 | 9,382 | 6,282 | 278 | 278 | 226 | 368 |
| pLOF(MAF<1%) | 21,612 | 14,823 | 7,394 | 8,855 | 5,998 | 20.4 | 20 | 6 | 93 |

Supplementary Table S6. Knock-out-tolerant genes.

| Gene | Drug name |
|---|---|
| AOC3 | hydralazine |
| BAZ2A | under development |
| CD33 | gemtuzumab ozogamicin,oncolysin m,hum195/rgel,hum-195-ac-225,hum-195-bi-213,bi-836858,lintuzumab,sgn-cd33a |
| CLCN3 | under development |
| CMA1 | km-01221,jnj-10311795,bay 11-42524,asb17061 |
| CNR2 | khk-6188,delta-9-tetrahydrocannabinol prodrugs,mda-19,kn-38-7271,ly-2828360,gw-42004,nabilone,jwh-051,rq-00202730,am-577,4-tetrahydro-1,s-777469,2-trimethyl-1,dronabinol oral solution,8-naphthyridine-4-carboxamide (enantiomeric mix),pxs-2076,sch-036,hu-433,842166x,6-(4-chlorophenyl)-7-(2,a-796260,3,4-dichlorophenyl)-n-(hydroxymethyl)-1,prs-211375 iv,prs-639058,tak-937,ar-xyz,research programme,2 |
| CPT1B | perhexiline,c75,etomoxir,l-carnitine,sdz-cpi-975,db-200 |
| CTSH | under development |
| CYP2D6 | isoquine,glutethimide,bms-694153 |
| GIPR | rg7697,rg7685 |
| HLA-B | velimogene aliplasmid |
| IFNGR2 | interferon gamma-1b |
| IL25 | alx-0761 |
| KCNA10 | dalfampridine |
| LDHB | stiripentol |
| LTK | under development |
| MMP8 | cipemastat,bb-1101,marimastat |
| SUCNR1 | under development |
| VEGFA | at001/r84,aflibercept,bevasiranib,bevacizumab + erlotinib,bevacizumab,bevacizumab + rituximab,bevacizumab + trastuzumab,rg7221,mp-0112,aln-veg01,avastin+/-tarceva,ranibizumab,ro5520985,ptc299,sflt-01,snn-0029 |
